# Distinct genetic liability profiles define clinically relevant patient strata across common diseases

**DOI:** 10.1101/2023.05.10.23289788

**Authors:** Lucia Trastulla, Sylvain Moser, Laura T. Jiménez-Barrón, Till F.M. Andlauer, Moritz von Scheidt, Schizophrenia Working Group of the Psychiatric Genomics Consortium, Monika Budde, Urs Heilbronner, Sergi Papiol, Alexander Teumer, Georg Homuth, Peter Falkai, Henry Völzke, Marcus Dörr, Thomas G. Schulze, Julien Gagneur, Francesco Iorio, Bertram Müller-Myhsok, Heribert Schunkert, Michael J. Ziller

## Abstract

Genome-wide association studies have unearthed a wealth of genetic associations across many complex diseases. However, translating these associations into biological mechanisms contributing to disease etiology and heterogeneity has been challenging. Here, we hypothesize that the effects of disease-associated genetic variants converge onto distinct cell type specific molecular pathways within distinct subgroups of patients. In order to test this hypothesis, we develop the CASTom-iGEx pipeline to operationalize individual level genotype data to interpret personal polygenic risk and identify the genetic basis of clinical heterogeneity. The paradigmatic application of this approach to coronary artery disease and schizophrenia reveals a convergence of disease associated variant effects onto known and novel genes, pathways, and biological processes. The biological process specific genetic liabilities are not equally distributed across patients. Instead, they defined genetically distinct groups of patients, characterized by different profiles across pathways, endophenotypes, and disease severity. These results provide further evidence for a genetic contribution to clinical heterogeneity and point to the existence of partially distinct pathomechanisms across patient subgroups. Thus, the universally applicable approach presented here has the potential to constitute an important component of future personalized medicine concepts.

## Introduction

Understanding the molecular basis of complex diseases, that result from the joint contribution of many genomic variants in conjunction with environmental factors, remains one of the major challenges of contemporary medical research. Genome-wide association studies (GWAS) identified hundreds of disease susceptibility loci across a spectrum of complex diseases^1,2^, but it remains challenging translate these associations into insights on molecular pathomechanisms. These challenges are rooted in the highly polygenic nature of complex diseases with small effect sizes of individual variants^3^, with most variants residing in the non-coding space of the genome with unknown function^4,5^. In addition, the high level of heterogeneity in symptoms, severity, and treatment response likely reflects differences in the underlying genetic basis^6^ of patient populations that are presently considered as homogeneous groups.

Thus, there is currently a critical gap between our insights into the disease association of individual genetic variants and the aggregated impact of these variants on biological processes and clinically relevant parameters. This gap constitutes one of the major obstacles on the road towards the implementation of personalized medicine and the operationalization of genetic information in clinical decision making^7^.

A key step towards resolving this problem was the development of transcriptome-wide association studies (TWAS). This approach combines genotype-based prediction of individual and tissue specific gene expression levels based on all SNPs within a large cis window of each gene with disease association testing. This strategy leverages either GWAS summary statistics^8^ or performs the association testing directly on cohorts with available disease/trait status information^9^. TWAS offers the great advantage of aggregating trait heritability using biologically meaningful concepts (gene regulation) and entities (genes), while at the same time dramatically reducing the multiple testing burden to discover novel gene-trait associations. At present, available strategies rely on gene level aggregation and do allow for further genetic risk aggregation on the individual level.

In parallel, distinct types of polygenic risk score (PRS) concepts were developed to resolve genetic heterogeneity among patients and identify individuals at higher risk for a particular diagnosis or trait expression. These concepts rely on the aggregation of SNP-trait association weights on the level of all SNPs below a certain association cutoff, the individual gene^10^ or pathway^11,12^ and enable systematic testing for PRS association with various other traits of interest. This approach of stratifying individuals according to genetic disease risk provided increased detection power to discover associations between different types of PRS and intermediate phenotypes or clinically relevant endpoints^11,13,14^ such as disease severity^15^.

Conversely, patient stratification on the clinical and endophenotype level found ample evidence for distinct clinical subgroups, such as e.g. in heart failure^16^, type 1 diabetes^17^, or recently for MDD and suggested distinct genetic liability profiles of overall PRS between these phenotypically defined groups^18^.

Alternative strategies to genetically resolve heterogeneity in clinical and intermediate phenotype variables leveraged genetic correlations with other traits to discover evidence for patient subgroups with specifically higher correlation for a particular trait of interest. However, the latter did not detect the presence of such groups in most analyzed traits such as SCZ, MDD or diabetes^19,20^.

Current approaches to genotype-based patient stratification rely on univariate genetic scores for a priori defined traits or specific hypothesis driven genes/pathways, resulting in a dichotomous classification of patients.

These strategies are supervised in nature and require detailed insights on potential disease mechanisms, rendering an unbiased discovery of subgroups and potential group specific genetic liabilities difficult. Moreover, traditional PRS based stratification approaches are agnostic to the underlying biological mechanisms, rendering the biological interpretation of patient strata challenging.

To overcome these current limitations, we sought to operationalize personal genetic profiles to stratify patients into biologically distinct subgroups in an unbiased and unsupervised manner and specifically address the question: How does heterogeneity in polygenic risk factor distribution contribute to heterogeneity in clinical parameters, severity, and treatment response across patient populations?

We therefore develop the CASTom-iGEx framework (**Supplementary Fig. 1**) that builds on an improved gene expression imputation method and on the concept of pathway activity association studies. We utilize this analytical approach to identify genes and pathways associated with complex diseases, providing novel associations when applied to coronary artery disease (CAD) and schizophrenia (SCZ). Finally, we leverage these results to perform unbiased, multivariate stratification of the patient population into distinct subgroups. We show that these groups differ with respect to the distribution of disease liability across disease relevant biological processes, intermediate phenotypes, and clinical outcome.

## Results

### Imputation of gene and pathway level activities

We first sought to better understand which genes and biological processes constitute targets of aggregated polygenic effects across individuals suffering from genetically complex diseases. We implemented an improved method to predict cell type specific gene expression from individual level genotype data (PriLer) and perform transcriptome and pathway wide association analyses (**Supplementary Fig. 1-6, Supplementary Tables 1** see Methods and Supplementary Text for details). The PriLer algorithm leverages matched transcriptome and genetic data as well as prior biological relevance of genetic variants. It iteratively adapts the relevance (weights) of all variants in a machine learning framework to model the cis-genetic component of gene expression. Importantly, PriLer allows to trace the impact of individual SNPs from their GWAS association signal to the final gene expression contribution.

Paradigmatic application of this method to coronary artery disease (CAD) using 19,026 CAD cases and 321,916 control individuals from the UK Biobank (UKBB) across 11 tissues from the GTEx project identified 180 genes across 83 loci (FDR ≤ 0.05, **Supplementary Fig. 7**). Of these, 48 genes across 33 loci were not previously implied in CAD^21^ (e.g. *NME7* and *NFU1*) (**Fig. 1a, Supplementary Data 2**). Importantly, the integration of GWAS signal, epigenetic annotation, and gene regulatory information effectively aids the priorization of disease relevant and functionally relevant SNPs consistent with all layers of information (**Fig. 1b**).

**Fig. 1:**
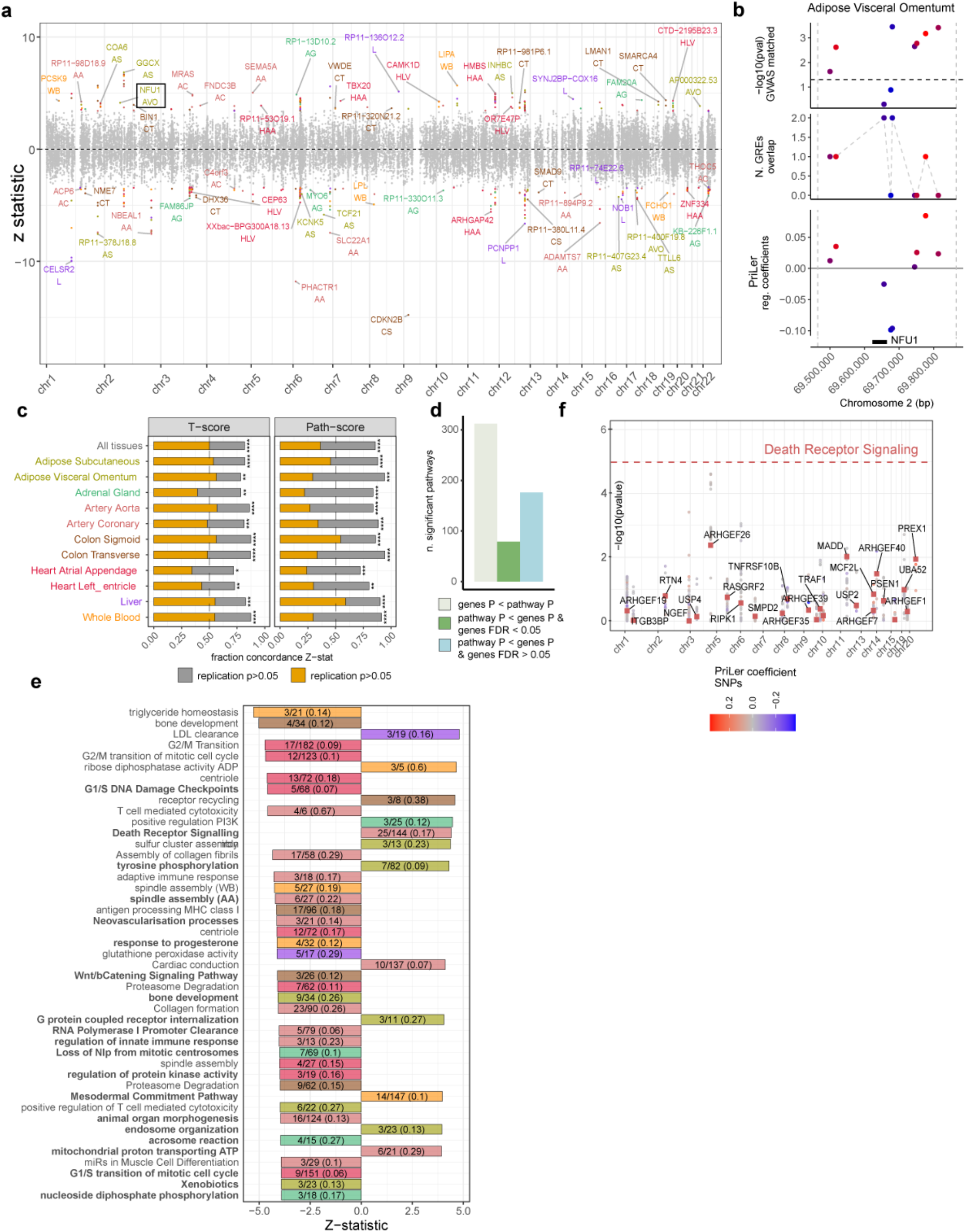
Genes and pathways associated with CAD. **a**. Manhattan plot showing Z-statistic across 11 tissues, colored dots refers to genes with tissue specific FDR ≤ 0.05. Acronyms in parenthesis indicate the initials of the tissue considered (AS = Adipose Subcutaneous, AVO = Adipose Visceral Omentum, AG = Adrenal Gland, AA = Artery Aorta, AC = Artery Coronary, CS = Colon Sigmoid, CT = Colon Transverse, HAA = Heart Atrial Appendage, HLV = Heart Left Ventricle, L = Liver, WB = Whole Blood). **b**. PriLer model for NFU1 in adipose visceral omentum. Each dot represents a variant having PriLer regression coefficient different from zero ordered on the x-axis according to its corresponding genomic position and colored based on PriLer coefficients values. Panels from the bottom to the top indicate: 1) genomic position of NFU1 with dashed lines representing TSS +/- 200kb window, 2) regression coefficient from PriLer model, 3) number of gene regulatory elements in the PriLer model that a variant intersects, 4) -log10 p-value from matched GWAS in UKBB (Methods). **c**. Reproducibility of gene levels T-scores (left) and pathway scores (right) via meta-analysis of CARDIoGRAM cohorts. X-axis shows the fraction of significant genes in UKBB that have the same effect sign (Z-statistic) in CARDIoGRAM meta-analysis, p-values are computed from one-sided sign test (* = P ≤ 0.05, ** = P ≤ 0.01, * * * = P ≤ 0.001, * * ** = P ≤ 0.0001). The fraction of genes concordant and nominal at a p-value threshold of 0.05 is shown in the yellow bar. **d**. Number of significant pathways (PALAS FDR ≤ 0.05) with at least one gene reaching better significance than the pathway (ivory), with all genes in the pathway less significant but with at least one gene having TWAS FDR ≤ 0.05 (green), and all genes less significant and not passing TWAS FDR 0.05 threshold (light blue). **e**. Reactome Death Receptor Signaling in artery aorta. The pathway significance is indicated by the dashed horizontal line, the coloured squares show genes included in that pathway and the corresponding TWAS p-value (y-axis) and the dots indicate the matched GWAS p-value of SNPs regulating those genes with colour reflecting PriLer regulatory coefficients. **f**. Among pathways more significant than any included gene, 45 prioritized pathways based on the following criteria: computed from more than 5 and less or equal than 200 t-score genes or more than 2 if pathway coverage is higher than 10%, originally including less than 200 genes and reaching at least 0.0001 as nominal significance. PALAS Z-statistic is shown in the x-axis color coded by tissue origin. Each pathway barplot contains the gene pathway coverage. The pathway name in bold reflects pathways without any significant gene (FDR > 0.05).

Replication analysis of these results based on a subset of the CARDIoGRAM cohorts^21^ including 13,279 CAD cases and 13,402 revealed high sign reproducibility of 82% (one-sided sign test P=4.35e-38) and an additional nominal reproducibility at 0.05 of 50% (**Fig. 1c**). Jointly, these analyses underscore the reliability of the PriLer approach to conducting TWAS studies and its improved capacity to pinpoint biologically relevant genetic variants contributing gene expression regulation (**Supplementary Fig. 3e, 6**).

In order to aggregate weak genetic effects further, we extended the concept of TWAS to the pathway/gene set level and performed a pathway level association study (PALAS). This methodology involves aggregating predicted gene-level scores into continuous pathway activity level scores at the individual level, using a predefined set of pathways from GO biological processes^22^, Reactome^23^ and WikiPathways^24^ (see Methods). Extensive validation of this approach on permuted data from CAD patients confirmed the well calibrated nature of this approach (**Supplementary Fig. 8**). This strategy identified 567 significant pathways across all tissues to be associated with CAD (FDR ≤ 0.05), with most pathways detected in artery aorta (**Supplementary Fig. 9a, Supplementary Data 3**).

Importantly, the PALAS methodology detected substantially more CAD associated pathways compared to more traditional pathway enrichment strategies such as hypergeometric testing of TWAS significant genes or MAGMA^25^ that each rely on summary statistics (**Supplementary Fig. 9b**). This increase in detection power resulted predominantly from the capacity of this approach to aggregate over weak association effects (**Fig. 1d**). For 45% of genome wide significant pathways, the PALAS p-value is lower than that of any included gene based on TWAS. Similarly, 31% of the identified pathways did contain not a single individually genome wide significant gene and thus also show an aggregation effect. Detailed analysis of well-known confounders in gene set analysis^26^ such as LD and gene-gene correlation confirmed that increased detection power did not result from these factors (**Supplementary Fig. 10**) with results being moderately sensitive to training sample size (**Supplementary Fig. 11**).

Following further priorization (Methods), we identified 45 CAD associated pathways showing an aggregation effect, of which 21 that did not contain a single individually significant gene below the FDR cutoff, and hence considered novel (**Fig. 1d**). Following a similar priorization strategy for the remaining pathways that are disrupted by at least one significant gene identified 63 pathways dysregulated by 23 distinct genes (**Supplementary Fig. 9c**).

Importantly, this approach recapitulated the key biology of CAD while at the same time discovering new biological themes (**Fig. 1e, Supplementary Fig. 9d**, e.g. Death Receptor Signaling or peptidyl-tyrosine phosphorylation). Replication analysis using the CARDIoGRAM cohorts^21^ confirmed high reproducibility of the results with 86% of pathway associations replicating based on direction of effect (one-side sign test P = 10e−63) as well as 22%-54% (dependent on tissue) based on nominal significance in the CARDIoGRAM dataset (**Fig. 1c**).

In summary, these results indicate the individual level convergence of a polygenic disease architecture driven by many small effect variants on biological processes in CAD. Moreover, they underscore the added value of the CASTom-iGEx pipeline, providing a strategy to successively aggregate polygenic signal from SNPs to genes to pathways.

### Deconstructing genetic heterogeneity in patient populations

Based on these results, we asked whether gene and pathway level liability profiles would be equally distributed across the patient population or rather cluster within specific patient subgroups, indicative of distinct genetically driven pathomechanisms.

To test this hypothesis, we performed unsupervised clustering based on imputed gene level scores from each CAD patient (n = In a) for each tissue separately, correcting for ancestry contribution. (see Methods). This analysis identified between 3 and 5 groups of CAD patients **(Fig. 2a, Supplementary Fig. 12a**) that largely overlap for clustering results from different tissues (**Supplementary Fig. 12b**). Against this background and the relevance of liver in CAD pathophysiology, we focused on patient stratification based on liver profiles. Careful evaluation of patient group structure revealed that the latter was not driven by single genes, but rather by a combination of CAD associated genes from multiple independent loci (**Supplementary Fig. 12c, Supplementary Tables 2**). Moreover, analysis of well known confounding factors showed that group structure was not was not driven by age, sex or ancestry contributions (see the detailed analysis of the latter factors in Supp. Text “Investigation of ancestry contribution to clustering structure” and **Supplementary Fig. 13, 14**).

**Fig. 2:**
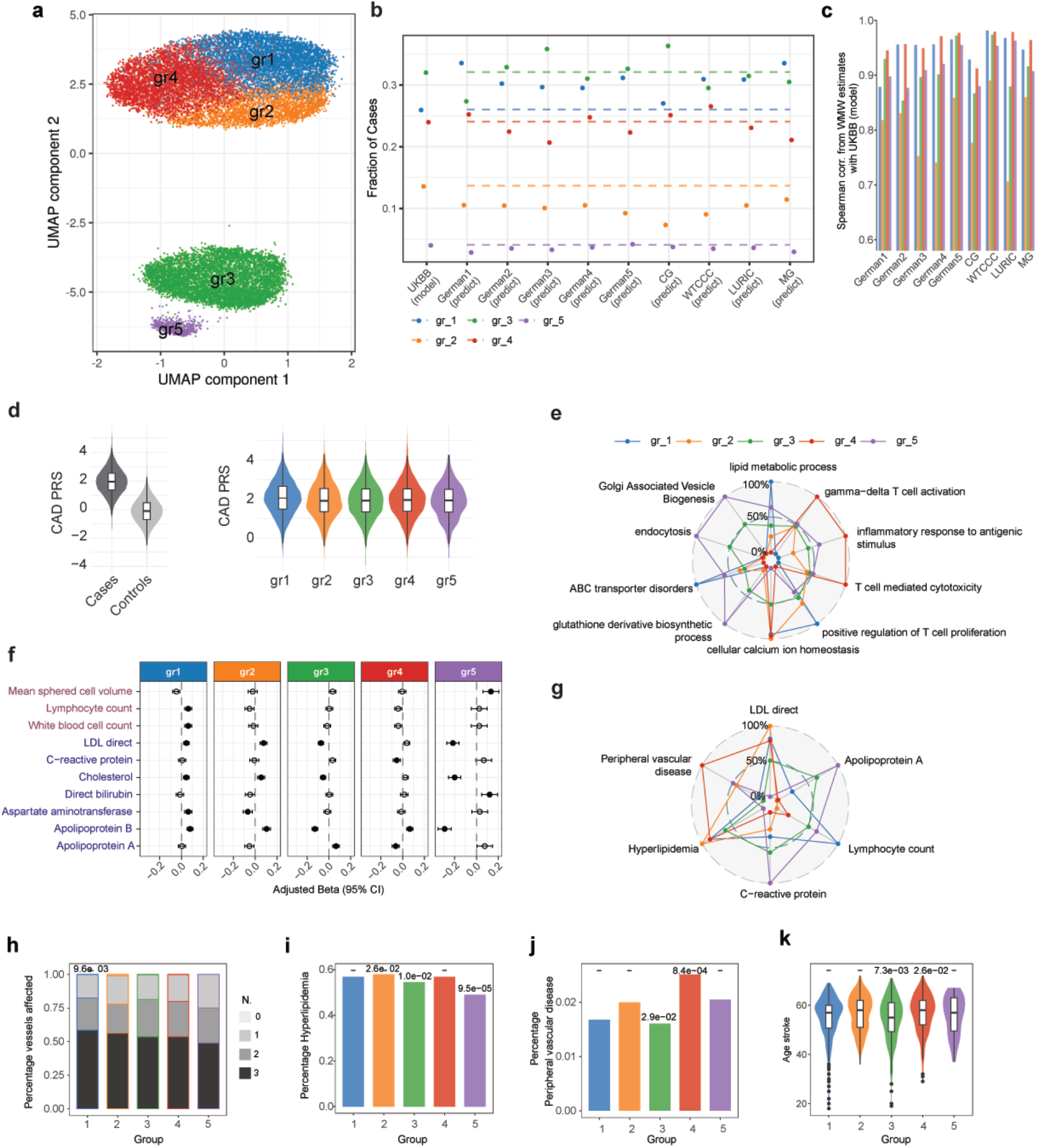
CAD patients genetically driven stratification from imputed gene expression in Liver. **a**. First 2 components of uniform manifold approximation and projection (UMAP) from gene T-scores in liver for CAD patients. Genes are clumped at 0.9 correlation, separately standardized and PCs corrected, and multiplied by Z-statistic CAD associations. Each dot represents a patient colored by the cluster membership. **b**. Prediction of clustering structure on 9 external CARDIoGRAM cohorts. Y-axis shows the fraction of cases assigned to each cluster in UKBB dataset and each external cohort for which the clustering structure was projected. The dashed lines indicate the fraction value for UKBB model clustering. **c**. For each group, Spearman correlation of WMW estimates in UKBB and each external cohort only from genes that are significantly associated with that group across all tissues. **d**. Left: Distribution of CAD polygenic risk score (PRS) for all UKBB individuals based on CAD GWAS summary statistics from UKBB CAD GWAS. Right: Distribution of CAD PRS across CAD cases divided by clustering group. **e**. Mean value of selected group-specific pathways in each group rescaled to 0-100 range. **f**. Among 212 endophenotypes measured in UKBB with at least one CAD associated and group specific pathway, forest plot shows significantly different ones (FDR ≤ 0.05) in at least one group (gr_i_ versus remaining patient) using Generalized Linear Model (GLM), indicating regression coefficient (β_GLM_) with 95% Confidence Interval (CI). Full dot means that β_GLM_ is significant (0.05 threshold) after BH correction performed separately for each group across all endophenotypes. The panel refers to continuous phenotypes, binary and ordinal categorical phenotypes are in Fig. S18. **g**. Mean value of selected group-specific endophenotypes in each group rescaled to 0-100 range. **h**. CAD severeness across projected clusters in GerMIFSV cohort. Y-axis indicates the percentage of patients with a certain number of vessel affected (grey shades). X-axis indicates the projected group. **i-j**. Percentage of patients in UKBB clustering with comorbidities ((i) hyperlipidemia, (j) peripheral vascular disease). **k**. Distribution of age of stroke for patients in UKBB. In (h-k) nominal p-values from group-wise GLM is shown at the top of the bar/violin plot. Boxplot elements include median as central line, 1^st^ and 3^rd^ quartiles as box limits, 1.5 interquartile ranges from 1^st^ and 3^rd^ quartiles as corresponding whiskers.

Strikingly, comparison of CAD polygenic risk scores (PRS) across groups revealed that the clustering results were highly distinct from PRS which were equally distributed across groups (**Fig. 2d**). These observations highlight the distinct layer of information captured by the CASTOM-iGEx stratification strategy.

To evaluate the generalizability and reproducibility of this patient stratification approach, we projected the imputed gene level score profiles from 9 independent CARDIoGRAM cohorts onto the clustering structure discovered on the UKBB dataset (see Methods). Subsequently, we determined the fraction of CARDIoGRAM CAD cases assigned to each cluster. This analysis revealed a virtually identical distribution of CAD cases across the clusters compared to the original UKBB dataset (**Fig. 2b**). Moreover, Spearman correlation analysis of the predicted gene expression profiles from the individual CARDIoGRAM cohorts and the UKBB dataset showed excellent concordance (cor. > 0.7, **Fig. 2c**), with WTCCC being the most consistent cohort (cor. > 0.88) and was not driven by a single locus (**Supplementary Fig. 13e**).

Subsequently, we determined the predicted differences in gene expression and pathway activity between all groups. We tested 36,397 genes for cluster specific association of predicted expression levels, identifying 887 genes-cluster associations across all tissues originating from 50 tissue-specific loci (**Supplementary Tables 2**), as well as 236 unique genes across all tissues originating from 16 loci (**Supplementary Fig. 15a**, FDR ≤ 0.01). PALAS analysis of 7,978 pathways across 11 tissues identified 271 unique pathways that exhibited a significantly different (FDR ≤ 0.01) liability distribution across patient groups (**Supplementary Fig. 15b**). Detailed evaluation of pathway association statistics across groups showed that cluster-specific gene and pathway association statistics were well-calibrated **(Supplementary Fig. 16a-e)**.

In order to further validate the molecular consequences of this increased genetic liability towards the de-regulation of genes and pathways, we employed an independent population-based cohort with genotyping as well as transcriptome data from whole blood (SHIP-TREND^27^). Projection of the respective individuals from the SHIP-TREND cohort onto the UKBB derived clustering structure using imputed gene expression profiles confirmed a distribution across the groups similar to the UKBB and CARDIoGRAM cohorts (**Supplementary Fig. 17a**). Differential gene level analysis between groups using the measured gene expression data confirmed 58% of predicted differentially expressed genes from imputation (Wilcoxon-Mann-Whitney (WMW) corrected FDR ≤ 0.05) based on sign concordance (64 out of 111 genes) and 18% on the nominal significance level in whole blood, with a general Pearson correlation of 0.38 (P=3.5e-5) (**Supplementary Fig. 17b**). Importantly, major drivers of the clustering were confirmed in the measured genes expression such as HLA-C in the MHC region, TMEM116 or PSRC1 genes (**Supplementary Fig. 17d**). This analysis revealed additional group specific transcriptome alterations indicative of genetically driven gene expression perturbations beyond cis-effects (**Supplementary Fig. 17e**). Groupwise pathway activity level effects showed an excellent correlation between predicted effects based on UKBB individuals and measured effects based on SHIP individuals (R_Spearman_=0.37, p=0.000396, **Supplementary Fig. 17c**).

These analyses support the existence of fundamental differences in genetic liability towards specific biological processes across individuals, suggesting genetically (partially) distinct patient subtypes.

### Genetically defined patient groups differ in disease relevant endophenotypes and clinical parameters

In order to understand the potential biological and phenotypic relevance of differences in pathway level liabilities, we performed association analysis of the latter pathways (PALAS) with 637 phenotypes from the UKBB. This analysis identified 212 endophenotypes (FDR ≤ 0.05) that were significantly connected to at least one group-specific and CAD associated pathway.

Based on these observations, we tested the hypothesis that the identified CAD patient groups would also differ on the phenotypic level with respect to any of the 212 endophenotypes, each connected to pathways with group-specific liabilities as well as 33 clinical phenotypes (see Methods). This analysis identified 36 cluster-specific endophenotype associations (FDR ≤ 0.05, **Supplementary Tables 3**) and 19 unique endophenotypes, all with high relevance to CAD biology (**Fig. 2e, Supplementary Fig. 18a-b**) and directly associated with the detected differences in underlying biological processes.

In particular, CAD group 1 and group 2 showed a specific reduction in Golgi Associated Vesicle Biogenesis, endocytosis, and endosome biology (**Fig. 2f**), concomitant with a significant increase of circulating LDL on the patient endophenotype level (**Fig. 2e,g**). This observation is consistent with the notion that vesicles filled with LDL are taken up by the cells via receptor-mediated endocytosis mechanisms^28^. Accordingly, circulating LDL level exhibit a strong genetic association with endocytosis related pathways (**Supplementary Fig. 18b**). Similarly, group 1 showed a significant increase in fatty acid and general lipid metabolic processes that were also significantly associated with circulating LDL levels and consistent with overall higher LDL and Cholesterol levels in patients of group 1 (**Fig. 2e-g**). Lastly, patients in group 1 exhibited an increase in immune cell populations, concomitant with a predicted increase in genes related to T cell proliferation (**Fig. 2f,g**).

We evaluated whether these differences in liabilities across genes, biological processes and endophenotypes were associated with differences in clinical parameters such as disease severity and/or trajectory (**Supplementary Tables 4**). To that end we leveraged additional clinical phenotypes collected on 2,383 CAD patients (GerMIFSV in CARDIoGRAM), evaluated between patient groups following their projection onto the UKBB clustering as well as 33 clinical parameters collected in UKBB. This analysis revealed that the projected patients of group 1 in GerMIFSV had a significantly higher number of vessels affected by CAD, indicative of a more severe disease course (**Fig. 2h**). In contrast, group 2 showed a higher incidence of hyperlipidemia (**Fig. 2i**). These observations are consistent with the overall higher levels of key CAD related endophenotypes (LDL, APOB) and elevated genetic liability towards the perturbation of lipid metabolism and endocytosis related pathways in group 2 (**Supplementary Fig. 18b**).

In contrast, group 3 showed comparatively low APOB and (LDL) cholesterol levels (**Fig. 2e,f**), an increased predicted activity of the N-acetyltransferase pathway (**Supplementary Fig. 15b**), previously implicated in cardiac dysfunction^29^. In addition, group 3 patients exhibited an increase in glutathione derivative biosynthetic processes, related to the cellular capacity to compensate against reactive oxygen species as well as a reduction in predicted gene activities related to ventricular system development (**Fig. 2f, Supplementary Fig. 15b**). Clinically, patients in group 3 demonstrate an increased frequency of chronic obstructive pulmonary disease and a decreased age of stroke (**Fig. 2k, Supplementary Tables 4**).

Conversely, patients assigned to group 4 were subject to an increased genetic liability towards many immune related pathways such as T cell activation or immunoglobulin mediated immune response (**Fig. 2f**), all of which were negatively associated with markers of inflammatory processes such as C-reactive protein (CRP, **Supplementary Fig. 18b)**. In line with this observation, group 4 patients showed decreased CRP levels compared to all other groups (**Fig. 2g**) and an increased frequency of peripheral vascular diseases and a slightly higher age of stroke (**Fig. 2j,k**). These observations suggest an increased relevance of inflammation related processes in CAD specifically in this subgroup of patients that is linked to distinct clinical characteristics.

Finally, group 5 showed the lowest levels CAD related endophenotypes, (**Fig. 2e,f**) as well as the lowest frequency of clinically relevant outcome parameters and other diseases (**Fig. 2h-j**). These observations suggest that group 5 represents the healthiest group of CAD patients. Simultaneously, group 5 exhibits the lowest genetic liabilities towards CAD associated biological processes but not with respect to PRS (**Fig. 2d**). Although not significant, we also observed a trend of increased CRP levels in group 5 (**Fig. 2e,g)** connected to endocytosis and glutathione biosynthesis liabilities (**Fig. 2f, Supplementary Fig. 18b)**. Interestingly, glucosamine consumption reduced CRP levels in group5 individuals, compared to all the other groups, where no decrease or even an opposite trend was observed (**Supplementary Fig. 18c-d**). This analysis suggests a possible cost-effective therapeutic strategy to decrease CRP for patients with precise genetic liabilities.

In order to further validate these observations, we leveraged the independent SHIP-Trend cohort and confirmed the observed groupwise differences in endophenotypes with respect to LDL distribution and CRP as well as additional differences in Carotid intima-media thickness (**Supplementary Fig. 17f)**. In contrast, random partitioning of patients into groups enabled very limited detection of endophenotypic differences in individual partitions (**Supplementary Fig. 16f)**.

In summary, the application of the unsupervised stratification strategy implemented in the CASTOM-iGEx framework suggests the existence of distinct CAD patient subtypes not identifiable by traditional PRS based approaches. These subgroups do not only differ with respect to their genetic risk distribution across biological processes, but also exhibit significant divergence in their disease relevant clinical and physiological parameters connected to these molecular differences.

### Identification of key biological processes impaired in SCZ

Going beyond well characterized CAD, we decided to evaluate the capacity of CASTOM-iGEx to obtain insights into the biology of a more enigmatic illness. To that end, we focused on SCZ, for which only limited insights into pathomechanisms, contributing endophenotypes, and disease subgroups are available. While the existence of clinical subtypes of SCZ patients is well known^30^, it is at present unclear, whether or not this phenotypic heterogeneity might result from a distinct genetic basis and potentially distinct biological mechanisms. To address these questions, we applied the CASTOM-iGEx pipeline to 36 European cohorts from Psychiatric Genomic Consortium (PGC) wave 2^31^ for a total of 24,764 cases and 30,655 controls, leveraging 9 GTEx tissues and DLPC (dorsolateral prefrontal cortex) gene expression data from the CommonMind consortium as training data^32^.

Similar to CAD, PALAS analysis identified 255 (Reactome), 692 (GO) and 125 (WikiPathways) unique pathways associated with SCZ (FDR ≤ 0.05), most detected in DLPC tissue (**Supplementary Fig. 19a, Supplementary Data 6**). Overall, 38% of the significant pathways showed an aggregation effect, with pathway level significance exceeding those of all participating genes (**Supplementary Fig. 19b**). Following prioritization and exclusion of genes located in the MHC locus (**Fig. 3a, Supplementary Fig. 19c**), this group (n=45, **Supplementary Data 6**) contained several genes sets not implied in SCZ through GWAS, but by other sources of evidence. These included genes related to Alzheimer’s disease^33^, degradation of the extracellular matrix, ErbB, and mTOR signaling related to myelination^34^ (**Fig. 3a)**. Similarly, we identified genetic evidence for TCA cycle, proteasome degradation, and impairment of the pyruvate dehydrogenase complex (**Fig. 3a**). Several pathways reflect clinically well-established comorbidities such as Insulin Signaling (e.g. Diabetes) or pathways in Pathogenesis of Cardiovascular Disease (**Fig. 3a**). Interestingly, we also identified genes enriched for de novo loss-of-function (LoF) mutations (**Supplementary Fig. 19d**) through rare variants in SCZ patients based on exome sequencing in multiple SCZ family studies^32^. This observation further supports the hypothesis of convergence between rare and common variants that affect the same genes and hence could be related to analogous pathomechanisms^35^.

**Fig. 3:**
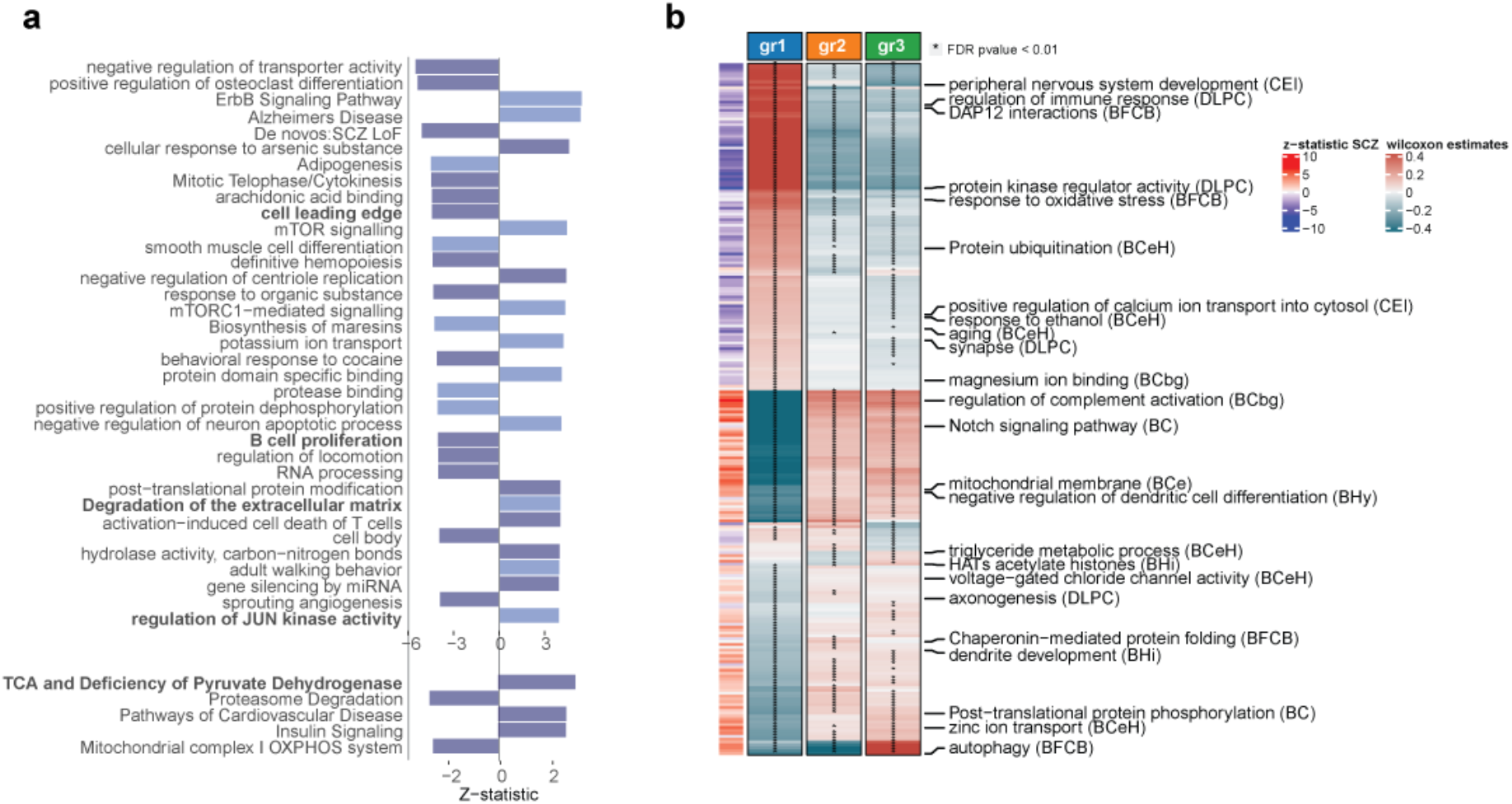
Impaired biological processes in SCZ. **a**. PALAS Z-statistic results for a selection of pathways. Among pathways more significant than any included gene and without any gene in the MHC locus, the top panel shows a subset of the 45 prioritized pathways based on the following criteria: computed from more than 5 and less or equal than 200 t-score genes or more than 2 if pathway coverage is higher than 10%, originally including less than 200 genes and reaching at least 0.0001 as nominal significance. PALAS Z-statistic is shown in the x-axis color coded by tissue origin (dark blue = DLPC in CMC, light blue = a brain region in GTEx). The bottom panel shows a selection of significant SCZ pathways in WikiPathway collection. The pathway name in bold reflects pathways without any significant gene (FDR > 0.05). **b**. Wilcoxon-Mann-Whitney (WMW) estimates for 241 group-specific pathways (FDR ≤ 0.05, Reactome and GO) including at least one gene in the MHC locus and considering only the most significant tissue per-pathways when repeated. The clustering is performed on SCZ patients in DLPC imputed gene expression, The row annotation on the left indicates the corresponding SCZ PALAS Z-statistics. The acronym in parenthesis in the pathway names refers to the tissue considered (DLPC = Dorsolateral Prefrontal Cortex in CMC, CEI = Cells EBV-transformed lymphocytes, BFBC = Brain Frontal Cortex BA9, BCeH = Brain Cerebellar Hemisphere, BCbg = Brain Caudate basal ganglia, BC = Brain Cortex, BCe = Brain Cerebellum, BHi = Brain Hippocampus, BHy = Brain Hypothalamus).

Importantly, a systematic analysis of this gene aggregation strategy (**Supplementary Fig. 19e**, Supplementary Text) revealed that the significance increment originated from the aggregation of distinct patients’ liability profiles that converge onto the same genetic targets, providing an explanation why this approach enables the implication of additional molecular processes in disease biology.

### Deconstructing heterogeneity among SCZ patients

Building on these results, we evaluated the hypothesis, whether or not genetic heterogeneity across gene and pathway level liability profiles would give rise to distinct SCZ patients subgroups. Following a similar strategy as applied for CAD (Methods), we detected 3 groups of SCZ patients based on clustering of 5,678 gene T-scores from DLPC (**Supplementary Fig. 20, Supplementary Fig. 21a**). Detailed analysis of potential confounders revealed minimal impact of ancestry and cohort membership on clustering structure as well as on detected gene associations (**Supplementary Fig. 21c-e, Supplementary Fig. 22**).

In total, we identified 594 cluster-specific genes (FDR 0.01) out of 26,836 tested across the 10 tissues distributed across 34 independent loci (**Supplementary Fig. 21b, Supplementary Tables 5**). The reproducibility of the observed clustering structure and identified group specific genes was high based on spearman correlation (> 95) of groupwise gene-expression profiles and distribution of patients across groups (**Supplementary Fig. 21f-g**).

Similarly, we identified 256 (+128 WikiPathway / CMC gene-set) unique pathways out of 6,120 (+3,571 WikiPathway / CMC gene-set) with differential liability profiles (**Fig. 3b, 4a, Supplementary Fig. 23, Supplementary Data 7**). Given the absence of large deeply phenotyped cohorts for SCZ, we turned to a different strategy for the identification of groupwise differences in endophenotypes and interpretation of pathway level liability profiles. Prior to application of SCZ, we carefully benchmarked this approach in CAD (see Methods, **Supplementary Fig. 24**).

These analyses resulted in 72 endophenotypes (out of 1000, see Materials & Methods) that differ in at least one SCZ patient group (**Fig. 3b, Supplementary Tables 6**). Jointly, these results support the notion of fundamental differences in endophenotype profiles across SCZ patient strata that were linked to distinct liabilities across multiple biological processes: Group 1 showed decreased estimated white blood cell counts and increased neutrophil to lymphocyte ratio (NLR) as well as lower estimated CRP levels, suggesting a lower inflammatory state. In line with these findings, group 1 showed decreased liability towards immune related pathways such as T-cell differentiation, Cytokines and Inflammatory Response and Complement Activation (**Fig. 4a,b**). Moreover, group 1 exhibited a decreased liability towards the development of depression (**Fig. 4a**, bottom) and an overall better estimated cognitive performance based on various indicators (**Fig. 4b, Supplementary Fig. 25**). This was accompanied by a lower predicted expression of presynaptic genes, genes related to synaptic density, and mitochondria as well as an increase in genes related to oxidative damage (**Fig. 4a**). Interestingly, group 1 also showed an increase in fractional anisotropy in the corpus callosum based on MRI^36^ and an opposite effect in group 3 was observed (**Fig. 4b)**. Previously, the latter was reported to be decreased in SCZ patients compared to controls. In summary, we conclude that group 1 represents a population of SCZ patients with a less severe disease status. However, group 1 was characterized by a significantly higher predisposition to metabolic syndrome (MetS) with higher levels of the 5 risk factors used to define MetS, including elevated cholesterol and triglyceride levels (**Fig. 4c**). It is well known that overall SCZ patients have an increased risk for MetS^37^, but unclear whether this comorbidity would result from a distinct genetic risk factor profile. In line with this observation, group 1 showed altered genetic liability with respect to insulin signalling related genes (**Fig. 4a**). These results supplement previous clinical observation on the existence of a MetS subgroup with a genetic and biological basis.

**Fig. 4:**
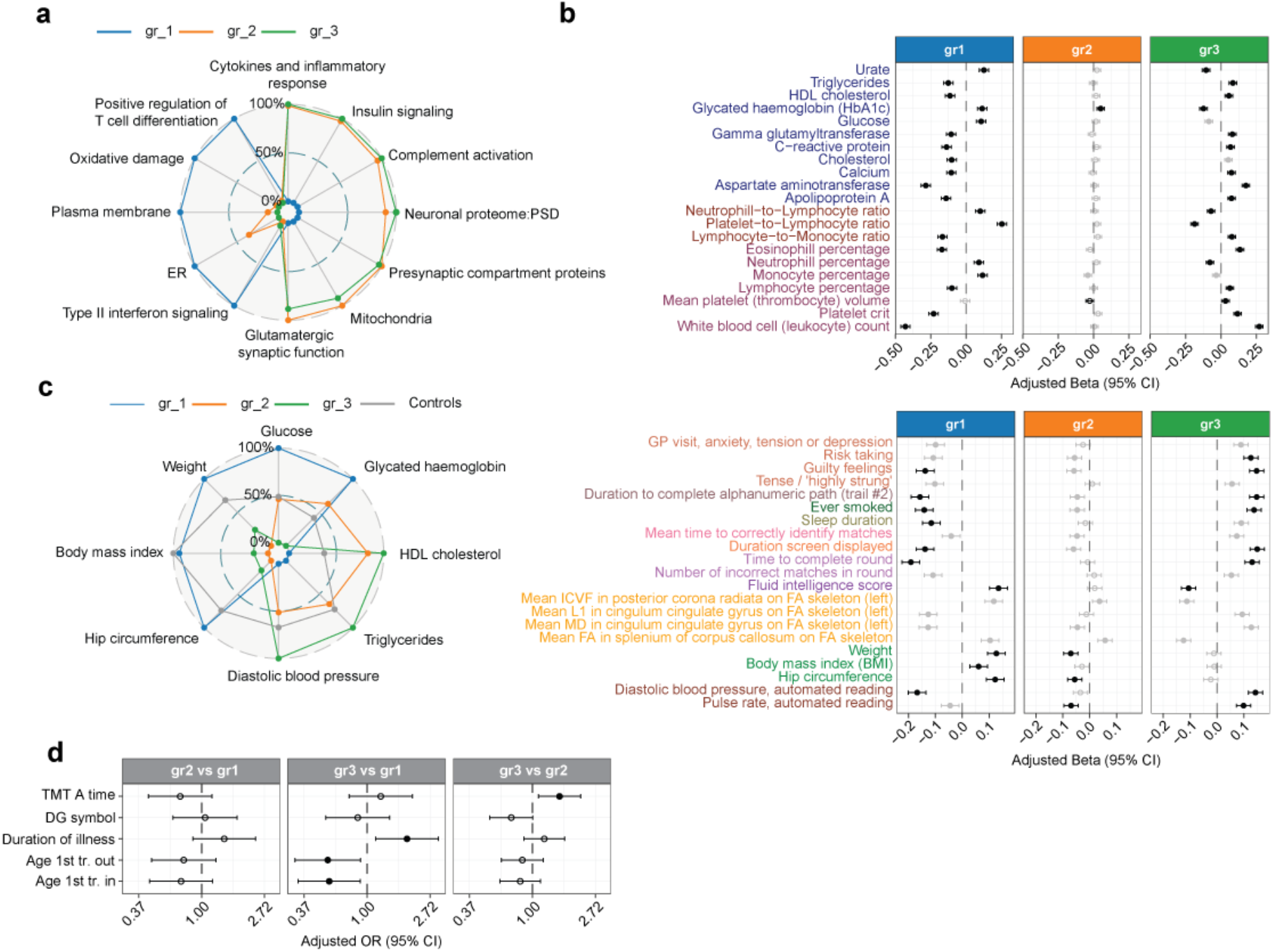
SCZ patients genetically driven stratification from imputed gene expression in DLPC. **a**. Mean value of selected group-specific pathways (Reactome and GO, WikiPathways and CMC Gene Set) in each group rescaled to 0-100 range. **b**. Forest plot of a selection of gene risk-scores (gene-RS) endophenotypes with FDR ≤ 0.05 and cluster reliable measure (CRM) > 500 in at least one group. X-axis shows the regression coefficient with 95% CI for the grouping variable (β_GLM_). Full dot indicates that β_GLM_ is significant after BH correction, performed separately for each group across all the endophenotype. Black dot indicates that the group-specific association is also reliable based on CRM threshold of 610. The top panel shows results in terms of blood count and blood biochemistry UKBB phenotype classes. **c**. Group-specific spider plot related to Metabolic Syndrome phenotypes. Mean value of group-specific gene-RS endophenotype related to metabolic syndrome across all cohorts. Grey chart refers to all control combined in PGC cohorts. In each endophenotype, SCZ groups plus controls group are rescaled to 0-100 range. **d**. Forest plot testing measured clinical differences across projected groups in SCZ PsyCourse cohort. The test based on GLM is performed for each pair of groups (label on top). Dot indicates significance (p ≤ 0.05). tr. out/in – treatment outpatient/inpatient.

Conversely, group 3 captures a patient group of severe SCZ, with increased inflammatory and substantially reduced cognitive performance parameters (**Fig. 4a, Supplementary Fig. 25**). These differences on the endophenotype level are reflected in a reduced expression of proteasome degradation, interferon II signalling, plasma membrane, and endoplasmatic reticulum (ER) related genes (**Fig. 4b**). In contrast, genes related to cytokine and inflammatory response, complement activation as well as related to the presynaptic compartment and postsynaptic density were upregulated (**Fig. 3b, 4a**).

Jointly, these observations suggest the existence of at least two SCZ patient populations with distinct endophenotypes and consistent biological liability profiles as well as another group that represents an intermediate between the two.

In order to validate these observations, we turned to a smaller but clinically phenotyped longitudinal cohort of SCZ patients (PsyCourse)^38^. Following the prediction of gene expression levels and projection of n=556 SCZ patients onto the PGC-SCZ patient derived clustering structure, we reproduced again 3 groups of patients with similar proportions (**Supplementary Fig. 26**). Comparison of differences in n=30 clinical phenotypes revealed a significantly longer duration of illness in group 3 compared to group 1 (p= 0.01) and a lower age of first treatment (p=0.02) indicative of an increased disease severity in group 3 (**Fig. 4d**). In addition, group 3 showed a significant reduction in one of the cognitive performance indicators (tmt_A_rt) compared to group 2 anda borderline significant a reduction in of an independent symbol test assessing cognitive performance (dg_sym, **Fig. 4d**, p=0.055).

In order to probe the possibility to operationalize insights the CASTOM-iGEx stratification strategy in the context of stratified medicine, we performed a drug repositioning analysis from pathways signatures^39^ (Methods). Based on the reversion signature principle^40^, we identified drugs that perturb cluster-specific up- or -down regulated pathways with an opposite effect on the transcriptionally derived pathway response. We found that treatments reverting the pathway signature for group 2 and group 3 predominantly were members of to the nervous system class based on Anatomical Therapeutic Chemical (ATC) classification system (**Supplementary Fig. 27a**). Conversely, none of the drugs associated to group 1 were related to nervous system treatments but instead were divided among other 4 ATC classes, in particular the cardiovascular system (**Supplementary Fig. 27a**). Jointly, these independent results further support the clinical relevance of the observed patient stratification structure.

In conclusion, CASTom-iGEx patient stratification methodology detected distinct patient groups exhibiting different genetic liabilities that translate into divergent clinical parameters across different complex diseases.

## Discussion

Here, we investigated how heterogeneity in polygenic risk factor distribution can contribute to heterogeneity in clinical parameters, severity, and treatment response across patients suffering from complex diseases.

We start to resolve this central problem on the road to stratification medicine by developing a multilayered machine learning approach that relies on the stepwise aggregation of genetic signal onto biological relevant entities (genes and pathways) on a per individuum level. We introduce the concept pathway level association studies and highlight the added value of this strategy in terms of identifying biologically directly interpretable, tissue specific associations and increased detection power. We show that a substantial number of pathway associations result from small, additive effects missed otherwise.

We show that aggregation of genetic liability through tissue specific gene expression enables the identification of distinct patient subgroups. This approach enables the unsupervised stratification of patients that exhibit distinct genetic liabilities across biological process into subgroups with diverse endophenotypic and clinical profiles. Importantly, this level of biologically and clinically relevant multivariate stratification was not achieved by traditional PRS analysis, highlighting the added value of the CASTom-iGEx approach.

Our results show that the effects of common disease associated genetic variants converge onto distinct cell type specific genes and molecular pathways within subgroups of patients. Most importantly, we extensively evaluate well known confounders in genetic stratification analyses and show that our discovered patient grouping is not compromised by the former.

We show the general feasibility of unbiased patient stratification by applying the CASTOM-iGEx pipeline to two fundamentally distinct complex diseases. Moreover, we demonstrate the added value of the biological concept informed genetic patient stratification through detailed clinical and endophenotypic characterization of the discovered patient strata.

In particular, we identify 5 groups of CAD patients with fundamentally distinct risk and disease relevant endophenotype profiles. This includes a healthier population, a population with reduced levels of blood-circulating LDL, and a decreased frequency of hyperlipidemia concomitant with higher predicted activities of vesicle mediated transport.

Finally, we identify a patient group that exhibits a stronger role of inflammatory processes, adding a genetic foundation to the debated role of inflammation in CAD. Similarly, stratification analysis of schizophrenic individuals revealed substantial heterogeneity in risk factor distribution related to pathomechanisms that have long been implicated to play a key role in SCZ. These include genes and pathways related to inhibitory and excitatory neurotransmission affecting excitatory/inhibitory balance in cortical microcircuits, glutamate metabolism as well as the so far barely recognized involvement of lipoprotein metabolism. These analyses also uncovered the existence of a SCZ patient group with substantially increased genetic loadings for better cognitive performance and lower liability for inflammatory processes, while at the same time showing a higher genetic risk profile for metabolic syndrome

These results showcase the general utility of the CASTom-iGEx approach in the deconstruction of phenotypic and clinical heterogeneity across patient populations and eventually facilitate precision medicine approaches. While the current results represent an important next step along this road, several key challenges remain.

First, the CASTom-iGEx strategy was only applied in the context of individuals with Caucasian ancestry. Application of European ancestry trained models to individuals with Indian ancestries showed overall poor performance and replication of results (**Supplementary Fig. 28**), consistent with previous observations^41,42^ and requires adaption to a trans-ancestry setting. However, the latter likely requires not only tailoring of statistical models but also generation of new cohorts: While most GWAS hits replicate across populations, there exists substantial variability in effect sizes^41^ and direction of effects for subthreshold associations, concomitant with limited transferability of PRS across populations^42^. The latter observations are consistent with considerations within the omnigenic model of complex traits, suggesting lower trans-population conservation of genes peripherally related to the phenotype of interest.

As consequence, the generalizability of gene risk score (GRS) based models such as CASTom-iGEx to a trans-ancestry setting through adapted statistical methods^43^ remains unclear and requires the careful calibration using ancestry specific and trans-ancestry GRS models. However, this endeavor requires in any case more diverse cohorts of matching genotype and gene expression data of disease relevant tissues of sufficient size across distinct populations^44^. Second, the approach presented here constitutes only one step forward towards the use biological and translational operationalization of common variant distribution, as it can only be truly effective when combined with other tools and data modalities. Environmental and lifestyle factors dramatically influence disease risk and disease course. Thus, it will be one of the critical next steps to integrate genetic based insights such as those provided by CASTom-iGEx with deep patient phenotyping information in the context of an unsupervised multi-modal patient clustering framework. In particular, integrating the present approach with multi-omic, imaging, clinical and exposome derived data modalities using e.g., network fusion methods represent promising avenues to increase the predictive power of patient stratification, specifically towards the prediction of treatment response.

## Data Availability

The software pipeline is based on R and is available at https://gitlab.mpcdf.mpg.de/luciat/castom-igex. The trained tissue specific PriLer models on GTEx v6p and CMC release 1 reference panels are available at https://doi.org/10.6084/m9.figshare.22347574.v2. TWAS and PALAS summary statistics for CAD and SCZ can be found at https://doi.org/10.6084/m9.figshare.22495561.v1.

## Acknowledgments

We thank all members of the Ziller, Gagneur and Schunkert labs for their support and critical feedback. We also thank Bernhard Baune and Monika Stoll for providing critical feedback on the manuscript.

This research has been conducted using the UK Biobank Resource under application numbers 34217 and 25214. We thank all participants, researchers, and support staff who make the study possible. Bona fide researchers can apply to use the UK Biobank data set by registering and applying at http://ukbiobank.ac.uk/ register-apply/.

The Genotype-Tissue Expression (GTEx) Project was supported by the Common Fund of the Office of the Director of the National Institutes of Health, and by NCI, NHGRI, NHLBI, NIDA, NIMH, and NINDS. The data used for the analyses described in this manuscript were obtained from: dbGaP accession number phs000424.v7.p2 on 11/28/2018.

This study used data from the CommonMind consortium provided through NIMH. Data for this publication were obtained from NIMH Repository & Genomics Resource, a centralized national biorepository for genetic studies of psychiatric disorders. Data were generated as part of the CommonMind Consortium supported by funding from Takeda Pharmaceuticals Company Limited, F. Hoffman-La Roche Ltd and NIH grants R01MH085542, R01MH093725, P50MH066392, P50MH080405, R01MH097276, RO1-MH-075916, P50M096891, P50MH084053S1, R37MH057881, AG02219, AG05138, MH06692, R01MH110921, R01MH109677, R01MH109897, U01MH103392, and contract HHSN271201300031C through IRP NIMH. Brain tissue for the study was obtained from the following brain bank collections: the Mount Sinai NIH Brain and Tissue Repository, the University of Pennsylvania Alzheimer’s Disease Core Center, the University of Pittsburgh NeuroBioBank and Brain and Tissue Repositories, and the NIMH Human Brain Collection Core. CMC Leadership: Panos Roussos, Joseph Buxbaum, Andrew Chess, Schahram Akbarian, Vahram Haroutunian (Icahn School of Medicine at Mount Sinai), Bernie Devlin, David Lewis (University of Pittsburgh), Raquel Gur, Chang-Gyu Hahn (University of Pennsylvania), Enrico Domenici (University of Trento), Mette A. Peters, Solveig Sieberts (Sage Bionetworks), Thomas Lehner, Stefano Marenco, Barbara K. Lipska (NIMH). The full list of PGC Schizophrenia working group members including affiliations can be found in Table S7.

## Author contributions

Conceptualization: MZ, LT; Methodology: LT, MZ; Investigation: LT, SM, LJT, TFMA, MS, AT, BMM, HS; Visualization: LT, Funding acquisition: MZ, JG, HS; Critical Resources: MB, PF, TG, HV, ST, UH, PGC, Supervision: MZ, Writing: LT, MZ

## Data availability

The UKBB data are privacy protected and access can be requested through the UKBB data portal. The GTEx data are available through dbGAP accession number phs000424.v7.p2. The PGC data are privacy protected and can be accessed through a secondary analysis proposal sponsored by a PGC-SCZ working group PI member that needs to be approved by the working group. The German cohorts of CARDIoGRAM consortium is privacy protected and can only be accessed through collaboration with PIs of the consortium, e.g. HS. The PsyCourse Study data are privacy protected but can be accessed by submitting a research proposal (see http://www.psycourse.de/openscience-en.html). The genotype and gene expression data from the CommonMind consortium is privacy protected and can be accessed via the CommonMind knowledge portal: http://dx.doi.org/10.7303/syn2759792. The SHIP-Trend study genotype data is privacy protected and can be accessed through the study PIs: https://www.maelstrom-research.org/study/ship. The PsyCourse data is privacy protected and can be accessed via an analysis request proposal through the study website: http://psycourse.de/openscience-de.html.

