## Supplementary Figures for "Distinct genetic liability profiles define clinically relevant patient strata across common diseases"

**This PDF file includes:**

Supplementary Fig. 1 to 28
Supplementary Tables 1 to 7
Captions for Supplementary Data 1 to 7

**Other Supplementary Materials for this manuscript include the following:**

Supplementary Data 1 to 7
Prior features construction in tissue-specific models
Significant genes associated with Coronary Artery Disease
Significant pathways associated with Coronary Artery Disease
Group-specific Pathways for CAD cases clustering in Liver
Significant genes associated with Schizophrenia
Significant pathways associated with Schizophrenia
Group-specific Pathways for SCZ cases clustering in Dorsolateral prefrontal cortex

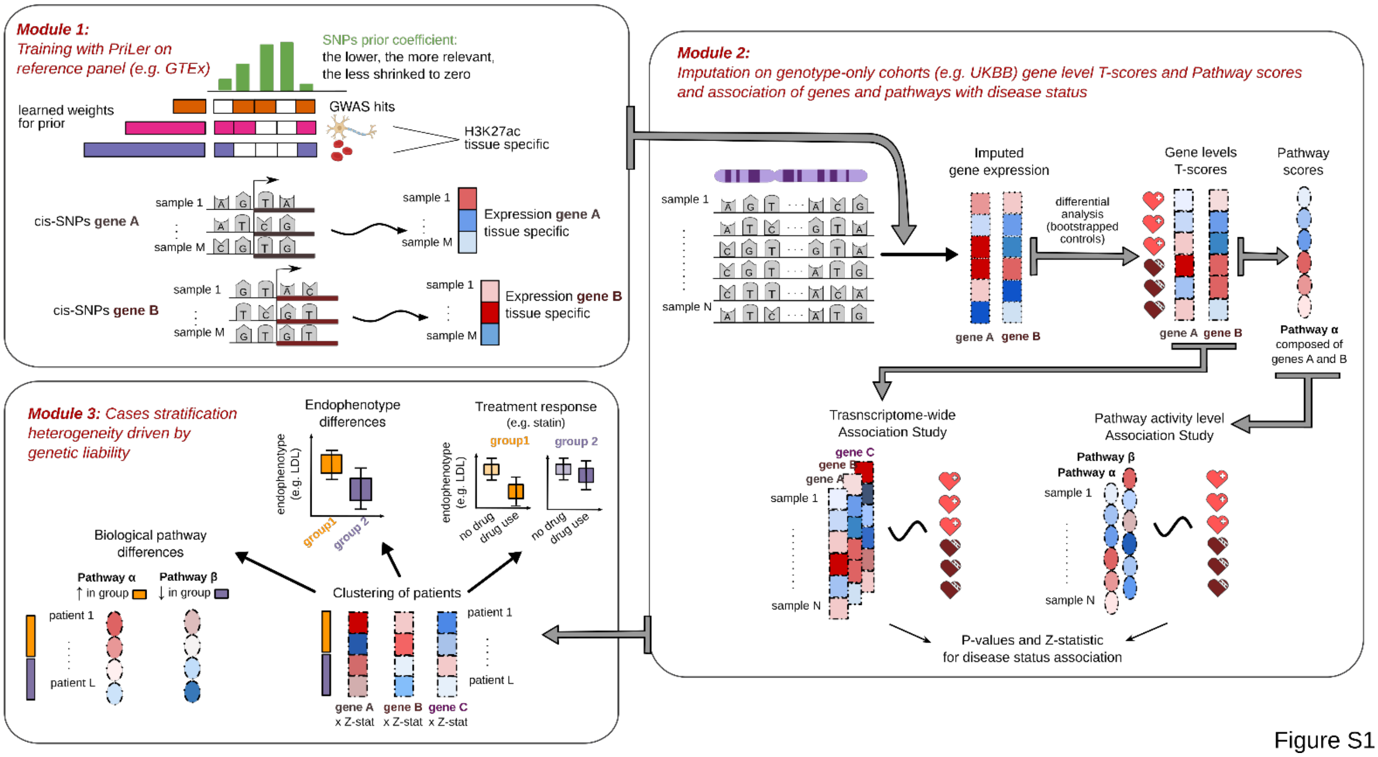


Figure S1

**Supplementary Fig. 1. Workflow CASTom-iGEx pipeline**.

CASTom-iGEx is composed of 3 modules: 1) Prior Learned elastic-net regression (PriLer) to train gene expression prediction models on reference panels integrating SNPs prior information, 2) application to genotype-only datasets, conversion to gene levels T-score and pathway levels scores and identification of differentially active genes and pathways between affected and unaffected individuals, 3) patient stratification based on gene levels T-score to characterize biological pathways and endophenotypic differences.

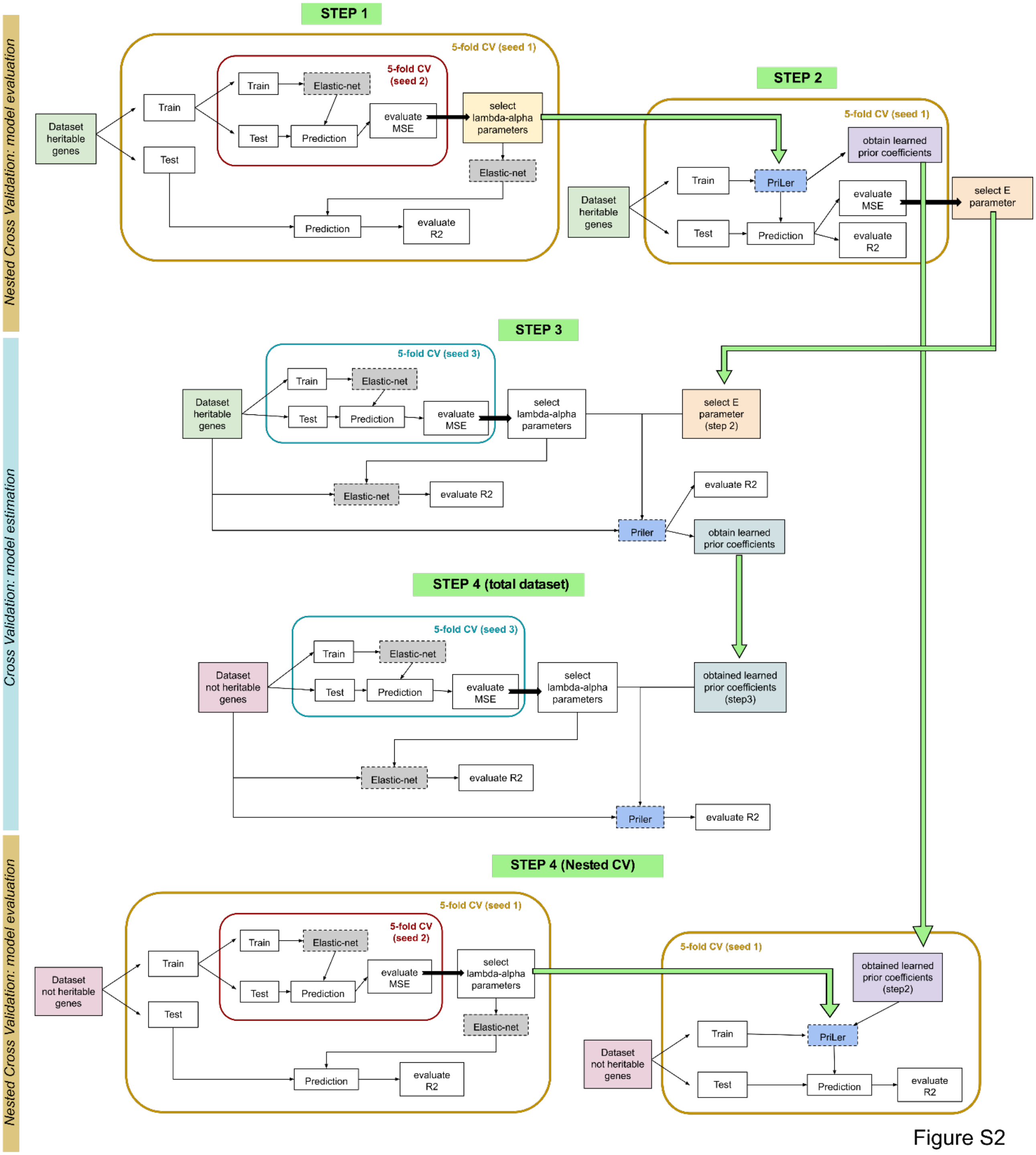
Nested Cross Validation: model evaluation
STEP 1
5-fold CV (seed 1)
5-fold CV (seed 2)
Dataset heritable genes
Train
Test
Elastic-net
Prediction
evaluate MSE
select lambda-alpha parameters
evaluate R2
STEP 2
obtain learned prior coefficients
PriLer
select E parameter
Cross Validation: model estimation
STEP 3
5-fold CV (seed 3)
select E parameter (step 2)
Priler
STEP 4 (total dataset)
Dataset not heritable genes
obtained learned prior coefficients (step3)
STEP 4 (Nested CV)
obtained learned prior coefficients (step2)

Figure S2

**Supplementary Fig. 2. PriLer steps to model gene expression integrating prior variant knowledge.**

The first step considers only heritable genes and builds an elastic-net regression model for each gene in a nested cross validation setting without prior information. The second step uses the optimal gene specific α-λ parameters combination found in step 1 to build a PriLer model in the same nested cross-validation frame in order to find the optimal E parameter that controls the magnitude of the prior weights. Step 1 and 2 are also used to evaluate the prediction models based on $R^2_{cv}$. The third step finds the optimal α-λ parameter on the entire set (single cross validation) from elastic-net regression and uses these α-λ pairs together with optimal E parameter from step 2 to build PriLer models on the entire dataset. The fourth step considers only genes not heritable, repeats step 3 but instead of deriving prior coefficients, it uses the ones computed from heritable genes. Finally, in order to evaluate the models based on $R^2_{cv}$ for genes not heritable, step 1 and 2 are also repeated with CV-specific prior coefficients derived in step 2.

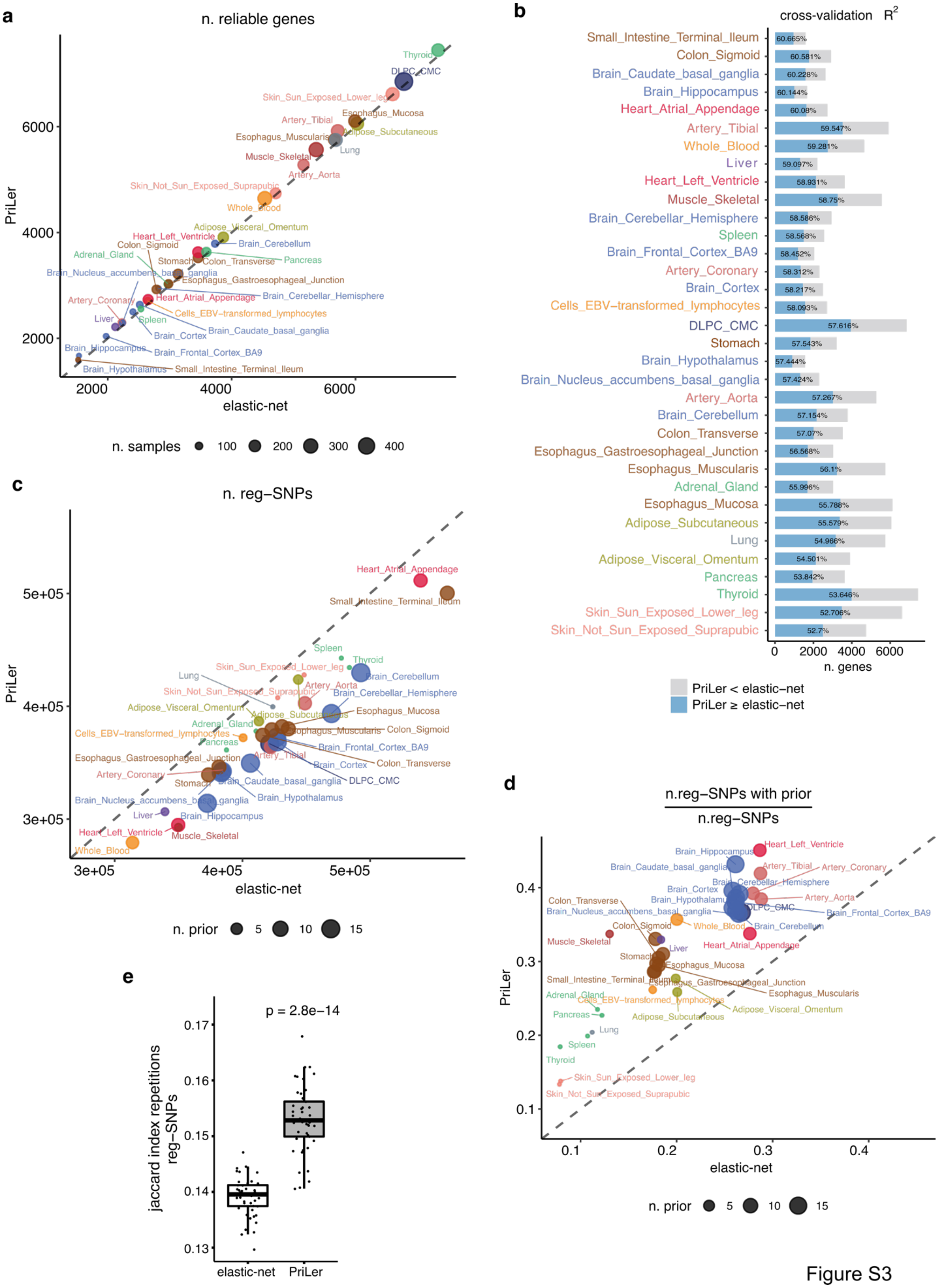
a
n. reliable genes
PriLer
elastic-net
n. samples 100 200 300 400
b
cross-validation $R^2$
n. genes
PriLer < elastic-net
PriLer ≥ elastic-net
c
n. reg−SNPs
PriLer
elastic-net
n. prior 5 10 15
d
n.reg−SNPs with prior / n.reg−SNPs
PriLer
elastic-net
n. prior 5 10 15
e
p = 2.8e−14
jaccard index repetitions reg−SNPs
elastic-net
PriLer

Figure S3

**Supplementary Fig. 3**. **Comparison PriLer and elastic-net regression imputation models.** For each tissue in GTEx/CMC panels comparison of PriLer and elastic-net regression without prior regarding **a.** number of reliable genes i.e. $R^2_{cv} > 0$ and $R^2 > 0.01$, **b.** number and percentage of genes among the reliable ones in PriLer models having better prediction performance ($R^2_{cv}$) in PriLer compared to elastic-net, **c.** number of regulatory variants i.e. variant that regulates at least 1 gene, **d**. fraction of regulatory variants that intersect with at least 1 prior feature used in PriLer tissue models, **e.** Regulatory variants robustness in whole blood measured via bootstrapping 100 individuals 10 times and computing Jaccard index in each pair of repetition for PriLer and elastic-net models (p-value from Wilcoxon-Mann-Whitney test). Boxplot elements include median as central line, $1^{st}$ and $3^{rd}$ quartiles as box limits, 1.5 interquartile ranges from $1^{st}$ and $3^{rd}$ quartiles as corresponding whiskers.

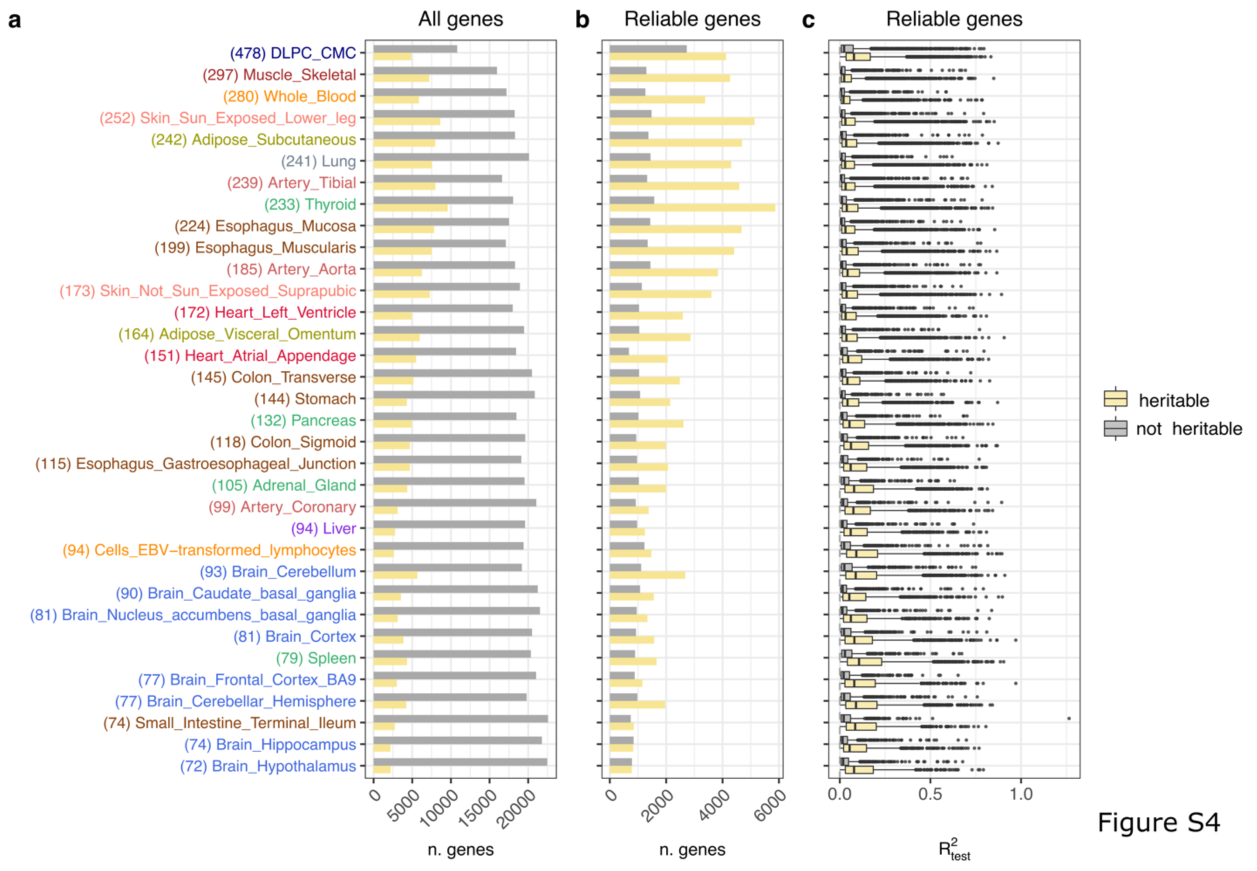


**Supplementary Fig. 4. Gene expression variability for heritable and not heritable genes explained by PriLer**

**a.** Number of total genes across the heritable and not heritable categories. **b.** Number of reliable genes across the heritable and not heritable categories. **c.** Average $R^2$ on test folds for reliable genes across all tissues, divided by heritable (yellow) and not heritable (grey) genes. The tissues are ordered on the y-axis according to their sample size. Boxplot elements include median as central line, $1^{st}$ and $3^{rd}$ quartiles as box limits, 1.5 interquartile ranges from $1^{st}$ and $3^{rd}$ quartiles as corresponding whiskers.

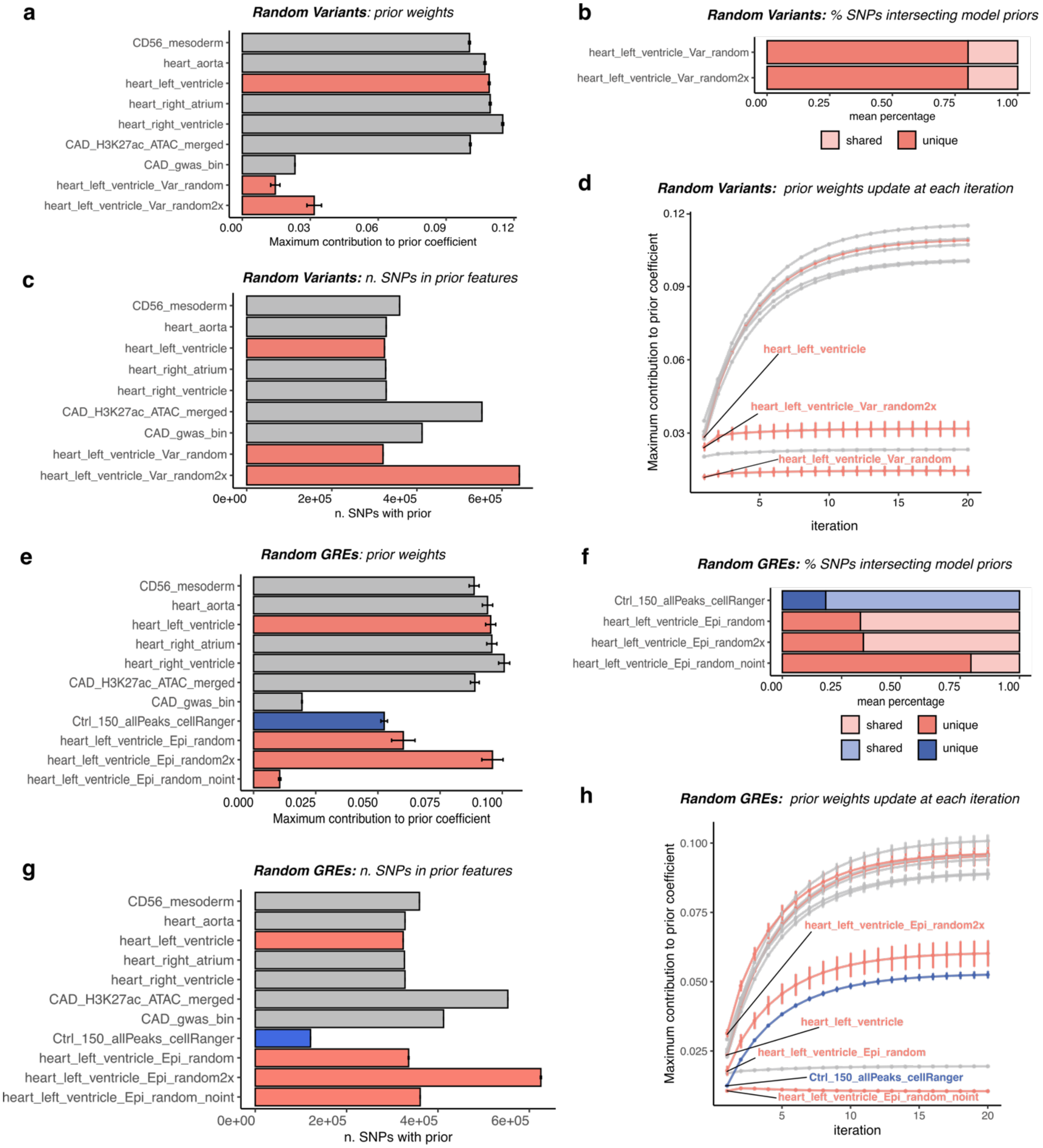


Figure S5

**Supplementary Fig. 5. Simulation of prior features to test weights their relevance in Artery Coronary tissue**.

In the first scenario (*Random Variants*), 2 prior features were randomly generated 50 times *heart_left_ventricle_Var_random* and *heart_left_ventricle_Var_random2x* using the same size/ twice the number of prior features as in *heart_left_ventricle* (pink). In the second scenario (*Random GREs*), open chromatin regions were randomly selected (i.e. gene regulatory elements GREs) from ChIP-Seq H3k27ac data 50 times using the same or twice feature set size as *heart_left_ventricle* and intersected the latter with variants location to create *heart_left_ventricle_Epi_random* and *heart_left_ventricle_Epi_random2x priors* (pink). This included also a prior feature related to brain tissue *Ctrl_150_allPeaks_cellRanger* (blue) as well as random prior *heart_left_ventricle_Epi_random_noint* sampling among GREs excluding the ones in the baseline prior features and in the same size as GREs for *heart_left_ventricle*. In both scenarios, we created PriLer models including other 6 fixed prior features for Artery Coronary. **a**, **e.** mean ± SD prior weights for each prior feature, **b**,. **f.** mean percentage of variants in the random priors features that are in common with fixed features used in the model, **c.**, **g.** mean ± SD number of variants associated to each prior features, **d.**, **h.** mean ± SD update of prior weights at each iterative step.

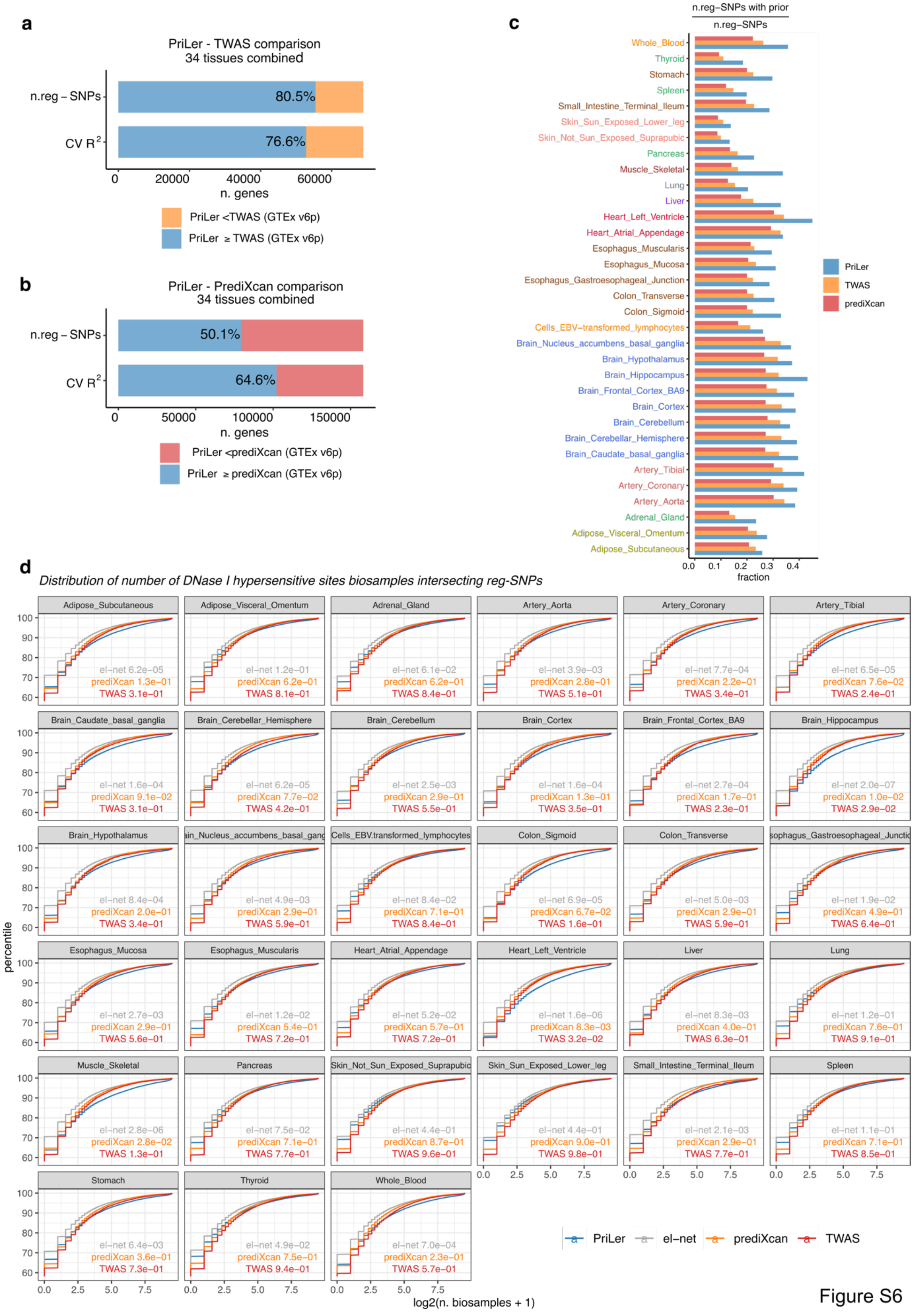
a
PriLer - TWAS comparison
34 tissues combined
n.reg – SNPs
CV R2
80.5%
76.6%
0
20000
40000
60000
n. genes
PriLer <TWAS (GTEx v6p)
PriLer ≥ TWAS (GTEx v6p)
b
PriLer - PrediXcan comparison
34 tissues combined
n.reg – SNPs
CV R2
50.1%
64.6%
0
50000
100000
150000
n. genes
PriLer <prediXcan (GTEx v6p)
PriLer ≥ prediXcan (GTEx v6p)
c
n.reg–SNPs with prior
n.reg–SNPs
fraction
PriLer
TWAS
prediXcan
d
Distribution of number of DNase I hypersensitive sites biosamples intersecting reg-SNPs
percentile
log2(n. biosamples + 1)
PriLer
el–net
prediXcan
TWAS

Figure S6

**Supplementary Fig. 6. Comparison PriLer with TWAS and prediXcan imputation models.** Comparison of PriLer to prediXcan and TWAS build on GTEx v6p 33 (tissues) and CMC datasets. Number and percentage of genes with better performance ($R^2_{cv}$) and higher number of regulatory variants in **a.** PriLer compared to TWAS and in **b.** PriLer compared to prediXcan. **c.** Fraction of regulatory variants intersecting tissue specific prior information used in PriLer model across the three methods. **d.** External validation of regulatory variants enrichment for PriLer in biologically meaningful regions using DNase I hypersensitive sites (DHSs): cumulative distribution of regulatory variants overlapping with a certain number of DHS biosamples, differences in distributions between PriLer and elasic-net, TWAS or prediXcan tested with Kolmogorov-Smirnoff test.

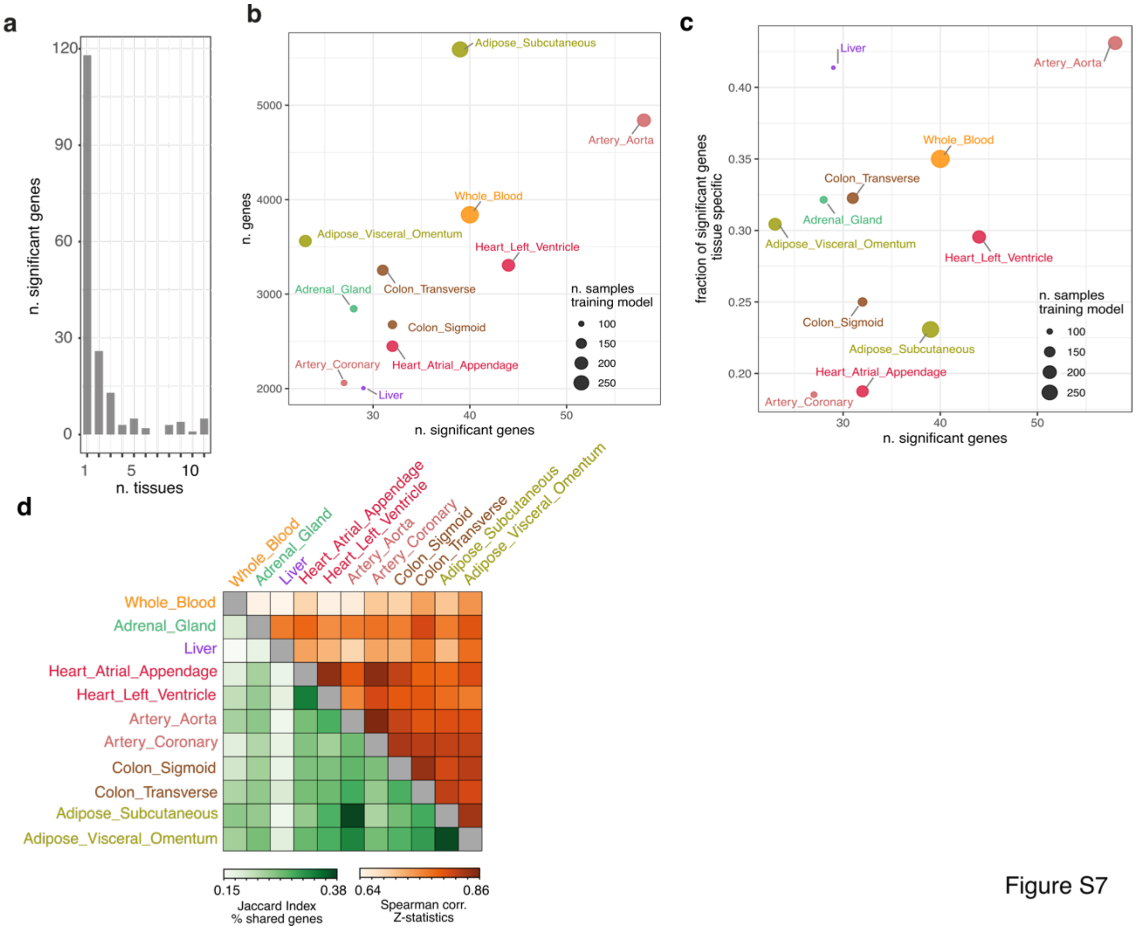


**Supplementary Fig. 7. TWAS summary results for CAD.**

**a.** Number of FDR 0.05 significant genes per number of tissues in which they are detected for each CAD related tissue **b.** number of detected reliable genes compared to the number of significant genes, **c.** fraction of significant genes uniquely detected in a tissue compared to number of significant genes. **d.** Lower-triangular (green): percentage of imputed genes that are in common between 2 tissues (Jaccard index), upper-triangular (orange): Spearman correlation of CAD Z-statistics among shared genes.

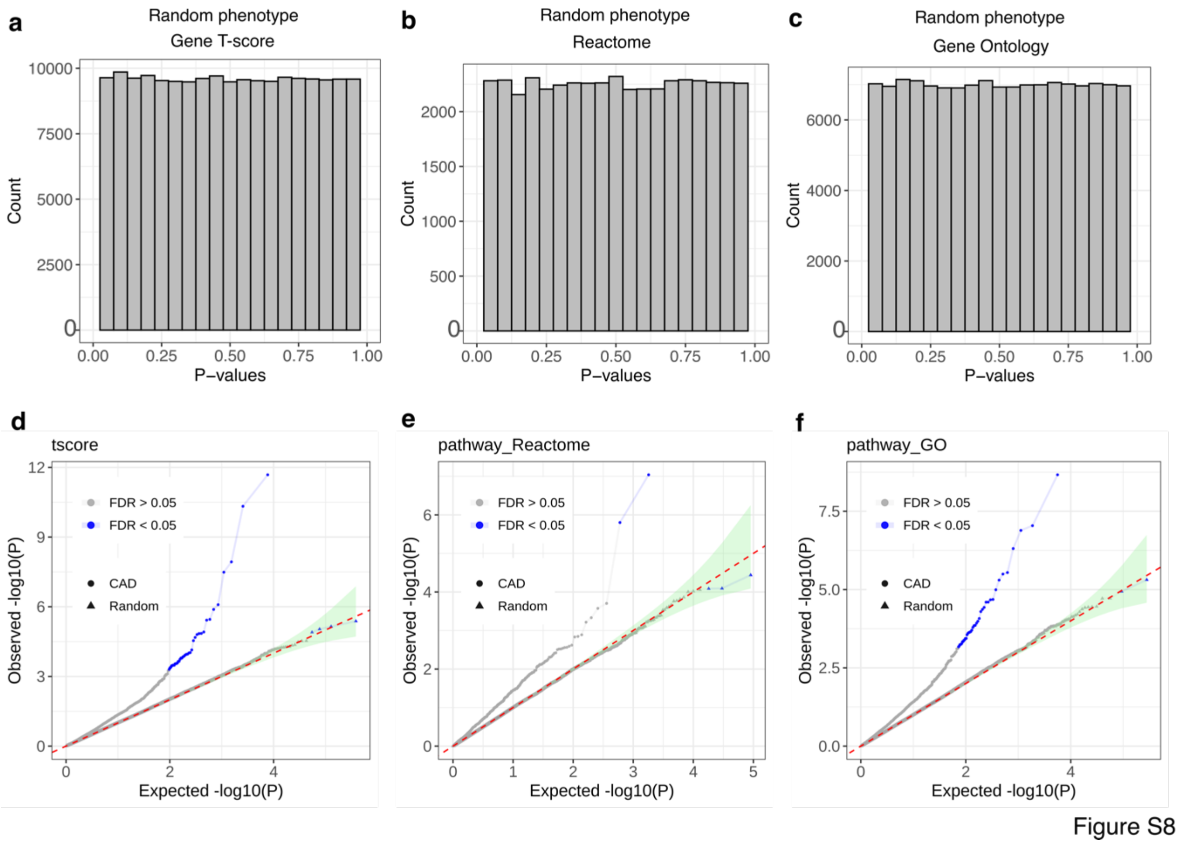


**Supplementary Fig. 8: P-value calibration for TWAS and PALAS in whole blood**

Distribution of p-values for two analyses (TWAS and PALAS) for random phenotypes in whole blood across 50 simulations, using size matched cases/control and the age/sex distributions. **a-c.** Count of p-values in specific intervals for gene T-scores, Reactome pathway-scores, and GO pathway-scores, respectively. **d-f.** Expected and observed p-value distributions for CAD (represented by dots) and random phenotypes (represented by triangles) for gene T-scores, Reactome pathway-scores, and GO pathway-scores. The diagonal line represents the expected distribution and the green shaded area shows the 95% confidence interval from a beta distribution. Blue points indicate genes that are significant at a 0.05 false discovery rate level, corrected separately for CAD and each simulation.

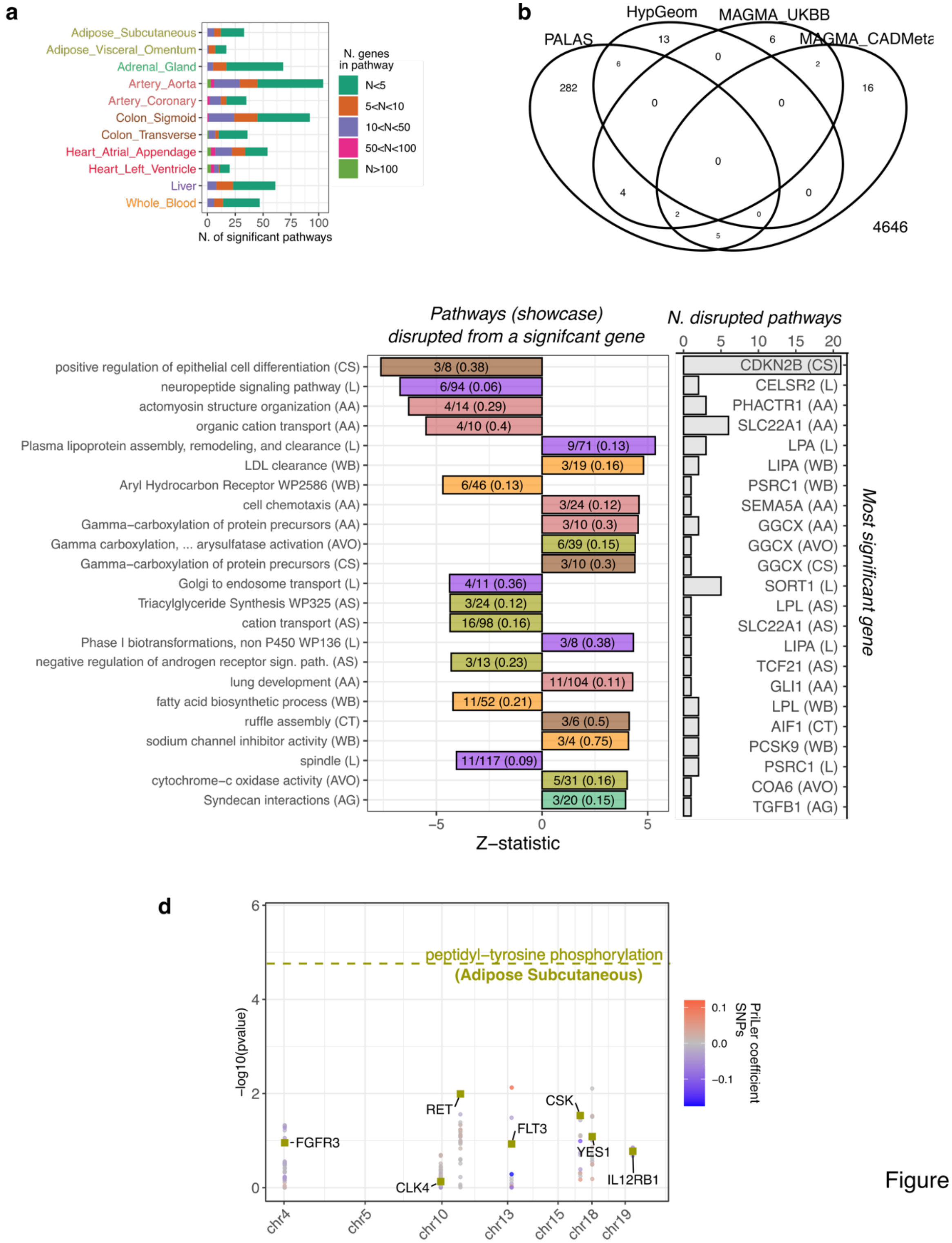


Figure S9

**Supplementary Fig. 9. PALAS summary results for CAD.**

**a.** Significant pathways linked to CAD in each tissue with tissue-specific FDR of less than or equal to 0.05. The bars represent the number of pathways and their color represents the number of genes that are reliably predicted in that tissue (based on T-score genes). **b.** Venn-diagram of PALAS significant pathways (FDR 0.05), pathways significant from hyper-geometric test (FDR 0.05) using TWAS significant genes and pathways significant from MAGMA (FDR 0.05) applied to GWAS performed on matched UK Biobank data (MAGMA_UKBB) or GWAS summary statistics from CAD GWAS in [20]. **c.** Pathways with at least one gene significantly associated with CAD based on TWAS and filtered based on the prioritization criteria (computed from more than 5 and less or equal than 200 T-score genes or more than 2 if pathway coverage is higher than 10%, originally including less than 200 genes and reaching at least 0.0001 as nominal significance). The left panel shows PALAS Z-statistic (x-axis) color coded by the tissue of origin and indicating the gene pathway coverage for only one exemplar (best coverage) per most significant gene (in the right panel). The right panel shows the number of disrupted pathways by a certain gene. Acronyms in parenthesis indicate the initials of the tissue considered (AS = Adipose Subcutaneous, AVO = Adipose Visceral Omentum, AG = Adrenal Gland, AA = Artery Aorta, AC = Artery Coronary, CS = Colon Sigmoid, CT = Colon Transverse, HAA = Heart Atrial Appendage, HLV = Heart Left Ventricle, L = Liver, WB = Whole Blood). **d.** GO peptidyl-tyrosine phosphorylation in adipose subcutaneous. The pathway significance is indicated by the dashed horizontal line, the coloured squares show genes included in that pathway and the corresponding TWAS p-value (y-axis) and the dots indicate the matched GWAS p-value of SNPs regulating those genes with colour reflecting PriLer regulatory coefficients.

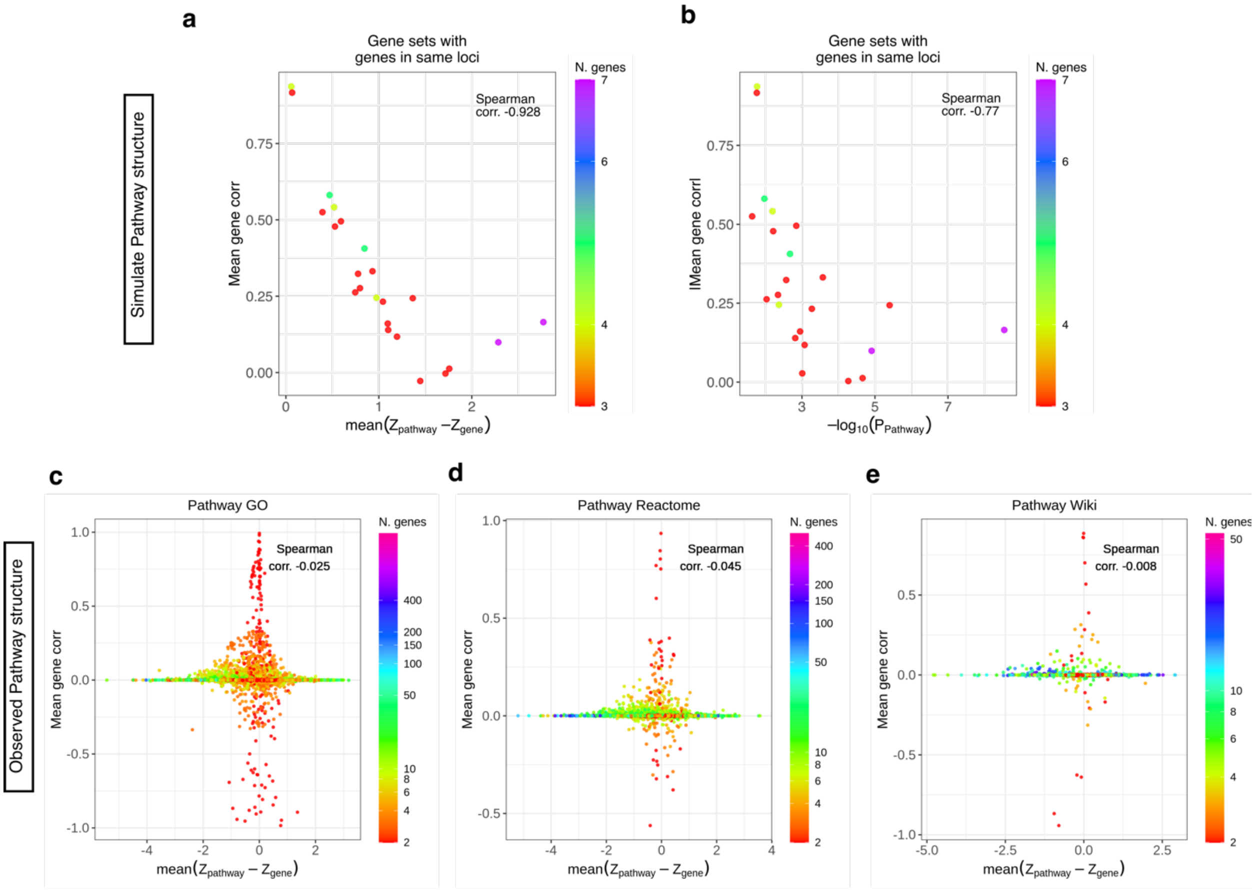


Figure S10

**Supplementary Fig. 10. Gene correlation relevance for PALAS results.**

**a-b.** This figure shows the simulation of pathway structure using gene expression data in whole blood from the same locus with a concordant effect size sign and TWAS nominal p-value of less than 0.1. The simulation covers 46 pathways in total. **a.** Each simulated pathway is a point with a color indicating the number of genes in the pathway. The X-axis represents the average difference in Z-statistic between the pathway and the included genes, while the Y-axis shows the absolute value of the mean correlation among the genes in the pathway. **b.** Similar to a., but with the X-axis showing -log 10 p-value from PALAS. **c-d.** Improvement in pathway significance related to the individual gene level p-values based on TWAS that are included in it as a function of the correlation between the genes (y-axis). Each point in the figure represents a pathway from GO **c.**, Reactome **d.**, or WikiPathways **e.**, and the color code shows the number of genes that were used to calculate the pathway-score. The X-axis displays the average difference in Z-statistics between a pathway and its corresponding genes, and the Y-axis shows the mean correlation between those genes.

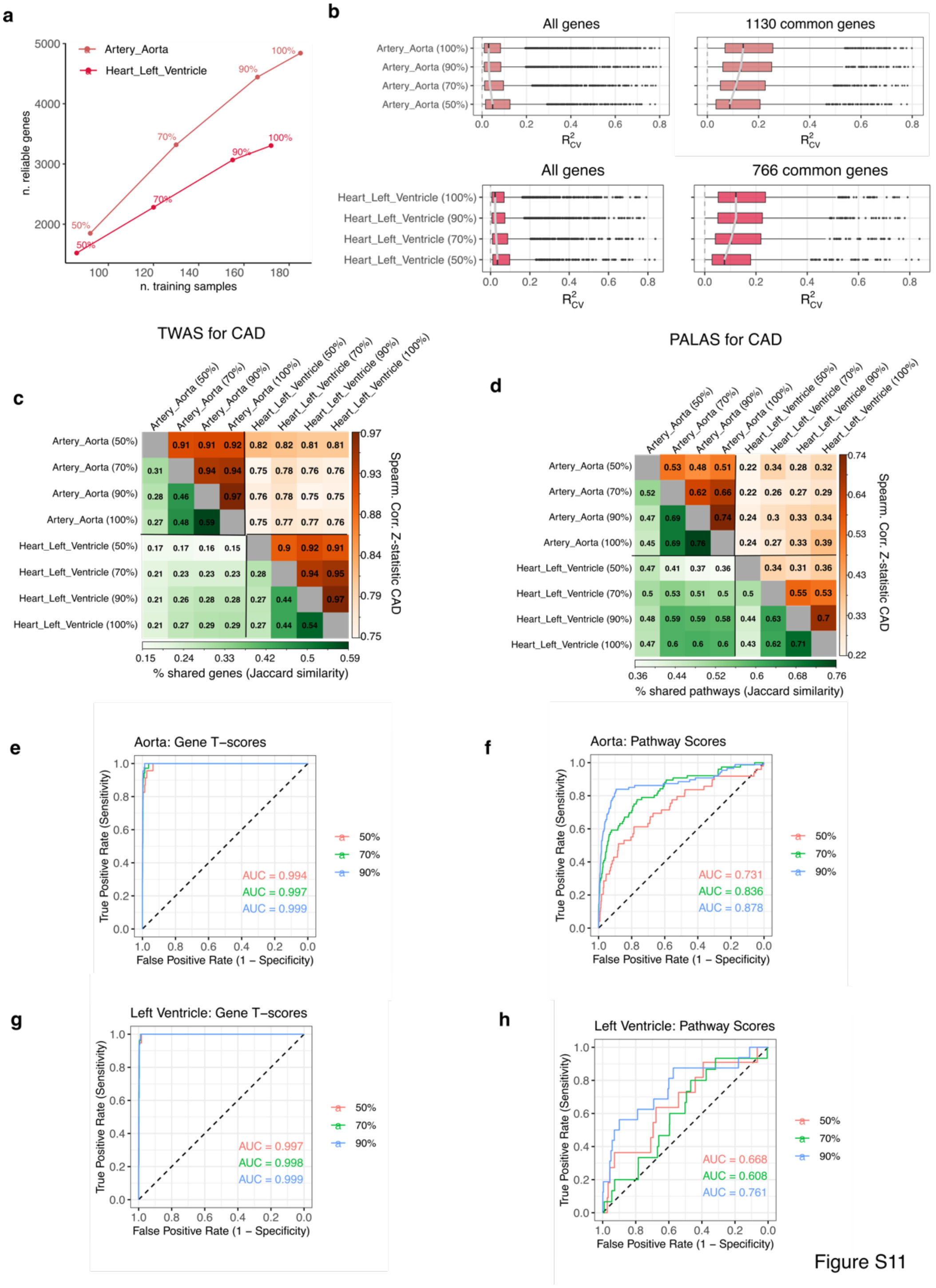


**Supplementary Fig. 11. Down-sampling of GTEx reference panel sample in artery aorta and heart left ventricle gene expression datasets**

**a.** Number of reliable genes from PriLer compared to the training sample size across the two sub-sampled tissues at 50%, 70% and 90%. **b.** PriLer model estimates in terms of average $R^2$ on test folds ($R^2_{CV}$) considering all reliable genes (left) and reliable genes in common of the tissue-specific sub-sampled models (right). The grey line connects the median distribution for the increasing model sample size. **c-d.** Heatmap with lower triangular part (green) indicating the percentage of shared genes/pathways computed via Jaccard similarity and the upper triangular part (orange) indicating the Spearman correlation of CAD Z-statistic for the genes/pathways in common. **c.** refers to TWAS results and **d.** to PALAS. **e-h.** Receiver operating characteristic (ROC) curve using significant genes/pathways (FDR 0.05) in the full sample size model (100%) as ground truth and the absolute Z-statistic as prediction values across sub-sampled percentages. For each comparison, only genes/pathways in common are considered. **e.** TWAS results in artery aorta, **f.** PALS results in artery aorta, **g.** TWAS results in heart left ventricle, **h.** PALS results in heart left ventricle. Area under the curve (AUC) is shown in each plot with color matching the sub-sampled model legend. Boxplot elements include median as central line, 1st and 3rd quartiles as box limits, 1.5 interquartile ranges from 1st and 3rd quartiles as corresponding whiskers.

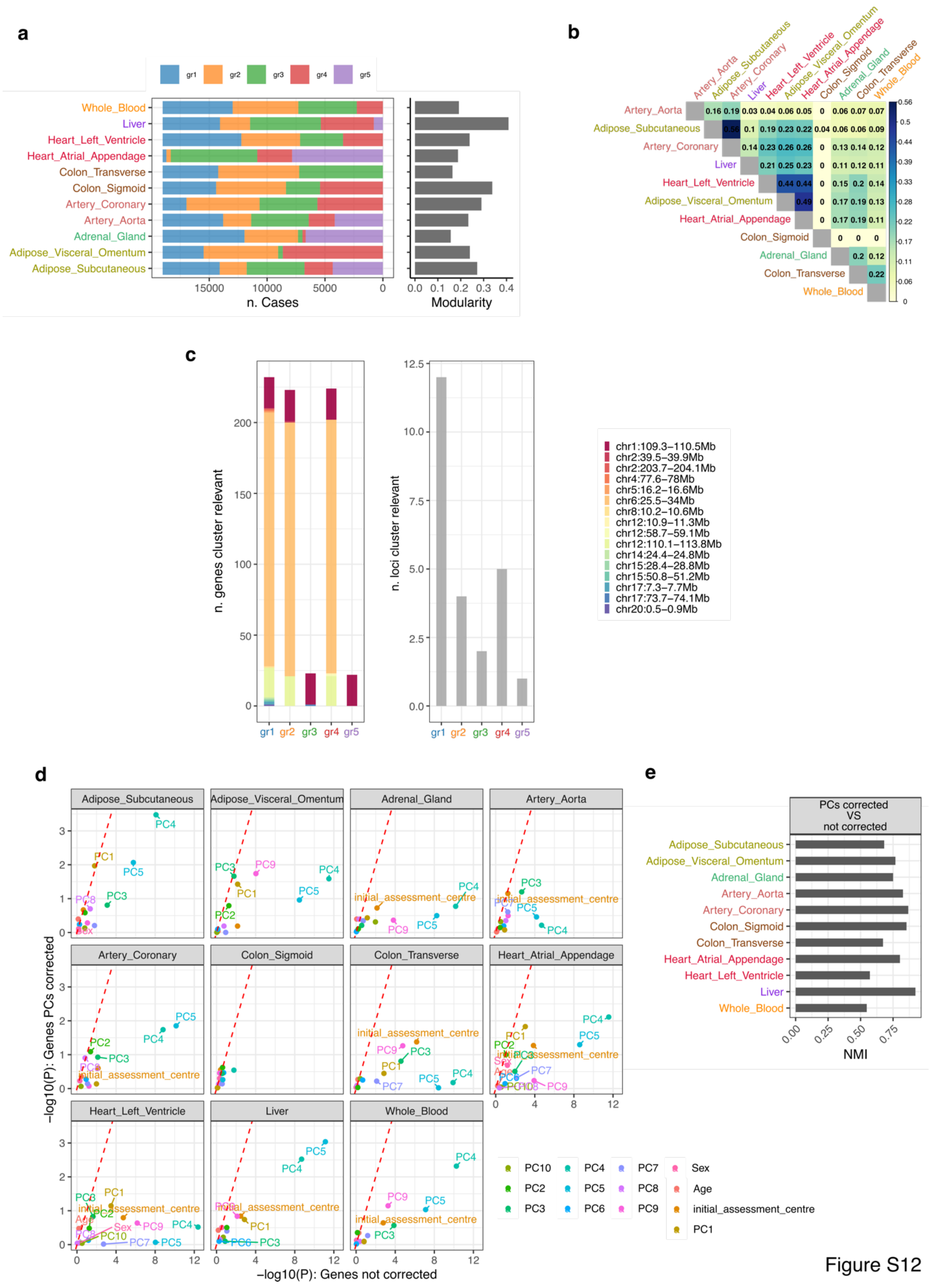

a
b
c
d
e
gr1
gr2
gr3
gr4
gr5
Whole_Blood
Liver
Heart_Left_Ventricle
Heart_Atrial_Appendage
Colon_Transverse
Colon_Sigmoid
Artery_Coronary
Artery_Aorta
Adrenal_Gland
Adipose_Visceral_Omentum
Adipose_Subcutaneous
n. Cases
Modularity
n. genes cluster relevant
n. loci cluster relevant
chr1:109.3–110.5Mb
chr2:39.5–39.9Mb
chr2:203.7–204.1Mb
chr4:77.6–78Mb
chr5:16.2–16.6Mb
chr6:25.5–34Mb
chr8:10.2–10.6Mb
chr12:10.9–11.3Mb
chr12:58.7–59.1Mb
chr12:110.1–113.8Mb
chr14:24.4–24.8Mb
chr15:28.4–28.8Mb
chr15:50.8–51.2Mb
chr17:7.3–7.7Mb
chr17:73.7–74.1Mb
chr20:0.5–0.9Mb
−log10(P): Genes PCs corrected
−log10(P): Genes not corrected
PC1
PC2
PC3
PC4
PC5
PC6
PC7
PC8
PC9
PC10
Sex
Age
initial_assessment_centre
PCs corrected
VS
not corrected
NMI


Figure S12

**Supplementary Fig. 12. Comparing clustering structure of CAD patients across tissues**.

**a.** Proportion of individuals in each tissue-specific cluster across 19,026 patients together with modularity from Louvain clustering. **b.** Normalized mutual information (NMI) for each pair of tissue-specific clustering structure. **c.** Number significant genes and loci (tissue specific FDR $\leq$ 0.01) associated with each group in liver clustering from Wilcoxon-Mann-Whithney (WMW) test of $gr_i$ versus remaining patients combing all tissues. **d.** -log10(p-value) testing clustering results with PCs from 1 to 10 and Age (Kruskal-Wallis test) and with Sex and Assessment Centre ($\chi^2$ test) before (x-axis) and after (y-axis) correction of gene T-scores for PCs. Red dashed line represents the intercept. **e.** NMI between clustering results using genes corrected for PCs or uncorrected.

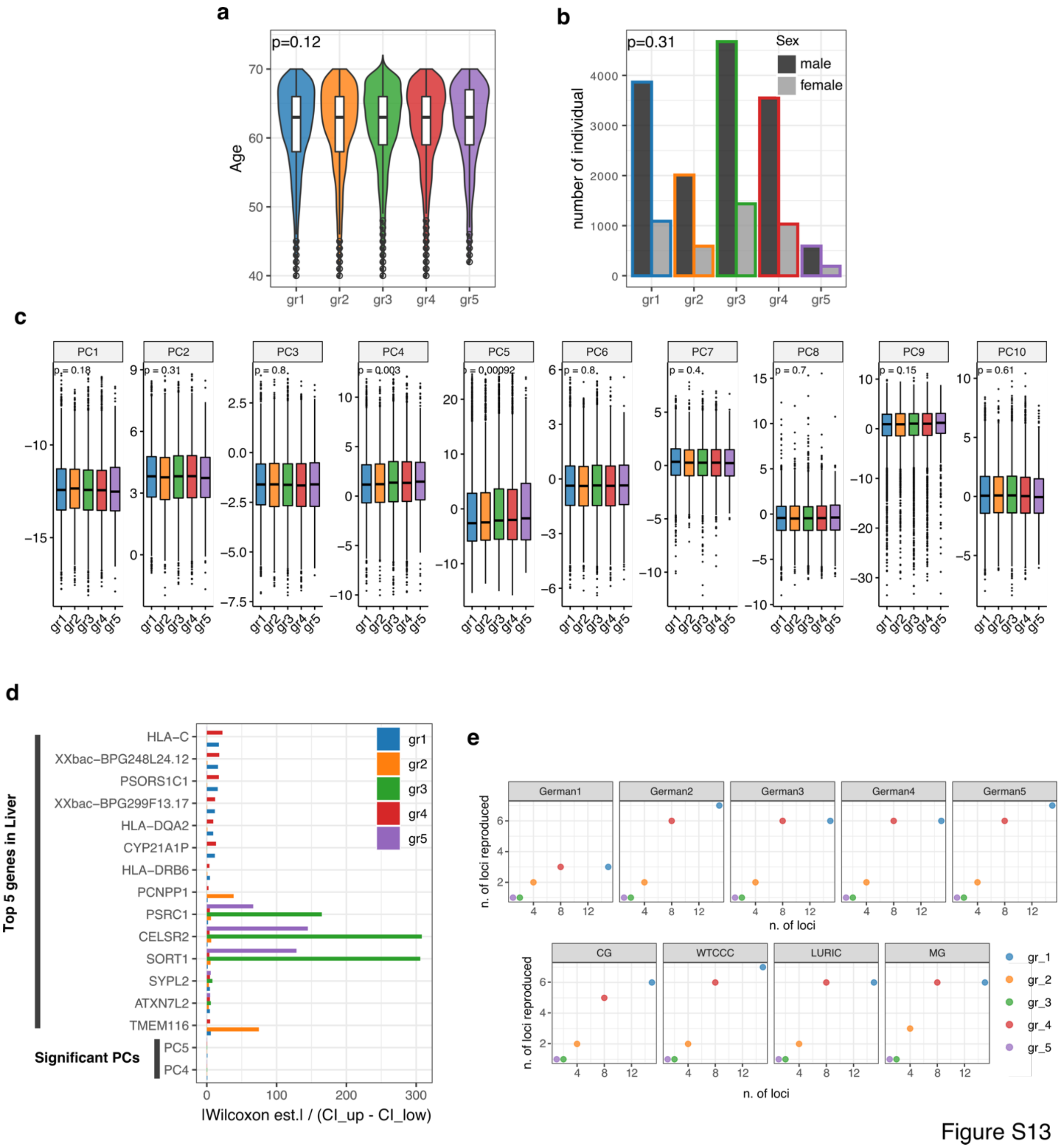


**Supplementary Fig. 13. Genes and covariates associated with clustering structure in liver.**

**a.** Age distribution for each group (p-values from Kruskal-Wallis) **b.** Sex distribution for each group (p-values from Chi-squared test). **c.** Distribution of PCs 1 to 10 for each group (p-values from Kruskal-Wallis test). **d.** X-axis shows the coefficient of variation, absolute value of WMW estimates divided by confidence interval ranges, for group-specific test. Y-axis indicates the first

5 genes per group based on WMW p-value and significant group-specific principal components (FDR ≤ 0.05). **e.** Reproducibility of group-specific loci on predicted groups in 9 external cohorts (CARDIoGRAM), the x-axis shows the number of loci across all tissues associated with each group in UKBB, the y-axis shows how many of these loci have the same sign and are significant at the nominal level of 0.05 for the strongest association of the WMW estimates in the predicted clustering structure. Boxplot elements include median as central line, $1^{st}$ and $3^{rd}$ quartiles as box limits, 1.5 interquartile ranges from $1^{st}$ and $3^{rd}$ quartiles as corresponding whiskers.

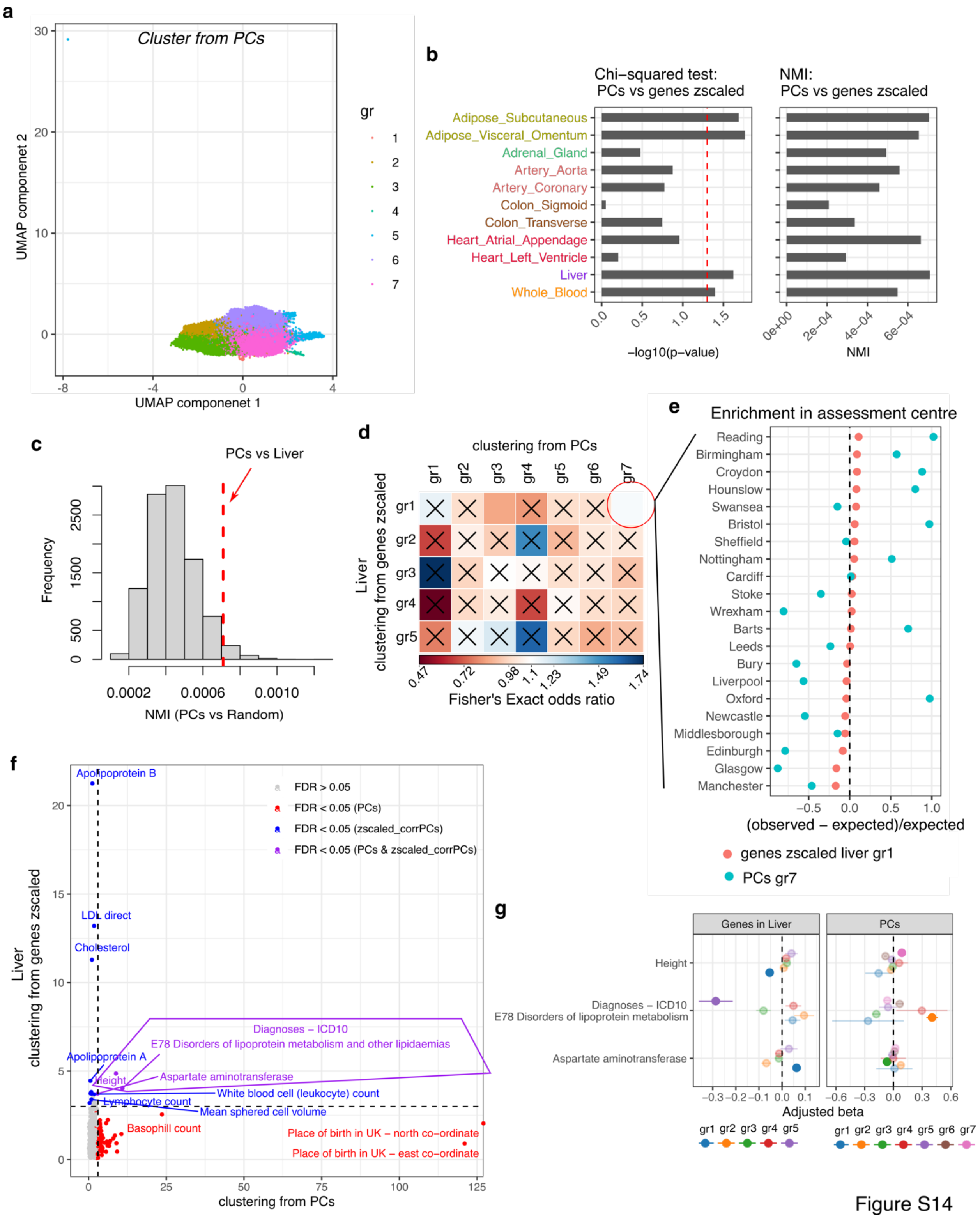


**Supplementary Fig. 14. Comparison of CAD clustering structure based on T-scores in liver and based on Principal Components from genotype data in UKBB.**

CAD cases are clustered using the first 40 PCs from UKBB (standardized). **a.** UMAP based on PCs, where the color refers to the assigned PC clustering. **b.** The comparison between PC clustering and gene T-score grouping is shown in terms of -log10 p-value of a chi-squared test (left panel) and NMI (right panel). The dashed line refers to the nominal p-value of 0.05, and the comparison is shown for each tissue. **c.** The histogram of NMI between the cluster from PCs and 10,000 randomly assigned groups with the same size as liver clustering is shown. The dashed line refers to the NMI comparing PCs and the actual liver clustering. **d.** The results of pairwise Fisher's Exact tests between a group detected in the PC clustering (columns) and a group detected in liver clustering (rows) are presented in a heatmap that indicates the computed odds ratio. Non-significant results at the nominal level of 0.01 are highlighted with an "x". **e.** Investigation of enrichment in the assessment center for groups 1 in liver and 7 in PC clustering. The x-axis indicates the fraction of (observed - expected)/expected counts as computed from the chi-squared statistic across the centers versus a group assignment (gr_i or not gr_i), while the y-axis indicates the center assignment. **f.** The results of testing each endophenotype are presented in dots that indicate the -log10 p-value of the most significant group-specific difference in PCs (x-axis) and liver (y-axis) clustering. The dashed lines refer to p-value = 0.001, and the color reflects the FDR significance threshold. **g.** Forest plot of the group-specific differences for the three endophenotypes that are significant in both PCs and liver clustering. The x-axis shows the regression coefficient from a GLM test of gr_i vs all remaining samples with 95% CI. The dots that are not shaded indicate the groups with the most significant association in terms of p-value.

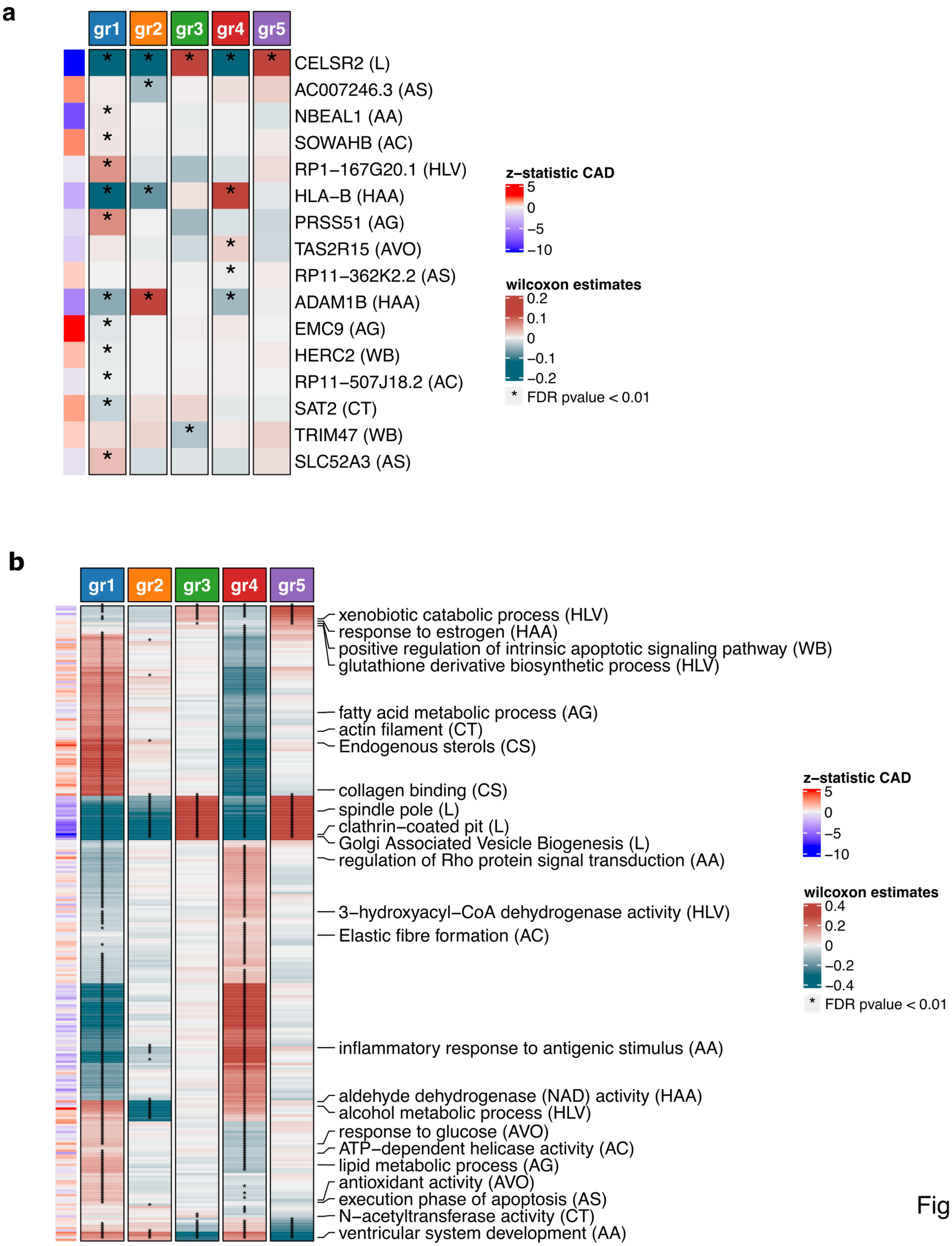


Figure S15

**Supplementary Fig. 15. Genes and pathways associated with CAD liver clustering structure**

**a.** Wilcoxon-Mann-Whithney (WMW) estimates for the most group-specific significant gene in the 16 group associated loci. In parenthesis the tissue considered with acronyms indicating to the tissue name initials. Row annotation on the left indicate the corresponding CAD Z-statistics from TWAS. **b.** WMW estimates (capped) for 271 significant pathways, considering only the most significant tissue per pathway. The rows are labeled with the names of the significant pathways, with the tissue considered indicated in parentheses. The left-side annotations show the corresponding CAD Z-statistics from PALAS.

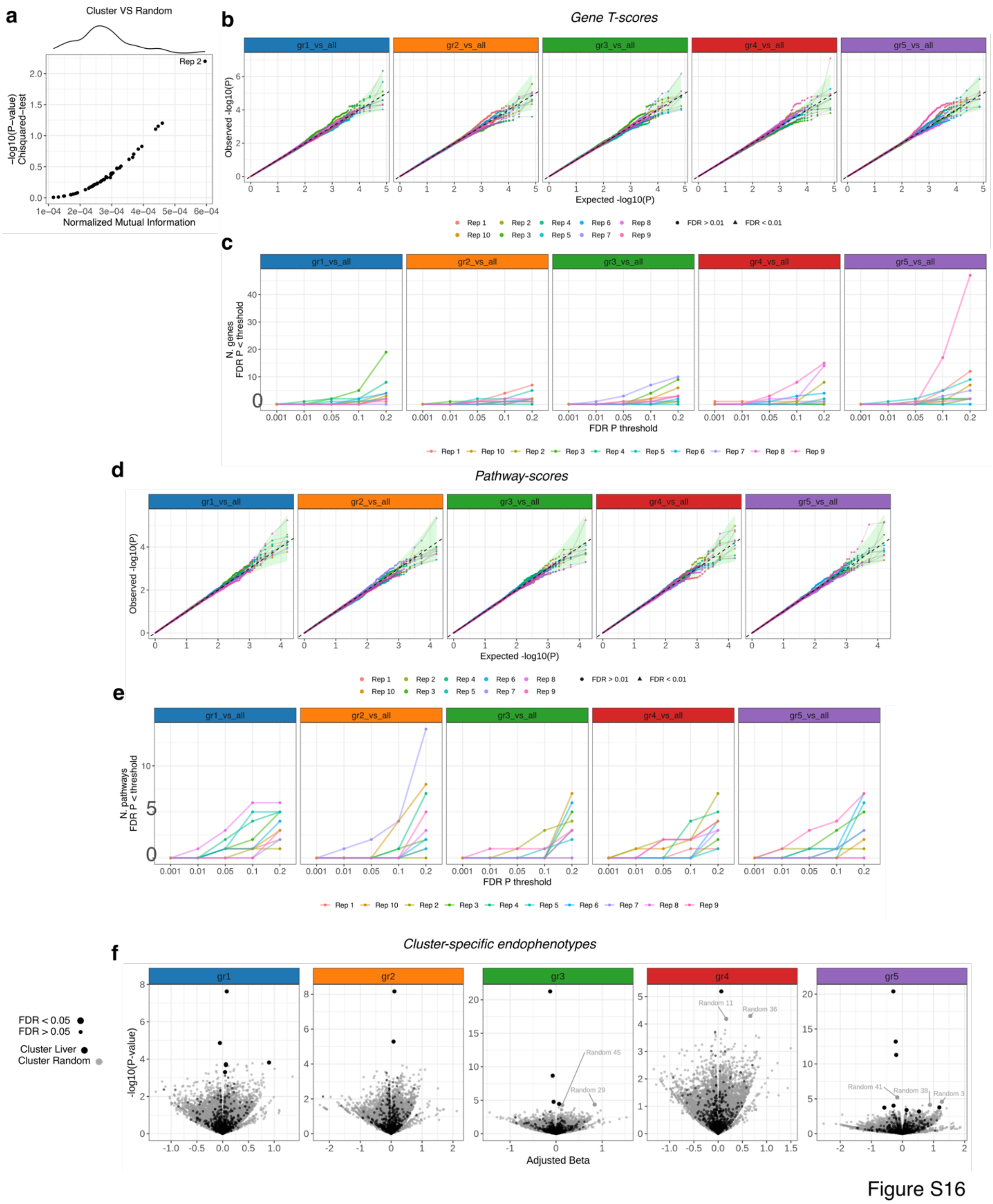


**Supplementary Fig. 16 Genes/pathways and endophenotype associations with random clustering structure**

Random clustering of CAD patients maintaining the same group size as liver grouping. **a.** X-axis NMI (x-axis) and -log10 p-values (y-axis) from $\chi 2$ statistics between liver clustering and random groupings. **b.** Quantile-quantile representation of the association between gene T-scores and group-wise specific (gri vs all remaining) testing, using the WMW method across all tissues. The expected p-values follow a uniform distribution, and the plot shows a dashed line along the diagonal, with a shaded green area representing a 95% confidence interval from the beta distribution. Each line in the plot corresponds to one of the 10 simulations. **c.** Number of significant associations that passed the FDR threshold for each of the 10 random clustering experiments as a function of varying FDR levels on the x-axis. **d.** Similar to b., pathway-scores group-specific differences for the selected gene-sets (Jaccard similarity $\leq 0.2$). **e.** Similar to c., pathway-scores group-specific counts based on FDR thresholds. **f.** Volcano plot of cluster-specific endophenotype differences. The x-axis represents the $\beta$ regression coefficient from the GLM, which refers to the features of gr_i versus the remaining cases, while the y-axis shows the corresponding -log10 p-value. Each grey dot represents an endophenotype among the 637 UK Biobank phenotypes that was tested for a random clustering configuration out of 50 repetitions. In contrast, each black dot refers to the endophenotype testing on the actual liver clustering. In both cases, the size of the dots corresponds to the significance of the test after correction (FDR $\leq 0.05$).

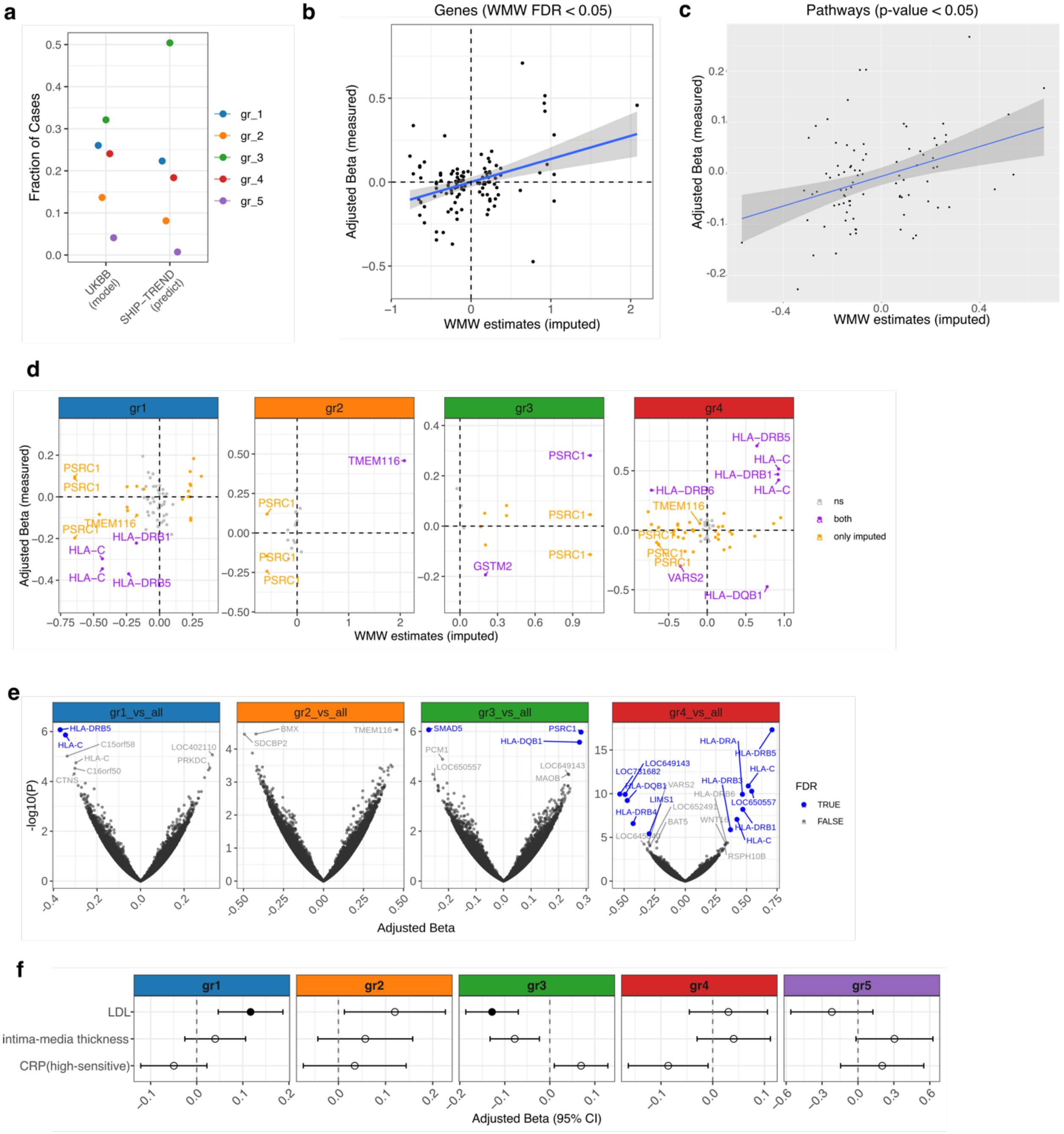


Figure S17

**Supplementary Fig. 17. Replication of CAD clustering on SHIP-Trend cohort**

Prediction of liver clustering structure on SHIP-Trend cohort. **a.** Y-axis indicates the fraction of CAD patients assigned to each group in UKBB data set and on the projected clustering for SHIP-Trend individuals. **b.** In whole blood, correlation of beta regression coefficients of group-wise gene level differences derived from predicted gene expression values (x-axis) and measured gene expression values in SHIP cohort (y-axis), considering only cluster-specific genes form imputed values in SHIP samples (WMW FDR 0.05 threshold). **c.** Similar to (B) but considering pathway-level group-specific differences and including only pathways with WMW nominal p-value below of 0.05, when testing imputed pathway values. **d.** Including only cluster-specific genes from the UKBB cohort, comparison between imputed gene expression (x-axis) and measured gene expression differences (y-axis), divided per group. Measured differences are adjusted for multiple-correction via BH procedure on this subset of genes. Each gene (dot) is colored by the significance (FDR 0.05) it reaches for both tests in SHIP cohort: grey not significant in both tests, purple significant in both and orange significant only when testing imputed gene expression differences. **e.** Volcano plots showing group-specific differences across all measured genes in whole blood for SHIP cohort. Dots in blue indicate genes passing FDR 0.05 threshold. **f.** Forest plot of cluster-specific endophenotype results in SHIP cohort. Out of the 20 clinical variable tested, the plot shows regression coefficient with 95% CI from GLM (x-axis) of those reaching nominal significance below 0.05. The full dots indicate results reaching FDR 0.05 threshold.

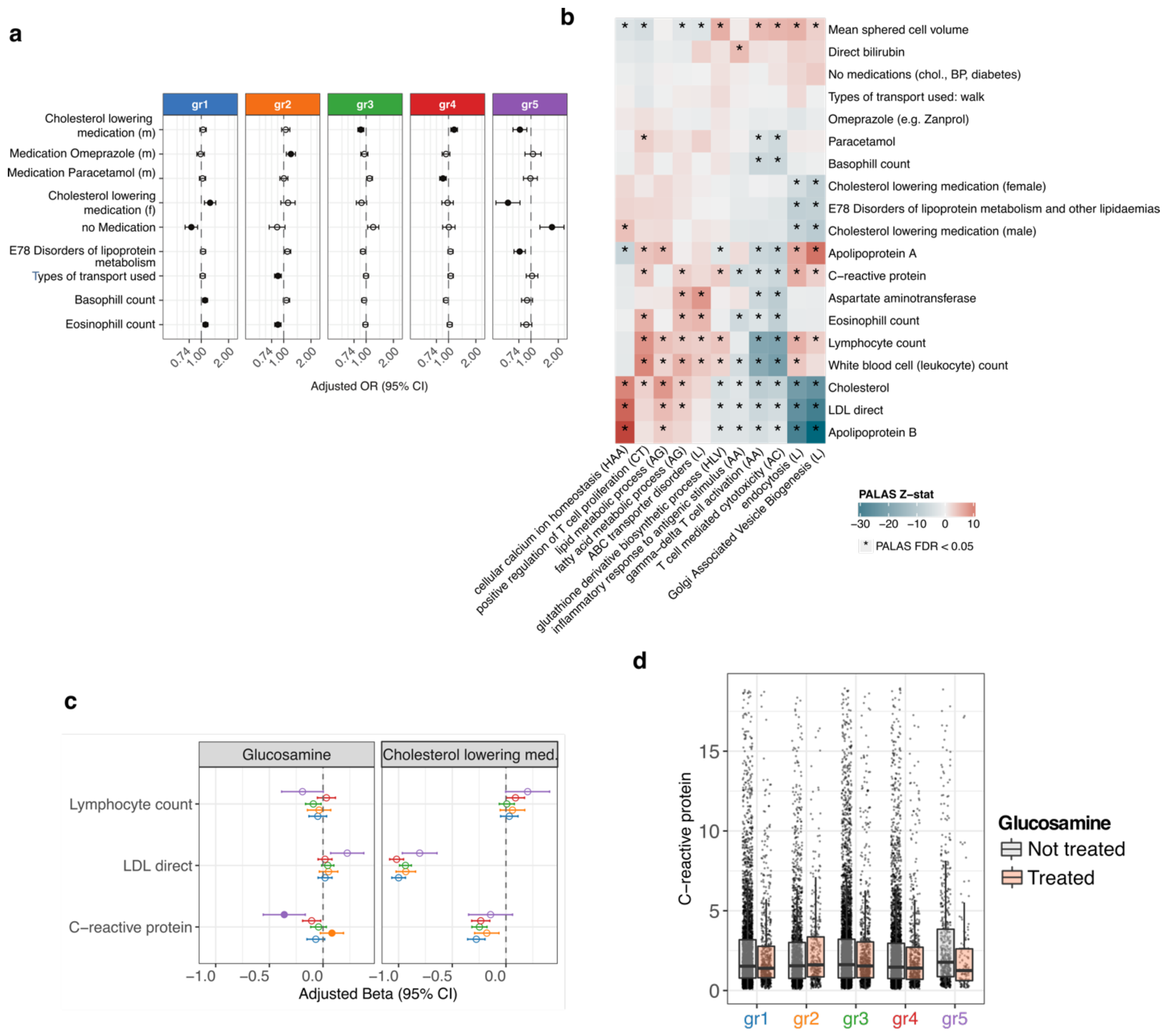


Figure S18

**Supplementary Fig. 18 Endophenotype and treatment response analyses on CAD clustering**

**a.** Among the 212 endophenotypes measured in UKBB with at least one CAD associated and group specific pathway, forest plot shows significantly different ones (FDR ≤ 0.05) in at least one group ($gr_i$ versus remaining patient) using Generalized Linear Model (GLM), indicating regression coefficient ($\beta_{GLM}$) with 95% Confidence Interval (CI). Full dot indicates that $\beta_{GLM}$ is significant after BH correction. This panel include binary or ordinal categorical phenotypes (m-male, f-female), continuous phenotypes are in Fig. 2e. **b.** Heatmap including PALAS Z-statistics for the

19 cluster-specific measured endophenotypes (rows) and a selection of cluster-specific pathways (columns). The * indicates associations that reach FDR 0.05 significance in PALAS. The capital abbreviation in the pathway names indicates the acronym of the tissue considered. HAA: Heart Atrial Appendage, CT: Colon Transverse, AG: Adrenal Gland, L: liver, HLV: Heart Left Ventricle, AA: Artery Aorta, AC: Artery Coronary. **c.** Treatment response showing the effect of glucosamine and cholesterol-lowering medications in each group for selected phenotypes. X-axis shows regression coefficient with 95% CI from GLM in each group, full dots indicate groups that are significantly different in a pairwise comparison after BH correction (pairwise comparison-specific and treatment-specific), tested using Z-test for comparing regression coefficients. **d.** Distribution of original CRP values in each group when taking or not glucosamine supplements, y-axis is cropped at CRP=20 mg/L excluding 330 outliers. Boxplot elements include median as central line, $1^{st}$ and $3^{rd}$ quartiles as box limits, 1.5 interquartile ranges from $1^{st}$ and $3^{rd}$ quartiles as corresponding whiskers.

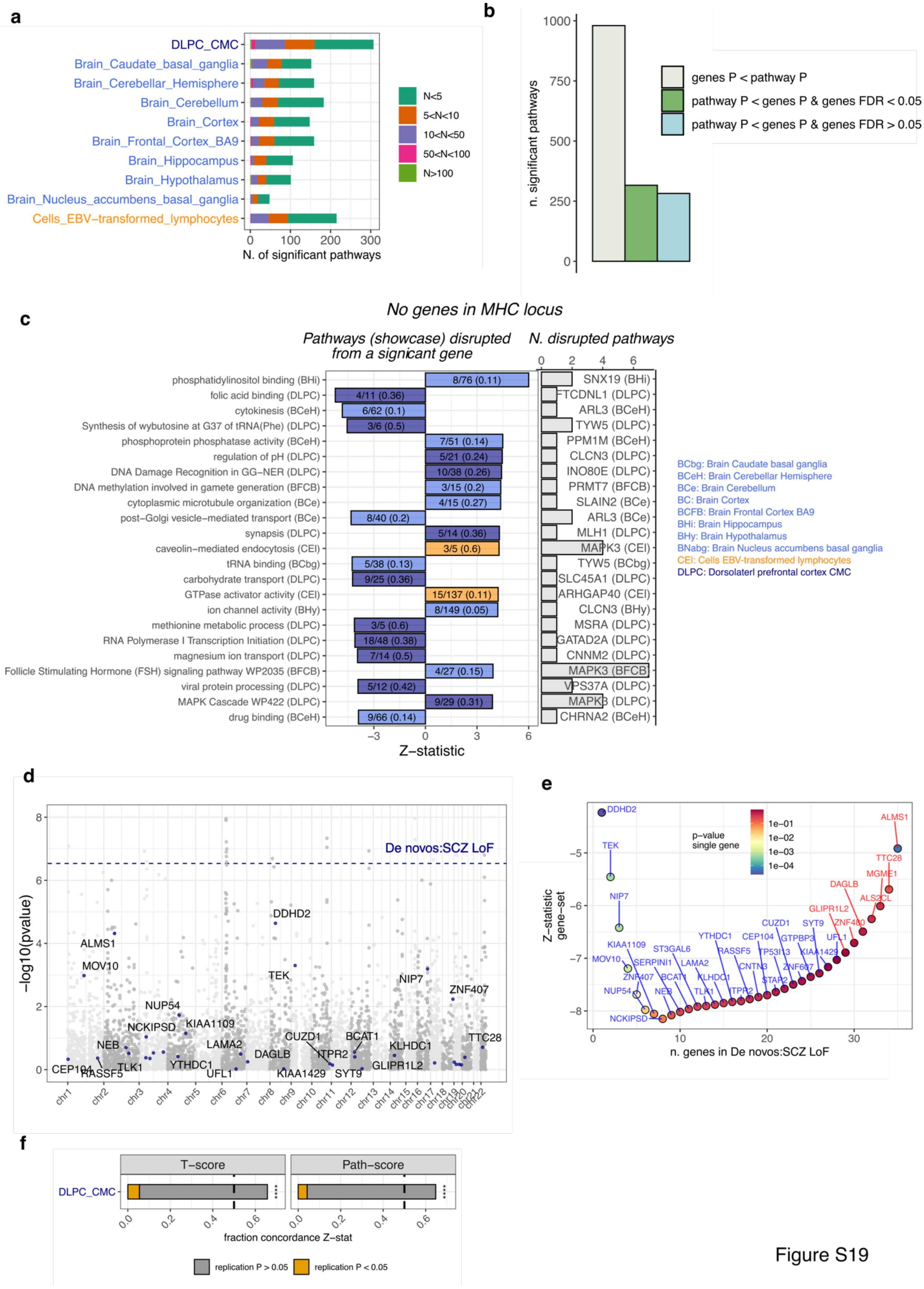
a
b
c
d
e
f
N. of significant pathways
n. significant pathways
genes P < pathway P
pathway P < genes P & genes FDR < 0.05
pathway P < genes P & genes FDR > 0.05
No genes in MHC locus
Pathways (showcase) disrupted from a signicant gene
N. disrupted pathways
Z−statistic
De novos:SCZ LoF
−log10(pvalue)
p−value single gene
Z−statistic gene−set
n. genes in De novos:SCZ LoF
T−score
Path−score
fraction concordance Z−stat
replication P > 0.05
replication P < 0.05
BCbg: Brain Caudate basal ganglia
BCeH: Brain Cerebellar Hemisphere
BCe: Brain Cerebellum
BC: Brain Cortex
BCFB: Brain Frontal Cortex BA9
BHi: Brain Hippocampus
BHy: Brain Hypothalamus
BNabg: Brain Nucleus accumbens basal ganglia
CEl: Cells EBV-transformed lymphocytes
DLPC: Dorsolaterl prefrontal cortex CMC

Figure S19

**Supplementary Fig. 19: PALAS summary results for SCZ and replicability.**

**a.** Significant pathways linked to SCZ in each tissue with tissue-specific FDR of less than or equal to 0.05. The bars represent the number of pathways, and their color represents the number of genes that are reliably predicted in that tissue (based on T-score genes). **b.** The number of pathways that are statistically significant (FDR $\leq$ 0.05) categorized into three groups based on the significance of their genes. The first group includes pathways where at least one gene is more significant than the pathway (ivory). The second group includes pathways where all genes are less significant than the pathway, but at least one gene has an FDR $\leq$ 0.05 (green). The third group includes pathways where all genes are less significant than the pathway and do not pass the FDR 0.05 threshold (light blue). **c.** Pathways with at least one gene more significant and no genes in the MHC locus, filtered using the prioritization criteria (computed from more than 5 and less or equal than 200 T-score genes or more than 2 if pathway coverage is higher than 10%, originally including less than 200 genes and reaching at least 0.0001 as nominal significance). The left panel shows PALAS Z-statistic (x-axis) color coded by the tissue of origin and indicating the gene pathway coverage for only one exemplar (best coverage) per most significant gene (in the right panel). The right panel shows the number of disrupted pathways by a certain gene. Acronyms in parenthesis indicate the initials of the tissue considered. **d.** CMC gene-set “De Novos: SCZ loss of function” in DLPC tissue. The dots indicate -log10(p-values) from TWAS for all tested genes in the given tissue. Genes that are included in the pathway are labelled and colored. The dashed line represents the -log10(p-value) of the considered pathway from PALAS. **e.** Incremental significance change of pathways is evaluated as genes from "De novos: SCZ LoF" are added one-by-one to the test gene set, with each addition leading to the computation of a pathway score from lowest to highest gene based on Z-statistic. The X-axis shows the number of genes in the incremental pathway analysis, while the Y-axis shows the corresponding Z-statistic level for each configuration. The labelled dot on the plot represents the gene that was added at each step, with the colour code indicating the actual TWAS p-value of that gene. Additionally, the color of the label indicates the sign of the gene-specific Z-statistic (blue for negative and red for positive). **f.** Reproducibility of gene levels T-scores (left) and pathway scores (right) on CMC data-set (478 individuals). X-axis shows the fraction of significant genes in PGC that have the same effect sign (Z-statistic) in CMC cohort, p-values are computed from one-sided sign test ($* * ** = P \leq 0.0001$). The fraction of genes concordant and nominal at a p-value threshold of 0.05 is shown in the yellow bar.

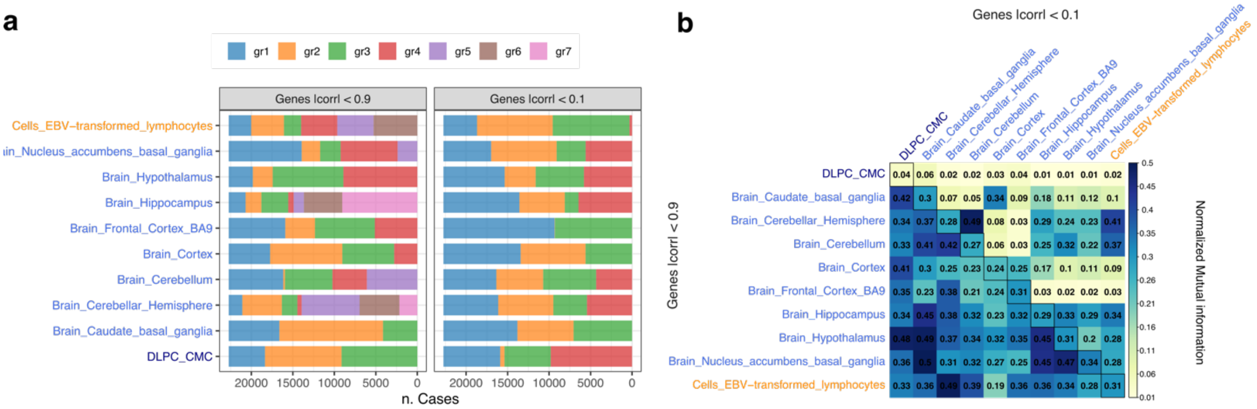


Figure S20

**Supplementary Fig. 20. Comparison of clustering structure for SCZ patients across tissues.** **a.** Proportion of individuals in each tissue-specific cluster among 22,687 patients for 2 different filtering strategies: genes are clumped based on imputed $R^2$ at Pearson correlation 0.9 (left) and correlation 0.1 (right). **b.** Normalized mutual information (NMI) for each pair of tissue-specific clustering structure in the 2 filtering strategies. Upper triangular matrix refers to |corr| < 0.9 preprocessing, lower triangular matrix to |corr| < 0.1 preprocessing, the diagonal shows NMI between the two filtering strategies in the same tissue.

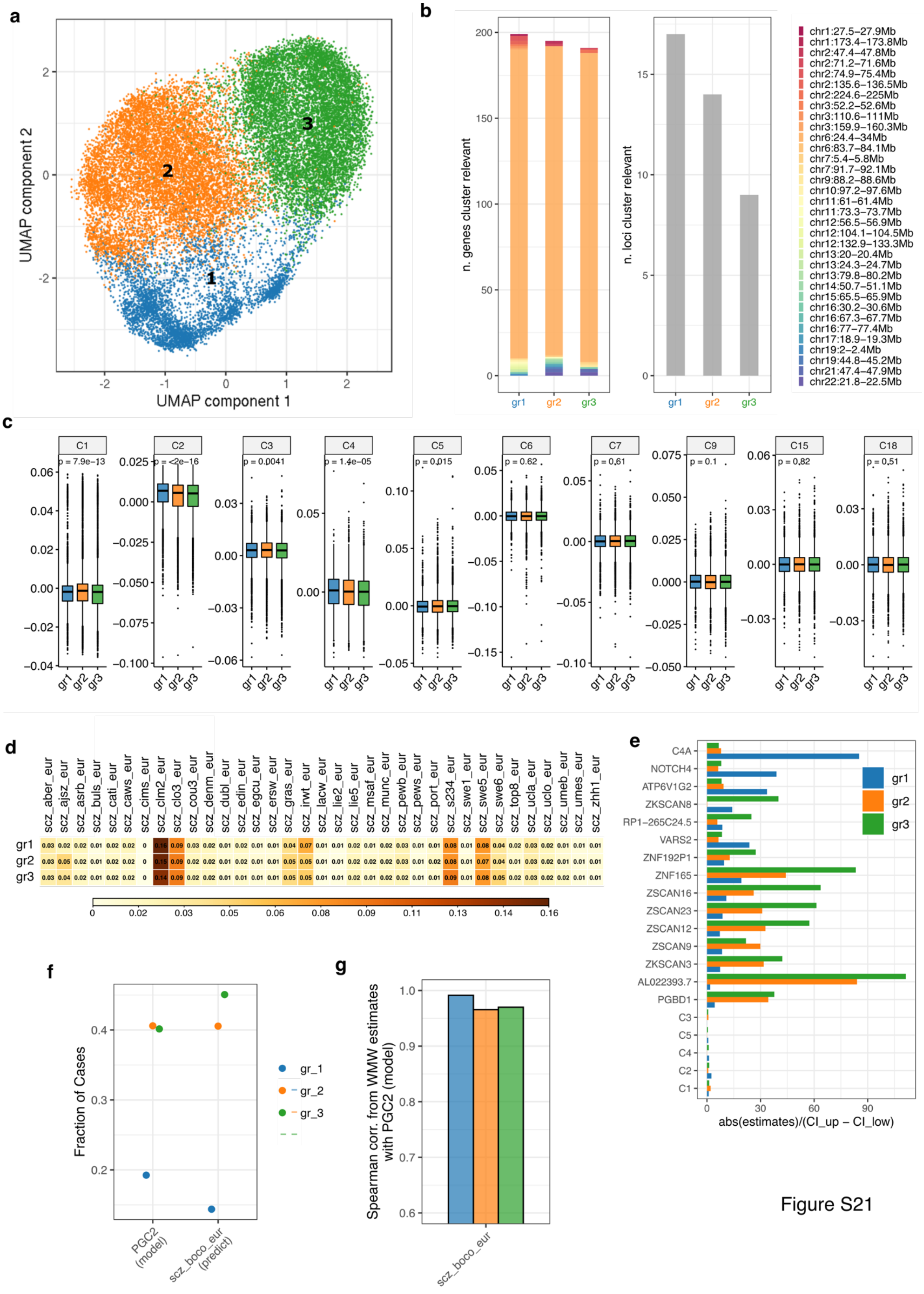
a
UMAP component 1
UMAP component 2
1
2
3
b
n. genes cluster relevant
n. loci cluster relevant
gr1
gr2
gr3
chr1:27.5–27.9Mb
chr1:173.4–173.8Mb
chr2:47.4–47.8Mb
chr2:71.2–71.6Mb
chr2:74.9–75.4Mb
chr2:135.6–136.5Mb
chr2:224.6–225Mb
chr3:52.2–52.6Mb
chr3:110.6–111Mb
chr3:159.9–160.3Mb
chr6:24.4–34Mb
chr6:83.7–84.1Mb
chr7:5.4–5.8Mb
chr7:91.7–92.1Mb
chr9:88.2–88.6Mb
chr10:97.2–97.6Mb
chr11:61–61.4Mb
chr11:73.3–73.7Mb
chr12:56.5–56.9Mb
chr12:104.1–104.5Mb
chr12:132.9–133.3Mb
chr13:20–20.4Mb
chr13:24.3–24.7Mb
chr13:79.8–80.2Mb
chr14:50.7–51.1Mb
chr15:65.5–65.9Mb
chr16:30.2–30.6Mb
chr16:67.3–67.7Mb
chr16:77–77.4Mb
chr17:18.9–19.3Mb
chr19:2–2.4Mb
chr19:44.8–45.2Mb
chr21:47.4–47.9Mb
chr22:21.8–22.5Mb
c
C1 p = 7.9e−13
C2 p = <2e−16
C3 p = 0.0041
C4 p = 1.4e−05
C5 p = 0.015
C6 p = 0.62
C7 p = 0.61
C9 p = 0.1
C15 p = 0.82
C18 p = 0.51
d
scz_aber_eur scz_ajsz_eur scz_asrb_eur scz_buls_eur scz_cati_eur scz_caws_eur scz_cims_eur scz_clm2_eur scz_clo3_eur scz_cou3_eur scz_denm_eur scz_dubl_eur scz_edin_eur scz_egcu_eur scz_ersw_eur scz_gras_eur scz_irwt_eur scz_lacw_eur scz_lie2_eur scz_lie5_eur scz_msaf_eur scz_munc_eur scz_pewb_eur scz_pews_eur scz_port_eur scz_s234_eur scz_swe1_eur scz_swe5_eur scz_swe6_eur scz_top8_eur scz_ucla_eur scz_uclo_eur scz_umeb_eur scz_umes_eur scz_zhh1_eur
gr1
gr2
gr3
e
C4A
NOTCH4
ATP6V1G2
ZKSCAN8
RP1−265C24.5
VARS2
ZNF192P1
ZNF165
ZSCAN16
ZSCAN23
ZSCAN12
ZSCAN9
ZKSCAN3
AL022393.7
PGBD1
C3
C5
C4
C2
C1
abs(estimates)/(CI_up − CI_low)
f
Fraction of Cases
PGC2 (model)
scz_boco_eur (predict)
gr_1
gr_2
gr_3
g
Spearman corr. from WMW estimates with PGC2 (model)
scz_boco_eur

Figure S21

**Supplementary Fig. 21: Genes and covariates associated with SCZ clustering structure in DLPC.**

**a.** Uniform manifold approximation and projection (UMAP) first 2 components of gene T-scores in DLPC standardized across SCZ patients, corrected for PCs, and multiplied by Z-statistic SCZ associations. Each dot represents a patient in the transformed UMAP space colored by the cluster membership. **b.** Number significant genes and loci (tissue specific FDR $\leq 0.01$) associated with each group from Wilcoxon-Mann-Whithney (WMW) test of $gr_i$ versus remaining patients combing all tissues. **c.** Distribution of PCs 1 to 10 for each group (p-values from Kruskal-Wallis test). **d.** Contingency table that displays the group and cohort structure, where each square in the table represents the fraction of patients in a group (rows) that belong to a particular cohort (columns). Each row of the table adds up to 1. **e.** X-axis shows the coefficient of variation, absolute value of WMW estimates divided by confidence interval ranges, for group-specific test. Y-axis indicates the first 5 genes per group based on WMW p-value and significant group-specific principal components (FDR $\leq 0.05$) **f-g.** Prediction of cluster membership for SCZ cases in external cohort (scz_boco_eur) through projection, validated via **f.** fraction of cases for each group in model (35 PGC2 cohorts) and external ones, **g.** group-specific Spearman correlation of WMW estimates in model and each external cohort only from genes that are significantly associated with that group across all tissues. Boxplot elements include median as central line, 1$^{st}$ and 3$^{rd}$ quartiles as box limits, 1.5 interquartile ranges from 1$^{st}$ and 3$^{rd}$ quartiles as corresponding whiskers.

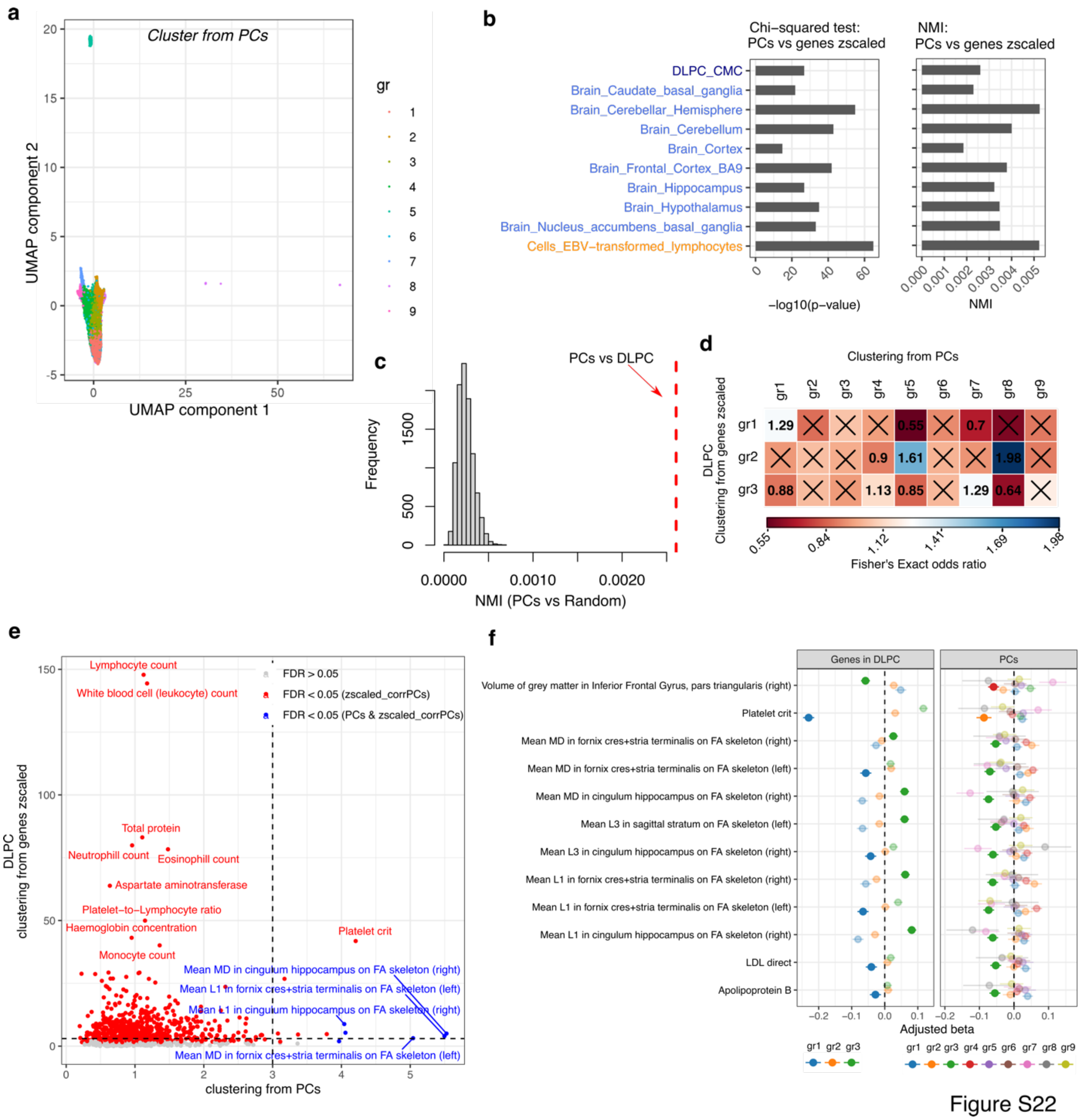


**Supplementary Fig. 22: Comparison SCZ clustering structure based on DLPC and based on PCs.**

SCZ cases clustered using the first 20 PGC PCs (standardized). **a.** UMAP based on PCs, where the color refers to the assigned PC clustering. **b.** For each tissue, comparison between PC clustering and gene T-score grouping is shown in terms of -log10 p-value of a chi-squared test (left panel) and NMI (right panel). **c.** The histogram of NMI between the cluster from PCs and 10,000 randomly assigned groups with the same size as liver clustering is shown. The dashed line refers to the NMI comparing PCs and the actual DLPC clustering. **d.** The results of pairwise Fisher's Exact tests between a group detected in PC clustering (columns) and a group detected in DLPC clustering (rows) are presented in a heatmap that indicates the computed odds ratio. Non-significant results at the nominal level of 0.01 are highlighted with an "x". **e.** The results of testing each endophenotype are presented in dots that indicate the -log10 p-value of the most significant group-specific difference in PCs (x-axis) and DLPC (y-axis) clustering. The dashed lines refer to p-value = 0.001, and the color reflects the FDR significance threshold. **f.** Forest plot of the group-specific differences for the endophenotypes that are significant in PCs cluster at FDR 0.1 threshold. The x-axis shows the regression coefficient from a GLM test of gr_i vs all remaining samples with 95% CI. The dots that are not shaded indicate the groups with the most significant association in terms of p-value.

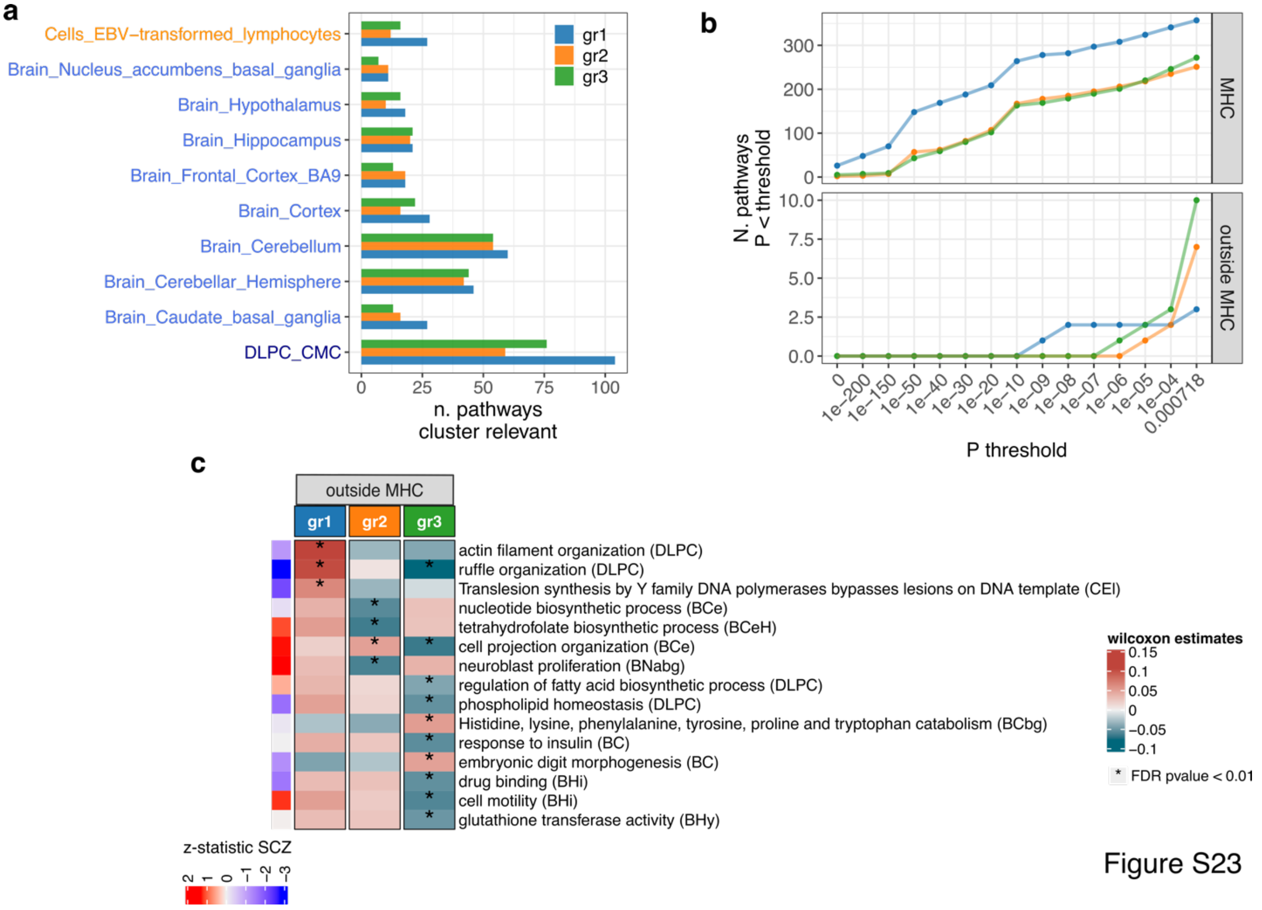


**Supplementary Fig. 23: Pathways associated with SCZ clustering structure in DLPC.**

**a.** The number of significant pathways (tissue specific FDR ≤ 0.01) associated with each group from the Wilcoxon-Mann-Whitney (WMW) test of group i versus the remaining patients. The pathways included in the analysis are from Reactome and GO and are filtered such that Jaccard similarity ≤ 0.2, retaining only the pathways with the highest coverage and removing significant pathways having discordant WMW estimates across tissues. **b.** For each group, the number of significant pathways passing the WMW p-value threshold is plotted on the y-axis against the p-value threshold on the x-axis, split into pathways that include at least one gene in the major histocompatibility complex (MHC) (top panel) and those that do not (bottom panel). **c.** The WMW estimates for the 15 significant pathways not including any genes in MHC (rows) are shown, testing each group against the rest (columns) and considering only the most significant tissue per pathway when repeated. The tissue tested is indicated by an acronym for the initial of the tissue name in parentheses. The row annotation on the left refers to the corresponding schizophrenia Z-statistics from the Psychiatric Genomics Consortium schizophrenia (SCZ) PGC2-PALAS dataset.

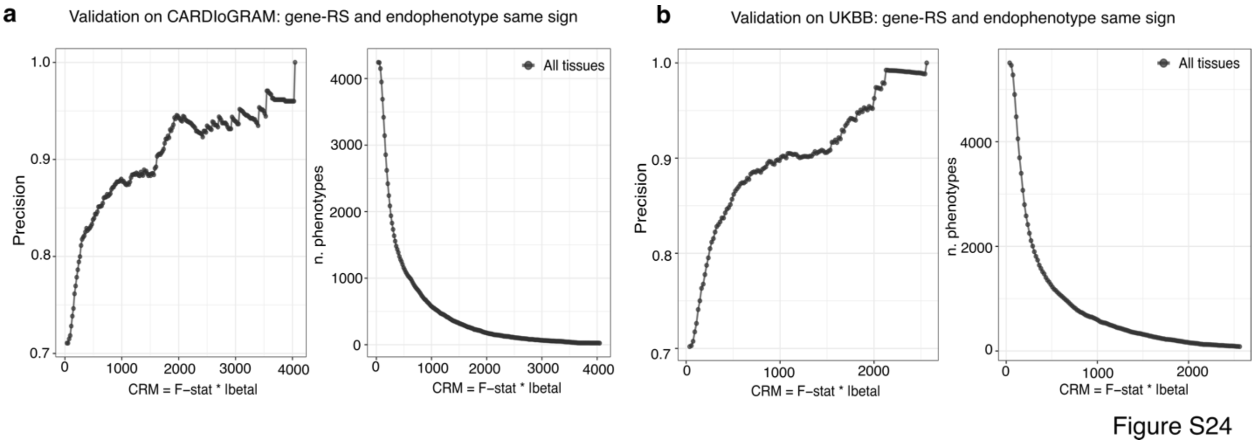


**Supplementary Fig. 24. Evaluation of gene-RS performance in resembling actual endophenotypes**

Validation of cluster-reliable measure (CRM) in CAD. **a.** Meta-analysis of gene-RS differences built on 9 CARDIoGRAM cohorts or **b.** UKBB. For the validation on UKBB, we included the same individuals used to build the original TWAS coefficients and to estimate R2. In both (A-B), CRM (x-axis) is compared to the actual CAD endophenotypic differences detected in each tissue-specific clustering, combined all together. On the left panel, the y-axis indicates the precision computed as the fraction of group-specific gene-RS differences having the same sign of GLM regression coefficient in the actual CAD endophenotype analysis, among all the endophenotypes passing CRM threshold. On the right panel, the y-axis indicates the number of phenotypes passing that CRM threshold.

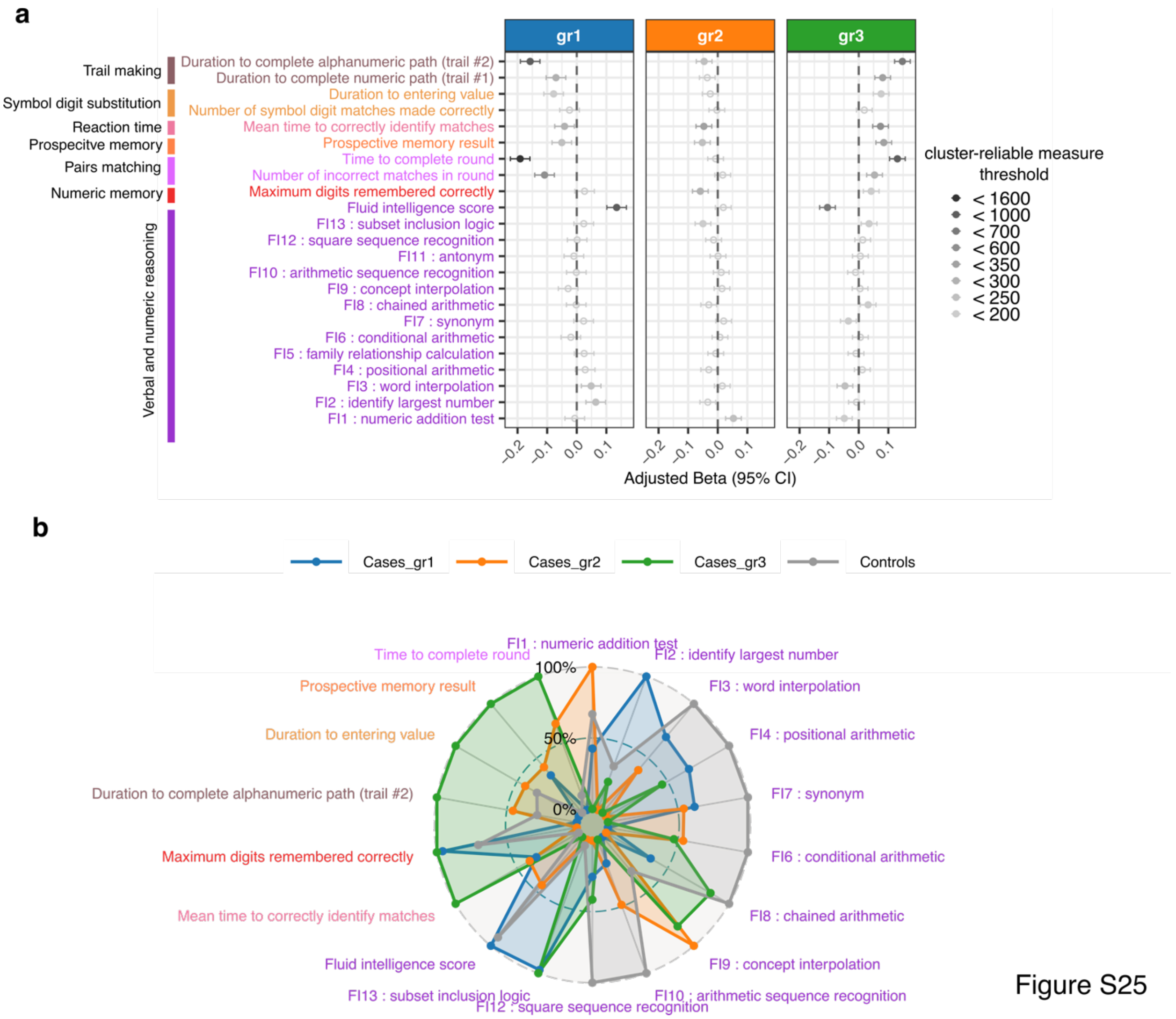


**Supplementary Fig. 25. Cognitive performance differences from gene-RS in DLPC SCZ clustering.**

**a.** Forest-plot of generalized linear model testing group-specific differences in gene-RS for a subset of cognitive performance phenotypes with 95% CI. Each dot in the plot represents the adjusted regression coefficient, which is filled if FDR is less than or equal to 0.05. The color of the dot corresponds to the credibility of the results according to the CRM threshold, with darker grey indicating more reliable results. **b.** Spider plot showing group-specific means (including controls) of gene-RS for cognitive performance phenotypes, rescaled to a range of 0-100. Each color on the chart represents a cognitive test class in the plot a.

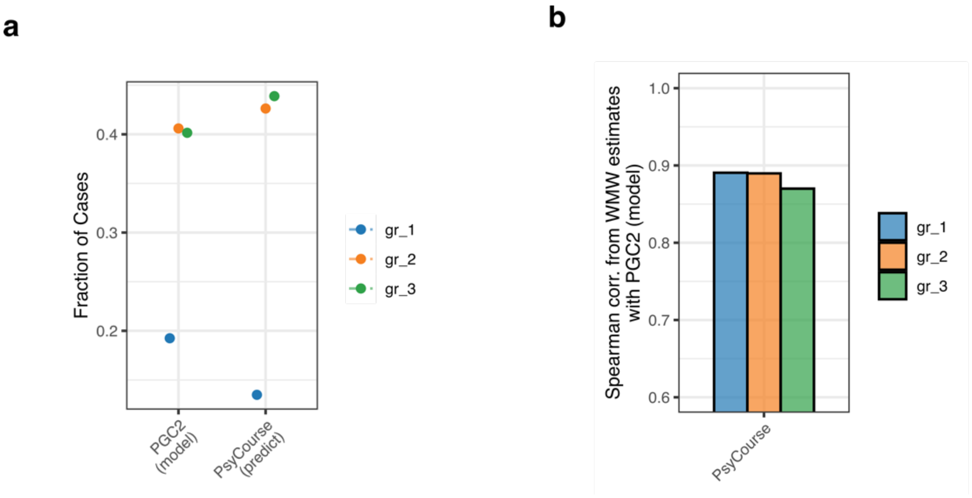


Figure S26

**Supplementary Fig. 26. Replication of PGC SCZ clustering on PsyCourse external cohort.** Prediction of cluster membership for SCZ cases in external cohort (PsyCourse) through projection, validated via **a.** fraction of cases for each group in model (35 PGC2 cohorts) and external ones, **b.** group-specific Spearman correlation of WMW estimates in model and each external cohort only from genes that are significantly associated with that group across all tissues.

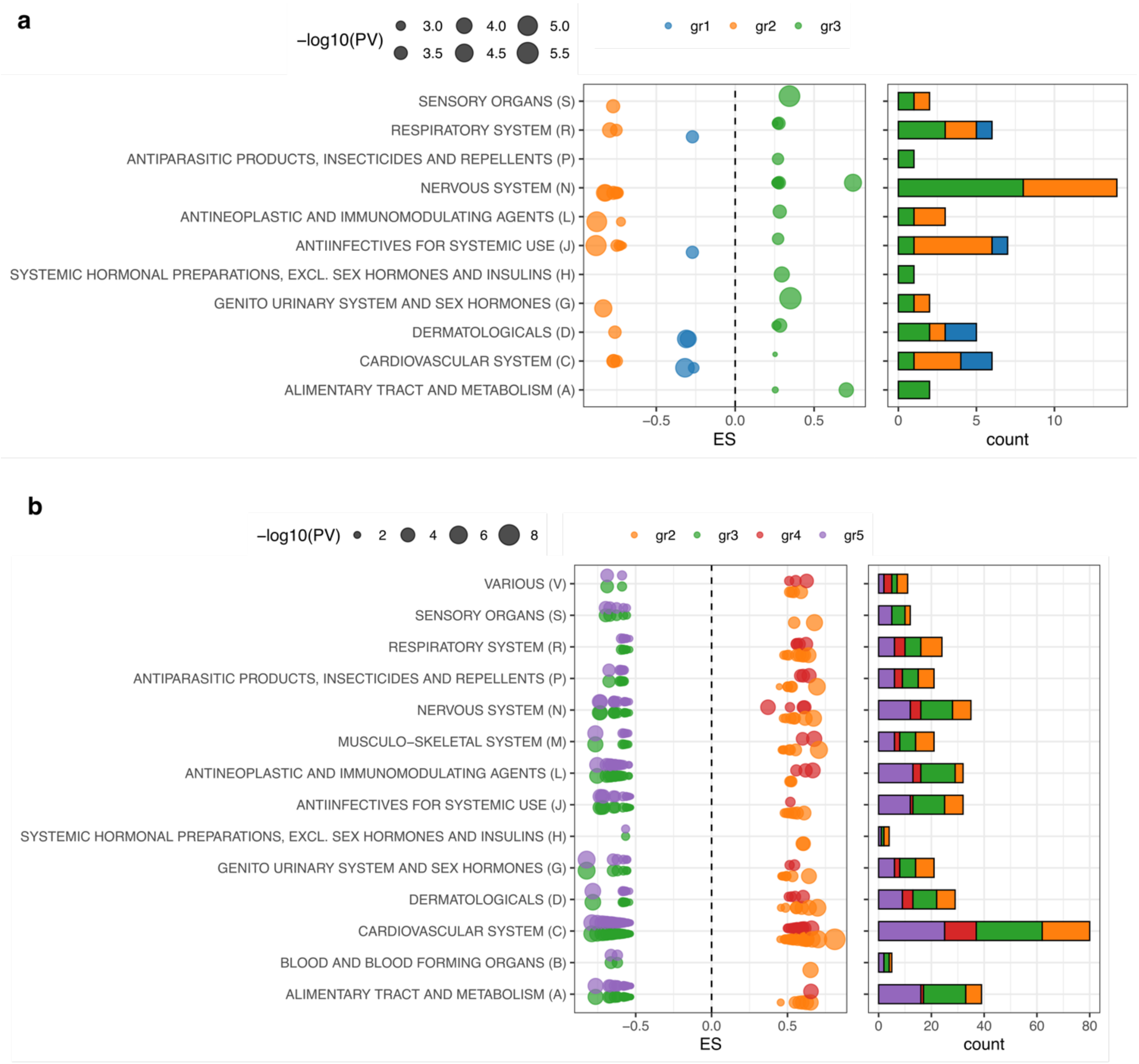


Figure S27

**Supplementary Fig. 27: Pathway analysis and drug response based on up- and down-regulated group-specific pathways.**

For each group in SCZ clustering **a.** and CAD clustering **b.**, the number of associated drugs from "gep2pep" output are shown (FDR $\leq$ 0.05, right panel) and the corresponding enrichment score from GSEA (left panel), including only those drugs whose name matched ATC annotation. The x-axis in the left panel indicates the enrichment score (ES) and dot size indicate the -log10 p-value of GSEA. The GSEA tests the cumulative changes of pathway response to a drug administration in a cluster-specific set of pathways (either up- or down-regulated, based on WMW estimate sign).

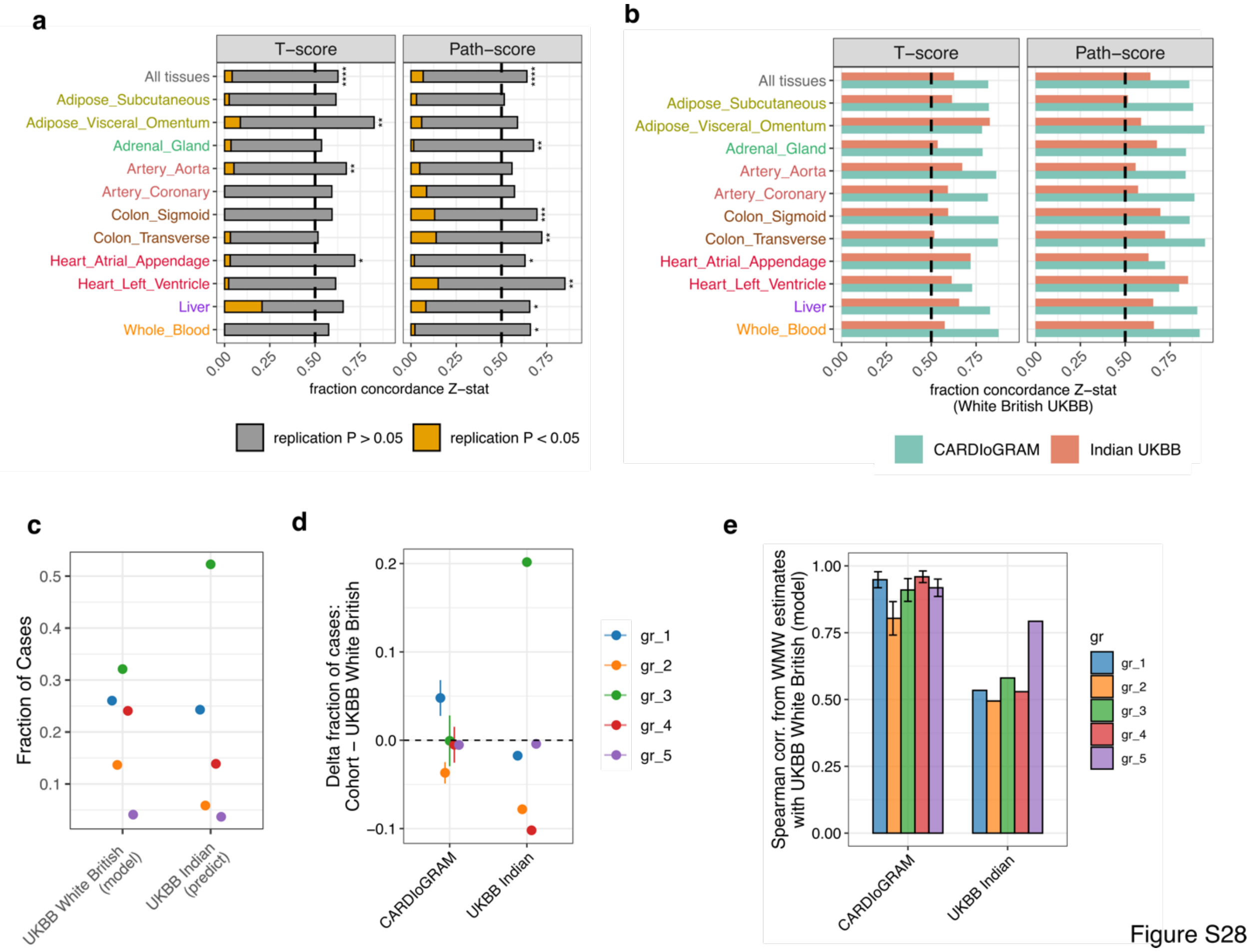


**Supplementary Fig. 28: Trans-ancestry application of CASTom-iGEx.**

CASTom-iGEx pipeline is applied in the context of CAD across Indian individuals in UKBB data set. **a.** Reproducibility of TWAS (left) and PALAS (right) significant results for UKBB white British (UKBB WB) cohort in the UKBB Indian population. X-axis shows the fraction of significant genes in UKBB WB that have the same effect sign (Z-statistic) in UKBB Indian. P-values are computed from one-sided sign test ($* = P \leq 0.05$, $** = P \leq 0.01$, $*** = P \leq 0.001$ , $**** = P \leq 0.0001$). The fraction of genes concordant and nominal at a p-value threshold of 0.05 is shown in the yellow bar. **b.** Fraction of reproduced significant results based on Z-statistic sign for UKBB WB in Indian UKBB cohort (red) and CARDIoGRAM meta-analysis (green). **c.** Projection of CAD cases clustering in liver from UKBB WB on UKBB Indian. Y-axis shows the fraction of cases assigned to each cluster in UKBB WB dataset and UKBB Indian. **d.** Comparison with CARDIoGRAM clustering projection. Y-axis shows the difference in fraction of cases between the external cohort into which the clustering was projected (CARDIoGRAM or UKBB Indian) and the model clustering cohort (UKBB WB). For CARDIoGRAM, average across 9 cohorts +/- standard deviation (error bars) is displayed. **e.** For each group, Spearman correlation of WMW estimates in UKBB WB and the external cohorts (CARDIoGRAM and UKBB Indian) only from genes that are significantly associated with that group across all tissues. For CARDIoGRAM, average across 9 cohorts +/- standard deviation (error bars) is displayed.

| Tissue | N of samples | N of genes | N of prior features | N of reliable genes | N of reg-SNPs | Percentage of reg-SNPs with Prior | Percentage of reg-SNPs in DHS |
|---|---|---|---|---|---|---|---|
| Dorsolateral Prefrontal Cortex | 478 | 15578 | 15 | 6854 | 366706 | 36.65 | NA |
| Adipose Subcutaneous | 242 | 25971 | 3 | 6058 | 423635 | 25.85 | 34.63 |
| Adipose Visceral Omentum | 164 | 25139 | 3 | 3910 | 386744 | 27.68 | 32.01 |
| Adrenal Gland | 105 | 23624 | 1 | 3027 | 377947 | 23.50 | 32.18 |
| Artery Aorta | 185 | 24274 | 7 | 5277 | 402688 | 38.45 | 33.31 |
| Artery Coronary | 99 | 23880 | 7 | 2298 | 343528 | 39.24 | 33.41 |
| Artery Tibial | 239 | 24335 | 7 | 5918 | 364109 | 41.93 | 34.86 |
| Brain Caudate basal ganglia | 90 | 24512 | 15 | 2635 | 349538 | 39.59 | 34.46 |
| Brain Cerebellar Hemisphere | 77 | 23762 | 15 | 2941 | 393519 | 39.13 | 34.69 |
| Brain Cerebellum | 93 | 24570 | 15 | 3788 | 429782 | 36.47 | 33.85 |
| Brain Cortex | 81 | 24110 | 15 | 2501 | 368497 | 38.62 | 34.66 |
| Brain Frontal Cortex BA9 | 77 | 23765 | 15 | 2041 | 372502 | 38.05 | 34.19 |
| Brain Hippocampus | 74 | 23723 | 15 | 1671 | 313967 | 43.17 | 36.49 |
| Brain Hypothalamus | 72 | 24426 | 15 | 1565 | 342292 | 37.26 | 33.86 |
| Brain Nucleus accumbens basal_ganglia | 81 | 24386 | 15 | 2290 | 341053 | 36.89 | 33.16 |
| Cells EBV-transformed lymphocytes | 94 | 21779 | 2 | 2706 | 372078 | 26.15 | 31.63 |
| Colon Sigmoid | 118 | 24051 | 8 | 2925 | 379960 | 33.05 | 35.08 |
| Colon Transverse | 145 | 25354 | 8 | 3522 | 374357 | 30.48 | 33.25 |
| Esophagus Gastroesophageal Junction | 115 | 23575 | 8 | 3030 | 346204 | 28.65 | 32.55 |
| Esophagus Mucosa | 224 | 25038 | 8 | 6107 | 381943 | 31.00 | 34.21 |
| Esophagus Muscularis | 199 | 24360 | 8 | 5754 | 379200 | 29.50 | 32.87 |
| Heart Atrial Appendage | 151 | 23666 | 7 | 2733 | 511496 | 33.77 | 32.41 |
| Heart Left Ventricle | 172 | 22681 | 7 | 3628 | 294768 | 45.07 | 36.77 |
| Liver | 94 | 22158 | 2 | 2215 | 306512 | 32.95 | 33.31 |
| Lung | 241 | 27372 | 1 | 5749 | 399559 | 20.40 | 31.66 |
| Muscle Skeletal | 297 | 22942 | 2 | 5566 | 292706 | 33.73 | 36.10 |
| Pancreas | 132 | 23153 | 1 | 3631 | 361244 | 22.70 | 32.41 |
| Skin Not Sun Exposed Suprapubic | 173 | 25922 | 1 | 4740 | 407576 | 13.36 | 30.82 |
| Skin Sun Exposed Lower leg | 252 | 26582 | 1 | 6614 | 427755 | 13.79 | 31.33 |
| Small Intestine Terminal Ileum | 74 | 25010 | 8 | 1594 | 500251 | 28.62 | 32.89 |
| Spleen | 79 | 24354 | 1 | 2556 | 442687 | 19.90 | 32.35 |
| Stomach | 144 | 24861 | 8 | 3215 | 339228 | 29.71 | 33.23 |
| Thyroid | 233 | 27305 | 1 | 7447 | 434307 | 18.44 | 31.73 |
| Whole Blood | 280 | 22805 | 6 | 4644 | 279175 | 35.69 | 35.75 |

**Supplementary Table 1. Summary of tissue-specific gene expression models via PriLer.**

Summary of tissue specific gene-expression models in terms of number of samples, genes, prior feature applied, reliable genes, regulatory variants (reg-SNPs), percentage of reg-SNPs intersecting a prior used in the model and a DNase I hypersensitive sites (DHS) in at least 1 biosample.

| Loci | N. genes | CAD best Z-statistic | Tissue | Group-specific differences | Best group-specific gene | WMW estimate best gene | WMW p-value best gene |
|---|---|---|---|---|---|---|---|
| chr1:109.3-110.5Mb | 8 | 3,411174828 | Artery_Aorta | gr1,gr2,gr3,gr4,gr5 | CELSR2 | 0.02761,0.02442,-0.06886,0.02981,-1.16541 | 1.99e-36,2.62e-20,2.37e-104,2.65e-40,3.39e-102 |
| chr1:109.4-110.5Mb | 7 | 4,97599405 | Heart_Atrial_Appendage | gr1,gr2,gr3,gr4,gr5 | SYPL2 | 0.26639,0.24292,-0.53989,0.27574,-1.11943 | 6.45e-84,4.18e-45,2.46e-290,5.4e-86,3.03e-127 |

| | | | | | | | |
|---|---|---|---|---|---|---|---|
| chr1:109.4-110.4Mb | 6 | 3,334446629 | Adrenal_Gland | gr1,gr2,gr3,gr4,gr5 | SARS | 0.40403,0.3579,-0.69966,0.44269,-1.2753 | 2.33e-127,7.25e-64,0,1.23e-146,2.05e-235 |
| chr1:109.4-110.5Mb | 6 | 4,583618248 | Colon_Sigmoid | gr1,gr2,gr3,gr4,gr5 | CELSR2 | 0.30218,0.25229,-0.63743,0.3044,-1.37487 | 2.56e-122,4.35e-58,0,2.92e-116,1.52e-242 |
| chr1:109.4-110.5Mb | 9 | -7,028664047 | Whole_Blood | gr1,gr2,gr3,gr4,gr5 | PSRC1 | -0.60816,-0.51929,1.15011,-0.58608,2.07084 | 0,2.69e-167,0,4.81e-306,0 |
| chr1:109.4-110.5Mb | 6 | -4,580688604 | Adipose_Subcutaneous | gr1,gr2,gr3,gr4,gr5 | RP5-1065J22.8 | -0.25132,-0.17771,0.48696,-0.27048,0.97473 | 3.77e-73,3.24e-24,4.8e-234,4.11e-81,1.16e-105 |
| chr1:109.6-110.5Mb | 4 | -1,666639122 | Adipose_Visceral_Omentum | gr1,gr2,gr3,gr4,gr5 | RP4-735C1.4 | -0.09594,-0.12862,0.19399,-0.12219,0.32612 | 3.66e-08,7.67e-09,3.59e-33,8.32e-12,4.36e-18 |
| chr1:109.6-110.3Mb | 6 | -10,00933509 | Liver | gr1,gr2,gr3,gr4,gr5 | CELSR2 | -0.2424,-0.12874,1.74223,-0.2003,3.42219 | 0,2.58e-259,0,0,0 |
| chr1:109.8-110.5Mb | 6 | 3,031988898 | Artery_Coronary | gr1,gr2,gr3,gr4,gr5 | SYPL2 | 0.13727,0.10924,-0.26075,0.13562,-0.60034 | 3.6e-24,7.55e-11,6.17e-68,2.29e-22,1.25e-42 |
| chr1:109.8-110.5Mb | 4 | -2,396183221 | Heart_Left_Ventricle | gr1,gr2,gr3,gr4,gr5 | SYPL2 | 0.14649,0.14693,-0.28754,0.15085,-0.59976 | 2.49e-22,1.01e-14,1.12e-71,8.84e-23,3.53e-44 |
| chr1:109.9-110.5Mb | 3 | -2,253282661 | Colon_Transverse | gr1,gr2,gr3,gr4,gr5 | AMIGO1 | -0.07133,-0.06855,0.16263,-0.09108,0.40657 | 9.36e-11,6.49e-07,3.74e-35,5.29e-15,1.21e-26 |
| chr2:39.5-39.9Mb | 1 | 1,24376926 | Adipose_Subcutaneous | gr2 | AC007246.3 | -0.03977 | 1.81e-05 |
| chr2:203.7-204.1Mb | 1 | -7,564368215 | Artery_Aorta | gr1 | NBEAL1 | 0.00876 | 7.56e-05 |
| chr4:77.6-78Mb | 1 | 1,355510651 | Artery_Coronary | gr1 | SOWAHB | 0.00809 | 2.04e-05 |
| chr5:16.2-16.6Mb | 1 | -0,430817975 | Heart_Left_Ventricle | gr1 | RP1-167G20.1 | 0.06094 | 7.27e-05 |
| chr6:25.5-33.7Mb | 68 | -4,379322809 | Adipose_Visceral_Omentum | gr1,gr2,gr4 | HLA-B | -0.85333,-0.08019,0.96545 | 0,2.62e-05,0 |
| chr6:25.5-33.9Mb | 63 | -4,110328139 | Adrenal_Gland | gr1,gr2,gr4 | HLA-DQB1-AS1 | -0.46386,-0.07382,0.55972 | 6.9e-198,7.7e-06,1.08e-256 |
| chr6:25.7-33.7Mb | 50 | -3,991651358 | Liver | gr1,gr2,gr4 | CYP21A2 | 0.28199,0.08718,-0.43291 | 4.89e-67,1.51e-05,2.41e-112 |
| chr6:25.8-33.9Mb | 82 | 4,580248435 | Artery_Aorta | gr1,gr2,gr4 | HCG27 | 0.52707,0.08608,-0.65365 | 6.51e-229,1.58e-05,4.73e-297 |
| chr6:25.8-33.4Mb | 69 | 4,312673204 | Colon_Transverse | gr1,gr2,gr4 | HLA-DQB1-AS1 | -0.27096,-0.06134,0.37388 | 2.42e-112,2.15e-05,6.71e-159 |
| chr6:25.8-33.8Mb | 73 | 4,373413098 | Whole_Blood | gr1,gr2,gr4 | HLA-B | -0.78835,-0.08126,0.92798 | 0,1.88e-05,0 |
| chr6:25.8-33.7Mb | 76 | -4,74790597 | Heart_Left_Ventricle | gr1,gr2,gr4 | HLA-DRB1 | -0.18475,-0.04601,0.281 | 2.62e-107,1.61e-06,1.78e-158 |
| chr6:25.8-33.7Mb | 98 | -4,285459753 | Adipose_Subcutaneous | gr1,gr2,gr4 | HLA-B | -0.77594,-0.08691,0.87628 | 0,1.99e-05,0 |
| chr6:26-34Mb | 63 | 4,487397064 | Colon_Sigmoid | gr1,gr2,gr4 | CYP21A2 | 0.5061,0.1101,-0.65137 | 1.06e-213,5.09e-08,7.66999999999999e-290 |
| chr6:26.2-28.4Mb | 6 | -1,19229995 | Artery_Coronary | gr1,gr4 | BTN3A2 | 0.12674,-0.13485 | 2.23e-55,3.07e-48 |
| chr6:26.2-33.8Mb | 50 | -4,138661172 | Heart_Atrial_Appendage | gr1,gr2,gr4 | HLA-B | -0.86251,-0.07401,0.9944 | 0,1.88e-05,0 |
| chr6:29.4-34Mb | 49 | -3,751226577 | Artery_Coronary | gr1,gr2,gr4 | HLA-DRB1 | -0.13472,-0.03993,0.28299 | 1.84e-66,1.94e-07,1.6e-114 |
| chr8:10.2-10.6Mb | 1 | -1,068128446 | Adrenal_Gland | gr1 | PRSS51 | 0.06507 | 0.000144 |
| chr12:10.9-11.3Mb | 1 | -1,620381613 | Adipose_Visceral_Omentum | gr4 | TAS2R15 | 0.02217 | 0.00016 |
| chr12:58.7-59.1Mb | 1 | 0,468416525 | Adipose_Subcutaneous | gr4 | RP11-362K2.2 | -0.00446 | 0.000179 |
| chr12:110.1-110.5Mb | 1 | -0,33645925 | Adipose_Subcutaneous | gr2 | GLTP | -0.11059 | 2.01e-07 |
| chr12:110.1-110.5Mb | 1 | -1,038637167 | Artery_Coronary | gr2 | GLTP | -0.01984 | 5.63e-06 |

| | | | | | | | |
|---|---|---|---|---|---|---|---|
| chr12:110.7-112.7Mb | 7 | -6,146002431 | Adipose_Visceral_Omentum | gr1,gr2,gr4 | TMEM116 | -0.14798,2.15816,-0.09355 | 3.98e-166,0,3.94e-99 |
| chr12:110.7-113.5Mb | 6 | -4,622736246 | Whole_Blood | gr1,gr2,gr4 | TMEM116 | -0.20588,2.0451,-0.17274 | 3.01e-192,0,2.34e-121 |
| chr12:110.7-112.7Mb | 4 | 3,751344091 | Colon_Transverse | gr1,gr2,gr4 | TMEM116 | -0.17116,2.00887,-0.15359 | 2.3e-139,0,2.81e-103 |
| chr12:111.2-112.7Mb | 6 | -5,856876271 | Heart_Left_Ventricle | gr1,gr2,gr4 | RP3-462E2.5 | 0.31558,-1.89141,0.22527 | 4.36e-189,0,3.67e-126 |
| chr12:111.9-112.7Mb | 2 | -6,644796997 | Liver | gr1,gr2,gr4 | TMEM116 | -0.07999,2.30002,-0.06325 | 8.15e-165,0,2.59e-103 |
| chr12:112-112.7Mb | 5 | -5,441387426 | Artery_Aorta | gr1,gr2,gr4 | TMEM116 | -0.10892,1.95019,-0.0882 | 1.74e-82,0,4.26e-56 |
| chr12:112-112.7Mb | 4 | -5,026537808 | Heart_Atrial_Appendage | gr1,gr2,gr4 | ADAM1B | -0.05466,2.40601,-0.04308 | 9.02e-211,0,1.93e-132 |
| chr12:112.1-113.5Mb | 6 | 5,289388769 | Adipose_Subcutaneous | gr1,gr2,gr4 | TMEM116 | -0.04571,2.36515,-0.03551 | 3.4e-125,0,6.22e-78 |
| chr12:112.1-113.7Mb | 4 | -4,949456162 | Artery_Coronary | gr1,gr2,gr4 | TMEM116 | -0.10179,2.14716,-0.08338 | 6.93e-83,0,2.89e-61 |
| chr12:112.1-113.8Mb | 2 | 3,231114658 | Adrenal_Gland | gr1,gr2,gr4 | ADAM1A | 0.03358,-1.59637,0.02995 | 4.79e-46,2.92e-212,5.79e-37 |
| chr12:112.1-112.9Mb | 3 | 5,111683548 | Colon_Sigmoid | gr1,gr2,gr4 | TMEM116 | -0.23844,1.90541,-0.17158 | 2.65e-163,0,3.29e-99 |
| chr14:24.4-24.8Mb | 1 | 2,368723351 | Adrenal_Gland | gr1 | EMC9 | -0.00907 | 0.00012 |
| chr15:28.4-28.8Mb | 1 | 0,692670835 | Whole_Blood | gr1 | HERC2 | -0.00543 | 2.41e-05 |
| chr15:50.8-51.2Mb | 1 | -0,5857644 | Artery_Coronary | gr1 | RP11-507J18.2 | -0.00415 | 0.000214 |
| chr17:7.3-7.7Mb | 1 | -0,039006589 | Adipose_Subcutaneous | gr1 | TNFSF12 | 0.01924 | 2.88e-05 |
| chr17:7.3-7.7Mb | 1 | 1,045534927 | Colon_Transverse | gr1 | SAT2 | -0.02237 | 5,00E-05 |
| chr17:73.7-74.1Mb | 1 | 0,445465707 | Whole_Blood | gr3 | TRIM47 | -0.03431 | 1.96e-05 |
| chr20:0.5-0.9Mb | 1 | -0,727776228 | Adipose_Subcutaneous | gr1 | SLC52A3 | 0.03398 | 8.95e-05 |

**Suppelemntary Table 2. Group-specific Genes for CAD cases clustering in Liver.**

Using the clustering structure of CAD cases from UKBB found in liver, the table reports genes that are significant (FDR 0.01) in at least one group across all tissues, tested via Wilcoxon-Mann-Whitney (WMW) for single a group against remaining cases after normalization. Significance is assessed after tissue-specific and group-specific multiple tested correction via Benjamini-Hochberg procedure. For each tissue, genes are combined into loci based on their genomic position (distance 1Mb of enlarged TSS window ± 200kb). For each group of genes in a tissue-specific locus, the table shows Z-statistic of the most significant gene for CAD, the groups with significantly different distribution in at least one gene, the common gene across groups with strongest association with a group distribution and the corresponding WMW estimate and p-value.

| UKBiobank Field | UKBiobank Meaning | UKBiobank Field class | Group-specific Test | Beta | SE beta | P-value | P-value BH corrected | Type phenotype | OR or Beta | Conf. Int. (lower) | Conf. Int. (upper) |
|---|---|---|---|---|---|---|---|---|---|---|---|
| Apolipoprotein B | NA | Blood_biochemistry | gr1_vs_all | 7.90e-02 | 1.41e-02 | 2.37e-08 | 3.91e-06 | CONTINUOUS | 7.90e-02 | 5.13e-02 | 1.07e-01 |
| Aspartate aminotransferase | NA | Blood_biochemistry | gr1_vs_all | 6.27e-02 | 1.68e-02 | 1.90e-04 | 8.66e-03 | CONTINUOUS | 6.27e-02 | 2.98e-02 | 9.57e-02 |
| White blood cell (leukocyte) count | NA | Blood_count | gr1_vs_all | 6.23e-02 | 1.68e-02 | 2.00e-04 | 8.66e-03 | CONTINUOUS | 6.23e-02 | 2.95e-02 | 9.52e-02 |
| Lymphocyte count | NA | Blood_count | gr1_vs_all | 6.20e-02 | 1.67e-02 | 2.10e-04 | 8.66e-03 | CONTINUOUS | 6.20e-02 | 2.92e-02 | 9.47e-02 |
| LDL direct | NA | Blood_biochemistry | gr1_vs_all | 4.57e-02 | 1.31e-02 | 4.99e-04 | 1.65e-02 | CONTINUOUS | 4.57e-02 | 2.00e-02 | 7.14e-02 |
| Cholesterol | NA | Blood_biochemistry | gr1_vs_all | 4.44e-02 | 1.32e-02 | 7.64e-04 | 2.10e-02 | CONTINUOUS | 4.44e-02 | 1.85e-02 | 7.02e-02 |
| Eosinophill count | NA | Blood_count | gr1_vs_all | 1.07e-01 | 3.24e-02 | 9.86e-04 | 2.32e-02 | CAT_ORD | 1.11e+00 | 1.04e+00 | 1.19e+00 |
| Medication for cholesterol, blood pressure, diabetes, or take exogenous hormones | None of the above | Medication | gr1_vs_all | -2.52e-01 | 8.10e-02 | 1.85e-03 | 3.82e-02 | CAT_MUL_BINARY_VAR | 7.77e-01 | 6.63e-01 | 9.11e-01 |
| Medication for cholesterol, blood pressure, diabetes, or take exogenous hormones | Cholesterol lowering medication | Medication | gr1_vs_all | 2.24e-01 | 7.30e-02 | 2.16e-03 | 3.96e-02 | CAT_MUL_BINARY_VAR | 1.25e+00 | 1.08e+00 | 1.44e+00 |
| Basophill count | NA | Blood_count | gr1_vs_all | 9.62e-02 | 3.22e-02 | 2.78e-03 | 4.58e-02 | CAT_ORD | 1.10e+00 | 1.03e+00 | 1.17e+00 |
| Urate | NA | Blood_biochemistry | gr1_vs_all | -4.51e-02 | 1.59e-02 | 4.50e-03 | 6.74e-02 | CONTINUOUS | -4.51e-02 | -7.63e-02 | -1.40e-02 |
| Mean sphered cell volume | NA | Blood_count | gr1_vs_all | -4.76e-02 | 1.70e-02 | 5.04e-03 | 6.93e-02 | CONTINUOUS | -4.76e-02 | -8.08e-02 | -1.43e-02 |
| Illnesses of siblings | Heart disease | Family_history | gr1_vs_all | 1.20e-01 | 4.44e-02 | 7.08e-03 | 8.99e-02 | CAT_MUL_BINARY_VAR | 1.13e+00 | 1.03e+00 | 1.23e+00 |
| Diagnoses - ICD10 | I20 Angina pectoris | ICD9-10_OPCS4 | gr1_vs_all | 8.67e-02 | 3.34e-02 | 9.42e-03 | 1.11e-01 | CAT_SINGLE_UNORDERED | 1.09e+00 | 1.02e+00 | 1.16e+00 |
| Apolipoprotein B | NA | Blood_biochemistry | gr2_vs_all | 1.05e-01 | 1.81e-02 | 6.92e-09 | 1.14e-06 | CONTINUOUS | 1.05e-01 | 6.92e-02 | 1.40e-01 |
| LDL direct | NA | Blood_biochemistry | gr2_vs_all | 7.64e-02 | 1.68e-02 | 5.19e-06 | 4.28e-04 | CONTINUOUS | 7.64e-02 | 4.36e-02 | 1.09e-01 |
| Eosinophill count | NA | Blood_count | gr2_vs_all | -1.48e-01 | 4.11e-02 | 3.24e-04 | 1.78e-02 | CAT_ORD | 8.63e-01 | 7.96e-01 | 9.35e-01 |
| Types of transport used (excluding work) | Walk | Physical_activity | gr2_vs_all | -1.46e-01 | 4.37e-02 | 8.25e-04 | 3.40e-02 | CAT_MUL_BINARY_VAR | 8.64e-01 | 7.93e-01 | 9.41e-01 |
| Cholesterol | NA | Blood_biochemistry | gr2_vs_all | 5.50e-02 | 1.69e-02 | 1.11e-03 | 3.66e-02 | CONTINUOUS | 5.50e-02 | 2.20e-02 | 8.81e-02 |
| Aspartate aminotransferase | NA | Blood_biochemistry | gr2_vs_all | -6.78e-02 | 2.15e-02 | 1.60e-03 | 4.39e-02 | CONTINUOUS | -6.78e-02 | -1.10e-01 | -2.57e-02 |
| Medication for pain relief, constipation, heartburn | Omeprazole (e.g. Zanprol) | Medication | gr2_vs_all | 1.83e-01 | 5.88e-02 | 1.87e-03 | 4.41e-02 | CAT_MUL_BINARY_VAR | 1.20e+00 | 1.07e+00 | 1.35e+00 |
| Reticulocyte count | NA | Blood_count | gr2_vs_all | -6.43e-02 | 2.13e-02 | 2.58e-03 | 5.04e-02 | CONTINUOUS | -6.43e-02 | -1.06e-01 | -2.25e-02 |
| Haemoglobin concentration | NA | Blood_count | gr2_vs_all | -5.67e-02 | 1.89e-02 | 2.75e-03 | 5.04e-02 | CONTINUOUS | -5.67e-02 | -9.38e-02 | -1.96e-02 |

| | | | | | | | | | | | |
|---|---|---|---|---|---|---|---|---|---|---|---|
| Eosinophill percentage | NA | Blood_count | gr2_vs_all | -6.32e-02 | 2.15e-02 | 3.33e-03 | 5.50e-02 | CONTINUOUS | -6.32e-02 | -1.05e-01 | -2.10e-02 |
| Alkaline phosphatase | NA | Blood_biochemistry | gr2_vs_all | -6.04e-02 | 2.15e-02 | 4.97e-03 | 7.46e-02 | CONTINUOUS | -6.04e-02 | -1.02e-01 | -1.83e-02 |
| Reticulocyte percentage | NA | Blood_count | gr2_vs_all | -5.84e-02 | 2.14e-02 | 6.32e-03 | 8.69e-02 | CONTINUOUS | -5.84e-02 | -1.00e-01 | -1.65e-02 |
| Haematocrit percentage | NA | Blood_count | gr2_vs_all | -5.18e-02 | 1.93e-02 | 7.35e-03 | 8.98e-02 | CONTINUOUS | -5.18e-02 | -8.96e-02 | -1.39e-02 |
| Diagnoses - ICD10 | I20 Angina pectoris | ICD9-10_OPCS4 | gr2_vs_all | 1.14e-01 | 4.27e-02 | 7.62e-03 | 8.98e-02 | CAT_SINGLE_UNORDERED | 1.12e+00 | 1.03e+00 | 1.22e+00 |
| Neutrophill percentage | NA | Blood_count | gr2_vs_all | 5.61e-02 | 2.14e-02 | 8.74e-03 | 9.61e-02 | CONTINUOUS | 5.61e-02 | 1.42e-02 | 9.81e-02 |
| Apolipoprotein B | NA | Blood_biochemistry | gr3_vs_all | -1.28e-01 | 1.33e-02 | 5.58e-22 | 9.21e-20 | CONTINUOUS | -1.28e-01 | -1.54e-01 | -1.02e-01 |
| LDL direct | NA | Blood_biochemistry | gr3_vs_all | -7.40e-02 | 1.23e-02 | 2.13e-09 | 1.76e-07 | CONTINUOUS | -7.40e-02 | -9.82e-02 | -4.98e-02 |
| Cholesterol | NA | Blood_biochemistry | gr3_vs_all | -5.34e-02 | 1.24e-02 | 1.69e-05 | 9.31e-04 | CONTINUOUS | -5.34e-02 | -7.78e-02 | -2.91e-02 |
| Apolipoprotein A | NA | Blood_biochemistry | gr3_vs_all | 6.50e-02 | 1.57e-02 | 3.48e-05 | 1.44e-03 | CONTINUOUS | 6.50e-02 | 3.43e-02 | 9.58e-02 |
| Medication for cholesterol, blood pressure or diabetes | Cholesterol lowering medication | Medication | gr3_vs_all | -1.35e-01 | 3.89e-02 | 5.42e-04 | 1.79e-02 | CAT_MUL_BINARY_VAR | 8.74e-01 | 8.10e-01 | 9.43e-01 |
| HDL cholesterol | NA | Blood_biochemistry | gr3_vs_all | 4.22e-02 | 1.54e-02 | 6.33e-03 | 1.74e-01 | CONTINUOUS | 4.22e-02 | 1.19e-02 | 7.25e-02 |
| Apolipoprotein B | NA | Blood_biochemistry | gr4_vs_all | 6.55e-02 | 1.45e-02 | 6.50e-06 | 1.07e-03 | CONTINUOUS | 6.55e-02 | 3.70e-02 | 9.39e-02 |
| Apolipoprotein A | NA | Blood_biochemistry | gr4_vs_all | -6.36e-02 | 1.71e-02 | 1.97e-04 | 1.63e-02 | CONTINUOUS | -6.36e-02 | -9.71e-02 | -3.01e-02 |
| Medication for pain relief, constipation, heartburn | Paracetamol | Medication | gr4_vs_all | -1.41e-01 | 4.26e-02 | 9.68e-04 | 4.09e-02 | CAT_MUL_BINARY_VAR | 8.69e-01 | 7.99e-01 | 9.44e-01 |
| Medication for cholesterol, blood pressure or diabetes | Cholesterol lowering medication | Medication | gr4_vs_all | 1.42e-01 | 4.32e-02 | 9.92e-04 | 4.09e-02 | CAT_MUL_BINARY_VAR | 1.15e+00 | 1.06e+00 | 1.25e+00 |
| C-reactive protein | NA | Blood_biochemistry | gr4_vs_all | -5.47e-02 | 1.71e-02 | 1.42e-03 | 4.69e-02 | CONTINUOUS | -5.47e-02 | -8.82e-02 | -2.11e-02 |
| Non-oily fish intake | NA | Diet | gr4_vs_all | -9.46e-02 | 3.27e-02 | 3.78e-03 | 1.04e-01 | CAT_ORD | 9.10e-01 | 8.53e-01 | 9.70e-01 |
| LDL direct | NA | Blood_biochemistry | gr4_vs_all | 3.76e-02 | 1.35e-02 | 5.21e-03 | 1.23e-01 | CONTINUOUS | 3.76e-02 | 1.12e-02 | 6.40e-02 |
| HDL cholesterol | NA | Blood_biochemistry | gr4_vs_all | -4.43e-02 | 1.68e-02 | 8.33e-03 | 1.72e-01 | CONTINUOUS | -4.43e-02 | -7.73e-02 | -1.14e-02 |
| Apolipoprotein B | NA | Blood_biochemistry | gr5_vs_all | -2.92e-01 | 3.10e-02 | 4.19e-21 | 6.92e-19 | CONTINUOUS | -2.92e-01 | -3.53e-01 | -2.32e-01 |
| LDL direct | NA | Blood_biochemistry | gr5_vs_all | -2.16e-01 | 2.88e-02 | 6.33e-14 | 5.22e-12 | CONTINUOUS | -2.16e-01 | -2.73e-01 | -1.60e-01 |
| Cholesterol | NA | Blood_biochemistry | gr5_vs_all | -2.00e-01 | 2.90e-02 | 5.11e-12 | 2.81e-10 | CONTINUOUS | -2.00e-01 | -2.57e-01 | -1.43e-01 |
| Diagnoses - ICD10 | E78 Disorders of lipoprotein metabolism and other lipidaemias | ICD9-10_OPCS4 | gr5_vs_all | -2.87e-01 | 7.34e-02 | 9.46e-05 | 3.90e-03 | CAT_SINGLE_UNORDERED | 7.51e-01 | 6.50e-01 | 8.67e-01 |
| Medication for cholesterol, blood pressure, diabetes, or take exogenous hormones | Cholesterol lowering medication | Medication | gr5_vs_all | -5.83e-01 | 1.56e-01 | 1.82e-04 | 6.02e-03 | CAT_MUL_BINARY_VAR | 5.58e-01 | 4.11e-01 | 7.58e-01 |
| Mean sphered cell volume | NA | Blood_count | gr5_vs_all | 1.33e-01 | 3.76e-02 | 4.00e-04 | 1.10e-02 | CONTINUOUS | 1.33e-01 | 5.94e-02 | 2.07e-01 |
| Medication for cholesterol, blood pressure, | None of the above | Medication | gr5_vs_all | 5.33e-01 | 1.56e-01 | 6.36e-04 | 1.50e-02 | CAT_MUL_BIN | 1.70e+00 | 1.26e+00 | 2.31e+00 |

| | | | | | | | | | | | |
|---|---|---|---|---|---|---|---|---|---|---|---|
| diabetes, or take exogenous hormones | | | | | | | | ARY_VAR | | | |
| Direct bilirubin | NA | Blood_biochemistry | gr5_vs_all | 1.21e-01 | 3.77e-02 | 1.34e-03 | 2.77e-02 | CONTINUOUS | 1.21e-01 | 4.70e-02 | 1.95e-01 |
| Medication for cholesterol, blood pressure or diabetes | Cholesterol lowering medication | Medication | gr5_vs_all | -2.80e-01 | 8.97e-02 | 1.77e-03 | 3.24e-02 | CAT_MUL_BINARY_VAR | 7.55e-01 | 6.34e-01 | 9.01e-01 |
| Diagnoses - ICD10 | I26 Pulmonary embolism | ICD9-10_OPCS4 | gr5_vs_all | 5.62e-01 | 1.93e-01 | 3.61e-03 | 5.96e-02 | CAT_SINGLE_UNORDERED | 1.75e+00 | 1.20e+00 | 2.56e+00 |
| Illnesses of siblings | High blood pressure | Family_history | gr5_vs_all | -2.76e-01 | 9.87e-02 | 5.10e-03 | 7.52e-02 | CAT_MUL_BINARY_VAR | 7.59e-01 | 6.25e-01 | 9.20e-01 |
| Eosinophill percentage | NA | Blood_count | gr5_vs_all | -1.03e-01 | 3.74e-02 | 5.88e-03 | 7.52e-02 | CONTINUOUS | -1.03e-01 | -1.76e-01 | -2.97e-02 |
| Diagnoses - ICD10 | I20 Angina pectoris | ICD9-10_OPCS4 | gr5_vs_all | -2.02e-01 | 7.34e-02 | 5.92e-03 | 7.52e-02 | CAT_SINGLE_UNORDERED | 8.17e-01 | 7.08e-01 | 9.44e-01 |
| Medication for pain relief, constipation, heartburn | Aspirin | Medication | gr5_vs_all | -2.03e-01 | 7.56e-02 | 7.15e-03 | 7.98e-02 | CAT_MUL_BINARY_VAR | 8.16e-01 | 7.03e-01 | 9.46e-01 |
| Albumin | NA | Blood_biochemistry | gr5_vs_all | -1.03e-01 | 3.84e-02 | 7.25e-03 | 7.98e-02 | CONTINUOUS | -1.03e-01 | -1.79e-01 | -2.79e-02 |

**Supplementary Table 3. Differences in endophenotypes for groups of CAD cases in Liver.** Group-specific endophenotype analysis using clustering of CAD cases in liver from UKBB with nominal p-value < 0.01. The tested endophenotypes are CAD related classes from UKBB. Differences are tested via Generalized Linear Model (GLM) with phenotype the dependent variable and group-specific clustering structure ($gr_i$ versus remaining cases) and covariates the independent variables. The regression coefficient $\boldsymbol{\beta}$ estimate, standard error and p-value refer to the grouping variable, estimates are corrected for multiple testing in a group-specific manner using Benjamini-Hochberg procedure. The family for GLM applied depends on the phenotype nature (continuous, binary or categorical ordinal). For non-continuous ones it is shown odds ratio and the corresponding 95% confidence interval, for continuous ones instead confidence intervals refer to $\boldsymbol{\beta}$ estimates.

| Dataset | ID | Group-specific Test | Beta | SE Beta | P-value | Type phenotype | OR or Beta | Conf. Int. (lower) | Conf. Int. (upper) | empirical P-value |
|---|---|---|---|---|---|---|---|---|---|---|
| GerMIFSV | Vessel_affected | gr1_vs_all | 0,228243267 | 0,088006103 | 0,009559527 | CAT_ORD | 1,256390927 | 1,057338048 | 1,492917203 | NA |
| UK Biobank | Chronic_kidney_disease | gr1_vs_all | -0,119178911 | 0,064234066 | 0,063541295 | CAT_SINGLE_UNORDERED | 0,887648977 | 0,78264555 | 1,006740159 | 0,012 |
| UK Biobank | Chronic_obstructive_pulmonary_disease | gr1_vs_all | -0,101587954 | 0,056233154 | 0,070832608 | CAT_SINGLE_UNORDERED | 0,903401718 | 0,809124152 | 1,008664321 | 0,004 |
| UK Biobank | Coronary_artery_bypass_graft | gr1_vs_all | 0,070257944 | 0,042418882 | 0,097663229 | CAT_SINGLE_UNORDERED | 1,072784864 | 0,987201075 | 1,16578 82 | 0,008 |
| UK Biobank | Smoking | gr2_vs_all | -0,130487812 | 0,046403812 | 0,004923354 | CAT_SINGLE_UNORDERED | 0,87766719 | 0,801365916 | 0,961233415 | 0,016 |
| UK Biobank | UAP | gr2_vs_all | 0,139843208 | 0,053370622 | 0,008787095 | CAT_SINGLE_UNORDERED | 1,150093459 | 1,035866952 | 1,276915883 | 0,004 |
| UK Biobank | Hyperlipidemia | gr2_vs_all | 0,095132326 | 0,042872399 | 0,026489149 | CAT_SINGLE_UNORDERED | 1,099804379 | 1,011165846 | 1,196212942 | 0,008 |
| UK Biobank | Acute_MI | gr2_vs_all | -0,078367963 | 0,043453883 | 0,071313752 | CAT_SINGLE_UNORDERED | 0,924624136 | 0,849135922 | 1,006823256 | 0,008 |
| UK Biobank | Cerebrovascular_disease | gr2_vs_all | 0,519833253 | 0,291991541 | 0,075026478 | CAT_SINGLE_UNORDERED | 1,6817472 | 0,948892607 | 2,980604574 | 0 |
| UK Biobank | Age_stroke | gr2_vs_all | 1,158971426 | 0,669161436 | 0,083608026 | CONTINUOUS | 1,158971426 | -0,15256089 | 2,470503741 | 0,004 |
| UK Biobank | Age_stroke | gr3_vs_all | -1,332380178 | 0,495304904 | 0,00727224 | CONTINUOUS | -1,332380178 | -2,303159951 | -0,361600404 | 0,004 |
| UK Biobank | Chronic_obstructive_pulmonary_disease | gr3_vs_all | 0,131308416 | 0,050647758 | 0,009525873 | CAT_SINGLE_UNORDERED | 1,14031942 | 1,032559463 | 1,25932542 | 0,004 |
| UK Biobank | Hyperlipidemia | gr3_vs_all | -0,080401614 | 0,031342123 | 0,01030898 | CAT_SINGLE_UNORDERED | 0,922745684 | 0,867767856 | 0,981206658 | 0,008 |
| UK Biobank | Peripheral_vascular_disease | gr3_vs_all | -0,260812053 | 0,119400731 | 0,028937114 | CAT_SINGLE_UNORDERED | 0,770425705 | 0,6096726 | 0,973564773 | 0,008 |
| UK Biobank | Age_heart_attack | gr3_vs_all | 0,335557819 | 0,178078086 | 0,059557749 | CONTINUOUS | 0,335557819 | -0,013468817 | 0,684584455 | 0,004 |
| UK Biobank | Smoking | gr3_vs_all | 0,064641379 | 0,03497599 | 0,064578568 | CAT_SINGLE_UNORDERED | 1,066776387 | 0,996097333 | 1,142470543 | 0,016 |
| UK Biobank | Coronary_artery_bypass_graft | gr3_vs_all | -0,072233624 | 0,040553764 | 0,074882763 | CAT_SINGLE_UNORDERED | 0,930313527 | 0,859230944 | 1,007276639 | 0,008 |
| GerMIFSV | Gensini_score | gr3_vs_all | -0,07311755 | 0,042976882 | 0,089013944 | CONTINUOUS | -0,07311755 | -0,15735069 | 0,011115591 | NA |
| UK Biobank | Peripheral_vascular_disease | gr4_vs_all | 0,38164915 | 0,114251454 | 0,000836483 | CAT_SINGLE_UNORDERED | 1,464698106 | 1,1708388 | 1,832310769 | 0,008 |
| UK Biobank | Age_stroke | gr4_vs_all | 1,183384837 | 0,52952493 | 0,025665976 | CONTINUOUS | 1,183384837 | 0,145535046 | 2,221234628 | 0,004 |
| UK Biobank | Poor_mobility | gr4_vs_all | -0,698973328 | 0,340657765 | 0,040185757 | CAT_SINGLE_UNORDERED | 0,497095396 | 0,254959575 | 0,969188284 | 0,012 |
| UK Biobank | UAP | gr4_vs_all | -0,090608124 | 0,044902955 | 0,043605271 | CAT_SINGLE_UNORDERED | 0,913375571 | 0,836426776 | 0,997403428 | 0,004 |
| UK Biobank | Age_heart_attack | gr4_vs_all | -0,359367995 | 0,195850066 | 0,066556719 | CONTINUOUS | -0,359367995 | -0,743227071 | 0,02449108 | 0,004 |
| UK Biobank | Coronary_artery_bypass_graft | gr4_vs_all | 0,072804504 | 0,043416156 | 0,093562253 | CAT_SINGLE_UNORDERED | 1,075520257 | 0,987785611 | 1,171047452 | 0,008 |
| UK Biobank | Age_angina | gr4_vs_all | -0,322694619 | 0,19583605 | 0,099446041 | CONTINUOUS | -0,322694619 | -0,706526223 | 0,061136986 | 0,008 |
| UK Biobank | Hyperlipidemia | gr5_vs_all | -0,286708045 | 0,073436728 | 9,46E-05 | CAT_SINGLE_UNORDERED | 0,750730876 | 0,650092 | 0,866949368 | 0,008 |
| UK Biobank | Coronary_artery_bypass_graft | gr5_vs_all | -0,199357064 | 0,100164947 | 0,046559249 | CAT_SINGLE_UNORDERED | 0,819257314 | 0,673224279 | 0,996967232 | 0,008 |

| | | | | | | | | | | |
|---|---|---|---|---|---|---|---|---|---|---|
| UK Biobank | UAP | gr5_vs_all | -0,188267582 | 0,100829004 | 0,061874126 | CAT_SINGLE_UNORDERED | 0,828393015 | 0,679846122 | 1,009397516 | 0,004 |
| UK Biobank | T1D | gr5_vs_all | 0,317530053 | 0,183017205 | 0,082745448 | CAT_SINGLE_UNORDERED | 1,373730529 | 0,959659182 | 1,966464349 | 0 |
| UK Biobank | Transient_cerebral_ischaemic_attacks | gr5_vs_all | -0,516004502 | 0,308035442 | 0,093905486 | CAT_SINGLE_UNORDERED | 0,596900705 | 0,3263637 | 1,091697551 | 0,004 |

**Supplementary Table 4. Hypothesis-driven endophenotype differences for CAD cases clustering in Liver.**

Group-specific clinical variables analysis using clustering of CAD cases in liver with nominal p-value < 0.1. For UKBB dataset, clustering of CAD cases in UKBB is considered and 33 clinical variables in UKBB are used without PHESANT preprocessing. For GerMIFSV dataset, clustering structure is obtained via projection from UKBB model clustering and 2 severeness index annotated for that dataset are tested. Differences are tested via Generalized Linear Model (GLM) with phenotype the dependent variable and group-specific clustering structure ($gr_i$ versus remaining cases) and covariates the independent variables. The regression coefficient $\boldsymbol{\beta}$ estimate, standard error and p-value refer to the grouping variable. The family for GLM applied depends on the phenotype nature (continuous, binary or categorical ordinal). For non-continuous ones it is shown odds ratio and the corresponding 95% confidence interval, for continuous ones instead confidence intervals refer to $\boldsymbol{\beta}$ estimates. Empirical p-value are computed for UKBB variables comparing to endophenotype associations of random clustering repetitions.

| Loci | N. genes | SCZ best Z-statistic | Tissue | Group-specific differences | Best group-specific gene | WMW estimate best gene | WMW p-value best gene |
|---|---|---|---|---|---|---|---|
| chr1:27.5-27.9Mb | 1 | 0,60937257 | Brain_Cerebellar_Hemisphere | gr1 | CD164L2 | 0.00634 | 1.28e-06 |
| chr1:173.4-173.8Mb | 1 | 4,119438257 | Brain_Nucleus_accumbens_basal_ganglia | gr2 | ANKRD45 | 0.00336 | 8.29e-05 |
| chr2:47.4-47.8Mb | 1 | -1,839532387 | DLPC_CMC | gr2 | MSH2 | 0.00784 | 8.23e-05 |
| chr2:71.2-71.6Mb | 1 | 0,352047327 | DLPC_CMC | gr2,gr3 | MPHOSPH10 | -0.01966,0.01665 | 2.85e-07,1.47e-05 |
| chr2:74.9-75.3Mb | 1 | -0,396267195 | Brain_Cerebellar_Hemisphere | gr1 | AC104135.3 | 0.06489 | 9.91e-05 |
| chr2:74.9-75.3Mb | 1 | -0,283647674 | Brain_Frontal_Cortex_BA9 | gr1 | AC104135.3 | 0.06382 | 0.000185 |
| chr2:74.9-75.4Mb | 3 | 0,412348847 | Cells_EBV-transformed_lymphocytes | gr1 | AC104135.2 | 0.06891 | 3.77e-05 |
| chr2:75-75.4Mb | 1 | -0,049025121 | Brain_Caudate_basal_ganglia | gr1 | AC104135.4 | 0.06252 | 0.000142 |
| chr2:135.6-136.5Mb | 2 | -0,94314726 | DLPC_CMC | gr1 | R3HDM1 | 0.01065 | 2.05e-06 |
| chr2:224.6-225Mb | 1 | -1,095143595 | DLPC_CMC | gr3 | WDFY1 | -0.01318 | 6.19e-08 |
| chr2:224.6-225Mb | 1 | -0,52061301 | Brain_Frontal_Cortex_BA9 | gr3 | WDFY1 | -0.0439 | 5.89e-05 |
| chr2:224.6-225Mb | 1 | 1,048818956 | Brain_Caudate_basal_ganglia | gr3 | AC073641.2 | 0.0565 | 3.87e-05 |
| chr2:224.6-225Mb | 1 | 1,194117805 | Cells_EBV-transformed_lymphocytes | gr3 | AC073641.2 | 0.05246 | 0.000183 |
| chr3:52.2-52.6Mb | 1 | -0,202460363 | Brain_Cerebellum | gr1 | DNAH1 | 0.01081 | 0.000164 |
| chr3:110.6-111Mb | 1 | -1,387956414 | DLPC_CMC | gr1 | PVRL3 | 0.00597 | 2.43e-10 |
| chr3:159.9-160.3Mb | 1 | 0,804017528 | Cells_EBV-transformed_lymphocytes | gr1 | SMC4 | -0.0169 | 9.73e-05 |
| chr6:24.4-34Mb | 84 | 9,11792427 | DLPC_CMC | gr1,gr2,gr3 | C4A | -2.31284,0.17878,0.14103 | 0,7.58e-269,8.64e-177 |
| chr6:24.8-26.9Mb | 8 | 7,492574253 | Cells_EBV-transformed_lymphocytes | gr1,gr2,gr3 | BTN3A2 | -0.67175,0.09297,0.09251 | 0,3.02e-71,1.12e-66 |
| chr6:24.9-33.7Mb | 44 | -8,509690388 | Brain_Cortex | gr1,gr2,gr3 | NOTCH4 | -1.91242,0.31281,0.28497 | 0,8.54e-243,4.19e-221 |
| chr6:25-34Mb | 50 | -8,765027816 | Brain_Caudate_basal_ganglia | gr1,gr2,gr3 | IER3 | 2.10795,-0.08578,-0.08574 | 0,5.48e-216,1.74e-209 |
| chr6:25.8-34Mb | 65 | 9,15699547 | Brain_Cerebellar_Hemisphere | gr1,gr2,gr3 | C4A | -2.20347,0.24164,0.27126 | 0,3e-219,8.82e-292 |
| chr6:25.8-34Mb | 84 | 8,567758457 | Brain_Cerebellum | gr1,gr2,gr3 | C4A | -2.0414,0.35472,0.33328 | 0,4.58e-267,2.43e-248 |
| chr6:25.8-28.5Mb | 11 | -7,45033741 | Brain_Nucleus_accumbens_basal_ganglia | gr1,gr2,gr3 | RP1-265C24.5 | -1.54759,-0.12148,1.60438 | 0,0,0 |
| chr6:25.8-33.6Mb | 44 | -9,091302526 | Brain_Frontal_Cortex_BA9 | gr1,gr2,gr3 | HLA-DMA | 1.7991,-0.17079,-0.14477 | 0,1.66e-159,1.1e-115 |
| chr6:25.9-34Mb | 43 | 9,425298026 | Brain_Hypothalamus | gr1,gr2,gr3 | NCR3 | 1.93175,-0.12358,-0.09018 | 0,2.71e-253,3.42e-155 |
| chr6:26-28.5Mb | 8 | -7,548006867 | Brain_Hippocampus | gr1,gr2,gr3 | AL022393.7 | 0.76927,1.15978,-1.59026 | 0,0,0 |
| chr6:27.9-34Mb | 51 | 9,140860601 | Cells_EBV-transformed_lymphocytes | gr1,gr2,gr3 | C4A | -2.31136,0.18873,0.20019 | 0,3.3e-236,1.9e-287 |
| chr6:29.6-33.6Mb | 26 | 9,479578005 | Brain_Hippocampus | gr1,gr2,gr3 | CYP21A1P | -2.1219,0.2312,0.26086 | 0,2.6e-215,8.8e-280 |
| chr6:29.6-33.6Mb | 31 | -8,516985031 | Brain_Nucleus_accumbens_basal_ganglia | gr1,gr2,gr3 | XXbac-BPG300A18.13 | 1.85146,-0.37392,-0.31261 | 0,1.39e-252,3.66e-189 |
| chr6:83.7-84.1Mb | 1 | -2,688743063 | Brain_Cerebellar_Hemisphere | gr2 | RWDD2A | -0.00957 | 4.68e-05 |
| chr7:5.4-5.8Mb | 1 | 1,539685934 | Brain_Hippocampus | gr3 | FSCN1 | -0.05445 | 8.72e-05 |
| chr7:91.7-92.1Mb | 1 | -0,96554596 | Brain_Cerebellum | gr1,gr3 | KRIT1 | 0.00678,-0.00481 | 7.45e-07,1.22e-05 |
| chr9:88.2-88.6Mb | 1 | -0,017199499 | DLPC_CMC | gr3 | RP11-213G2.3 | 0.01069 | 3.29e-07 |
| chr10:97.2-97.6Mb | 1 | -1,618003931 | Brain_Cerebellum | gr1 | ALDH18A1 | 0.07352 | 2.6e-05 |
| chr11:61-61.4Mb | 1 | 0,215306172 | DLPC_CMC | gr1 | TMEM216 | -0.00411 | 2.89e-05 |
| chr11:73.3-73.7Mb | 1 | 2,671221059 | Brain_Cerebellar_Hemisphere | gr2 | MRPL48 | 0.00619 | 0.000106 |
| chr12:56.5-56.9Mb | 1 | 1,371344018 | Brain_Hypothalamus | gr1 | RP11-977G19.11 | 0.00557 | 8.43e-05 |
| chr12:104.1-104.5Mb | 1 | -1,360871384 | Brain_Cortex | gr1 | MIR3652 | -0.00802 | 6.05e-05 |

| | | | | | | | |
|---|---|---|---|---|---|---|---|
| chr12:132.9-133.3Mb | 1 | -1,001149867 | Brain_Cerebellar_Hemisphere | gr1 | FBRSL1 | 0.01188 | 0.000132 |
| chr13:20-20.4Mb | 1 | 0,868280955 | Cells_EBV-transformed_lymphocytes | gr1 | MPHOSPH8 | 0.0077 | 5.07e-05 |
| chr13:24.3-24.7Mb | 1 | 1,28914845 | Brain_Cerebellar_Hemisphere | gr1 | MIPEP | 0.05857 | 9.67e-05 |
| chr13:79.8-80.2Mb | 1 | -0,325824917 | Brain_Cerebellum | gr2 | RBM26-AS1 | 0.00716 | 0.000118 |
| chr14:50.7-51.1Mb | 1 | -0,247051358 | Brain_Cerebellum | gr2 | CDKL1 | 0.03716 | 0.000197 |
| chr15:65.5-65.9Mb | 1 | -1,264254803 | DLPC_CMC | gr2 | IGDCC4 | 0.0499 | 9.84e-05 |
| chr16:30.2-30.6Mb | 1 | -2,104837054 | Brain_Hypothalamus | gr3 | ZNF48 | 0.01108 | 7.79e-05 |
| chr16:67.3-67.7Mb | 1 | -2,598944984 | Brain_Hippocampus | gr1 | HSD11B2 | -0.00499 | 0.000204 |
| chr16:77-77.4Mb | 1 | 0,330733192 | Brain_Cerebellum | gr2 | SYCE1L | 0.05207 | 0.000219 |
| chr16:77-77.4Mb | 1 | -0,159949579 | Brain_Frontal_Cortex_BA9 | gr2 | SYCE1L | 0.01571 | 0.000203 |
| chr17:18.9-19.3Mb | 1 | 1,04212852 | DLPC_CMC | gr2 | SNORD3A | 0.00429 | 5.95e-05 |
| chr19:2-2.4Mb | 1 | -0,841315918 | Brain_Nucleus_accumbens_basal_ganglia | gr1 | SF3A2 | -0.007 | 2.09e-14 |
| chr19:44.8-45.2Mb | 1 | 0,760791203 | Brain_Hypothalamus | gr2 | ZNF180 | -0.02512 | 0.000113 |
| chr21:47.4-47.8Mb | 1 | -1,891164482 | Brain_Nucleus_accumbens_basal_ganglia | gr2 | FTCD | -0.03959 | 5.26e-05 |
| chr21:47.5-47.9Mb | 1 | -1,110019041 | Brain_Cerebellum | gr2,gr3 | MCM3AP | -0.05892,0.05559 | 1.81e-05,5.48e-05 |
| chr22:21.8-22.2Mb | 1 | 2,21030452 | Cells_EBV-transformed_lymphocytes | gr2 | YDJC | 0.00947 | 3.52e-05 |
| chr22:22.1-22.5Mb | 1 | -1,136840537 | Brain_Cerebellar_Hemisphere | gr3 | PPM1F | 0.02444 | 0.000164 |

**Supplementary Table 5. Group-specific Genes for SCZ cases clustering in Dorsolateral prefrontal cortex.**

Using the clustering structure of SCZ cases found in DLPC, the table reports genes that are significant in at least one group across all tissues (FDR 0.01), tested via Wilcoxon-Mann-Whitney (WMW) for single a group against remaining cases after concatenation and normalization of 35 SCZ cohorts. Significance is assessed after tissue-specific and group-specific multiple tested correction via Benjamini-Hochberg procedure. For each tissue, genes are combined into loci based on their genomic position (distance 1Mb of enlarged TSS window ± 200kb). For each group of genes in a tissue-specific locus, the table shows Z-statistic of the most significant gene for SCZ, the groups with significantly different distribution in at least one gene, the common gene across groups with strongest association with a group distribution and the corresponding WMW estimate and p-value.

| Macro | UKBiobank Class | UKBiobank Field | UKBiobank Meaning | CRM | Group-specific Test | Beta | SE Beta | P-value | P-value BH corrected | Conf. Int. (lower) | Conf. Int. (upper) |
|---|---|---|---|---|---|---|---|---|---|---|---|
| | Blood_count | White blood cell (leukocyte) count | NA | 6215,1946 | gr1_vs_all | -0,4296964 | 0,016630125 | 4,12E-145 | 2,06E-142 | -0,462290845 | -0,397101955 |
| | Blood_count | Red blood cell (erythrocyte) count | NA | 966,045493 | gr1_vs_all | -0,070507801 | 0,016874932 | 2,95E-05 | 0,000109204 | -0,103582061 | -0,037433541 |
| | Blood_count | Haemoglobin concentration | NA | 2167,89944 | gr1_vs_all | -0,193736284 | 0,016826393 | 1,37E-30 | 1,37E-28 | -0,226715408 | -0,16075716 |
| | Blood_count | Haematocrit percentage | NA | 1664,15071 | gr1_vs_all | -0,157496525 | 0,016841135 | 9,38E-21 | 4,47E-19 | -0,190504542 | -0,124488507 |
| | Blood_count | Mean corpuscular volume | NA | 1389,44082 | gr1_vs_all | -0,073763607 | 0,016875434 | 1,24E-05 | 5,05E-05 | -0,10683885 | -0,040688364 |
| | Blood_count | Mean corpuscular haemoglobin | NA | 1941,67865 | gr1_vs_all | -0,103446856 | 0,016863294 | 8,69E-10 | 9,65E-09 | -0,136498305 | -0,070395408 |
| | Blood_count | Mean corpuscular haemoglobin concentration | NA | 891,882849 | gr1_vs_all | -0,135407535 | 0,016848677 | 9,68E-16 | 2,76E-14 | -0,168430334 | -0,102384735 |
| | Blood_count | Red blood cell (erythrocyte) distribution width | NA | 2960,83729 | gr1_vs_all | 0,165192699 | 0,016838319 | 1,13E-22 | 5,63E-21 | 0,1321902 | 0,198195198 |
| | Blood_count | Platelet count | NA | 3413,39561 | gr1_vs_all | -0,183125728 | 0,016826197 | 1,62E-27 | 1,08E-25 | -0,216104469 | -0,150146987 |
| | Blood_count | Platelet crit | NA | 3709,90552 | gr1_vs_all | -0,230277839 | 0,016805408 | 1,45E-42 | 1,81E-40 | -0,263215833 | -0,197339845 |
| | Blood_count | Platelet distribution width | NA | 1600,10493 | gr1_vs_all | 0,101245965 | 0,016876212 | 2,01E-09 | 2,05E-08 | 0,068169198 | 0,134322732 |
| | Blood_count | Lymphocyte count | NA | 6154,96497 | gr1_vs_all | -0,434440564 | 0,01661248 | 1,51E-148 | 1,51E-145 | -0,467000426 | -0,401880701 |
| | Blood_count | Monocyte count | NA | 3199,60529 | gr1_vs_all | -0,225478206 | 0,016816392 | 7,74E-41 | 8,60E-39 | -0,258437729 | -0,192518682 |
| | Blood_count | Neutrophill count | NA | 4303,24735 | gr1_vs_all | -0,31976882 | 0,01674706 | 1,22E-80 | 3,04E-78 | -0,352592455 | -0,286945185 |
| | Blood_count | Eosinophill count | NA | 3856,98493 | gr1_vs_all | -0,316434279 | 0,016739838 | 4,36E-79 | 8,73E-77 | -0,349243758 | -0,283624799 |
| | Blood_count | Basophill count | NA | 647,725412 | gr1_vs_all | -0,115564617 | 0,016849513 | 7,13E-12 | 1,15E-10 | -0,148589055 | -0,08254018 |
| | Blood_count | Lymphocyte percentage | NA | 1249,27198 | gr1_vs_all | -0,100746387 | 0,016865119 | 2,35E-09 | 2,31E-08 | -0,133801412 | -0,067691362 |
| | Blood_count | Monocyte percentage | NA | 1593,07132 | gr1_vs_all | 0,115599375 | 0,016852674 | 7,09E-12 | 1,15E-10 | 0,08256874 | 0,148630009 |
| | Blood_count | Neutrophill percentage | NA | 1054,03742 | gr1_vs_all | 0,089886155 | 0,016874367 | 1,01E-07 | 6,82E-07 | 0,056813003 | 0,122959308 |
| | Blood_count | Eosinophill percentage | NA | 2318,06347 | gr1_vs_all | -0,171396111 | 0,016817763 | 2,44E-24 | 1,44E-22 | -0,20435832 | -0,138433902 |
| | Blood_count | Reticulocyte percentage | NA | 2486,76083 | gr1_vs_all | -0,18362411 | 0,016838487 | 1,28E-27 | 9,12E-26 | -0,216626939 | -0,150621281 |
| | Blood_count | Reticulocyte count | NA | 2621,5937 | gr1_vs_all | -0,191921891 | 0,016837587 | 5,14E-30 | 4,67E-28 | -0,224922955 | -0,158920826 |
| | Blood_count | Mean reticulocyte volume | NA | 2879,2199 | gr1_vs_all | 0,190342318 | 0,016840995 | 1,53E-29 | 1,18E-27 | 0,157334574 | 0,223350062 |
| | Blood_count | Mean sphered | NA | 1382,1474 | gr1_vs_all | 0,096530276 | 0,016874651 | 1,08E-08 | 8,82E-08 | 0,063456569 | 0,129603983 |

| | | | | | | | | | | | |
|---|---|---|---|---|---|---|---|---|---|---|---|
| | | cell volume | | | | | | | | | |
| | Blood_count | High light scatter reticulocyte percentage | NA | 1636,95418 | gr1_vs_all | -0,116035625 | 0,016865369 | 6,14E-12 | 1,02E-10 | -0,149091141 | -0,082980109 |
| | Blood_count | High light scatter reticulocyte count | NA | 1794,91908 | gr1_vs_all | -0,126640176 | 0,016865127 | 6,18E-14 | 1,40E-12 | -0,159695219 | -0,093585134 |
| | Blood_count_ratio | Lymphocyte-to-Monocyte ratio | NA | 2320,66891 | gr1_vs_all | -0,167469414 | 0,016829559 | 2,79E-23 | 1,47E-21 | -0,200454743 | -0,134484085 |
| | Blood_count_ratio | Platelet-to-Lymphocyte ratio | NA | 4025,76637 | gr1_vs_all | 0,252102244 | 0,016795754 | 1,11E-50 | 1,58E-48 | 0,219183172 | 0,285021316 |
| | Blood_count_ratio | Neutrophill-to-Lymphocyte ratio | NA | 1197,18849 | gr1_vs_all | 0,098883997 | 0,016868862 | 4,64E-09 | 4,03E-08 | 0,065821634 | 0,13194636 |
| | Blood_count_ratio | Eosinophill-to-Lymphocyte ratio | NA | 1753,51356 | gr1_vs_all | -0,131459782 | 0,016834342 | 6,01E-15 | 1,62E-13 | -0,164454487 | -0,098465078 |
| | Blood_pressure | Diastolic blood pressure, automated reading | NA | 1566,79006 | gr1_vs_all | -0,167673583 | 0,016829832 | 2,47E-23 | 1,37E-21 | -0,200659447 | -0,134687719 |
| | Body_size_measures | Hip circumference | NA | 1485,29756 | gr1_vs_all | 0,122304872 | 0,01685701 | 4,13E-13 | 8,79E-12 | 0,089265738 | 0,155344005 |
| | Body_size_measures | Body mass index (BMI) | NA | 765,359357 | gr1_vs_all | 0,060823964 | 0,01686283 | 0,000310441 | 0,000929465 | 0,027773425 | 0,093874503 |
| | Body_size_measures | Weight | NA | 1833,73636 | gr1_vs_all | 0,126830998 | 0,016854639 | 5,47E-14 | 1,29E-12 | 0,093796512 | 0,159865484 |
| | Sleep | Sleep duration | NA | 711,344318 | gr1_vs_all | -0,115188714 | 0,016869119 | 8,80E-12 | 1,40E-10 | -0,14825158 | -0,082125848 |
| | Smoking | Past tobacco smoking | NA | 936,547806 | gr1_vs_all | 0,149693771 | 0,016858097 | 7,19E-19 | 2,77E-17 | 0,116652508 | 0,182735034 |
| | Smoking | Smoking status | Never | 858,691374 | gr1_vs_all | 0,12710887 | 0,016856707 | 4,86E-14 | 1,23E-12 | 0,094070331 | 0,160147409 |
| | Smoking | Smoking status | Previous | 647,211773 | gr1_vs_all | -0,120313346 | 0,016865828 | 1,01E-12 | 2,01E-11 | -0,153369761 | -0,087256931 |
| | Smoking | Ever smoked | NA | 844,809172 | gr1_vs_all | -0,141499279 | 0,016863366 | 5,10E-17 | 1,70E-15 | -0,17455087 | -0,108447688 |
| | Blood_biochemistry | Alanine aminotransferase | NA | 876,626767 | gr1_vs_all | -0,089627621 | 0,01688176 | 1,11E-07 | 7,32E-07 | -0,122715263 | -0,056539978 |
| | Blood_biochemistry | Apolipoprotein A | NA | 1693,68578 | gr1_vs_all | -0,141953307 | 0,016829421 | 3,51E-17 | 1,21E-15 | -0,174938365 | -0,108968248 |
| | Blood_biochemistry | Aspartate aminotransferase | NA | 3101,81243 | gr1_vs_all | -0,285435133 | 0,016768439 | 1,42E-64 | 2,36E-62 | -0,318300669 | -0,252569597 |
| | Blood_biochemistry | Calcium | NA | 924,234146 | gr1_vs_all | -0,105111813 | 0,01686928 | 4,72E-10 | 5,42E-09 | -0,138174994 | -0,072048632 |
| | Blood_biochemistry | Cholesterol | NA | 996,651892 | gr1_vs_all | -0,102662302 | 0,016871716 | 1,18E-09 | 1,29E-08 | -0,135730258 | -0,069594347 |
| | Blood_biochemistry | Creatinine | NA | 1874,36596 | gr1_vs_all | 0,139589639 | 0,016847108 | 1,24E-16 | 4,00E-15 | 0,106569915 | 0,172609364 |
| | Blood_biochemistry | C-reactive protein | NA | 1812,01963 | gr1_vs_all | -0,138810432 | 0,016859108 | 1,91E-16 | 5,98E-15 | -0,171853677 | -0,105767187 |
| | Blood_biochemistry | Cystatin C | NA | 3014,23785 | gr1_vs_all | 0,145591187 | 0,016843833 | 5,80E-18 | 2,15E-16 | 0,11257788 | 0,178604494 |
| | Blood_biochemistry | Gamma glutamyltransferase | NA | 1535,12266 | gr1_vs_all | -0,10608428 | 0,016874961 | 3,31E-10 | 3,93E-09 | -0,139158596 | -0,073009964 |
| | Blood_biochemistry | Glucose | NA | 727,808265 | gr1_vs_all | 0,105655085 | 0,016867578 | 3,82E-10 | 4,45E-09 | 0,07259524 | 0,13871493 |
| | Blood_biochemistry | Glycated haemoglobin (HbA1c) | NA | 1535,20329 | gr1_vs_all | 0,112719874 | 0,016861989 | 2,37E-11 | 3,48E-10 | 0,079670983 | 0,145768764 |
| | Blood_biochemistry | HDL cholesterol | NA | 1531,47566 | gr1_vs_all | -0,114480876 | 0,016845384 | 1,10E-11 | 1,70E-10 | -0,147497221 | -0,08146453 |

| | | | | | | | | | | | |
|---|---|---|---|---|---|---|---|---|---|---|---|
| | Blood_biochemistry | IGF-1 | NA | 2192,81323 | gr1_vs_all | -0,156108828 | 0,016835747 | 1,98E-20 | 8,24E-19 | -0,189106285 | -0,123111371 |
| | Blood_biochemistry | Lipoprotein A | NA | 741,840486 | gr1_vs_all | -0,053533043 | 0,016883135 | 0,00152222 | 0,003843991 | -0,086623381 | -0,020442706 |
| | Blood_biochemistry | Phosphate | NA | 708,58569 | gr1_vs_all | -0,081250948 | 0,016884403 | 1,50E-06 | 7,55E-06 | -0,114343769 | -0,048158126 |
| | Blood_biochemistry | Total bilirubin | NA | 1379,72003 | gr1_vs_all | -0,085853638 | 0,016874199 | 3,65E-07 | 2,15E-06 | -0,118926459 | -0,052780816 |
| | Blood_biochemistry | Testosterone | NA | 696,114707 | gr1_vs_all | -0,100323636 | 0,016874821 | 2,80E-09 | 2,68E-08 | -0,133397678 | -0,067249594 |
| | Blood_biochemistry | Total protein | NA | 3626,34477 | gr1_vs_all | -0,325983133 | 0,016733992 | 7,82E-84 | 2,61E-81 | -0,358781155 | -0,293185111 |
| | Blood_biochemistry | Triglycerides | NA | 1495,38511 | gr1_vs_all | -0,126817575 | 0,016856493 | 5,54E-14 | 1,29E-12 | -0,159855695 | -0,093779456 |
| | Blood_biochemistry | Urate | NA | 1563,39911 | gr1_vs_all | 0,125299095 | 0,016859143 | 1,11E-13 | 2,41E-12 | 0,092255781 | 0,158342408 |
| | Mental_health | Guilty feelings | NA | 692,786428 | gr1_vs_all | -0,137779094 | 0,016865126 | 3,26E-16 | 9,58E-15 | -0,170834134 | -0,104724054 |
| Cognitive Tests | Fluid_intelligence | FI2 : identify largest number | NA | 231,972423 | gr1_vs_all | 0,064151986 | 0,016878654 | 0,000144621 | 0,000471636 | 0,031070432 | 0,09723354 |
| | Fluid_intelligence | FI3 : word interpolation | NA | 222,090715 | gr1_vs_all | 0,048566995 | 0,016884286 | 0,004025362 | 0,008985184 | 0,015474402 | 0,081659588 |
| | Fluid_intelligence | Fluid intelligence score | NA | 848,166598 | gr1_vs_all | 0,13472979 | 0,01686812 | 1,45E-15 | 4,01E-14 | 0,101668882 | 0,167790698 |
| | Fluid_intelligence | Number of fluid intelligence questions attempted within time limit | NA | 790,49429 | gr1_vs_all | 0,175294397 | 0,016847 | 2,68E-25 | 1,67E-23 | 0,142274884 | 0,20831391 |
| | Fluid_intelligence | FI1 : numeric addition test (Online) | NA | 196,680295 | gr1_vs_all | -0,055083648 | 0,016869462 | 0,00109515 | 0,002866885 | -0,088147186 | -0,02202011 |
| | Fluid_intelligence | FI2 : identify largest number (Online) | NA | 267,504429 | gr1_vs_all | 0,074709309 | 0,016866639 | 9,49E-06 | 3,97E-05 | 0,041651304 | 0,107767314 |
| | Fluid_intelligence | FI3 : word interpolation (Online) | NA | 364,099428 | gr1_vs_all | 0,093045703 | 0,016879748 | 3,58E-08 | 2,50E-07 | 0,059962005 | 0,126129401 |
| | Fluid_intelligence | FI5 : family relationship calculation (Online) | NA | 206,336585 | gr1_vs_all | 0,052550872 | 0,016867113 | 0,001838113 | 0,004549784 | 0,019491938 | 0,085609806 |
| | Fluid_intelligence | FI7 : synonym (Online) | NA | 193,326431 | gr1_vs_all | -0,052148225 | 0,016891399 | 0,00202251 | 0,004926984 | -0,085254758 | -0,019041692 |
| | Fluid_intelligence | FI10 : arithmetic sequence recognition (Online) | NA | 143,372913 | gr1_vs_all | -0,039565841 | 0,016867396 | 0,018999981 | 0,034734883 | -0,072625329 | -0,006506352 |
| | Fluid_intelligence | FI11 : antonym (Online) | NA | 167,389508 | gr1_vs_all | 0,044626588 | 0,016883763 | 0,008219119 | 0,016705526 | 0,011535021 | 0,077718154 |
| | Fluid_intelligence | Fluid intelligence score (Online) | NA | 816,624537 | gr1_vs_all | 0,156540736 | 0,016857352 | 1,74E-20 | 7,56E-19 | 0,123500935 | 0,189580538 |
| | Pairs_matching | Number of correct matches in round | NA | 426,205786 | gr1_vs_all | 0,113181863 | 0,016845518 | 1,88E-11 | 2,84E-10 | 0,080165255 | 0,146198472 |
| | Pairs_matching | Number of incorrect matches in round | NA | 598,110195 | gr1_vs_all | -0,108805048 | 0,016873304 | 1,15E-10 | 1,48E-09 | -0,141876116 | -0,075733979 |
| | Pairs_matching | Time to complete round | NA | 1260,47539 | gr1_vs_all | -0,191167865 | 0,016843297 | 8,94E-30 | 7,45E-28 | -0,224180119 | -0,15815561 |
| | Pairs_matching | Number of correct | NA | 200,667152 | gr1_vs_all | 0,084659542 | 0,01687283 | 5,27E-07 | 2,93E-06 | 0,051589401 | 0,117729682 |

| | | | | | | | | | | | |
|---|---|---|---|---|---|---|---|---|---|---|---|
| | | matches in round | | | | | | | | | |
| | Pairs_matching | Time to complete round | NA | 185,529311 | gr1_vs_all | -0,043388034 | 0,016870043 | 0,010120568 | 0,02004073 | -0,076452711 | -0,010323357 |
| | Pairs_matching | Pairs matching completion status | Completed | 193,629619 | gr1_vs_all | 0,054130279 | 0,016868114 | 0,001333689 | 0,003402267 | 0,021069383 | 0,087191175 |
| | Pairs_matching | Pairs matching completion status | Abandoned | 199,762088 | gr1_vs_all | -0,056061333 | 0,01688079 | 0,000898327 | 0,002389168 | -0,089147074 | -0,022975592 |
| | Prospective_memory | Time to answer | NA | 427,30017 | gr1_vs_all | -0,093516694 | 0,016867396 | 2,99E-08 | 2,18E-07 | -0,126576183 | -0,060457205 |
| | Prospective_memory | Duration screen displayed | NA | 675,631116 | gr1_vs_all | -0,138482731 | 0,016857423 | 2,24E-16 | 6,77E-15 | -0,171522673 | -0,105442789 |
| | Prospective_memory | Number of attempts | NA | 220,00093 | gr1_vs_all | -0,051419346 | 0,016865489 | 0,002300291 | 0,00551005 | -0,084475097 | -0,018363595 |
| | Prospective_memory | Final attempt correct | yes | 190,823575 | gr1_vs_all | 0,049148414 | 0,016869742 | 0,003578579 | 0,008059862 | 0,016084327 | 0,082212501 |
| | Prospective_memory | Final attempt correct | no | 180,620097 | gr1_vs_all | -0,0468676 | 0,016873614 | 0,005481245 | 0,011687091 | -0,079939276 | -0,013795924 |
| | Prospective_memory | Prospective memory result | NA | 202,5425 | gr1_vs_all | -0,050415327 | 0,016864201 | 0,002797449 | 0,006536095 | -0,083468553 | -0,017362101 |
| | Reaction_time | Number of times snap-button pressed | NA | 167,773622 | gr1_vs_all | 0,041849673 | 0,016884191 | 0,013195996 | 0,025328208 | 0,008757266 | 0,074942079 |
| | Reaction_time | Duration to first press of snap-button in each round | NA | 315,304238 | gr1_vs_all | -0,045910432 | 0,016885069 | 0,006553024 | 0,013567337 | -0,079004559 | -0,012816304 |
| | Reaction_time | Mean time to correctly identify matches | NA | 285,321916 | gr1_vs_all | -0,041268023 | 0,016879298 | 0,014497196 | 0,027199242 | -0,07435084 | -0,008185207 |
| | Symbol_digit_substitution | Duration to entering value | NA | 65,1102018 | gr1_vs_all | -0,07810027 | 0,016878184 | 3,73E-06 | 1,70E-05 | -0,111180902 | -0,045019638 |
| | Symbol_digit_substitution | Symbol digit completion status | Completed | 107,394139 | gr1_vs_all | 0,047299617 | 0,016882618 | 0,005088105 | 0,010965744 | 0,014210293 | 0,080388941 |
| | Symbol_digit_substitution | Symbol digit completion status | Abandoned | 46,2979171 | gr1_vs_all | -0,049029644 | 0,016889767 | 0,003700571 | 0,008297245 | -0,08213298 | -0,015926308 |
| | Trail_making | Errors before selecting correct item in alphanumeric path (trail #2) | NA | 156,666422 | gr1_vs_all | -0,043108371 | 0,016874599 | 0,010636519 | 0,02093803 | -0,076181978 | -0,010034764 |
| | Trail_making | Interval between previous point and current one in numeric path (trail #1) | NA | 288,053412 | gr1_vs_all | -0,069221746 | 0,016885147 | 4,15E-05 | 0,000148878 | -0,102316026 | -0,036127466 |
| | Trail_making | Interval between previous point and current one in alphanumeric path (trail #2) | NA | 731,756627 | gr1_vs_all | -0,153773405 | 0,016857775 | 7,99E-20 | 3,19E-18 | -0,186814037 | -0,120732772 |
| | Trail_making | Duration to complete numeric path (trail #1) | NA | 291,760566 | gr1_vs_all | -0,070272747 | 0,016883311 | 3,16E-05 | 0,000115851 | -0,103363428 | -0,037182067 |

| | | | | | | | | | | | |
|---|---|---|---|---|---|---|---|---|---|---|---|
| | Trail_making | Duration to complete alphanumeric path (trail #2) | NA | 740,691497 | gr1_vs_all | -0,157328118 | 0,016854272 | 1,10E-20 | 5,02E-19 | -0,190361883 | -0,124294352 |
| | Trail_making | Trail making completion status | Completed | 217,22551 | gr1_vs_all | 0,057915552 | 0,016880636 | 0,000602683 | 0,001697698 | 0,024830113 | 0,09100099 |
| | Trail_making | Trail making completion status | Abandoned | 386,898843 | gr1_vs_all | -0,100623413 | 0,016866774 | 2,47E-09 | 2,40E-08 | -0,133681682 | -0,067565144 |
| | Trail_making | Total errors traversing numeric path (trail #1) | NA | 187,574422 | gr1_vs_all | -0,052127427 | 0,016886656 | 0,00202499 | 0,004926984 | -0,085224664 | -0,01903019 |
| | Blood_count | Haemoglobin concentration | NA | 702,749659 | gr2_vs_all | -0,062801855 | 0,013529949 | 3,48E-06 | 0,000386112 | -0,089320069 | -0,036283642 |
| | Blood_count | Mean corpuscular volume | NA | 749,641305 | gr2_vs_all | -0,039797483 | 0,013539453 | 0,003292101 | 0,026834717 | -0,066334323 | -0,013260642 |
| | Blood_count | Mean corpuscular haemoglobin | NA | 884,222648 | gr2_vs_all | -0,04710875 | 0,013534194 | 0,000501007 | 0,008491636 | -0,073635283 | -0,020582216 |
| | Blood_count | Platelet count | NA | 783,79288 | gr2_vs_all | 0,042049812 | 0,013529146 | 0,001885331 | 0,020562255 | 0,015533172 | 0,068566451 |
| | Blood_pressure | Pulse rate, automated reading | NA | 680,157931 | gr2_vs_all | -0,069281045 | 0,013540131 | 3,13E-07 | 7,84E-05 | -0,095819213 | -0,042742877 |
| | Body_size_measures | Hip circumference | NA | 675,306352 | gr2_vs_all | -0,055607212 | 0,013532197 | 3,98E-05 | 0,00163312 | -0,082129832 | -0,029084593 |
| | Body_size_measures | Weight | NA | 1012,90754 | gr2_vs_all | -0,070058094 | 0,01352852 | 2,26E-07 | 7,52E-05 | -0,096573507 | -0,043542682 |
| | Blood_biochemistry | Cystatin C | NA | 838,833157 | gr2_vs_all | -0,040516615 | 0,01353054 | 0,002752306 | 0,02457416 | -0,067035986 | -0,013997244 |
| | Blood_biochemistry | Glycated haemoglobin (HbA1c) | NA | 688,531651 | gr2_vs_all | 0,050554348 | 0,013534701 | 0,000188051 | 0,004792804 | 0,024026822 | 0,077081874 |
| | Blood_biochemistry | Total bilirubin | NA | 796,25843 | gr2_vs_all | -0,049547503 | 0,013539065 | 0,000253186 | 0,005725301 | -0,076083584 | -0,023011423 |
| Cognitive Tests | Fluid_intelligence | FI1 : numeric addition test | NA | 203,782634 | gr2_vs_all | 0,052984133 | 0,013534442 | 9,08E-05 | 0,002927659 | 0,026457115 | 0,079511152 |
| | Fluid_intelligence | FI13 : subset inclusion logic | NA | 184,738836 | gr2_vs_all | -0,049893521 | 0,013542997 | 0,000230061 | 0,005369735 | -0,076437306 | -0,023349736 |
| | Fluid_intelligence | Number of fluid intelligence questions attempted within time limit | NA | 177,716474 | gr2_vs_all | 0,039409143 | 0,013543204 | 0,003619151 | 0,028723417 | 0,012864951 | 0,065953334 |
| | Fluid_intelligence | FI1 : numeric addition test (Online) | NA | 133,69316 | gr2_vs_all | -0,037443034 | 0,013532438 | 0,005663624 | 0,03872651 | -0,063966124 | -0,010919943 |
| | Fluid_intelligence | FI2 : identify largest number (Online) | NA | 220,949391 | gr2_vs_all | 0,061707301 | 0,013528924 | 5,11E-06 | 0,000511431 | 0,035191097 | 0,088223505 |
| | Fluid_intelligence | FI6 : conditional arithmetic (Online) | NA | 141,468414 | gr2_vs_all | 0,037227362 | 0,013541965 | 0,005981795 | 0,039661277 | 0,010685597 | 0,063769126 |
| | Numeric_memory | Maximum digits remembered correctly | NA | 263,142047 | gr2_vs_all | -0,058879892 | 0,013537406 | 1,37E-05 | 0,000806423 | -0,08541272 | -0,032347064 |

| | | | | | | | | | | | |
|---|---|---|---|---|---|---|---|---|---|---|---|
| | Pairs_matching | Pairs matching completion status | Completed | 149,793444 | gr2_vs_all | -0,041875623 | 0,013530675 | 0,001971415 | 0,020972503 | -0,068395259 | -0,015355988 |
| | Pairs_matching | Pairs matching completion status | Completed with pause | 164,318816 | gr2_vs_all | 0,045693634 | 0,013537081 | 0,000738207 | 0,010275584 | 0,019161443 | 0,072225825 |
| | Prospective_memory | Duration screen displayed | NA | 292,48497 | gr2_vs_all | -0,059950047 | 0,013536131 | 9,52E-06 | 0,000732012 | -0,086480377 | -0,033419718 |
| | Prospective_memory | Number of attempts | NA | 223,506634 | gr2_vs_all | -0,052238711 | 0,01352668 | 0,00011282 | 0,003318237 | -0,078750517 | -0,025726905 |
| | Prospective_memory | Final attempt correct | yes | 142,259777 | gr2_vs_all | 0,036640349 | 0,013532111 | 0,006781025 | 0,042645239 | 0,010117899 | 0,0631628 |
| | Prospective_memory | Final attempt correct | no | 153,587557 | gr2_vs_all | -0,039853152 | 0,013534587 | 0,003237618 | 0,026834717 | -0,066380455 | -0,013325848 |
| | Prospective_memory | Prospective memory result | NA | 208,562545 | gr2_vs_all | -0,051913791 | 0,013525595 | 0,000124281 | 0,003550888 | -0,078423469 | -0,025404113 |
| | Reaction_time | Duration to first press of snap-button in each round | NA | 302,192109 | gr2_vs_all | -0,044001217 | 0,013543117 | 0,001159911 | 0,014145258 | -0,070545238 | -0,017457196 |
| | Reaction_time | Mean time to correctly identify matches | NA | 323,91803 | gr2_vs_all | -0,046850438 | 0,013537642 | 0,000539657 | 0,0087845 | -0,073383729 | -0,020317147 |
| | Trail_making | Interval between previous point and current one in numeric path (trail #1) | NA | 154,949199 | gr2_vs_all | -0,037235643 | 0,013546883 | 0,005988853 | 0,039661277 | -0,063787047 | -0,01068424 |
| | Trail_making | Interval between previous point and current one in alphanumeric path (trail #2) | NA | 217,505393 | gr2_vs_all | -0,045707198 | 0,013543543 | 0,000739842 | 0,010275584 | -0,072252055 | -0,019162341 |
| | Trail_making | Duration to complete alphanumeric path (trail #2) | NA | 216,31847 | gr2_vs_all | -0,045947574 | 0,01354186 | 0,00069249 | 0,010275584 | -0,072489132 | -0,019406017 |
| | Blood_count | White blood cell (leukocyte) count | NA | 3924,09399 | gr3_vs_all | 0,27129787 | 0,013426495 | 5,36E-90 | 2,68E-87 | 0,244982424 | 0,297613317 |
| | Blood_count | Red blood cell (erythrocyte) count | NA | 798,186923 | gr3_vs_all | 0,058256475 | 0,013547978 | 1,72E-05 | 5,98E-05 | 0,031702927 | 0,084810023 |
| | Blood_count | Haemoglobin concentration | NA | 2101,44285 | gr3_vs_all | 0,187797331 | 0,013491236 | 7,26E-44 | 1,21E-41 | 0,161354994 | 0,214239667 |
| | Blood_count | Haematocrit percentage | NA | 1626,73656 | gr3_vs_all | 0,15395562 | 0,013508572 | 5,23E-30 | 5,03E-28 | 0,127479306 | 0,180431935 |
| | Blood_count | Mean corpuscular volume | NA | 1646,6323 | gr3_vs_all | 0,087417569 | 0,013541961 | 1,10E-10 | 1,15E-09 | 0,060875813 | 0,113959325 |
| | Blood_count | Mean corpuscular haemoglobin | NA | 2137,42729 | gr3_vs_all | 0,113875761 | 0,013529057 | 4,08E-17 | 1,07E-15 | 0,087359297 | 0,140392225 |
| | Blood_count | Mean corpuscular haemoglobin concentration | NA | 814,172658 | gr3_vs_all | 0,12360941 | 0,013521566 | 6,65E-20 | 2,22E-18 | 0,097107627 | 0,150111192 |

| | | | | | | | | | | | |
|---|---|---|---|---|---|---|---|---|---|---|---|
| | Blood_count | Red blood cell (erythrocyte) distribution width | NA | 1680,09206 | gr3_vs_all | -0,09373664 | 0,013533235 | 4,43E-12 | 6,24E-11 | -0,120261293 | -0,067211988 |
| | Blood_count | Platelet count | NA | 1415,02143 | gr3_vs_all | 0,075914679 | 0,013534998 | 2,06E-08 | 1,27E-07 | 0,04938657 | 0,102442787 |
| | Blood_count | Platelet crit | NA | 1882,21561 | gr3_vs_all | 0,116831154 | 0,013525964 | 6,12E-18 | 1,80E-16 | 0,090320752 | 0,143341556 |
| | Blood_count | Mean platelet (thrombocyte) volume | NA | 911,654404 | gr3_vs_all | 0,032182103 | 0,013551211 | 0,017564317 | 0,032647429 | 0,005622217 | 0,058741989 |
| | Blood_count | Lymphocyte count | NA | 3878,59923 | gr3_vs_all | 0,273766113 | 0,013414539 | 9,44E-92 | 9,44E-89 | 0,2474741 | 0,300058127 |
| | Blood_count | Monocyte count | NA | 2481,75364 | gr3_vs_all | 0,174890746 | 0,013504851 | 3,20E-38 | 3,56E-36 | 0,148421724 | 0,201359769 |
| | Blood_count | Neutrophill count | NA | 2607,48253 | gr3_vs_all | 0,1937587 | 0,013492043 | 1,46E-46 | 2,91E-44 | 0,167314782 | 0,220202618 |
| | Blood_count | Eosinophill count | NA | 2681,79886 | gr3_vs_all | 0,220019809 | 0,013466256 | 1,15E-59 | 2,87E-57 | 0,193626432 | 0,246413186 |
| | Blood_count | Lymphocyte percentage | NA | 788,339167 | gr3_vs_all | 0,063574886 | 0,013544476 | 2,70E-06 | 1,08E-05 | 0,0370282 | 0,090121571 |
| | Blood_count | Neutrophill percentage | NA | 914,26812 | gr3_vs_all | -0,077966915 | 0,013546415 | 8,75E-09 | 5,91E-08 | -0,104517401 | -0,051416429 |
| | Blood_count | Eosinophill percentage | NA | 1819,82393 | gr3_vs_all | 0,134556603 | 0,013503741 | 2,44E-23 | 1,11E-21 | 0,108089756 | 0,161023449 |
| | Blood_count | Reticulocyte percentage | NA | 1993,09759 | gr3_vs_all | 0,147171681 | 0,013519147 | 1,57E-27 | 1,05E-25 | 0,12067464 | 0,173668722 |
| | Blood_count | Reticulocyte count | NA | 2103,59617 | gr3_vs_all | 0,154000276 | 0,013518346 | 5,53E-30 | 5,03E-28 | 0,127504805 | 0,180495748 |
| | Blood_count | Mean reticulocyte volume | NA | 1969,98469 | gr3_vs_all | -0,130233697 | 0,013531455 | 6,94E-22 | 2,89E-20 | -0,156754862 | -0,103712532 |
| | Blood_count | High light scatter reticulocyte percentage | NA | 1526,82601 | gr3_vs_all | 0,108229181 | 0,013535667 | 1,35E-15 | 2,93E-14 | 0,081699761 | 0,134758601 |
| | Blood_count | High light scatter reticulocyte count | NA | 1650,67182 | gr3_vs_all | 0,116462838 | 0,013535162 | 8,16E-18 | 2,33E-16 | 0,089934409 | 0,142991268 |
| | Blood_count_ratio | Lymphocyte-to-Monocyte ratio | NA | 1075,56633 | gr3_vs_all | 0,077617476 | 0,013531504 | 9,81E-09 | 6,54E-08 | 0,051096215 | 0,104138736 |
| | Blood_count_ratio | Platelet-to-Lymphocyte ratio | NA | 2974,09436 | gr3_vs_all | -0,186244257 | 0,013494977 | 3,76E-43 | 5,37E-41 | -0,212693925 | -0,159794588 |
| | Blood_count_ratio | Neutrophill-to-Lymphocyte ratio | NA | 850,769053 | gr3_vs_all | -0,070270844 | 0,013545633 | 2,15E-07 | 1,07E-06 | -0,096819797 | -0,043721891 |
| | Blood_count_ratio | Eosinophill-to-Lymphocyte ratio | NA | 1483,00528 | gr3_vs_all | 0,111179951 | 0,013513705 | 2,02E-16 | 4,92E-15 | 0,084693575 | 0,137666327 |
| | Blood_pressure | Pulse rate, automated reading | NA | 968,209157 | gr3_vs_all | 0,09862201 | 0,013544491 | 3,41E-13 | 5,33E-12 | 0,072075296 | 0,125168724 |
| | Blood_pressure | Diastolic blood pressure, automated reading | NA | 1333,67708 | gr3_vs_all | 0,142726471 | 0,013508408 | 4,94E-26 | 2,74E-24 | 0,116250478 | 0,169202463 |
| | Smoking | Past tobacco smoking | NA | 845,078859 | gr3_vs_all | -0,135073768 | 0,013528578 | 2,00E-23 | 9,50E-22 | -0,161589295 | -0,108558242 |
| | Smoking | Smoking status | Never | 937,307483 | gr3_vs_all | -0,138746118 | 0,013519256 | 1,17E-24 | 6,17E-23 | -0,165243372 | -0,112248864 |
| | Smoking | Ever smoked | NA | 821,904775 | gr3_vs_all | 0,137662962 | 0,013529155 | 2,88E-24 | 1,44E-22 | 0,111146305 | 0,164179619 |
| | Blood_biochemistry | Alkaline phosphatase | NA | 807,253246 | gr3_vs_all | 0,042726668 | 0,013550862 | 0,001617796 | 0,003974928 | 0,016167467 | 0,069285869 |

| | | | | | | | | | | | |
|---|---|---|---|---|---|---|---|---|---|---|---|
| | Blood_biochemistry | Apolipoprotein A | NA | 889,589018 | gr3_vs_all | 0,074559345 | 0,01352386 | 3,56E-08 | 2,10E-07 | 0,048053067 | 0,101065623 |
| | Blood_biochemistry | Aspartate aminotransferase | NA | 1916,77697 | gr3_vs_all | 0,176385743 | 0,013497782 | 6,95E-39 | 8,69E-37 | 0,149930576 | 0,202840911 |
| | Blood_biochemistry | Direct bilirubin | NA | 776,944535 | gr3_vs_all | 0,066755705 | 0,013547572 | 8,39E-07 | 3,71E-06 | 0,040202951 | 0,093308459 |
| | Blood_biochemistry | Calcium | NA | 651,531229 | gr3_vs_all | 0,074097704 | 0,013546402 | 4,55E-08 | 2,61E-07 | 0,047547245 | 0,100648164 |
| | Blood_biochemistry | Creatinine | NA | 1706,15009 | gr3_vs_all | -0,127062101 | 0,013520104 | 6,07E-21 | 2,33E-19 | -0,153561017 | -0,100563185 |
| | Blood_biochemistry | C-reactive protein | NA | 886,846034 | gr3_vs_all | 0,067937168 | 0,013548283 | 5,36E-07 | 2,44E-06 | 0,04138302 | 0,094491315 |
| | Blood_biochemistry | Cystatin C | NA | 1102,58691 | gr3_vs_all | -0,053256228 | 0,013540951 | 8,42E-05 | 0,000256556 | -0,079796005 | -0,026716451 |
| | Blood_biochemistry | Gamma glutamyltransferase | NA | 1163,57644 | gr3_vs_all | 0,080408668 | 0,013549591 | 2,99E-09 | 2,14E-08 | 0,053851957 | 0,106965379 |
| | Blood_biochemistry | Glycated haemoglobin (HbA1c) | NA | 1679,36829 | gr3_vs_all | -0,123304961 | 0,01352647 | 8,44E-20 | 2,72E-18 | -0,149816354 | -0,096793568 |
| | Blood_biochemistry | HDL cholesterol | NA | 748,226476 | gr3_vs_all | 0,055931429 | 0,013533228 | 3,60E-05 | 0,000117486 | 0,029406789 | 0,082456069 |
| | Blood_biochemistry | IGF-1 | NA | 1294,62741 | gr3_vs_all | 0,092165975 | 0,013528583 | 9,82E-12 | 1,33E-10 | 0,06565044 | 0,118681509 |
| | Blood_biochemistry | Total bilirubin | NA | 1687,06864 | gr3_vs_all | 0,104978529 | 0,013537493 | 9,23E-15 | 1,85E-13 | 0,07844553 | 0,131511529 |
| | Blood_biochemistry | Total protein | NA | 2526,17423 | gr3_vs_all | 0,227085466 | 0,013462874 | 1,90E-63 | 6,34E-61 | 0,200698719 | 0,253472214 |
| | Blood_biochemistry | Triglycerides | NA | 996,033959 | gr3_vs_all | 0,08446962 | 0,013538743 | 4,48E-10 | 3,83E-09 | 0,057934171 | 0,111005069 |
| | Blood_biochemistry | Urate | NA | 1308,98956 | gr3_vs_all | -0,104909364 | 0,013534166 | 9,47E-15 | 1,86E-13 | -0,131435842 | -0,078382886 |
| | Mental_health | Guilty feelings | NA | 739,921618 | gr3_vs_all | 0,147153186 | 0,013525074 | 1,68E-27 | 1,05E-25 | 0,120644528 | 0,173661845 |
| | Mental_health | Risk taking | NA | 646,178341 | gr3_vs_all | 0,125697375 | 0,013520764 | 1,58E-20 | 5,84E-19 | 0,099197165 | 0,152197585 |
| Cognitive Tests | Fluid_intelligence | Attempted fluid intelligence (FI) test. | NA | 114,440391 | gr3_vs_all | 0,031061556 | 0,013536226 | 0,021759501 | 0,039490928 | 0,004531041 | 0,057592072 |
| | Fluid_intelligence | FI1 : numeric addition test | NA | 187,46388 | gr3_vs_all | -0,048741205 | 0,013547507 | 0,000321604 | 0,000881107 | -0,075293831 | -0,022188579 |
| | Fluid_intelligence | FI3 : word interpolation | NA | 212,902115 | gr3_vs_all | -0,046557624 | 0,013554743 | 0,000594085 | 0,0015588 | -0,073124432 | -0,019990816 |
| | Fluid_intelligence | FI7 : synonym | NA | 143,151513 | gr3_vs_all | -0,034979835 | 0,013556452 | 0,009877419 | 0,019598054 | -0,061549993 | -0,008409678 |
| | Fluid_intelligence | FI8 : chained arithmetic | NA | 140,269551 | gr3_vs_all | 0,031534875 | 0,013560739 | 0,02005647 | 0,036800862 | 0,004956315 | 0,058113435 |
| | Fluid_intelligence | FI13 : subset inclusion logic | NA | 128,695614 | gr3_vs_all | 0,034757593 | 0,013557449 | 0,010361892 | 0,020478048 | 0,008185481 | 0,061329706 |
| | Fluid_intelligence | Fluid intelligence score | NA | 664,99273 | gr3_vs_all | -0,10563294 | 0,013543699 | 6,49E-15 | 1,35E-13 | -0,132178102 | -0,079087778 |
| | Fluid_intelligence | Number of fluid intelligence questions attempted within time limit | NA | 687,586997 | gr3_vs_all | -0,152474407 | 0,013520251 | 2,03E-29 | 1,69E-27 | -0,178973611 | -0,125975203 |
| | Fluid_intelligence | FI1 : numeric addition test (Online) | NA | 260,715802 | gr3_vs_all | 0,073017876 | 0,013538399 | 6,98E-08 | 3,92E-07 | 0,046483102 | 0,099552651 |
| | Fluid_intelligence | FI2 : identify largest number (Online) | NA | 393,784044 | gr3_vs_all | -0,109976997 | 0,01352779 | 4,52E-16 | 1,05E-14 | -0,136490978 | -0,083463016 |
| | Fluid_intelligence | FI3 : word interpolation (Online) | NA | 313,515908 | gr3_vs_all | -0,080119071 | 0,013550788 | 3,42E-09 | 2,42E-08 | -0,106678127 | -0,053560015 |
| | Fluid_intelligence | FI4 : positional | NA | 119,840361 | gr3_vs_all | -0,0309929 | 0,013550295 | 0,022190455 | 0,040054974 | -0,057550991 | -0,004434809 |

| | | | | | | | | | | | |
|---|---|---|---|---|---|---|---|---|---|---|---|
| | | arithmetic (Online) | | | | | | | | | |
| | Fluid_intelligence | FI5 : family relationship calculation (Online) | NA | 250,754482 | gr3_vs_all | -0,063863453 | 0,013538266 | 2,40E-06 | 9,70E-06 | -0,090397967 | -0,037328938 |
| | Fluid_intelligence | FI6 : conditional arithmetic (Online) | NA | 223,06332 | gr3_vs_all | -0,058699032 | 0,013550981 | 1,49E-05 | 5,25E-05 | -0,085258467 | -0,032139596 |
| | Fluid_intelligence | FI7 : synonym (Online) | NA | 212,336699 | gr3_vs_all | 0,05727609 | 0,013559023 | 2,41E-05 | 8,22E-05 | 0,030700893 | 0,083851286 |
| | Fluid_intelligence | FI13 : subset inclusion logic (Online) | NA | 185,265692 | gr3_vs_all | 0,049906095 | 0,013551778 | 0,000231382 | 0,000661091 | 0,023345099 | 0,07646709 |
| | Fluid_intelligence | Fluid intelligence score (Online) | NA | 651,030292 | gr3_vs_all | -0,124797574 | 0,013534531 | 3,20E-20 | 1,14E-18 | -0,151324767 | -0,098270381 |
| | Numeric_memory | Maximum digits remembered correctly | NA | 187,877141 | gr3_vs_all | 0,042038838 | 0,013552538 | 0,001925082 | 0,004649957 | 0,015476352 | 0,068601323 |
| | Pairs_matching | Number of correct matches in round | NA | 329,272107 | gr3_vs_all | -0,087440461 | 0,013525666 | 1,04E-10 | 1,09E-09 | -0,11395028 | -0,060930643 |
| | Pairs_matching | Number of incorrect matches in round | NA | 292,996805 | gr3_vs_all | 0,053300431 | 0,013554769 | 8,44E-05 | 0,000256573 | 0,026733572 | 0,079867291 |
| | Pairs_matching | Time to complete round | NA | 860,516339 | gr3_vs_all | 0,130508753 | 0,013533515 | 5,78E-22 | 2,51E-20 | 0,103983551 | 0,157033956 |
| | Pairs_matching | Number of correct matches in round | NA | 132,549638 | gr3_vs_all | -0,055921417 | 0,013549026 | 3,68E-05 | 0,00011919 | -0,082477021 | -0,029365814 |
| | Pairs_matching | Time to complete round | NA | 179,499221 | gr3_vs_all | 0,041977832 | 0,013543467 | 0,001940917 | 0,00467691 | 0,015433124 | 0,06852254 |
| | Pairs_matching | Pairs matching completion status | Completed with pause | 117,663709 | gr3_vs_all | -0,032719822 | 0,013551099 | 0,015762467 | 0,029462556 | -0,059279488 | -0,006160157 |
| | Prospective_memory | Time to answer | NA | 421,395744 | gr3_vs_all | 0,092224482 | 0,013537566 | 9,83E-12 | 1,33E-10 | 0,06569134 | 0,118757624 |
| | Prospective_memory | Duration screen displayed | NA | 728,525288 | gr3_vs_all | 0,149324341 | 0,013518037 | 2,70E-28 | 2,07E-26 | 0,122829475 | 0,175819208 |
| | Prospective_memory | Number of attempts | NA | 365,725603 | gr3_vs_all | 0,085478599 | 0,013531582 | 2,72E-10 | 2,49E-09 | 0,058957186 | 0,112000012 |
| | Prospective_memory | Final attempt correct | yes | 265,522875 | gr3_vs_all | -0,068387924 | 0,013539041 | 4,43E-07 | 2,05E-06 | -0,094923956 | -0,041851893 |
| | Prospective_memory | Final attempt correct | no | 270,294269 | gr3_vs_all | 0,070136401 | 0,013541527 | 2,25E-07 | 1,10E-06 | 0,043595497 | 0,096677305 |
| | Prospective_memory | Prospective memory result | NA | 339,500664 | gr3_vs_all | 0,084505904 | 0,013530709 | 4,30E-10 | 3,71E-09 | 0,057986202 | 0,111025606 |
| | Reaction_time | Duration to first press of snap-button in each round | NA | 505,986503 | gr3_vs_all | 0,07367506 | 0,013549806 | 5,46E-08 | 3,12E-07 | 0,047117928 | 0,100232193 |
| | Reaction_time | Mean time to correctly identify matches | NA | 508,425755 | gr3_vs_all | 0,073537028 | 0,013544779 | 5,72E-08 | 3,25E-07 | 0,046989749 | 0,100084308 |
| | Symbol_digit_substitution | Duration to entering value | NA | 63,0453263 | gr3_vs_all | 0,075623433 | 0,013547989 | 2,41E-08 | 1,44E-07 | 0,049069863 | 0,102177004 |
| | Trail_making | Errors before selecting correct item in alphanum | NA | 179,730267 | gr3_vs_all | 0,049454624 | 0,013545988 | 0,000261942 | 0,000735792 | 0,022904976 | 0,076004272 |

| | | | | | | | | | | | |
|---|---|---|---|---|---|---|---|---|---|---|---|
| | | eric path (trail #2) | | | | | | | | | |
| | Trail_making | Interval between previous point and current one in numeric path (trail #1) | NA | 340,909003 | gr3_vs_all | 0,081923405 | 0,013550595 | 1,51E-09 | 1,15E-08 | 0,055364726 | 0,108482083 |
| | Trail_making | Interval between previous point and current one in alphanumeric path (trail #2) | NA | 689,586791 | gr3_vs_all | 0,144911716 | 0,013525132 | 1,01E-26 | 5,95E-25 | 0,118402944 | 0,171420488 |
| | Trail_making | Duration to complete numeric path (trail #1) | NA | 335,512404 | gr3_vs_all | 0,08081071 | 0,013549568 | 2,50E-09 | 1,80E-08 | 0,054254044 | 0,107367376 |
| | Trail_making | Duration to complete alphanumeric path (trail #2) | NA | 694,157041 | gr3_vs_all | 0,14744387 | 0,013522273 | 1,29E-27 | 9,25E-26 | 0,120940702 | 0,173947037 |
| | Trail_making | Total errors traversing alphanumeric path (trail #2) | NA | 160,754247 | gr3_vs_all | 0,043470479 | 0,013555037 | 0,001343311 | 0,003325027 | 0,016903094 | 0,070037863 |

**Supplementary Table 6. Differences in gene-risk scores approximating endophenotypes for groups of SCZ cases in Dorsolateral prefrontal cortex.**

Group-specific gene-risk scores (gene-RS) analysis using clustering of SCZ cases in DLPC from 35 PGC cohorts. Gene-RS are imputed in each cohort using UKBB estimates for SCZ related phenotypes and genes in DLPC tissue. Differences in gene-RS approximating a phenotype are tested via Generalized Linear Model (GLM) with gene-RS the dependent variable and group-specific clustering structure ($gr_i$ versus remaining cases) and covariates the independent variables. Each cohort is tested separately and results are summarized via meta-analysis. The regression coefficient $\boldsymbol{\beta}$ estimate, standard error, p-value and 95% confidence interval refer to the grouping variable, estimates are corrected for multiple testing in a group-specific manner using Benjamini-Hochberg procedure. The family for GLM applied is always Gaussian due to the continuous nature of gene-RS. Each phenotype-group test is annotated with the corresponding cluster-reliability metric (CRM). The table shows significant and reliable results

(FDR < 0.05 and CRM > 610) for all phenotype classes but cognitive ones and only significant (FDR < 0.05) for cognitive classes.

**Supplementary Table 7 (separate file) List of PGC Schizophrenia Working Group members with affiliations**

**Supplementary Data 1. (separate file)**

**Prior features construction in tissue-specific models.**

H3K27ac ChIP-Seq and ATAC-Seq prior features as well as GWAS derived for CAD and SCZ. The table contains prior features with corresponding GEO access number and data type used to build tissue specific PriLer gene expression models as well as the tissue-model in which they are included. When multiple features share the same PriLer name, the final prior was built as union of gene regulatory regions.

**Supplementary Data 2. (separate file)**

**Significant genes associated with Coronary Artery Disease.**

Significant genes after tissue-specific multiple testing correction via Benjamini-Hochberg (FDR $\leq 0.05$) associated with CAD across 11 tissues. Gene expression models are built from PriLer in GTEx reference panel and imputed in UKBB and CARDIoGRAM cohorts. UKBB is used as discovery dataset composed of 19,026 cases and 321,916 controls, replication results are derived from meta-analysis of 9 CARDIoGRAM cohorts (13,279 cases and 13,402 controls). Each gene is tested via logistic regression correcting for additional covariates. Z-statistic is defined as the gene estimator divided by its standard error $\frac{\beta}{SE\ \beta}$. Each gene is annotated with GWAS from [20] and from matched GWAS in UKBB (Methods) considering the best result around the TSS gene (200kb window). In both cases, GWASs results are considered significant based on FDR correction. Hence, a gene is considered in a new loci if GWAS best hit in that loci is not significant at 0.05 FDR level, divided per published results[20] and matched GWAS respectively.

**Supplementary Data 3. (separate file)**

**Significant pathways associated with Coronary Artery Disease.**

Significant pathways after tissue-specific and database-specific multiple testing correction via Benjamini-Hochberg (FDR ≤ 0.05) associated with CAD across 11 tissues. The number of genes detected depends on the intersection between genes in a pathway and reliable genes predicted with PriLer in a certain tissue. Each pathway is tested via logistic regression correcting for additional covariates. Z-statistic is defined as the gene estimator divided by its standard error $\frac{\beta}{SE\ \beta}$. In addition, each pathway is assigned to one of the possible 3 classes: genes P < pathway P i.e. there is at least one gene in that gene-set more significant than the overall pathway; pathway P < genes P & genes FDR > 0.05 i.e. the pathway is more significant than any gene in that gene-set and no genes is significant after multiple testing correction; pathway P < genes P & genes FDR < 0.05 i.e. the pathway is more significant than any gene in that gene-set and at least one gene pass FDR 0.05 threshold.

**Supplementary Data 4. (separate file)**

**Group-specific Pathways for CAD cases clustering in Liver.**

Using the clustering structure of CAD cases from UKBB found in liver, the table reports pathways that are significant (FDR 0.01) in at least one group across all tissues, tested via

Wilcoxon-Mann-Whitney (WMW) for single a group against remaining cases after normalization. The pathways are initially clumped based on gene set definition at jaccard similarity threshold 0.2. Significance is assessed after tissue-specific and group-specific multiple tested correction via Benjamini-Hochberg procedure. The table shows Z-statistic from PALAS for CAD, the genes composing the pathway, the groups with significantly different distribution, and the corresponding WMW estimate and p-value. "Improvement wrt genes" indicates for each group if the p-value of the pathway is lower than the p-value reached by the corresponding genes.

**Supplementary Data 5. (separate file)**

**Significant genes associated with Schizophrenia.**

Significant genes after tissue-specific multiple testing correction via Benjamini-Hochberg (FDR $\leq 0.05$) associated with SCZ across 10 tissues. Gene expression models are built from PriLer in GTEx and CMC reference panels and imputed in 36 European PGC cohorts and CMC dataset itself. Results from meta-analysis in PGC multiple cohorts are used as discovery for a total of 24,764 cases and 30,655 controls, replication results are derived only for DLPC tissue in CMC from CMC dataset itself (212 controls and 266 cases). Each gene is tested via logistic regression correcting for additional covariates. Z-statistic is defined as the gene estimator divided by its standard error $\frac{\boldsymbol{\beta}}{\boldsymbol{SE}\,\boldsymbol{\beta}}$. Each gene is annotated with GWAS best result around its TSS (200kb window) and GWAS results are considered significant based on FDR correction. Hence, a gene is considered in a new locus if GWAS best hit in that loci is not significant at 0.05 FDR level.

**Supplementary Data 6. (separate file)**

**Significant pathways associated with Schizophrenia.**

Significant pathways after tissue-specific and database-specific multiple testing correction via Benjamini-Hochberg (FDR $\leq 0.05$) associated with SCZ across 11 tissues. The number of genes detected depends on the intersection between genes in a pathway and reliable genes predicted with PriLer in a certain tissue. Each pathway is tested via logistic regression correcting for additional covariates. Z-statistic is defined as the gene estimator divided by its standard error $\frac{\boldsymbol{\beta}}{\boldsymbol{SE\ \beta}}$. Gene-sets databases included are Reactome, Gene Ontology, WikiHuman Pathway and a gene-set for DLPC in CMC panel specifically customized in a previous study[7].

**Supplementary Data 7. (separate file)**

**Group-specific Pathways for SCZ cases clustering in Dorsolateral prefrontal cortex.**

Using the clustering structure of SCZ cases from 35 cohorts of PGC2 found in DLPC, the table reports pathways that are significant (FDR 0.01) in at least one group across all tissues, tested via Wilcoxon-Mann-Whitney (WMW) for single a group against remaining cases after normalization. The pathways in Reactome and GO databases are initially clumped based on gene set definition at jaccard similarity threshold 0.2. The pathways from WikiPathway and CMC Gene Set [7] are filtered considering only pathways composed of at least 2 genes. Significance is assessed after tissue-specific and group-specific multiple tested correction via Benjamini-Hochberg procedure, separately for Reactome and GO selection, WikiPathway and CMC Gene Set. The table shows Z-statistic from PALAS for SCZ, the genes composing the pathway, the groups with significantly different distribution, and the corresponding WMW estimate and p-value. "Improvement wrt genes" indicates for each group if the p-value of the pathway is lower than the p-value reached by the corresponding genes.
