## Supplementary Text and Methods for "Distinct genetic liability profiles define clinically relevant patient strata across common diseases"

**This PDF file includes:**

Methods
Supplementary Text
List of the Schizophrenia Working Group Psychiatric Genomics Consortium members

### Methods

<u>Prior Learned elastic-net regression to model gene expression</u>

We developed a methodology called PriLer (*Prior Learned elastic-net regression*) that estimates gene expression from cis-acting SNPs, combining elastic-net regression with biological annotation of individual genetic variants defined as prior. This includes for example annotation information such as cell type specific chromatin state or GWAS association signal. Since the relevance of each considered biological annotations is a priori unknown, we implemented an iterative learning procedure to obtain optimized weights for each prior in a nested cross-validation fashion (**Supplementary Fig. 1** Module 1, **Supplementary Fig. 2**).

Namely, let $N$ be the total number of genes expressed in a tissue across $M$ individuals, $P$ the total amount of SNPs and indels across all genome and $K$ the number of prior features included. For $n = 1, \dots, N$, we indicate with $Y_n$ the $M$-length vector of expression of gene $n$ and with $X_n$ the genotype matrix $M \times P_n$ of cis-effects for gene $n$ where $P_n$ is the number of cis-variants distant from the corresponding transcription starting site (TSS) not more than 200kb. Prior information is modelled as a $P \times K$ binary matrix $A$ where 1 indicates that variant $p$ intersects prior feature $k$ (e.g. is in an open chromatin region of cell type $k$).

In elastic-net regression without prior information, gene expression is modeled as a function of cis-variants effects, where the regression coefficients for each gene $n$ are found by solving

$$\min_{\boldsymbol{\beta}_n} \left[\frac{1}{M} \| \boldsymbol{Y_n} - X_n \boldsymbol{\beta_n} \|_2^2 + \sum_{p=1,\dots,P_n} L(\beta_{n,p}, \lambda_n, \alpha_n)\right]$$

with $L$ being the elastic-net penalty function specific for variant $p$:

$$L(\beta_{n,p}, \lambda_n, \alpha_n) = \lambda_n\left(\frac{1-\alpha_n}{2}\, \beta_{n,p}^2 + \alpha_n |\beta_{n,p}|\right)$$

The problem is solved separately for each gene using glmnet R package [1] with $\lambda_n$ and $\alpha_n$ hyperparameters controlling shrinkage of regression coefficients and ridge/lasso contribution and are optimally found via nested 5-fold cross validation.

In PriLer instead, we hypothesize that variants carrying biological prior information are more likely to be putative regulatory variants (reg-SNPs) i.e. regulating at least one gene. To that end, each variant $p$ is multiplied by a prior coefficient $v_p$obtained as a nonlinear combination through the sigmoid function of prior information in matrix $A$:

$$v_p = 1 - \frac{1}{1 + exp\left(-\sum_{k=1,\ldots,K} \gamma_k \, A_{pk}\right)}$$

where $\gamma_k$ represents the prior weight associated to prior feature class $k$ (vector form $\gamma$) and is automatically learned by PriLer through an iterative procedure. Thus, PriLer aims at solving the following problem with respect to $\beta_n$for all the genes and the $\gamma$ prior weights vector:

$$\min_{\gamma,\boldsymbol{\beta_n},n=1,\ldots,N} \ \{ \sum_{n=1,\ldots,N} [\frac{1}{M} \parallel \boldsymbol{Y_n} - X_n\boldsymbol{\beta_n} \parallel_2^2 + \sum_{p=1,\ldots,P} v_p \; L(\beta_{n,p}, \lambda_n, \alpha_n)] + E \parallel \boldsymbol{\gamma} \parallel_2^2 \}$$

Note that since we consider all the genes together, we now iterate through $P$ variants although regression coefficients for variants not in cis-regions of a certain gene $n$ are set to 0. The last term of the objective function represents a regularization term for prior weights and the number of hyperparameters is $2N + 1$ i.e. gene-specific $\lambda_n, \alpha_n$ pairs and $E$.

The problem is solved in a 2-step iterative procedure. Initially, prior weights are set to 0 for all the $K$ features. The first step minimizes PriLer function with respect to $\beta_n$ separately for each gene keeping $\boldsymbol{\gamma}$ as fixed (hence $v_p$) via cyclical coordinate descendent algorithm as implemented in glmnet R package; the second step minimizes the PriLer function with respect to $\gamma_k$ for $k = 1,\ldots,K$ keeping $\boldsymbol{\beta_n}$ fixed through globally-convergent method-of-moving-asymptotes implemented in nloptr R package [2]. The algorithm stops until convergence is reached in term of the maximum number of iterations or minimal decrease of the objective function from previous step.

In general, the lower the prior coefficient $v_p$, the less will the corresponding regression coefficient for variant $p$ shrink to zero for all the genes. Hence, the more relevance the variant will have in the gene expression prediction. On the other hand, the weights for the prior features $\gamma_k$ are dependent on putative reg-SNPs across all the genes that have prior information not zero: the more there are reg-SNPs intersecting a certain prior feature, the higher the correspondent prior weight will be. It is also worth noting that, for prior features intersecting a considerable higher number of variants, the corresponding prior weight will be higher since by chance that prior feature intersects more reg-SNPs. However, in the iterative procedure, if that prior feature is not actually relevant for that tissue-regression model, the corresponding weight remains stable and does not increase (see "Evaluation of prior weights selection in PriLer through random prior simulation" section).

Since PriLer uses the combined information across all genes to derive prior weights, we do not want to introduce noise in that estimation due to genes that are poorly explained by cis-effects. Hence, we estimate prior weights using only heritable genes for which a non-null proportion of variation in gene expression is determined by genetic effects. The list of heritable genes for GTEx and CMC are downloaded from http://gusevlab.org/projects/fusion/ database of TWAS method [3] (reference functional data), where heritability is estimated for each gene from cis-SNPs via REML algorithm implemented in GCTA [4]. Heritable genes are defined as those having heritability p-value $< 0.01$ estimated in GTEx v7 (https://gusevlab.org/projects/fusion/weights/GTEX7.txt) and CMC (https://data.broadinstitute.org/alkesgroup/FUSION/WGT/CMC.BRAIN.RNASEQ.tar.bz2). A gene expression prediction model is built for all the genes that have cis-variants in the predefined window. In case of not heritable genes, we use prior coefficients $v_p$ estimated from heritable genes only.

To find an optimal hyperparameter configuration and evaluate gene expression prediction models, we implemented PriLer in a nested 5-fold cross-validation (CV) setting dividing the procedure in 4 steps (**Supplementary Fig. 2**). The first step involves heritable genes only and estimates gene expression using elastic-net regression (enet) without prior information. The inner CV finds the optimal $\alpha_n, \lambda_n$ combination for each gene $n$ separately that minimizes the mean squared error (MSE) on test folders, the outer CV instead builds enet models based on the optimal hyperparameters and evaluates each gene-model via average $R^2$on the test folders ($R_{cv}^2$).

The second step uses $\alpha_n, \lambda_n$ combination found in step 1 and builds PriLer models in the outer CV across all heritable genes for different values of hyperparameter $E$, which controls $\gamma$ module. The optimal $E$ parameter is chosen as the one minimizing MSE on the test folds and for that hyperparameters combination $\alpha_n, \lambda_n$ and $E$ we evaluate PriLer performance based on $R_{cv}^2$. The third step creates a final model for each gene applied to all $M$ samples that will be further used in the external prediction to genotype-only data. Hence, from a single CV, optimal $\alpha_n, \lambda_n$ combination for enet is found and used in PriLer together with optimal $E$ parameter found in step 2. Finally, the fourth step is used to build PriLer (and enet) models for not heritable genes: step from 1 to 3 are repeated but prior weights $\gamma_k$ and consequentially prior coefficients $v_p$ are kept fixed as obtained in step 2 and step 3 (for evaluation and final model creation).

In summary, we obtain $R_{cv}^2$ that estimates PriLer and enet performance, gene expression prediction models together with the corresponding $R^2$computed across all samples and for all the genes having cis-variants in 200kb window.

The algorithm we implemented is inspired by the Lirnet algorithm described in [5], however PriLer is adapted to large reference panels of matched genotype and gene expression data, uses a simplified formula for computing the prior coefficients and optimizes α and λ penalty parameters instead of using the same penalty across all genes, thus allowing for differences in gene sparsity.

We introduce in PriLer the possibility to model also effects from cofounders to gene expression and variant-gene interaction in a linear manner. In this case, the first term of the objective function representing the prediction squared error becomes:

$$\| \boldsymbol{Y_n} - X_n \boldsymbol{\beta_n} - Z\, \boldsymbol{\mu_n} \|_2^2$$

With $Z$ the $M \times C$ confounder matrix unique to all the genes and $\mu_n$ the corresponding regression coefficient specific to gene-model $n$. The penalty factor term however does not change, being applied only to genotype data. This is practically achieved via the *penalty.factor* option of glmnet set to zero in correspondence of the confounders position so that they are included in all the models for gene expression.

Finally, in order to evaluate PriLer performance as well as enet, we used $R^2$ in the sense of fraction of deviance explained by the model as implemented in glmnet (dev.ratio). In our model, we explicitly account for linear confounder effects as well as their interaction with cis-variants due to the probable not orthogonal effect especially between variants and genetically derived ancestry components. However, we are mostly interested in the variance that can be explained by genotype only. Consider $\widehat{Y}$ as the predicted gene expression vector estimated by the model for a certain gene

$$\boldsymbol{\widehat{Y}} \coloneqq X\boldsymbol{\widehat{\beta}} + Z\boldsymbol{\widehat{\mu}}$$

and $\bar{Y}$ the mean original gene expression, let $\|\cdot\|_2$ be the Euclidean norm operator and $\langle\cdot,\cdot\rangle$ be the scalar product operator among 2 vectors, then $R^2$ can be formulated as

$$1 - \frac{\| \boldsymbol{Y} - \boldsymbol{\widehat{Y}} \|_2^2}{\| \boldsymbol{Y} - \bar{Y} \|_2^2} = \frac{\| \boldsymbol{\widehat{Y}} - \bar{Y} \|_2^2 + 2\, \langle \boldsymbol{Y} - \boldsymbol{\widehat{Y}},\ \boldsymbol{\widehat{Y}} - \bar{Y} \rangle}{\sigma_{\boldsymbol{Y}}^2}$$

For this reason, we split $R^2$ in three components: $R_g^2 + R_c^2 + R_{g,c}^2$ (see Appendix A) with

$$R_g^2 = \frac{\| \widehat{\boldsymbol{W}} - \overline{W} \|_2^2 + 2 < \boldsymbol{W} - \widehat{\boldsymbol{W}}, \widehat{\boldsymbol{W}} - \overline{W} >}{\sigma_{\boldsymbol{Y}}^2}$$

$$R_c^2 = \frac{\| \widehat{\boldsymbol{V}} - \bar{\bar{V}} \|_2^2}{\sigma_{\boldsymbol{Y}}^2}$$

$$R_{g,c}^2 = \frac{2 < \boldsymbol{W} - \overline{W}, \widehat{\boldsymbol{V}} - \bar{\bar{V}} >}{\sigma_{\boldsymbol{Y}}^2}$$

where $\widehat{\boldsymbol{W}} \coloneqq X\widehat{\boldsymbol{\beta}}$ is the predicted genotype effect, $\boldsymbol{W} \coloneqq \boldsymbol{Y} - Z\widehat{\boldsymbol{\mu}}$ is the gene expression vector corrected for the confounder effect hence carrying supposedly only the genotype effect and $\overline{W}$ the corresponding mean, $\widehat{\boldsymbol{V}} \coloneqq Z\widehat{\boldsymbol{\mu}}$ is the predicted confounder contribution and $\bar{\bar{V}}$ the corresponding mean. Hence, $R_g^2$ represents the part of the variance in gene expression that is due to the genetic component, $R_c^2$ is the contribution of confounders and $R_{g,c}^2$ represents the joint effect between two. For simplicity, throughout the text we will refer to $R_g^2$ as $R^2$ and average $R_g^2$ in cross validation as $R_{cv}^2$.

Reference panels for training gene expression models

Gene expression prediction models are built based on matched data composed of gene expression and genotype individual dosages, also referred to as reference panels. We used GTEx v6p[6] that includes donors across 44 non-diseased post-mortem tissues and cell lines and CommonMind Consortium (CMC) Release1[7] composed of RNA-Seq data extracted from post-mortem dorsolateral prefrontal cortex (DLPC) for patients with schizoaffective disorders and controls.

For genotype preprocessing, REF and ALT alleles were aligned to human reference genome hg19 and variants were filtered out based on imputation quality score (INFO) < 0.8, minor allele frequency (MAF) < 0.05 and deviation from Hardy-Weinberg Equilibrium (HWE) P < 5e-5 as well as removal of multiallelic position. Since GWAS data is optionally used as prior information in PriLer, genotype data was matched with CAD and SCZ GWAS summary statistic obtained from [8] and [9] in case of GTEx and only SCZ in case of CMC such that only variants with the same position and REF/ALT annotations are kept. Genotype probabilities were then converted to 0-2 dosages where 0 refers to REF/REF configuration and the final number of variants was 6,486,416 and 6,491,178 for GTEx and CMC respectively across 22 autosomal chromosomes.

For RNA-sequencing data, we followed the respective guidelines used to process data for eQTL analysis by the 2 consortia. In case of CMC, we used 'SVA corrected excluded ancestry' gene expression processed data that includes residuals from weighted regression through voom-based log transformed CPM (read counts per million total reads) and correspondent observation weights corrected for chosen confounders (see [7] for details). In case of GTEx instead, we excluded poor quality samples (sample attributes SMAFRZE column equals to 'EXCLUDE'), considered only the ones matching genotype data and excluded tissues with less than 70 resulting samples. We then followed the GTEx guidelines for eQTL analysis[6] i.e. for each tissue, genes such that RPKM > 0.1 in at least 10 individuals and number of reads ≥ 6 in at least 10 individuals were retained, RPKM expression values were quantile normalized to the average empirical distribution observed across samples and expression values were inverse quantile normalized to a standard normal distribution for each gene across samples. We additionally excluded from the analysis tissues sex specific and tissues not matching any prior features (see below) resulting in a total of 33 tissues. Finally, genes were annotated using Ensembl on GRCh37 via biomaRt (Bioconductor), in order to define transcription starting site (TSS).

For covariates included in the PriLer model, we followed again the guidelines for eQTL analysis in the respective consortia. In particular, for CMC we used 5 ancestry components provided and computed via GemTools based on a set of high-quality autosomal SNPs from pre-imputed data. For GTEx instead, we included as covariates individual sex, genotype array platform, PEER components calculated from normalized expression matrices for each tissue separately with the number of PEER factors determined as a function of the tissue sample size (N): 15 factors for $N < 150$, 30 factors for $150 \leq N < 250$ and 35 factors for $N \geq 250$ and finally the first 3 principal components (PCs) from genotype data computed using EIGENSTRAT as implemented in Ricopili (see (*5*) for details). We included in our analysis only samples with Caucasian ancestry: CMC ethnicity 'Caucasian' and GTEx reported race 'white' for a total of 478 samples (212 controls and 266 cases) and 377 respectively.

Our methodology incorporates prior information into elastic-net regression. To that end, we used as prior features cell-type specific open chromatin regions one-hot encoded and included CAD GWAS summary statistic [8] for tissues related to CAD and SCZ GWAS summary statistic [10] for brain lines and immunological cell types. GWAS information is converted into binary using 0.05 and 0.01 nominal p-values threshold respectively.

The resulting prior matrix is a binary format with dimension n. of variants times n. of prior features included in the tissue specific model with 1 indicating either the variant intersects an open chromatin region for that cell type or it passes the nominal GWAS threshold. Open chromatin regions are derived from H3K27ac ChIP-seq data obtained from the Epigenome Roadmap Project as well as ENCODE and merged together (see **Data S1** for full sample list). In addition, H3K27ac and ATAC-Seq feature based profiles are combined and included for heart related tissues, obtained from [11] (GSE72696). For SCZ and brain related tissues, we used ATAC-Seq profiles from human post mortem prefrontal cortex neuronal cells from[12] (GSE83345). All annotation information can be downloaded from the supplemental website at https://gitlab.mpcdf.mpg.de/luciat/castom-igex/-

/tree/master/refData/prior_features/. The brain related prior features from ATAC-Seq (*FPC_neuronal_ATAC_R2* and *FPC_neuronal_ATAC_R4*) were modified due to the reduced number of included putative gene regulatory elements (GREs) compared to the H3K27ac derived features (number of GREs 44,475 and 34,883 versus mean number 128,817.3) and a consequence reduction in the number of variants with those priors that would have greatly penalized the correspondent PriLer prior weight (see below for detail). Hence, for each GREs of these 2 prior features, we extended it by half median length of GREs in H3K27ac data (1,192) in both directions. With the purpose of not introducing noise in the selection of these prior features, the weights are solely estimated from heritable genes (see "Prior Learned elastic-net regression to model gene expression" section). The complete list of tissue-specific gene expression model, number of samples, number of genes and prior features can be found in **Table S1** and tissue specific usage for each prior in **Data S1**. Tissue-specific trained models are also available here https://doi.org/10.6084/m9.figshare.22347574.v2.

Genotype-only datasets preprocessing

To impute gene expression from PriLer in large-scale genotype-only datasets, the first step is to match genetic data with reference panels (GTEx and CMC). In particular, for UK Biobank (UKBB), we used imputed data from third release, aligned REF and ALT allele to hg19 and excluded samples due to non-white British ancestry and withdrawn consent. As post-imputation QC, we filtered variants based on SNP call rate < 0.98, INFO < 0.8, MAF < 0.05 and HWE p-value < 1e-6 as well as multiallelic positions. We then excluded relatives up to 3rd degree based on kinship matrix such that the largest amount of samples not related would be retained, following UKBB guidelines [13]. Additional samples with no matching submitted and inferred gender and poor-quality ones being outliers for heterozygosity and missing rates are excluded. Our final set after quality control included 340,939 individuals. Genotype data was separately matched with previously processed GTEx and CMC imputed genotype excluding variants having differences in ALT frequency > 0.15 resulting in 5,728,140 and 5,774,100 variants respectively. For CAD application, we used as replication 9 case-control European ancestry cohorts from CARDIoGRAM consortium: German Myocardial Infarction Family Studies (GerMIFS) I[14], II[15], III[16], IV[8], V[17], the LUdwigshafen RIsk and Cardiovascular Health Study (LURIC)[18], Cardiogenics (CG), Wellcome Trust Case Control Consortium (WTCCC), Myocardial Infarction Genetics Consortium (MIGen)[19]. Pre-imputation QC was performed on each cohort separately using the following criteria: individual call rate ≥ 0.98, SNP call rate > 0.98, minor allele frequency (MAF) > 0.01, concordant recorded and genotype-derived gender, population outliers excluded (deviate beyond mean ± 5x standard deviation) for top two dimensions from the multidimensional scaling (MDS) analysis, PI_HAT < 0.0625 (individuals more distant away than fourth-degree relatives) in the identity-by-descent (IBD) analysis, heterozygosity rate within mean ± 3 x standard deviation, and HWE p-value > 1e-6. Imputation was performed on each cohort separately using the Haplotype Reference Consortium panel on the Sanger Imputation Server (https://www.sanger.ac.uk/science/tools/sanger-imputation-service). Post-imputation QC was then performed with the following criteria; SNP call rate > 0.98, MAF > 0.05, HWE p-value >1e-6, INFO score ≥ 0.8, multiallelic position excluded and PI_HAT < 0.0625 in IBD analysis for individuals. We then considered all the cohorts together to remove up to fourth-degree relatives (PI_HAT < 0.0625), keeping if possible individuals annotated as cases and/or with the lowest missing rate. Finally, only variants in common across all the cohorts were retained as well as with the aforementioned UKBB-GTEx matched genotype set and such that ALT frequency differences for each pair of cohort/UKBB/GTEx dataset did not exceed 0.15. This procedure yield to a total of 26,681 individuals across the 9 cohorts and 4,257,718 variants matching CARDIoGRAM cohorts, UKBB and GTEx genotyping data. GTEx tissue models adopted for CAD analysis are composed of 2 adipose tissues (subcutaneous and visceral omentum), adrenal gland, 2 artery tissues (aorta and coronary), 2 colon tissues (sigmoid and transverse), 2 heart tissues (atrial appendage and left ventricle), liver and whole blood.

For SCZ application instead, we used 36 PGC cohorts of European ancestry from Psychiatric Genomic Consortium (PGC) for SCZ wave2[10]. Following PGC guidelines, for each cohort we excluded imputed variants based on MAF < 0.01, INFO < 0.6, multiallelic positions and variants that were missing in at least 20 samples (genotype certainty < 0.8). Prior to matching variants with GTEx and CMC, we filtered the reference panels such that INFO ≥ 0.6 and MAF ≥ 0.01 based on Caucasian individuals. Finally, variants with ALT frequency differences across all possible pair of dataset > 0.15 are excluded, obtaining a total of 5,912,207 and 5,934,252 SNPs and Indels when matching GTEx and CMC respectively. Individuals across all the cohorts are excluded if diagnosis is not available and samples are duplicated/related or a total of 55,419 individuals. GTEx tissue models adopted for SCZ analysis are composed of 8 brain tissues (caudate basal ganglia, cerebellar hemisphere, cerebellum, cortex, frontal cortex BA9, hippocampus, hypothalamus, and nucleus accumbens basal ganglia) and cell EBV transformed lymphocytes while CMC tissue model is based on dorsolateral prefrontal cortex.

UKBB phenotype pre-processing and coronary artery disease diagnosis definition

UK Biobank is a large-scale biomedical database and research resource containing genetic, lifestyle and health information from half a million UK participants[13]. We used the available deep phenotyping in two different contexts: i) to define CAD and extract CAD related phenotypes in order to perform TWAS and PALAS as well as detect endophenotype differences and treatment response in CAD cases using as genotype data the matched dataset with CARDIoGRAM cohorts, ii) to perform TWAS and PALAS analysis for SCZ related phenotypes and build gene risk scores (gene-RS) weights to model gene-RS in external cohorts such as PGC.

Similarly to previous CAD HARD definition[20], CAD diagnosis was determined by either hospital episode or self-reported via questionnaire combining ICD10 and ICD9 codes for myocardial infarction and ischaemic heart diseases (I21-I24 and 410-412), old myocardial infarction (I25.2), OPCS-4 codes for procedures for coronary artery bypass graft surgery (CABG) (K40-K46), percutaneous transluminal coronary angioplasty (PTCA) (K49-K50, K75) and self-reported heart attack, PTCA, CABG and triple heart bypass. In addition, we used CAD SOFT definition[20] to define reference set composed of controls for gene T-scores computation (see "From imputed gene expression to gene T-scores"). CAD SOFT phenotype was defined with the same requirement of CAD HARD plus individuals reporting ICD9 codes for angina pectoris and coronary atherosclerosis (413-414), ICD10 codes for angina pectoris and chronic ischemic heart disease (I20, I25), and self-reported angina.

Phenotypes we had access under application numbers 34217 and 25214 were processed for subsequent analysis using PHESANT software[21]. PHESANT automatically converts UKBB phenotypes distribution to continuous inverse-rank normalized, ordered categorical, unordered categorical or binary, depending on original data type (continuous, integer, categorical single or multiple). Based on the final category, the correct generalized linear model was applied during TWAS and PALAS: Gaussian for continuous, logistic for unordered categorical and binary or ordinal logistic regression for ordered categorical. In addition, PHESANT automatically removes phenotypes recorded for less than 500 individuals and constant ones across the samples.

Original phenotypes not converted via PHESANT are only used in hypothesis-driven CAD endophenotype analysis in which clinical phenotypes are tested (35 in total, nominal significant results are shown in **Table S4**).

SHIP-Trend cohort preprocessing

The Study of Health in Pomerania (SHIP-Trend) is a population-based cohort study in West Pomerania (northeast of Germany) and is focused on the prevalence and incidence of common population-relevant diseases and their risk factors. Baseline examinations for SHIP-Trend were carried out between 2008 and 2012, comprising 4,420 participants aged 20 to 81 years. Study design and sampling methods were previously described[22].

Regarding genotyping, data was collected from nonfasting blood samples. A subset of the SHIP-Trend samples was genotyped using the Illumina Human Omni 2.5 array, while the majority of samples were genotypes using Global Screening Array (GSA-24v1). Genotypes were determined using the GenomeStudio 2.0 Genotyping Module (GenCall algorithm). Individuals with a genotyping call rate < 94%, duplicates (based on estimated IBD), and mismatches between reported and genotypedwere removed. Genotypes were imputed using the HRCv1.1 reference panel and using the Eagle and minimac3 software implemented in the Michigan Imputation Server for pre-phasing and imputation, respectively. Before imputation QC steps include the removal of SNPs with a HWE p-value < 0.0001, call rate < 0.95, monomorphic SNPs, variants having position mapping problem from genome build b36 to b37, duplicate IDs, or with inconsistent reference site alleles. As post-imputation QC steps, variants with MAF > 0.05, HWE p-value >1e-6, INFO score ≥ 0.8 were retained and multi-allelic positions were excluded. Individuals more distant away than fourth-degree relatives in the identity-by-descent (IBD) analysis were kept (PI_HAT < 0.0625). The resulting variants were matched with the final set of 4,257,718 variants harmonized for

CARDIoGRAM cohorts, UKBB and GTEx genotyping data (CAD-matched variants). SHIP-Trend variants were matched based on same position and REF/ALT annotation. Variants with ALT frequency differences between SHIP-Trend cohort and GTEx not exceeding 0.15 were kept. This procedure yield to 4,240,949 SNPs in the SHIP-Trend cohort also available in the CAD-matched variants set across 4,119 individuals. Finally, gene expression was imputed based on previously trained models of liver and whole blood tissues using CAD-matched variants (see "*From imputed gene expression to gene T-scores"*).

Regarding transcriptome analysis, RNA was prepared from whole blood under fasting conditions using the PAXgene Blood miRNA Kit (Qiagen, Hilden, Germany). 500ng of RNA was reverse transcribed into cRNA and biotin-UTP-labeled via Illumina TotalPrep-96 RNA Amp Kit (Ambion). 3000ng of cRNA were hybridized to the Illumina HumanHT-12 v3 Expression BeadChips, followed by washing steps as described in the Illumina protocol. Gene expression raw intensity data was generated with the expression arrays were exported from Illumina's GenomeStudio V 2010.1 Gene Expression Module to the R environment and processed (quantile normalization and log2-transformation) with the lumi 1.12.4 package from the Bioconductor open source software as described elsewhere[23]. Quality-controlled gene expression data and genotyping data were available for 976 SHIP-TREND samples.

PsyCourse Study pre-processing

The PsyCourse Study is a longitudinal, multi-center observational study of patients suffering from severe mental disorders (mainly schizophrenia, bipolar disorder, and recurrent depression) as well as healthy control that were subjected to comprehensive neuropsychological testing[24] and assessment of disease history. All participants were subjected to genotyping using the Infinium

Global Screening Array-24 Kit, version 3.0. Prior to imputation, SNPs were filtered based on MAF≥ 0.01, removal of SNPs HWE P < 0.0001, palindrom SNPs and SNPs with MAF deviating more than 10% for EUR reference populations. Subjects were Sex checked and individuals were filtered based on SNP call rate > 98%, individual call rate > 98% and excluding MDS outliers. Genotypes were imputed using the HRCv1.1 reference panel and using the Eagle and minimac3 software implemented in the Michigan Imputation Server for pre-phasing and imputation, respectively, resulting in 7,712,287 SNPs dosages. Subsequently, SNP names were changed to rsID and duplicate rsIDs removed (multiallelic markers and SNP annotation duplicates). This procedure left 556 individuals with suffering from SCZ or schizoaffective disorder. The resulting variants were matched with the final set of 5,934,252 variants harmonized for PGC2 cohorts and CMC genotyping data (SCZ-matched variants). Variants with ALT frequency differences between the PsyCourse Study and CMC not exceeding 0.15 were kept, yielding to 5,094,785 SNPs in the PsyCourse Study also available in the SCZ-matched variants set. Finally, gene expression was imputed based on previously trained models of DLPC tissue using SCZ-matched variants (see "*From imputed gene expression to gene T-scores"*).

From imputed gene expression to gene T-scores

After the gene expression prediction model is built on reference panels, the first step is to impute tissue-specific gene expression on genotype-only cohorts based on PriLer models (**Supplementary Fig. 1** Module 2). Let $\tilde{X}$ be the $L \times P$ matrix of dosages for $L$ new individuals. For each reliable gene $n$ ($R^2 > 0.01$ and $R^2_{cv} > 0$) in a certain tissue, we predict gene expression for $L$ individuals based on cis-effects estimated via PriLer

$$\widehat{\boldsymbol{W}}_n \coloneqq \tilde{X}\,\widehat{\boldsymbol{\beta}}_n.$$

In all applications with the only exception of SHIP-Trend Trend cohort and the PsyCourse Study, $P$ variants in the genotype-only datasets and reference panels are matched via the harmonization process described in *"Genotype-only datasets preprocessing"*. Thus, $\widehat{\boldsymbol{\beta}}_n$is a P-length vector with non-zero entries only in correspondence of the cis-variants in 200kb window of the gene $n$ TSS. Instead, the genotype matrix of SHIP-Trend and PsyCourse are composed of a subset of the original CAD-matched variants or SCZ-matched respectively, of dimension $Q < P$. In these cases, gene expression is imputed using $Q$ regression coefficients $\widehat{\boldsymbol{\beta}}_{\boldsymbol{n}}^{\boldsymbol{Q}}$ also available in $\widehat{\boldsymbol{\beta}}_n$.

We do not use directly predicted gene expression to test for disease association but convert the imputed expression to gene t-scores for each individual. T-scores are generated as individual moderated t-statistic or ordinary t-statistic depending on the sample size due to computational feasibility. For each cohort in PGC and CARDIoGRAM, the samples are divided in a reference set comprising randomly selected 80% of the control individuals as well as the comparison set, composed of the remaining controls plus all the cases. A moderate t-statistic is computed using *eBayes* function from limma R package[25] between each individual in the comparison set and all the other samples in the reference set, bootstrapping over the controls and averaging across 40 folds. The same procedure is used in SHIP-Trend cohort and the PsyCourse Study however without a priori cases-controls division. Instead, in each repetition 20% of the individuals were randomly selected as reference set.

In UKBB, due to the large sample size (~340,000) we defined gene t-score as the ordinary t-statistic for each sample $l$ in the comparison set as $\frac{\bar{C}_n}{sd(\boldsymbol{C_n})/\sqrt{L_{ref}}}$ where $\boldsymbol{C_n} \coloneqq \widehat{W}_n\,(l) - \widehat{\boldsymbol{W}}_{\boldsymbol{n}}\,(ref)$ is the vector of singular differences between current sample $l$ and the samples in reference set of size $L_{ref}$. For CAD analysis, we adopted bootstrapping technique over 10 folds and used as reference set 30% of individuals not annotated as CAD (SOFT) for a total of 92,784 individuals.

For SCZ related phenotypes analysis in UKBB instead, we did not use a priori cases-controls division but randomly selected 10 times 20% of the individuals (68,190 in total) as reference set. Differently from the large incidence of CAD in UKBB cohort, individuals with registered schizophrenia disorders were limited to 1022 out of 340,939 considered samples (ICD10 F20-F29, ICD9 295, self-reported schizophrenia). Because they only compose the 0.29% of the total cohort, they are negligible to the actual reference set size, and we simply sampled across the entire population.

Using gene T-scores instead of imputed gene expression allows both to obtain a similar distribution for all the genes that now are not scaled according to the predictive performance and variance explained of the corresponding PriLer model.

Computation of individual-level pathway-scores

From the gene T-scores, we subsequently computed individual level pathway scores. In contrast to previous approaches[26–28], we do not set a cut-off for gene level significance or perform an enrichment analysis. Instead, for each sample a representative score for the pathway activity is computed as the mean across gene T-scores that belong to a certain pathway. We used as pathway databases Reactome[29] and Gene Ontology[30] as default in CASTom-iGEx pipeline and additionally considered Human WikiPathways[31] as custom gene-sets. In each tissue, gene-sets are defined based on the reliable set in that tissue ($R^2 \geq 0.01$ and $R^2_{cv} > 0$) and only pathways that are not redundant (i.e. composed by the same set of genes) are retained, giving priority to more specific gene-sets being composed of a lower number of genes. The advantage of gene T-scores in the computation of pathways instead of directly imputed gene expression relies on the new scaling space such that each gene can contribute equally regardless the variance explained in the model.

Association of genes and pathways with a trait

For both gene T-scores and pathway scores, we separately tested the association of each gene/pathway with a certain trait (**Supplementary Fig.** Module 2), using *glm* (Gaussian or logistic regression for continuous or binary trait) or *polr* (ordinal logistic regression for ordered categorical) functions in R and correcting for additional covariates. In case of CARDIoGRAM cohorts and UKBB for CAD analysis, we corrected for sex and first 10 Principal Components (PCs) estimated from pre-imputed data. In case of SCZ cohorts, we corrected for 10 PCs (from 1 to 7, 9,15 and 18) as suggested in [10], correcting for biases due array type and to population structure, that are partially reflected in the phenotypic variability. We used additional covariates in UKBB dataset for CAD analysis when testing blood biochemistry (category 17518) and blood count (category 100081) phenotypes to correct for medication effect affecting blood levels: medication for pain relief, constipation, heartburn (Field 6154), dietary supplements (Field 6155, 6179) and medication for cholesterol, blood pressure and diabetes (Field 6153, 6177). When using UKBB for SCZ related phenotypes instead, we considered as confounders first 10 PCs, age, sex and phenotype specific covariates: for 'Maximum digits remembered correctly' (Field 4282) additional covariates are fields 4250, 4253, 4283 and 4285; for Symbol digit substitution (category 122) we tested fields 20158, 20230 and 20245 additionally correcting for fields 20195 and 20200; for T1 structural brain MRI (category 110) we tested all data fields and regional grey matter volumes subclass correcting for scanner coordinates (fields 25756-25759). In general, we refer as gene/pathway Z-statistics as the estimated effect for trait association divided by its standard error. In case of multiple cohorts (CARDIoGRAM and PGC), we implemented an approach for meta-analysis similar to GWAMA[32]. Namely, a fixed-effect meta-analysis is initially performed for each gene/pathway weighted by the inverse of their variance. In the presence of heterogeneity effects between cohorts tested via Cochran's statistic ($P \leq 0.001$), we adopted a random-effects meta-analysis calculating the random-effects variance component.

Genes and pathways are finally corrected for multiple testing controlling false discovery rate (FDR) using Benjamini-Hochberg procedure for each tissue, removing pathways composed of a single gene and considering each pathway database separately.

Finally, to identify loci harboring associated genes, we defined loci based on gene TSS position, using a window of 200kb in both directions and merging genes with overlapping window or with boundaries not distant more than 1Mb.

GWAS for coronary artery disease

We compare our TWAS and PALAS with two GWAS summary statistics. The first GWAS (simply referred as "GWAS") is a recent meta-analysis of UK Biobank SOFT CAD GWAS with CARDIoGRAMplusC4D 1000 Genomes-based GWAS and the Myocardial Infarction Genetics and CARDIoGRAM Exome[20] downloaded from www.CARDIOGRAMPLUSC4D.ORG. The second GWAS, also called "matched GWAS" is performed on UKBB data set using PLINK (v2.00a2LM) software[33] via --glm option using the same individuals, case-control distribution, covariates as well as SNPs and indels. In both cases, GWAS p-values are adjusted with Benjamini-Hochberg (BH) procedure to be consistent with the correction adopted for TWAS and PALAS results. The first GWAS is used study the novelty of the identified loci from our TWAS. The matched GWAS instead is used to compare GWAS, TWAS and PALAS summary statistics, having kept the same sample size and variants, and to investigate the aggregation of small effects variants into biological mechanisms, i.e. genes and pathways.

Additional pathway-detection methods

We applied other two state-of-the-art strategies to detect significant pathways in CAD.

The first is based on hyper-geometric test using significantly associated genes from TWAS. For each tissue, we considered genes reliable in a tissue as background. For each pathway detected in a tissue based on the reliably expressed genes, we computed an hypergeometric test using fisher-exact test R function (alternative=”greater”). We considered as genes in a pathway those genes that are also reliably expressed in the considered tissue and we intersect this set with the genes FDR 0.05.

The second method is based on MAGMA[34] using a matched GWAS from the UKBB or GWAS results from the summary statistics of a recent large GWAS[35]. MAGMA analysis was performed by first annotating all SNP locations with genes in vicinity using standard parameters and magma –annotate. Subsequently, we performed gene analysis on SNP p-value data using the European reference panel from Phase 3 of the 1000 Genomes project and GO as well as Reactome pathways for subsequent pathway level analysis leaving all parameters at their standard values. Only pathways significant below an FDR of 0.05 were retained for further analysis.

Pathway characterization and prioritization

To further characterize the significant pathways identified, we split them into two classes based on the corresponding genes significance. Let $\Omega$ be a significant pathway with $\text{FDR}(\Omega) \leq 0.05$. Suppose $\Omega$ is defined from $\{g_1, \dots, g_n\}$ genes (called original genes) of which $\{g_1, \dots, g_{\tilde{n}}\}$ $(\tilde{n} \leq n)$ are those also reliable in the tissue considered (called T-score genes) and hence used to compute the corresponding pathway score. We divided pathways into two categories. The first category is composed of pathways with at least one gene more significant than the pathway association, i.e.

it exists a gene $g_i \in \{g_1, \dots, g_{\tilde{n}}\}$ such that p-value $(g_i) \leq$ p-value (Ω). The remaining significant pathways (second category) are then formed by genes all less significant than the pathway itself, i.e. for all $g_i \in \{g_1, \dots, g_{\tilde{n}}\}$ it results p-value$(g_i) >$ p-value(Ω). These are further split in those including at least one gene significant at FDR 0.05 (green) and those having no gene passing FDR 0.05 threshold, hence considered "novel". Pathways in the first category are perturbed by the action one or more strong effect genes with non-concordant effects, whereas pathways in the second category are disrupted by the aggregation of effects, either from putative targets identified from TWAS or from completely weak signals that would be missed using a p-value cut-off strategy, hence novel.

To prioritize the associated pathways (FDR 0.05), we apply the following strategy to focus on more plausible candidates. We select only pathways computed from T-score genes $5 < \tilde{n} \leq 200$ or $3 \leq \tilde{n} \leq 200$ if pathway coverage $\tilde{n}/n \geq 0.1$, that originally included genes $n < 200$ and reaching nominal significance p-value(Ω) $\leq 0.0001$.

Patient stratification based on gene T-scores

For the purpose of stratifying patients based solely on genetically derived data (**Supplementary Fig. S1** Module 3), we adopted a graph-based clustering approach similar to the PhenoGraph method[36] developed in Seurat for single-cell data. Cases are represented as a node and connected to their neighbors via edges with corresponding weights defined as the similarity between individuals. We apply for each tissue the following pre-processing steps to perform features filtering and normalization, and reduce ancestry contribution. First, gene T-scores are clumped at absolute Pearson correlation of 0.9, directly estimated from the considered cases and giving priority to genes that are more significant with respect to the disease of interest. In details, genes are sorted from the most to the least significantly associated with the phenotype of interest (CAD or SCZ) based on the TWAS p-value. All genes are initially assigned to a "current set" and the first gene in this list is compared to all the others based on Pearson correlation estimated from that set of samples, the genes with an absolute Pearson correlation > 0.9 are included in the "remove set". The "current set" is then updated removing the considered genes and the correlated ones above 0.9 threshold and the entire procedure is repeated until "current set" coincides with an empty set. Finally, the set of clumped genes is obtained discarding the genes in the "remove set" from those initially available in the tissue. Second, each gene is standardized removing the average and dividing for sample standard deviation computed across cases ($\frac{x-\mu}{\sigma}$). Third, standardized gene T-scores are independently corrected for the same PCs considered in TWAS/PALAS, taking the residuals of the gene-specific linear model. This step is crucial to reduce the relevance of population structure in the final clustering (see **Supplementary Fig. 12d**). Fourth, the corrected gene T-scores are multiplied by the corresponding Z-statistic for trait association (CAD or SCZ) such that i) differences between patients are enhanced and ii) genes that are more relevant for a certain trait will have a higher impact in the clustering decision, despite retaining all the information. For SCZ clustering on PGC cohorts, the different data sets are merged together via juxtaposition and the same steps descried before are applied, even PCs correction on the merged data set due to PCs estimation on the merged cohorts in PGC wave2. Given the data heterogeneity of the different PGC cohorts, we additionally perform outlier removal. In particular, the four steps previously described are performed and outliers are detected as a union across 10 tissues and 2 clumping strategy (0.9 and 0.1) of samples that deviate beyond median ± 6x s.d. for the first 2 UMAP components[37] (minimum distance = 0.01 and n. of neighbor = 30). These SCZ affected individuals are excluded from further analysis and the pre-processing steps are performed again on the filtered set of samples. Across the 36 PGC cohorts, 35 were used for clustering, filtering 259 outliers for a total of 22,732 cases and 1 cohort (scz_boco_eur, 1,773 cases) was used for external validation. In SCZ analysis, the set of variants of PGC cohorts was not harmonized with UKBB data set that is used to approximate missing phenotype information (see "Risk scores computation"). Thus, to ensure a consistent imputation of the genetic variables, we computed Pearson correlation of impute gene expression and imputed pathway scores between the models built from UKBB and PGC. Genes and pathways are included in the clustering analysis if the correlation between imputation on the reference panels GTEx and CMC between the two genotype-only data sets is higher than 0.8. After pre-processing, we construct a sparse similarity matrix for each pair of samples based on the number of shared nearest neighbor (SNN). We initially computed scaled exponential similarity kernel[38] between samples $i$ and $j$ as

$$K(i,j) = \exp\left(-\frac{ed^2(\boldsymbol{Z}_i, \boldsymbol{Z}_j)}{0.5\sigma_{i,j}}\right)$$

with $ed(\boldsymbol{Z}_i, \boldsymbol{Z}_j)$ the Euclidean distance between normalized gene-level t-scores and

$$\sigma_{i,j} = \frac{mean(ed(\boldsymbol{Z}_i, N_i)) + mean\left(ed(\boldsymbol{Z}_j, N_j)\right) + ed(\boldsymbol{Z}_i, \boldsymbol{Z}_j)}{3}$$

where $mean(ed(\boldsymbol{Z}_i, N_i))$ is the averaged Euclidean distance between sample $i$ its k=30 closest neighbors. Hence, this initial similarity matrix depends already on the local density of the data due to the customized scaling parameter $\sigma_{i,j}$. However, to sparsify the similarity and give information only on the local interactions, we used the similarity kernel defined above to compute the percentage of shared nearest neighbor (SNN) between samples $i$ and $j$:

$$S(i,j) = \frac{|v_i \cap v_j|}{|v_i \cup v_j|}$$

with $v_i$ the set of k=30 nearest neighbor based on $K$. $S$ matrix represents the weight for edges in the patient graph structure. We finally applied Louvain Method[39] implemented in *igraph* R package[40] to detect communities that would maximize modularity based on SNN graph. The number of groups is automatically detected by the algorithm in an unsupervised manner, however it depends on the hyperparamter k. We fixed the number of nearest neighbors to consider a priori 30 since it represented a good compromise between being large enough to estimate the local geometry and small enough to avoid large neighborhoods.

Polygenic risk score computation in CAD cases

To compute polygenic risk score (PRS) for individuals in UKBB related to CAD phenotype, we used PRSice2 software[41] with default parameters. We considered as base and target data sets the UKBB cohort with CAD phenotype. The GWAS results for --base input are the matched GWAS summary statistics as described in "GWAS for coronary artery disease". Distributions among cases and controls division as well as clusters were obtained after standardization of best-fit PRS across all individuals. Of note, the use of the same data set for base (GWAS summary statistic) and target (prediction) cohort leads to overfit in the separation between cases and controls. Nevertheless, the focus of this analysis is not the variance explained by PRS but rather the similar distribution and non-stratification of the identified cluster of cases.

Detection of genes and biological pathways associated with clustering structure

In order to test for genes and pathways associated with detected clustering structure, we considered each tissue separately and test differences of a certain gene/pathway in $gr_g$ versus the remaining patients via Wilcoxon-Mann-Whitney (WMW) test implemented in rstatix R package[42]. In each test, the WMW estimates and confidence intervals are computed corresponding to the median difference of the location parameter (Hodges-Lehmann estimator). Let $G$ be the total number of clusters detected, for each group $g \; in \; 1, \dots, G$ in a tissue, p-values were corrected for multiple comparison using Benjamini-Hochberg procedure to control for false discovery rate. Note that, although the clustering is tissue specific, we tested for differences in gene and molecular pathways across all tissues. Cluster-specific genes were subsequently combined across tissues in loci based on physical location (TSS window 200kb, merged if distance < 1Mb). To identify cluster-specific pathways, we tested only pathways filtered with the following strategy. For each tissue, we considered pathways both in Reactome and GO composed of at least 3 genes and no more than 200 (both original genes and T-score genes in the pathway). These pathways are then clumped giving priority to those with the highest coverage (ratio between T-score genes and original genes) and highest number of genes used to compute the pathway (T-score genes). The resulting set of pathways have a pairwise Jaccard Index not exceeding 0.2.

In addition, we tested pathways in WikiPathway and CommonMind gene-sets[7] in SCZ without this initially filtering but using all the available pathways.

Predict cluster structure and validate gene signature

Similarly to PhenoGraph approach, we implemented a projection method based on the percentage of SNN in order to use the detected clustering structure from one cohort to predict groups on external cohorts such as CARDIoGRAM for CAD and scz_boco_eur for SCZ. In particular, for each cohort we considered only genes used in the clustering model and repeated the gene-specific standardization, correction for PCs and Z-statistic multiplication as described in the clustering pre-processing procedure. The Z-statistic for the projection coincides with the one used in the initial clustering and is obtained from the general TWAS. Then, we computed the percentage of SNN based on the exponential similarity kernel as previously described among each pair of individuals in the combined datasets (model plus external cohort). For each sample in the external cohort, the assigned label is based on the probability that a random walk originating at external sample will first reach a labeled sample in the model clustering for each group $G$. The problem is solved via a system of linear equations based on graph Laplacian of the enlarged sample network and each new sample is then allocated to the group that it reaches first with highest probability, see[36] for details. We evaluated the projected clustering on external cohorts based on i) the fraction of cases assigned to a certain cluster both in model clustering and projected and ii) the correlation among cluster-relevant genes. The latter is computed for each group as the Spearman correlation of WMW estimates for model clustering and external cohort across all tissues, including only genes that are cluster-relevant (FDR < 0.01) in the model. In addition, we estimated the number of reproduced loci in the external cohort using the identified loci of cluster-relevant genes. For each group $g$, we considered each relevant locus and retained the most significant gene in that locus, we then annotated the locus as replicated if the WMW estimate for that gene has the same sign in model and external cohort.

<u>Detection of endophenotype differences across patient strata</u>

To test for differences among trait related endophenotypes across patient clusters, we applied generalized linear models to detect group-specific differences, comparing group $g$ ($gr_g$) versus the remaining samples. More specifically, we applied this strategy for the CAD analysis, leveraging the UKBB deep phenotyping and 637 phenotypes included the following categories: alcohol, arterial stiffness, blood biochemistry, blood count, blood pressure, body size measures, diet, hand grip strength, impedance measures, physical activity, sleep, and smoking (class 1 phenotypes). We also included additional clinical information such as family history, medications, ICD10 diagnosis related to anemia, circulatory system, respiratory system, and endocrine system (class 2 phenotypes). The following phenotypes were excluded: all phenotypes having less than 100 values, binary phenotypes with less than 50 true values and categorical ordinal phenotypes with less than 10 samples in the base category both inside and outside the considered group. Continuous phenotypes were initially standardized $\left(\frac{x-\mu}{\sigma}\right)$. Depending on the nature of the phenotype (continuous, binary or categorical ordinal) and similarly to trait-gene/pathway association, for endophenotype $j$ and group $g$, we applied the following generalized linear model (GLM):

$$pheno_j \sim gr_g + cov_1 + \cdots + cov_l$$

with $gr_g$ a binary n. of cases-vector having 1 in correspondence individuals clustered in group $g$. In both class 1 and 2 phenotypes, the covariates included first 10 PCs, age and sex. Additionally, for class 1 we also corrected for medication usage: pain relief medication (aspirin, ibuprofen, paracetamol), vitamin supplements (A, B, C, D, E, folic acid), mineral and dietary supplements (glucosamine, calcium, zinc, iron, selenium), blood pressure medication, cholesterol lowering medication and insulin usage (part of Fields 6154, 6155, 6179, 6153, 6177). Hence, for each endophenotype $j$ and group $g$ we obtained an estimate of group $g$ impact with respect to all the other cases in the form of adjusted regression coefficient $\beta_{GLM}$ and corresponding p-value tested from normality assumption. Subsequently, we filtered endophenotypes for those that showed evidence for association from PALAS with at least one group associated pathway (pathway-group association FDR $\leq 0.05$) and corrected group-specific p-values considering both class 1 and 2 endophenotypes for multiple testing using the Benjamini-Hochberg procedure.

In case of the hypothesis-driven analysis for CAD, we tested with the same procedure 33 clinical variables among UKBB (BMI, unstable angina pectoris, history of myocardial infarction, coronary artery bypass graft, percutaneous coronary intervention, history of bleeding, heart function severity, hypertension, hyperlipidemia, diabetes, diabetes type 1, diabetes type 2, insulin mediation, peripheral vascular disease, cerebrovascular disease, cerebral stroke, transient cerebral ischaemic attacks, chronic obstructive pulmonary disease, chronic kidney disease, dialysis, atherosclerotic heart disease, poor mobility, pulmonary hypertension, left ventricular ejection fraction, history of cancer, smoking, age of angina diagnosis, age of heart attack, age of stroke, death due to acute myocardial infarction, death due to chronic ischemic heart disease, death due to stroke, age of death) and 2 endophenotypes registered for GerMIFSV (Gensini score and n. of vessel affected). In contrast to the general analysis, clinical variables in UKBB were not converted via PHESANT software but directly used relying on a permutation based p-value. To that end, individuals were randomly assigned to any of the 5 CAD clusters, respecting the original group followed by the same GLM based endophenotype analysis, this was repeated 50 times (see "Patients clustering simulation in CAD" Supplementary Text). We then determined the frequency that a particular clinical variable was nominally (p-value $\leq 0.01$) associated with any of the groups in any of the 50 partitions and used this frequency to determine an empirical p-value by dividing by the number of tests. We then retain only clinical variables with an empirical p-value below 0.01.

For the SHIP Trend cohort, both 20 collected clinical variables (imt_auto_t0, ldlch, hdlch, tg_s, igf1, hba1c, crp_hs_re_z, bmi_t0, bia_magermasse, sysbp_t0, diabp_t0, hyp_t0, mi_first_t0, stroke_first_t0, plaque_t0, stenos_t0, fmd_reduced, abi_pathol, mort_all, mort_cvd) and 24,925 measured gene transcripts across 975 samples were tested with the previously described procedure.

We included as covariates testing group-specific clinical variable differences the first 10 PCs, sex, genotype array type and medication info for blood pressure, cholesterol lowering and insulin. In addition to these covariates, we also included in the cluster-specific measured gene expression analysis RNA integrity number, amplification batch (96 well plates), sample storage time, white blood cell count, hematocrit, red blood cell count, platelet count as well as neutrophils, lymphocytes, monocytes, and basophiles percentages. To compare the differences in actual gene expression with the imputed one, we considered only group-wise significant genes from UKBB at FDR 0.01 in whole blood. Measured transcripts were restricted to the set of group-specific significant genes from UKBB matched by not null ENTREZ gene ID. P-values for adjusted beta in this subset of transcripts were corrected via Benjamini-Hochberg procedure. In addition, we built pathway-scores in SHIP-Trend cohort from the measured gene expression (called measured pathway-scores) and tested group-specific differences via GLM. These measured pathway-scores are obtained in a similar manner to the predicted gene expression but using all measured genes in the whole blood microarray dataset based on the quantile normalized, z-scored residuals after correction for covariates.

For the PsyCourse Study, we tested the following phenotypes using the same GLM based procedure evaluating the following variables: v1_nrpsy_tmt_A_rt, v1_dur_illness, v1_age_1st_inpat_trm, v1_age_1st_out_trm, v1_nrpsy_dg_sym, v1_chol_trig, v1_panss_sum_pos, v1_tms_daypat_outpat_trm, v1_1st_ep, v1_bmi, v1_nrpsy_tmt_B_rt, v1_diabetes, v1_cat_daypat_outpat_trm, v1_cgi_s, v1_nrpsy_mtv, v1_kid_fail, v1_outpat_psy_trm, v1_gaf, v1_stroke, v1_epilepsy, v1_hyperten, v1_nrpsy_mwtb, v1_panss_sum_neg, v1_nrpsy_dgt_sp_bck, v1_fam_hist, v1_nrpsy_dgt_sp_frw, v1_autoimm, v1_ang_pec, v1_heart_att, v1_liv_cir_inf, including Age, Sex, center of patient recruitment and the first two PCs from the genotype analysis as covariates.

Group-specific treatment response analysis in CAD

Taking advantage of the treatment annotation in UKBB data, we investigated whether cases from different genetically detect groups exhibited a different treatment response. For this purpose, we regarded as response phenotypes the categories of arterial stiffness, blood biochemistry, blood count, blood pressure, body size measure, hand grip strength and impedance measures; and we considered as treatments the 17 medications previously described for endophenotype differences analysis (pain relief, vitamin supplements, mineral and dietary supplements, blood pressure medication, cholesterol lowering medication and insulin). Consider group $g$ composed of $n_g$ cases and consider phenotype $j$ values in corresponding of group $g$ ($pheno_j(gr_g)$). Phenotypes with less the 300 available values were excluded, and continuous ones were normalized. The response for medication $i$ (e.g. cholesterol lowering medication) in group $g$ measured based on phenotype $j$ is tested via GLM

$$pheno_j(gr_g) \sim med_i(gr_g) + cov_1(gr_g) + \cdots + cov_l(gr_g)$$

and we denote as $\hat{\beta}_{i,j,g}$ regression coefficient representing treatment $i$ effect on phenotype $j$ in group $g$. We used as covariates first 10 PCs, age, sex as well as all the other treatment binary categories. In order to test differences among treatment-phenotype effects across groups, for each pair of groups $(g, h)$ we evaluated regression coefficient differences using Z-test[43]:

$$Z_{i,j}\,(g,h) = \frac{\hat{\beta}_{i,j,g} - \hat{\beta}_{i,j,h}}{\sqrt{\left(SE\,\hat{\beta}_{i,j,g}\right)^2 + \left(SE\,\hat{\beta}_{i,j,h}\right)^2}}$$

where $SE$ is the standard error for regression coefficient computed from GLM. P-values were computed under the assumption of normal distribution and corrected for multiple testing across all the phenotypes but separately for each group-pair $(g, h)$ and treatment $j$ taken into consideration.

Risk scores computation and differences detection in cases stratification

In order to test for endophenotypic differences in datasets without any endophenotypic information such as PGC cohorts, we developed a strategy to annotate patient with endophenotypes from genetic information using tissue-specific gene-risk scores (gene-RS). For each tissue, gene-phenotype association was estimated (TWAS) as previously described in UKBB for phenotype $j$, obtaining for each gene $n$ association Z-statistic $Z_n^j = \frac{\beta_n^j}{SE\ \beta_n^j}$. Secondly, we filtered redundant genes due to LD structure clumping genes at 0.1 squared Pearson correlation cut-off and giving priority to those with higher genotype $R^2$ imputation. The correlation among genes was estimated via a subset of UKBB samples without CAD HARD diagnosis. Finally, for an external cohort composed of $L$ individuals, gene-RS is defined as the $L$-vector of weighted sum for gene t-scores previously corrected for PCs ($\boldsymbol{T_n}$ $L$-vector, for $n = 1, \cdots, N$) multiplied by gene-phenotype Z-statistic $Z_n^j$:

$$\boldsymbol{RS^j} = \sum_{n=1,\dots,N} \boldsymbol{T_n} Z_n^j$$

Hence, we obtained a continuous risk score that mimics the actual phenotype not available for PGC cohorts, which was then tested for group-specific differences. Namely, PGC cohorts are combined, and each gene is corrected for PCs as described in the clustering procedure. Gene-RS are then computed with phenotype effect estimated from UKBB and standardized. Finally, cluster differences are tested via GLM with gaussian link function including PCs as covariates and considering the partition of SCZ cases previously computed on PGC cohorts. In SCZ analysis, we leveraged TWAS results for 1,000 phenotypes from UKBB among the categories of alcohol use, anxiety, blood biochemistry, blood count, blood count ratio, blood pressure, body size measure, cannabis use, depression, dMRI skeleton, happiness and well-being, mental distress and health, sleep, smoking, social support, susceptibility weighted brain MRI, T1 structural brain MRI, task functional brain MRI, traumatic events. In hypothesis-driven analysis, we specifically investigated cognitive function and used TWAS Z-statistic from numeric memory, pairs matching, prospective memory, reaction time, fluid intelligence, symbol digit substitution, trail making.

The reliability of the gene-RS to estimate the actual endophenotype differences depends on i) the number of samples in the gene-endophenotype association analysis together with the genetic heritability of the phenotype and ii) the effect size of the cluster specific difference. The former was measured in UKBB via F-test statistic: gene-RS ability to model actual phenotype was estimated via nested linear models of phenotype predicted via gene-RS plus covariates or only covariates. The latter was estimated via the absolute value of the regression coefficient from GLM cluster differences for gene-RS ($|\beta_g|$ for $g\ in\ 1, \cdots, G$ groups). Hence, we defined a cluster-reliable non-negative measure (CRM) for each endophenotype $i$ and group $g$ as the product of F-statistic and cluster-specific coefficient: $CRM(j,g) = Fstat_j \cdot |\beta_g|$ (see Supplementary Text for validation).

Pathway analysis and drug response

We utilized gene2drug tool[44] for drug repositioning for each group levering the group-specific signature of up-regulated and down-regulated pathways. Briefly, this method uses genome-wide transcriptional response to treatments of 1,309 small molecules measured on 5 cell lines called Connectivity Map (CMap)[45]. The results across CMap multiple experiments are merged using

Prototype Ranked Lists approach as described in [46] to obtain a summary matrix of measured transcriptional changes (genes) for each tested drug. This information is then converted into pathway expression profiles as signed enrichment score (ES) from Gene Set Enrichment Analysis (GSEA), that indicates how much the expression of genes in a pathway is perturbed by the drug administration [47]. Afterwards, pathways enrichment scores are ranked for each drug according to their p-values, with most significantly up-regulated pathways at the top and down-regulated at the bottom. Given a set of pathways, gene2drug uses these ranked pathway expression profiles to compute for each drug an enrichment score and a p-value via GSEA that represent the extent of those pathways to be up- or down-regulated by each drug. Thus, the aim of gene2drug method is to predict drugs that can target the provided set of pathways. In our application to group-specific pathway signatures, we first removed shared significant pathways across tissues that showed a discordant WMW estimates sign. We used the gene2drug R Bioconductor package (gep2pep) and considered precomputed pathway expression profile of the CMap available from http://dsea.tigem.it/data/Cmap_MSigDB_v6.1_PEPs.tar.gz. We matched the filtered group-specific pathways with those available in the CMap annotation for Reactome ("C2_CP:REACTOME"), Gene Ontology Biological Process ("C5_BP"), Molecular Function ("C5_MF"), and Cellular Component ("C5_CC"). Following the transcription reversion signature principle for drug repositioning [48], we searched for drugs with inhibiting effects ($ES < 0$) of group-specific up-regulated pathways and, vice-versa, activating effects ($ES > 0$) for group-specific down-regulated ones. Thus, for each group and pathway database we performed two analyses, separately providing up-regulated and down-regulated pathway in a group and searching for drugs that target those pathways but induce an opposite effect. The resulting p-values from GSEA were corrected for multiple testing via BH procedure and results with $FDR \leq 0.05$ were retained. Finally, we annotated drug names with ATC codes by matching names, using summary tables generated via https://github.com/fabkury/atcd (date 2021-12-03).

Clustering based on genotype derived principal components

To study the ancestry contribution to tissue-specific clustering, we separately cluster cases (CAD or SCZ) solely based on the PCs derived from genotype data. For CAD, we considered the first 40 PCs available in UKBB data set. For SCZ instead we considered the first 20 PCs available and computed jointly in the PGC cohorts. In both diseases, we separately standardized each PCs to mean 0 and standard deviation 1 and performed Louvain clustering on shared nearest neighbor network built from the available PCs. We then compared the obtained clustering structure to those obtained from the actual tissues via NMI and compared it to the 10,000 random partitions of cases of the same size **(Supplementary Fig. S14c, Supplementary Fig. S22c)**. To investigate the overlap at the single group level, we additionally computed the odds ratio from Fisher's Exact test comparing each pair of groups from PCs and imputed gene expression, namely individuals in $gr_i$ (PC) and outside $gr_i$ (PC) with individuals in $gr_j$ (imputed expression) and outside $gr_j$ (imputed expression) **(Supplementary Fig. S14d, Supplementary Fig. S22d)**. Finally, endophenotype differences in PC clustering was performed via previously described GLM approach but only correcting for age and sex covariates. To compare endophenotype differences, we considered for each endophenotype tested the group reaching highest significance (lowest p-value) and compared -log10 p-value between clustering based on PCs and based on imputed gene expression **(Supplementary Fig. S14f, Supplementary Fig. S22e)**.

Appendix A

We explicit $R^2$ as 1 minus the ratio between the variance explained by the model and the original one:

$$1 - \frac{\| \boldsymbol{Y} - \widehat{\boldsymbol{Y}} \|_2^2}{\| \boldsymbol{Y} - \bar{Y} \|_2^2} = \frac{\| \widehat{\boldsymbol{Y}} - \bar{Y} \|_2^2 + 2 \langle \boldsymbol{Y} - \widehat{\boldsymbol{Y}},\ \widehat{\boldsymbol{Y}} - \bar{Y} \rangle}{\sigma_{\boldsymbol{Y}}^2}$$

with $\widehat{\boldsymbol{Y}} \coloneqq X\widehat{\boldsymbol{\beta}} + Z\widehat{\boldsymbol{\mu}}$ the predicted gene expression, $\bar{Y}$ the mean original gene expression, $X$ the cis-variant dosage matrix for the gene in consideration and $Z$ the covariate matrix also including all-one vector to account for intercept term.

Let $\widehat{\boldsymbol{W}} \coloneqq X\widehat{\boldsymbol{\beta}}$ be the predicted genotype effect, $\boldsymbol{W} \coloneqq \boldsymbol{Y} - Z\widehat{\boldsymbol{\mu}}$ the gene expression vector corrected for the confounder effect and $\bar{W}$ the corresponding mean, $\widehat{\boldsymbol{V}} \coloneqq Z\widehat{\boldsymbol{\mu}}$ the predicted confounder contribution and $\bar{\hat{V}}$ the corresponding mean. Thus by definition, $\boldsymbol{Y} = \boldsymbol{W} + \widehat{\boldsymbol{V}}$ and $\bar{Y} = \bar{W} + \bar{\hat{V}}$, hence the first term of $R^2$ nominator can be written as

$\| \widehat{\boldsymbol{Y}} - \bar{Y} \|_2^2 = \| \widehat{\boldsymbol{W}} + \widehat{\boldsymbol{V}} - \bar{W} - \bar{\hat{V}} \|_2^2 = \| \widehat{\boldsymbol{W}} - \bar{W} \|_2^2 + \| \widehat{\boldsymbol{V}} - \bar{\hat{V}} \|_2^2 + 2\langle \widehat{\boldsymbol{W}} - \bar{W}, \widehat{\boldsymbol{V}} - \bar{\hat{V}} \rangle$. Since by definition $\boldsymbol{Y} - \widehat{\boldsymbol{Y}} = \boldsymbol{W} - \widehat{\boldsymbol{W}}$, the second term of $R^2$ nominator becomes

$$\langle \boldsymbol{Y} - \widehat{\boldsymbol{Y}}, \widehat{\boldsymbol{Y}} - \bar{Y} \rangle = \left\langle \boldsymbol{W} - \widehat{\boldsymbol{W}}, \widehat{\boldsymbol{W}} + \widehat{\boldsymbol{V}} - \bar{W} - \bar{\hat{V}} \right\rangle = \langle \boldsymbol{W} - \widehat{\boldsymbol{W}}, \widehat{\boldsymbol{W}} - \bar{W} \rangle + \left\langle \boldsymbol{W} - \widehat{\boldsymbol{W}}, \widehat{\boldsymbol{V}} - \bar{\hat{V}} \right\rangle$$

Hence, $R^2$ can be expressed as

$$\frac{\| \widehat{\boldsymbol{W}} - \bar{W} \|_2^2 + 2\langle \boldsymbol{W} - \widehat{\boldsymbol{W}}, \widehat{\boldsymbol{W}} - \bar{W} \rangle + \| \widehat{\boldsymbol{V}} - \bar{\hat{V}} \|_2^2 + 2\left\langle \boldsymbol{W} - \bar{W}, \widehat{\boldsymbol{V}} - \bar{\hat{V}} \right\rangle}{\sigma_{\boldsymbol{Y}}^2}$$

which we grouped in 3 components $R_g^2, R_c^2$ and $R_{g,c}^2$.

### Supplementary Text

Validation and comparison of Priler against elastic-net regression

Since PriLer is an extension of elastic-net regression (enet) that incorporates prior knowledge on individual variants, we initially benchmarked PriLer against enet across 34 tissue-specific models (33 GTEx and 1 CMC). First, we compare PriLer and enet in terms of reliable genes i.e. genes predicted from genetic data having $R^2 \geq 0.01$ and $R^2_{cv} > 0$. The number of reliable genes is very similar (**Supplementary Fig. 3a**) but always higher for PriLer for a total of 2,922 additional genes (mean ± sd: 85.94 ± 47.39). In addition, for reliable genes in PriLer, we observed an increase in number of genes having higher $R^2_{cv}$ in PriLer compared to enet (**Supplementary Fig. 3b**), showing an overall better prediction performance. The number of genes with improved prediction performance is partly correlated with number of priors used in the model across tissues (Pearson corr. 0.48) and negatively with the number of training samples (corr. -0.28). PriLer not only increases the number of genes that can be accurately predicted, but also decreases the number of reg-SNPs (**Supplementary Fig. 3c**), with a total decrease across all genes of 1,462,466 variants (mean ± s.d. 43,014 ±14,530). The difference in number of reg-SNPs significantly depends on the number of prior features (corr. -0.68). Moreover, we observe an increase in fraction of reg-SNPs that contain any prior information used in PriLer model (**Supplementary Fig. 3d**). The mean increase is 11% (sd 3.32%) with the difference in fraction of reg-SNPs with prior being partly dependent on the number of prior included (corr. 0.26). In addition, we compared reg-SNPs robustness in whole blood tissue, downsampling to 100 individuals 10 times and comparing reg-SNPs selection in each pair of repetition using Jaccard index (**Supplementary Fig. 3e**), PriLer shows a significant increase in terms of concordance of selection with respect to enet (Wilcoxon-Mann-Whitney P=2.8e-14). In summary, PriLer generates better performing models of genotype-based expression imputation, using a reduced amount of variants but more biologically meaningful and robust compared to elastic-net regression without prior information.

Finally, we observe the differences in terms of predictive performances for heritable and not heritable genes defined a priori via GCTA software in PriLer. The majority of expressed genes are not heritable across all tissues (**Supplementary Fig. 4a**). Thus, prior weights are calibrated on a smaller set of genes whose size varies with the training sample size. On the other hand, as expected heritable genes constitute most of the reliable genes defined by PriLer **(Supplementary Fig. 4b**), and the variance explained for heritable genes is always significantly higher compared to not heritable ones in the same tissue (**Supplementary Fig. 4c**, Wilcoxon-Mann-Whitney p-value < $2.21^{-49}$). Overall, median prediction accuracy of heritable-vs-non heritable genes differs by 0.0398 on average, inversely dependent on overall training sample size (Spear. correlation = -0.8128) and ranging from 0.0125 for whole blood to 0.077 for spleen. Similarly, the proportion of heritable-vs-non-heritable genes depends on sample size (Spear. correlation = 0.8105), with proportions ranging from 49% for hippocampus to 78% for thyroid.

Evaluation of prior weights selection in PriLer through random prior simulation

To examine whether the learned weights for prior features were meaningful for the model tissue considered, we simulated random prior features using as example artery coronary tissue and focusing on *heart_left_ventricle* prior features that indicate whether a variant is located in an open chromatin position based on H3K27ac for heart left ventricle cell type. We define as baseline prior 7 priors that are normally adopted in artery coronary model tissue (**Data S1**).

First, we define two new prior features called heart_left_ventricle_Var_random and heart_left_ventricle_Var_random2x randomly selecting variants in the same size or twice respectively of the original prior feature heart_left_ventricle (**Supplementary Fig. 5c**). The aim is to emulate a prior that is not biologically meaningful but contain the same amount of information or twice of an existing one. The estimates for prior weights across 50 repetitions are close to zero although different from it (mean ± sd = 0.0145 ± 2.04e-03 and 0.0317 ± 3.22e-03) (**Supplementary Fig. 5a**) with Var_random2x increased compared to Var_random but still lower than the original prior heart_left_ventricle (mean ± sd = 0.109 ± 4.01e-04). Indeed, when a prior feature intersects SNPs that are used even to a small extent in a gene regression model, the initial estimate cannot be exactly zero and the bigger the prior size (number of variants it intersects), the more likely is that prior to be relevant just because of randomly intersecting reg-SNPs. In addition, just by chance, the variants randomly selected still intersects baseline prior features that are used in the model (mean sharing 20%, **Supplementary Fig. 5b**). However, in the iterative procedure, the weights for the randomly created priors remain fixed instead of increasing until convergence as it happens for the original prior (**Supplementary Fig. 5d**). This means that the use of variants intersecting heart_left_ventricle in the gene regression models increases the performance, which does not happen for the randomly generated priors.

Second, we generated random prior features that resemble ChIP-Seq H3k27ac data used to build prior information. To this end, we randomly select open chromatin regions i.e. gene regulatory elements (GREs) from the original data in the same size or twice as *heart_left_ventricle* and intersected with variants location to create *heart_left_ventricle_Epi_random* and *heart_left_ventricle_Epi_random2x* priors. In addition, we included *Ctrl_150_allPeaks* which is a prior feature related to brain tissue. Differently from the first scenario that just extrapolates variants by chance, sampling GREs allows taking into consideration genomic positions and LD structure. The randomly selected GREs across 50 repetitions partly overlap with baseline GREs used in artery coronary tissue (**Supplementary Fig. 5f**) resulting in a sharing of 67% and 65% of variants in *Epi_random* and *Epi_random2x* respectively as well as 81% shared variants with *Ctrl_150_allPeaks*. Thus, to generate a random prior that would not show a high sharing with the baseline model, we randomly selected GREs excluding the ones used in the baseline prior features and in the same size as GREs for *heart_left_ventricle*. The newly created prior feature (*Epi_random_noint*) only shares 20% of the variants detected among the baseline priors due to GREs possible overlapping. The number of variants from randomly generated priors are similar to the original *heart_left_ventricle* for *Epi_random* and *Epi_random_noint* while twice the amount for *Epi_random_2x* (**Supplementary Fig. 5g**). Differently from the first scenario, the estimate for prior weights *Epi_random Epi_random_2x* and *Ctrl_150_allPeaks* are very different from zero (mean ± sd = 0.06 ± 0.005, 0.096 ± 0.004, 0.052 ± 0.001). The only random prior reaching the similar weight as *heart_left_ventricle* (0.095 ± 0.002) is *Epi_random_2x*, which includes twice the amount of the information than the original (**Supplementary Fig. 5e**), while *Epi_random_noint* estimates are very close to zero (0.01 ± 0.0004). Although the new prior included in the model are not related to artery coronary, the relevance can be explained by the high sharing in terms of variants with respect to the baseline model. Indeed, when the percentage is reduced as in the case of *Epi_random_noint,* the associated weight is close to zero. Regardless *Epi_random_2x* starting at higher relevance due to the increased size, it just reaches the same value of original *heart_left_ventricle* at convergence (**Supplementary Fig. 5h**).

We conclude that the weights reflect a tissue specific configuration of gene expression regulation that can be partially confounded by high sharing of variants with actual relevant prior features. However, not relevant prior weights are reduced to the minimum when their sharing with relevant priors is only marginal, even in case of existing GREs reflecting genome structure.

Comparison of Priler against existing methods: TWAS and prediXcan

We compared PriLer to prediXcan[49] and TWAS[50] methods build on GTEx v6p and CMC datasets. Summary of tissue models for prediXcan are downloaded from https://s3.amazonaws.com/predictdb2/deprecated/download-by-tissue-HapMap/ and https://github.com/laurahuckins/CMC_DLPFC_prediXcan/blob/master/DLPFC_oldMetax.db.tar.gz and for TWAS from https://data.broadinstitute.org/alkesgroup/FUSION/WGT/GTEx.ALL.tar and https://data.broadinstitute.org/alkesgroup/FUSION/WGT/CMC.BRAIN.RNASEQ.tar.bz2. In order to compare PriLer performance with previous methods we used $cor_{cv}^2$ defined as squared correlation between $\boldsymbol{W}_{\boldsymbol{test}}$ and $\widehat{\boldsymbol{W}}_{\boldsymbol{test}}$ defined as adjusted gene expression and predicted expression from genetic effects respectively combing all test folds. Since we restrict our analysis to Caucasian only, the number of individuals used in PriLer is lower (mean decrease 22 ± 17 and 19 ± 18 respect prediXcan and TWAS), slightly decreasing the overall power. We then consider only genes in PriLer having any 200kb cis-variants and being also present in prediXcan or TWAS summary statistics. Combining all the tissues together, PriLer shows an increase in term of predictive performance: the percentage of genes with higher $cor_{cv}^2$ in PriLer is 64.6% compared to prediXcan out of 158,249 and 76.6% compared to TWAS out of 68,891 (**Supplementary Fig. 6a-b**). The number of genes that uses more reg-SNPs in PriLer is instead similar to prediXcan (50.1% higher in PriLer) but particularly greater compared to TWAS (80.5% higher in PriLer). A possible reason for this increase is that TWAS choses the best model among 5 different ones which also include best eQTL. Therefore, PriLer outperforms TWAS and prediXcan even including a reduced amount of training samples and with a number of selected variants that is higher only compared to TWAS, resulting in an even improved $cor_{cv}^2$. Similar to the comparison with elastic-net regression, the fraction of reg-SNPs intersecting tissue specific prior information is increased for PriLer (**Supplementary Fig. 6c**). In order to externally validate the enrichment of reg-SNPs for PriLer in biologically meaningful regions, we use recently annotated map of DNase I hypersensitive sites (DHSs) across 733 human biosamples encompassing 438 cell and tissue types and states [51], including reg-SNPs from reliable genes for PriLer and enet as well as reg-SNPs in TWAS and prediXcan. Intersecting their location with DHSs, each reg-SNP is annotated with the number of biosamples it overlaps with respect to a DHS. We tested the differences between PriLer and enet, prediXcan and TWAS in term of distribution of number of reg-SNPs intersecting DHSs biosamples using Kolmogorov-Smirnoff test (**Supplementary Fig. 6d**). Although TWAS shows the highest percentage of reg-SNPs intersecting at least 1 biosample DHSs, PriLer displays an increased percentage of reg-SNPs intersecting multiple biosample DHSs with a significant improvement for brain hippocampus, heart left ventricle and muscle skeletal tissues. Interestingly, these tissues already showed the strongest improvement in term of increased fraction of reg-SNPs having prior information for PriLer with respect to the other methods (**Supplementary Fig. 3d, Supplementary Fig. 6c**). In summary, we developed a gene expression imputation method that offers multiple and incremental improvements over existing strategies.

<u>Calibration of type 1 error in TWAS and PALAS</u>

In order to determine whether the approach to TWAS and PALAS proposed here provided well-calibrated p-values both at the gene and pathway-score levels, we considered whole blood as exemplar tissue that included 3,840 genes, 902 Reactome pathways and 2,803 GO pathways and simulated random phenotypes 50 times. In detail, we created binary vectors that resembled CAD phenotype keeping the same case/control size, i.e. 19,026 cases and 321,913 controls. To create random phenotypes that resembled as closely as possible the same confounders as the actual CAD classification, we selected the same number of female/males and the same age compared to the actual CAD phenotype among the case/control classes. We then performed TWAS and PALAS and tested for associations between the randomly created phenotypes and gene T-scores and pathway-scores that were previously computed for CAD (i.e. considering as reference set a subset of individuals non-affected by CAD). Finally, multiple-testing correction is performed via BH procedure, correcting for each simulation separately. Combing all the simulations, we observed that p-value distribution approximates a uniform distribution in (0,1) range (**Supplementary Fig. 8a-c**), validated also via Kolmogorov-Smirnoff test that compared a random uniform distribution with the simulated one form gene associations (p-value=0.17), pathway associations in Reactome (p-value=0.87) and pathway associations in GO (p-value=0.5). The same conclusions can be drawn from quantile-quantile plots in **Supplementary Fig. 8d-f,** with the expected distribution of p-value extracted from a uniform one. The association signal with the actual CAD phenotype greatly diverged from the simulated ones, with very few genes/pathways passing the FDR 0.05 threshold in the simulated phenotypes (blue points). However, all simulation results remain in the 95% confidence intervals of the standard uniform order statistics that follows a beta distribution. We can then conclude that CASTom-iGEx strategy for TWAS and PALAS returns well-calibrated p-values.

Relevance of genes correlation in pathway significance

Next, we investigated whether gene correlation and LD structure were connected to the increase observed in pathway-score significance. To this aim, we performed two analyses:

1. Simulation of pathway structure from actual gene T-scores in whole blood, creating gene-sets composed of 3 or more genes located in the same loci, for a total of 46 simulated pathways. In this case, the goal was to understand how the loci structure can influence the pathway significance.

2. Estimation of relationship between pathway significance increase and average gene correlation across all detected pathways with $n \geq 2$, to observe the actual relevance and extent of genes correlation in pathway significance.

For 1., we only considered actual genes in whole blood that were showing a certain level of significance i.e. nominal TWAS p-value $\leq 0.01$ and created simulated gene-sets from those genes that were also in the same loci and had the same effect size sign in CAD associations (all Z-stat genes >0 or < 0), to avoid a compensatory effect for gene relevance. This procedure led to a total of 46 simulated pathways with the number of genes included varying from 3 to 7. Although all the genes were in the same loci, the increase in pathway significance was dependent and inversely proportional to the estimated average genes correlation (**Supplementary Fig. 10a**), with almost no increase for pathways that included highly correlated genes, This resulted in a general lower significance of pathways composed of correlated genes (**Supplementary Fig. 10b**). Based on these findings we concluded that the gene correlation due to the regulation from the same variants (or in LD with them) rather than the vicinity of genomic coordinates is relevant in the observed pathway significance and that genes in the same loci not correlated do still lead to an improvement in the information captured by the pathway scores.

For 2., we finally considered the actual pathway-scores and increase or decrease in pathway association level compared to the average genes correlation included in the pathways. Across all the pathways databases, there was no rank correlation between average differences of significance in pathways versus genes and genes correlation (**Supplementary Fig. 10c-e**, absolute Spearman corr.

< 0.045). Indeed, we observed that pathways with highly correlated genes (> |0.5|), usually including less than 4, showed only marginal improvement in pathway significance. In contrast, pathways with a striking effect of increased significance were those formed by more than 10 genes and having and average correlation around zero. Hence, we conclude that the increase in pathway relevance with respect to single genes became maximal when the correlation among genes was minimal. Overall, genes correlation due to LD structure did not increase pathway significance nor pathway improvement compared to single genes. Finally, observing actual pathway structures, the gene-sets with best improvement were formed by not correlated genes.

Training sample size effect on TWAS and PALAS results for CAD

To assess the robustness of TWAS and PALAS results based on the reference panel sample size, we performed a down-sampling analysis and applied PriLer on 50% 70% and 90% of samples in artery aorta (AA) and heart left ventricle (HLV) tissues. Afterwards, we imputed gene expression on same UKBB data set used for CAD, converted them into gene T-scores and pathway-scores. Finally, we tested the reliable genes and computed pathways for association with CAD phenotype as previously described.

Consistently to what was observed across different tissues, the number of reliable gene decrease with the decrease of the same size (**Supplementary Fig. 11a**). When comparing model performances in terms of $R^2_{CV}$, considering all reliable genes the $R^2$ estimates remained stable across sampling percentages but drastically improved for those genes in common (**Supplementary Fig. 11b**). This indicates that increasing the sample size 1) we can reliable predict more genes that have a weaker genetic component of expression regulation and 2) we can better estimate the genetic dependencies of more heritable genes. We then predicted gene expression on UKBB for the corresponding reliable genes and computed gene T-scores and pathways scores as previously explained. Finally, we run TWAS and PALAS analysis, testing for CAD associations. The percentage of shared genes as well as the correlation of genes Z-statistics from TWAS were higher for the same tissue across sub-sampling percentages, with an increase when increasing the sampling percentage (**Supplementary Fig. 11c**). The trend was similar for pathways associations but with a general lower correlation (**Supplementary Fig. 11d**). This is due to the fact that the same pathway across sub-sampled tissues can have different coverages and can be composed of a different gene set, depending on the tissue-specific reliable gene set.

Finally, we investigated the concordance in prediction of significant genes and pathways (FDR 0.05) across the sub-sampled models compared to the full model (100%) using all samples (**Supplementary Fig. 11e-h**). For each comparison (sub-sampled vs full model), we considered only predicted genes/pathways in common and we computed the receiver operating characteristic (ROC) curve and the corresponding area under the curve (AUC) considering significant genes/pathways as real positives, and absolute value of CAD Z-statistic in each sub-sampled model as prediction score. The concordance is particularly high for TWAS results, with AUC > 0.99 for all the comparison in both tissues and increasing with the training sample size (**Supplementary Fig. 11e,g**). A similar increase is observed for PALAS results but with a lower prediction performance, leading to a AUC > 0.73 in aorta and > 0.6 in left ventricle (**Supplementary Fig. 11f,h**). In conclusion, a decrease in sample size leads to a lower number of reliable genes and worse model performance, as expected. The TWAS and PALAS association are highly correlated across sub-sampled models in the same tissue, although less for pathways than genes. Similarly, the prediction of significant genes is highly consistent between the full and the sub-sampled models, however lower performances were observed for pathways. This is related to the difference in considered gene-sets given fewer genes that can be reliably estimated at lower sample size. Nevertheless, these trends (correlation and prediction) are increasing with the sub-sampled training size.

<u>Clustering simulation in CAD</u>

To generate an empirical null-distribution of gene, pathway, and endophenotype associations with clustering structure, we randomly partitioned UKBB CAD patients 50 times into similar sized groups compared to actual liver-based clustering. All random partitions but one were independent (**Supplementary Fig. 16a**), with repetition 2 only showing a mild association (P=0.0063) and not passing FDR 0.05 threshold (FDR = 0.32). Considering the first 10 repetitions due to computational time constraints, we then detected the group-specific genes and pathways across clusters. The WMW p-value distributions mostly did not deviate from the expected uniform distribution (**Supplementary Fig. 16b,d**) with some exception for genes such as repetition 9 in group 5. Observing the number of association passing FDR thresholds, across each group the 0.01 upper bound identifies 1 gene significant in 1 out of 10 repetitions (**Supplementary Fig. 16c**) and 1 pathway significant in at max 2 repetitions (**Supplementary Fig. 16e**). Thus, to reduce the number of false-positives, we used as FDR threshold for cluster-specific genes and pathways of 0.01 instead of the otherwise used 0.05. Finally, we computed the endophenotype differences in each cluster via GLM across the 50 random clustering testing 637 UKBB phenotypes. We compared the effect size $\beta_{GLM}$ and the corresponding -log10 p-value from random clustering repetitions and liver cluster (**Supplementary Fig. 16f**). Extremely significant results were only achieved for liver clusters and maximum 1 endophenotype was significant (FDR 0.05 ) in 7 different repetitions. In conclusion, FDR cut-off of 0.01 ensures a reduction of false positives for cluster-specific genes and pathways. Moreover, the empirical null-distribution of endophenotype associations leads to almost no significant results and greatly different compared to the actual clustering in liver. Endophenotype associations in random clusters were also tested for the 33 hypothesis-driven clinical phenotypes and used to compute the empirical p-value (see "Detection of endophenotype differences across patient strata" in Methods).

Investigation of ancestry contribution to clustering structure

To reduce possible biases in clustering structure given by ancestry, we correct imputed gene expression for 10 genotype-based PCs as pre-processing step prior clustering. in the context of CAD, we observed that clustering of the data on the residuals compared to no correction showed no/minimal remaining associations with PCs (**Supplementary Fig. 12d**). This step only minimally changed the clustering structure compared to no correction, indicating that the overall impact of population structure as captured by PCs was already small (NMI > 0.5, **Supplementary Fig. 12e**). In the context of CAD clustering in liver and SCZ clustering in DLPC, PCs association reduced after correction but was still significantly associated with the detected clusters (**Supplementary Fig. 13c, Supplementary Fig. 21c**), with a stronger effect in SCZ. Nevertheless, in both CAD and SCZ we observed that the actual PC distribution across clusters (**Supplementary Fig. 13c, Supplementary Fig. 21c**) was not driving the partitioning compared to the imputed gene expression based on effect size estimates (30-50 fold differences between each contributing gene and PCs **Supplementary Fig. 13d, Supplementary Fig. 21e**), hence not separating the patient space. In fact, the coefficients of variations (effect-sizes divided by confidence interval range) specific to each group for the strongest associated PCs (PC4-PC5 in CAD and PC1-PC5 in SCZ) were below 0.92 and 2.6 in absolute value compared to the top 5 genes per group (P < 1e-100) that showed a coefficient of variation > 4 and 23 respectively, with a peak of 300 for SORT1 and 85 for C4A, respectively for CAD and SCZ **(Supplementary Fig. 13d, Supplementary Fig. 21e**). To better characterize the PCs contribution, we performed clustering of individuals based solely on PCs and repeated the endophenotype analysis (see "Cluster cases from genetic principal components" in Methods, **Supplementary Fig. 14, Supplementary Fig. 22**). Both for CAD and SCZ, we found marginal overlap between tissue based clusters and PCs based ones (NMI < 0.0052, **Supplementary Fig. 14b, Supplementary Fig. 22b),** although significant based on chi-squared test and with a stronger signal in SCZ. Specifically in liver for CAD and DLPC for SCZ, the minimal overlap was not null and greater than what is reached by a randomly assigned clustering structure (**Supplementary Fig. 14c, Supplementary Fig. 22c**). To understand which groups from tissue based and PCs based shared a higher by chance number of individuals, we compared pairwise Fisher's Exact test odds ratio (**Supplementary Fig. 14d, Supplementary Fig. 22d**). For CAD, an enrichment was detected between gr7 in PCs and gr1 in liver, with a consequential depletion between gr3 in PCs and gr1 in liver. This could be related to a higher fraction of samples in gr7 PCs and gr1 liver originally from Reading and Birmingham surroundings (**Supplementary Fig. 14e**). For SCZ instead, 10 pairs showed either a significant enrichment or depletion (p-value < 0.01), with strongest enrichment among gr2 in DLPC and gr5 in PCs (**Supplementary Fig. 22d**). Most importantly, we observed that the minimal overlap found between tissue-derived and ancestry-derived clustering did not influence the group-specific endophenotype differences (**Supplementary Fig. 14f-g**, **Supplementary Fig. 22e-f**). For CAD, we observed different endophenotype significance among the two clustering structures (**Supplementary Fig. 14e**), with place of birth in UK being the strongest signal in PCs clustering not significant for liver clustering, as expected (**Supplementary Fig. 14f**). For the three endophenotypes significant in both clustering configurations (height, comorbidity with lipidaemia and aspartate aminotransferase), we additionally examined whether this was related to the mild overlap between gr1 in liver and gr7 in PCs. Looking at cluster-specific effect-sizes (**Supplementary Fig. 14g**), hyper lipidaemia diagnosis showed an opposite effect in the two not enriched nor depleted groups (gr5 in liver and gr2 in PCs), height strongest associations were referring to overlapping gr1 liver and gr7 PCs but with an opposite effect, and aspartate aminotransferase was strongest in two depleted groups (gr3 PCs and gr1 liver) with an opposite effect, thus being the only concordant result with clustering overlap (OR = 0.86, P = 0.0002). Similarly, for SCZ the trend among best p-value endophenotype association was different between DLPC and PCs cluster (**Supplementary Fig. 14e)** and with a great variability in magnitude. However, 6 endophenotypes passing FDR 0.05 threshold were identified in both partitions. Considering results with FDR 0.1 threshold, we then investigated the group-specific differences for the strongest association in each endophenotype (**Supplementary Fig. 14f**). Platelet crit is lowest in two not enriched nor depleted groups (DLPC gr1 and PCs gr2), similar to LDL direct and apolipoprotein B decreased in DLPC gr1 and PCs gr3 as well as the following diffusion magnetic resonance imaging (dMRI) phenotypes: Mean L1 in fornix cres+stria terminalis on FA skeleton (left), Mean L3 in cingulum hippocampus on FA skeleton (right) and Mean MD in fornix cres+stria terminalis on FA skeleton (left). The remaining dMRI phenotypes showed strongest but opposite effect for DLPC gr3 and PCs gr3 that again show evidence of no significant overlap nor depletion. The only endophenotype with concordant result based on group overlap was Volume of grey matter in Inferior Frontal Gyrus that had strongest associations with concordant sing in PCs gr4 and DLPC gr3 and were actually enriched for shared individuals. Jointly, these results show that patient groups and detected endophenotype differences in our analysis were not driven by PCs as would be expected if population structure had a major impact on overall clustering.

Incremental effect from pathway-scores: case study "De novo loss-of-function" gene-set

In order to evaluate the power gain using gene set based aggregation in more detail, we performed a high resolution analysis of the De novo LoF gene-set in DLPC in SCZ. This gene-set is a collection of genes harboring rare variants detected in probands from multiple SCZ family studies. In DLPC, this pathway was composed of 35 genes and reached a significance of $P = 2.92e-07$, exceeding that of any individual gene (genes $P \geq 2.29e-05$). We sorted the 35 pathway member genes with respect to SCZ Z-statistic and added one gene at a time to the gene-set structure, computing at each increment the gene-set association with SCZ (**Supplementary Fig. 19e**). This analysis showed that an increment in the pathway level significance corresponding to the best incremental gene-set configuration was achieved when adding same directional effect genes even with very low effect (i.e. until nominal $P < 0.1$), supporting notion of the importance of the small effect variants in SCZ architecture. In addition, significant opposite sign association can disrupt the overall pathway signal. For instance, the overall pathway significance drastically decreased when adding ALMS1 gene that was positively and significantly associated with SCZ, hence with an opposite sign with respect to the majority of genes (**Supplementary Fig. 19e**). On the other hand, genes with a negative Z-statistic but not associated with SCZ even at the nominal level contributed only to the gene-set signal and thus slowly decreased the overall level of significance (from NEB to ULF1 genes). Importantly, the considered genes were independent and mostly located in different loci with only ALS2CL and NCKIPSD both in 3p21.31 and indeed showing the highest interaction (Pearson corr.= −0.03, data not shown).

Validation of gene risk scores to mimic actual phenotype in cluster-specific differences

To evaluate the reliability of cluster-specific differences for gene-RS in term of actual differences in corresponding endophenotype, we defined a cluster-reliable measure (CRM). We calibrated a reasonable threshold for CRM based on CAD analysis and UKBB phenotyping. This threshold is subsequently applied to SCZ cluster-specific gene-RS differences to highlight significant results that are likely to be observed also in the actual phenotype.

First, we build gene-RS weights (Z-statistics) for 369 CAD related phenotypes in UKBB in 10 GTEx tissues. Then, we compute gene-RS separately on each CARDIoGRAM cohort and tissue, correcting each gene for the cohort-specific first 10 PCs and considered the projected clustering structure based on UKBB CAD tissue-specific results. Via meta-analysis similar to TWAS and PALAS, gene-RS group-specific differences across all cohorts are summarized and CRM for each group-endophenotype combination is computed as described in Methods ("Risk scores computation and differences detection in cases stratification") for those passing FDR 0.05 cluster-specific significance. Success rate of gene-RS reliability, i.e. actual endophenotype differences detected for the same clustering structure in UKBB, is measure in term of precision (**Supplementary Fig. 24**). This is computed based on the fraction of group-specific gene-RS differences having same sign of $\beta_{GLM}$ in gene-RS and actual endophenotype analysis in UKBB among all the endophenotypes passing a certain CRM threshold:

$$Precision = \frac{\#(\beta_{GLM}^{gene-RS} \cdot \beta_{GLM}^{pheno} > 0 \quad \wedge \quad CRM_{gene-RS} > thr_{CRM})}{\#(CRM_{gene-RS} > thr_{CRM})}$$

Combining all tissues together, CRM cut-offs on CARDIoGRAM of 610 or 265 lead to precision > 0.85 or > 0.8 respectively (**Supplementary Fig. 24a)** for CAD. A similar trend was observed when comparing cluster-specific results from gene-RS and endophenotype on UKBB (**Supplementary Fig. 24b)**, with increased precision performances having estimated F-statistic on the same samples where actual endophenotypes where measured. Thus, we adopted those thresholds to define strongly reliable and reliable cluster-specific results in SCZ.

Application of CASTom-iGEx to non-european individuals in UKBB

To test trans-ancestry performances of CASTom-iGEx trained on European samples, we applied CASTom-iGEx pipeline to individuals from UKBB of Indian origins. In particular, we filtered samples using ethnic background (data-field 21000) coded as "Indian" (code 3100), being the largest non white British population. In addition, we removed individuals that withdraw consent, non-imputed ones, having a discordant genetically inferred and reported gender as well as relatives up to 3rd degree. The final cohort included 5,236 individuals among which 461 were satisfying the CAD HARD definition (see Methods). We considered only variants used for CAD analysis in UKBB white British cohort harmonized with GTEx v6p reference panel and CARDIoGRAM cohorts, matched by SNP IDs. On this genotype-only dataset, we imputed gene expression across the 10 CAD related tissues trained on GTEx v6p European samples and performed TWAS and PALAS testing for CAD phenotype. Fraction of concordance based on Z-statistic sign for UKBB white British (UKBB WB) significant results indicated an overall mild replication (< 0.65) combining all tissues, that however was not significant in some of the tissues (**Supplementary Fig. 28a**) and lower than the replication reached in the European based CARDIoGRAM meta-analysis (**Supplementary Fig. 28b**). In addition, we projected Indian UKBB cohort into the clustering structure computed on UKBB WB in liver. The fraction of cases assigned to each group differed in UKBB Indian from clustering model more than what was observed across CARDIoGRAM cohorts (**Supplementary Fig. 28c-d).** Similarly, the Spear. correlation of cluster-specific genes was different from null but strongly reduced in UKBB Indian compared to CARDIoGRAM (**Supplementary Fig. 28e**). In conclusion, the performances and replications were overall poor when using CASTom-iGEx European trained models on different ancestry population.

**List of the Schizophrenia Working Group Psychiatric Genomics Consortium members**

Stephan Ripke[1,2], Benjamin M. Neale[1,2,3,4], Aiden Corvin[5], James T. R. Walters[6], Kai-How Farh[1], Peter A. Holmans[6,7], Phil Lee[1,2,4], Brendan Bulik-Sullivan[1,2], David A. Collier[8,9], Hailiang Huang[1,3], Tune H. Pers[3,10,11], Ingrid Agartz[12,13,14], Esben Agerbo[15,16,17], Margot Albus[18], Madeline Alexander[19], Farooq Amin[20,21], Silviu A. Bacanu[22], Martin Begemann[23], Richard A Belliveau Jr[2], Judit Bene[24,25], Sarah E. Bergen [2,26], Elizabeth Bevilacqua[2], Tim B Bigdeli [22], Donald W. Black[27], Richard Bruggeman[28], Nancy G. Buccola[29], Randy L. Buckner[30,31,32], William Byerley[33], Wiepke Cahn[34], Guiqing Cai[35,36], Murray J. Cairns[39,120,170], Dominique Campion[37], Rita M. Cantor[38], Vaughan J. Carr[39,40], Noa Carrera[6], Stanley V. Catts[39,41], Kimberly D. Chambert[2], Raymond C. K. Chan[42], Ronald Y. L. Chen[43], Eric Y. H. Chen[43,44], Wei Cheng[45], Eric F. C. Cheung[46], Siow Ann Chong[47], C. Robert Cloninger[48], David Cohen[49], Nadine Cohen[50], Paul Cormican[5], Nick Craddock[6,7], Benedicto Crespo-Facorro[210], James J. Crowley[51], Michael Davidson[54], Kenneth L. Davis[36], Franziska Degenhardt[55,56], Jurgen Del Favero[57], Lynn E. DeLisi[128,129] , Ditte Demontis[17,58,59], Dimitris Dikeos[60], Timothy Dinan[61], Srdjan Djurovic[14,62], Gary Donohoe[5,63], Elodie Drapeau[36], Jubao Duan[64,65], Frank Dudbridge[66], Naser Durmishi[67], Peter Eichhammer[68], Johan Eriksson[69,70,71], Valentina Escott-Price[6], Laurent Essioux[72], Ayman H. Fanous[73,74,75,76], Martilias S. Farrell[51], Josef Frank[77], Lude Franke[78], Robert Freedman[79], Nelson B. Freimer[80], Marion Friedl[81], Joseph I. Friedman[36], Menachem Fromer[1,2,4,82], Giulio Genovese[2], Lyudmila Georgieva[6], Elliot S. Gershon[209], Ina Giegling[81,83], Paola Giusti-Rodríguez[51], Stephanie Godard[84], Jacqueline I. Goldstein[1,3], Vera Golimbet[85], Srihari Gopal[86], Jacob Gratten[87], Lieuwe de Haan[88], Christian Hammer[23], Marian L. Hamshere[6], Mark Hansen[89], Thomas Hansen[17,90], Vahram Haroutunian[36,91,92], Annette M. Hartmann[81], Frans A. Henskens[39,93,94], Stefan Herms[55,56,95], Joel N. Hirschhorn[3,11,96], Per Hoffmann[55,56,95], Andrea Hofman[55,56], Mads V. Hollegaard[97], David M.

Hougaard[97], Masashi Ikeda[98], Inge Joa[99], Antonio Julià[100], René S. Kahn[34], Luba Kalaydjieva[101,102], Sena Karachanak-Yankova[103], Juha Karjalainen[78], David Kavanagh[6], Matthew C. Keller[104], Brian J. Kelly[120], James L. Kennedy[105,106,107], Andrey Khrunin[108], Yunjung Kim[51], Janis Klovins[109], James A. Knowles[110], Bettina Konte[81], Vaidutis Kucinskas[111], Zita Ausrele Kucinskiene[111], Hana Kuzelova-Ptackova[112], Anna K. Kähler[26], Claudine Laurent[19,113], Jimmy Lee Chee Keong[47,114], S. Hong Lee[87], Sophie E. Legge[6], Bernard Lerer[115], Miaoxin Li[43,44,116] Tao Li[117], Kung-Yee Liang[118], Jeffrey Lieberman[119], Svetlana Limborska[108], Carmel M. Loughland[39,120], Jan Lubinski[121], Jouko Lönnqvist[122], Milan Macek Jr[112], Patrik K. E. Magnusson[26], Brion S. Maher[123], Wolfgang Maier[124], Jacques Mallet[125], Sara Marsal[100], Manuel Mattheisen[17,58,59,126], Morten Mattingsdal[14,127], Robert W. McCarley[128,129], Colm McDonald[130], Andrew M. McIntosh[131,132], Sandra Meier[77], Carin J. Meijer[88], Bela Melegh[24,25], Ingrid Melle[14,133], Raquelle I. Mesholam-Gately[128,134], Andres Metspalu[135], Patricia T. Michie[39,136], Lili Milani[135], Vihra Milanova[137], Younes Mokrab[8], Derek W. Morris[5,63], Ole Mors[17,58,138], Kieran C. Murphy[139], Robin M. Murray[140], Inez Myin-Germeys[141], Bertram Müller-Myhsok[142,143,144], Mari Nelis[135], Igor Nenadic[145], Deborah A. Nertney[146], Gerald Nestadt[147], Kristin K. Nicodemus[148], Liene Nikitina-Zake[109], Laura Nisenbaum[149], Annelie Nordin[150], Eadbhard O'Callaghan[151], Colm O'Dushlaine[2], F. Anthony O'Neill[152], Sang-Yun Oh[153], Ann Olincy[79], Line Olsen[17,90], Jim Van Os[141,154], Psychosis Endophenotypes International Consortium[155], Christos Pantelis[39,156], George N. Papadimitriou[60], Sergi Papiol[23], Elena Parkhomenko[36], Michele T. Pato[110], Tiina Paunio[157,158], Milica Pejovic-Milovancevic[159], Diana O. Perkins[160], Olli Pietiläinen[158,161], Jonathan Pimm[53], Andrew J. Pocklington[6], John Powell[140], Alkes Price[3],[162], Ann E. Pulver[147], Shaun M. Purcell[82], Digby Quested[163], Henrik B. Rasmussen[17,90], Abraham Reichenberg[36], Mark A. Reimers[164], Alexander L. Richards[6], Joshua L. Roffman[30,32], Panos Roussos[82,165], Douglas M. Ruderfer[6,82],

Veikko Salomaa[71], Alan R. Sanders[64,65], Ulrich Schall[39,120], Christian R. Schubert[166], Thomas G. Schulze[77,167], Sibylle G. Schwab[168], Edward M. Scolnick[2], Rodney J. Scott[39,169,170], Larry J. Seidman[128,134], Jianxin Shi[171], Engilbert Sigurdsson[172], Teimuraz Silagadze[173], Jeremy M. Silverman[36,174], Kang Sim[47], Petr Slominsky[108], Jordan W. Smoller[2,4], Hon-Cheong So[43], Chris C. A. Spencer[175], Eli A. Stahl[3,82], Hreinn Stefansson[176], Stacy Steinberg[176], Elisabeth Stogmann[177], Richard E. Straub[178], Eric Strengman[179,34], Jana Strohmaier[77], T. Scott Stroup[119], Mythily Subramaniam[47], Jaana Suvisaari[122], Dragan M. Svrakic[48], Jin P. Szatkiewicz[51], Erik Söderman[12], Srinivas Thirumalai[180], Draga Toncheva[103], Paul A. Tooney[39,120,170] , Sarah Tosato[181], Juha Veijola[182,183], John Waddington[184], Dermot Walsh[185], Dai Wang[86], Qiang Wang[117], Bradley T. Webb[22], Mark Weiser[54], Dieter B. Wildenauer[186], Nigel M. Williams[6], Stephanie Williams[51], Stephanie H. Witt[77], Aaron R. Wolen[164], Emily H. M. Wong[43], Brandon K. Wormley[22], Jing Qin Wu[39,170], Hualin Simon Xi[187], Clement C. Zai[105,106], Xuebin Zheng[188], Fritz Zimprich[177], Naomi R. Wray[87], Kari Stefansson[176], Peter M. Visscher[87], Wellcome Trust Case-Control Consortium 2[189], Rolf Adolfsson[150], Ole A. Andreassen[14,133], Douglas H. R. Blackwood[132], Elvira Bramon[190], Joseph D. Buxbaum[35,36,91,191], Anders D. Børglum[17,58,59,138], Sven Cichon[55,56,95,192], Ariel Darvasi[193], Enrico Domenici[194], Hannelore Ehrenreich[23], Tõnu Esko[3,11,96,135], Pablo V. Gejman[64,65], Michael Gill[5], Hugh Gurling[53], Christina M. Hultman[26], Nakao Iwata[98], Assen V. Jablensky[39,102,186,195], Erik G. Jönsson[12,14], Kenneth S. Kendler[196], George Kirov[6], Jo Knight[105,106,107], Todd Lencz[197,198,199], Douglas F. Levinson[19], Qingqin S. Li[86], Jianjun Liu[188,200], Anil K. Malhotra[197,198,199], Steven A. McCarroll[2,96], Andrew McQuillin[53], Jennifer L. Moran[2], Preben B. Mortensen[15,16,17], Bryan J. Mowry[87,201], Markus M. Nöthen[55,56], Roel A. Ophoff[38,80,34], Michael J. Owen[6,7], Aarno Palotie[2,4,161], Carlos N. Pato[110], Tracey L. Petryshen[2,128,202], Danielle Posthuma[203,204,205], Marcella Rietschel[77], Brien P. Riley[196], Dan Rujescu[81,83], Pak C. Sham[43,44,116]

Pamela Sklar[82,91,165], David St Clair[206], Daniel R. Weinberger[178,207], Jens R. Wendland[166], Thomas Werge[17,90,208], Mark J. Daly[1,2,3], Patrick F. Sullivan[26,51,160] & Michael C. O'Donovan[6,7]

[1]Analytic and Translational Genetics Unit, Massachusetts General Hospital, Boston, Massachusetts 02114, USA.

[2]Stanley Center for Psychiatric Research, Broad Institute of MIT and Harvard, Cambridge, Massachusetts 02142, USA.

[3]Medical and Population Genetics Program, Broad Institute of MIT and Harvard, Cambridge, Massachusetts 02142, USA.

[4]Psychiatric and Neurodevelopmental Genetics Unit, Massachusetts General Hospital, Boston, Massachusetts 02114, USA.

[5]Neuropsychiatric Genetics Research Group, Department of Psychiatry, Trinity College Dublin, Dublin 8, Ireland.

[6]MRC Centre for Neuropsychiatric Genetics and Genomics, Institute of Psychological Medicine and Clinical Neurosciences, School of Medicine, Cardiff University, Cardiff, CF24 4HQ, UK.

[7]National Centre for Mental Health, Cardiff University, Cardiff, CF24 4HQ, UK.

[8]Eli Lilly and Company Limited, Erl Wood Manor, Sunninghill Road, Windlesham, Surrey, GU20 6PH, UK.

[9]Social, Genetic and Developmental Psychiatry Centre, Institute of Psychiatry, King's College London, London, SE5 8AF, UK.

[10]Center for Biological Sequence Analysis, Department of Systems Biology, Technical University of Denmark, DK-2800, Denmark.

[11]Division of Endocrinology and Center for Basic and Translational Obesity Research, Boston Children's Hospital, Boston, Massachusetts, 02115USA.

[12]Department of Clinical Neuroscience, Psychiatry Section, Karolinska Institutet, SE-17176 Stockholm, Sweden.

[13]Department of Psychiatry, Diakonhjemmet Hospital, 0319 Oslo, Norway.

[14]NORMENT, KG Jebsen Centre for Psychosis Research, Institute of Clinical Medicine, University of Oslo, 0424 Oslo, Norway.

[15]Centre for Integrative Register-based Research, CIRRAU, Aarhus University, DK-8210 Aarhus, Denmark.

[16]National Centre for Register-based Research, Aarhus University, DK-8210 Aarhus, Denmark.

[17]The Lundbeck Foundation Initiative for Integrative Psychiatric Research, iPSYCH, Denmark.

[18]State Mental Hospital, 85540 Haar, Germany.

[19]Department of Psychiatry and Behavioral Sciences, Stanford University, Stanford, California 94305, USA.

[20]Department of Psychiatry and Behavioral Sciences, Atlanta Veterans Affairs Medical Center, Atlanta, Georgia 30033, USA.

[21]Department of Psychiatry and Behavioral Sciences, Emory University, Atlanta Georgia 30322, USA.

[22]Virginia Institute for Psychiatric and Behavioral Genetics, Department of Psychiatry, Virginia Commonwealth University, Richmond, Virginia 23298, USA.

[23]Clinical Neuroscience, Max Planck Institute of Experimental Medicine, Göttingen 37075, Germany.

[24]Department of Medical Genetics, University of Pécs, Pécs H-7624, Hungary.

[25]Szentagothai Research Center, University of Pécs, Pécs H-7624, Hungary.

[26]Department of Medical Epidemiology and Biostatistics, Karolinska Institutet, Stockholm SE-17177, Sweden.

[27]Department of Psychiatry, University of Iowa Carver College of Medicine, Iowa City, Iowa 52242, USA.

[28]University Medical Center Groningen, Department of Psychiatry, University of Groningen NL-9700 RB, The Netherlands.

[29]School of Nursing, Louisiana State University Health Sciences Center, New Orleans, Louisiana 70112, USA.

[30]Athinoula A. Martinos Center, Massachusetts General Hospital, Boston, Massachusetts 02129, USA.

[31]Center for Brain Science, Harvard University, Cambridge, Massachusetts, 02138 USA.

[32]Department of Psychiatry, Massachusetts General Hospital, Boston, Massachusetts, 02114 USA.

[33]Department of Psychiatry, University of California at San Francisco, San Francisco, California, 94143 USA.

[34]University Medical Center Utrecht, Department of Psychiatry, Rudolf Magnus Institute of Neuroscience, 3584 Utrecht, The Netherlands.

[35]Department of Human Genetics, Icahn School of Medicine at Mount Sinai, New York, New York 10029 USA.

[36]Department of Psychiatry, Icahn School of Medicine at Mount Sinai, New York, New York 10029 USA.

[37]Centre Hospitalier du Rouvray and INSERM U1079 Faculty of Medicine, 76301 Rouen, France.

[38]Department of Human Genetics, David Geffen School of Medicine, University of California, Los Angeles, California 90095, USA.

[39]Schizophrenia Research Institute, Sydney NSW 2010, Australia.

[40]School of Psychiatry, University of New South Wales, Sydney NSW 2031, Australia.

[41]Royal Brisbane and Women's Hospital, University of Queensland, Brisbane, St Lucia QLD 4072, Australia.

[42]Institute of Psychology, Chinese Academy of Science, Beijing 100101, China.

[43]Department of Psychiatry, Li Ka Shing Faculty of Medicine, The University of Hong Kong, Hong Kong, China.

[44]State Key Laboratory for Brain and Cognitive Sciences, Li Ka Shing Faculty of Medicine, The University of Hong Kong, Hong Kong, China.

[45]Department of Computer Science, University of North Carolina, Chapel Hill, North Carolina 27514, USA.

[46]Castle Peak Hospital, Hong Kong, China.

[47]Institute of Mental Health, Singapore 539747, Singapore.

[48]Department of Psychiatry, Washington University, St. Louis, Missouri 63110, USA.

[49]Department of Child and Adolescent Psychiatry, Assistance Publique Hopitaux de Paris, Pierre and Marie Curie Faculty of Medicine and Institute for Intelligent Systems and Robotics, Paris, 75013, France.

[50] Blue Note Biosciences, Princeton, New Jersey 08540, USA

[51]Department of Genetics, University of North Carolina, Chapel Hill, North Carolina 27599-7264, USA.

[52]Department of Psychological Medicine, Queen Mary University of London, London E1 1BB, UK.

[53]Molecular Psychiatry Laboratory, Division of Psychiatry, University College London, London WC1E 6JJ, UK.

[54]Sheba Medical Center, Tel Hashomer 52621, Israel.

[55]Department of Genomics, Life and Brain Center, D-53127 Bonn, Germany.

[56]Institute of Human Genetics, University of Bonn, D-53127 Bonn, Germany.

[57]Applied Molecular Genomics Unit, VIB Department of Molecular Genetics, University of Antwerp, B-2610 Antwerp, Belgium.

[58]Centre for Integrative Sequencing, iSEQ, Aarhus University, DK-8000 Aarhus C, Denmark.

[59]Department of Biomedicine, Aarhus University, DK-8000 Aarhus C, Denmark.

[60]First Department of Psychiatry, University of Athens Medical School, Athens 11528, Greece.

[61]Department of Psychiatry, University College Cork, Co. Cork, Ireland.

[62]Department of Medical Genetics, Oslo University Hospital, 0424 Oslo, Norway.

[63]Cognitive Genetics and Therapy Group, School of Psychology and Discipline of Biochemistry, National University of Ireland Galway, Co. Galway, Ireland.

[64]Department of Psychiatry and Behavioral Neuroscience, University of Chicago, Chicago, Illinois 60637, USA.

[65]Department of Psychiatry and Behavioral Sciences, NorthShore University HealthSystem, Evanston, Illinois 60201, USA.

[66]Department of Non-Communicable Disease Epidemiology, London School of Hygiene and Tropical Medicine, London WC1E 7HT, UK.

[67]Department of Child and Adolescent Psychiatry, University Clinic of Psychiatry, Skopje 1000, Republic of Macedonia.

[68]Department of Psychiatry, University of Regensburg, 93053 Regensburg, Germany.

[69]Department of General Practice, Helsinki University Central Hospital, University of Helsinki P.O. Box 20, Tukholmankatu 8 B, FI-00014, Helsinki, Finland

[70]Folkhälsan Research Center, Helsinki, Finland, Biomedicum Helsinki 1, Haartmaninkatu 8, FI-00290, Helsinki, Finland.

[71]National Institute for Health and Welfare, P.O. BOX 30, FI-00271 Helsinki, Finland.

[72]Translational Technologies and Bioinformatics, Pharma Research and Early Development, F. Hoffman-La Roche, CH-4070 Basel, Switzerland.

[73]Department of Psychiatry, Georgetown University School of Medicine, Washington DC 20057, USA.

[74]Department of Psychiatry, Keck School of Medicine of the University of Southern California, Los Angeles, California 90033, USA.

[75]Department of Psychiatry, Virginia Commonwealth University School of Medicine, Richmond, Virginia 23298, USA.

[76]Mental Health Service Line, Washington VA Medical Center, Washington DC 20422, USA.

[77]Department of Genetic Epidemiology in Psychiatry, Central Institute of Mental Health, Medical Faculty Mannheim, University of Heidelberg, Heidelberg , D-68159 Mannheim, Germany.

[78]Department of Genetics, University of Groningen, University Medical Centre Groningen, 9700 RB Groningen, The Netherlands.

[79]Department of Psychiatry, University of Colorado Denver, Aurora, Colorado 80045, USA.

[80]Center for Neurobehavioral Genetics, Semel Institute for Neuroscience and Human Behavior, University of California, Los Angeles, California 90095, USA.

[81]Department of Psychiatry, University of Halle, 06112 Halle, Germany.

[82]Division of Psychiatric Genomics, Department of Psychiatry, Icahn School of Medicine at Mount Sinai, New York, New York 10029, USA.

[83]Department of Psychiatry, University of Munich, 80336, Munich, Germany.

[84]Departments of Psychiatry and Human and Molecular Genetics, INSERM, Institut de Myologie, Hôpital de la Pitiè-Salpêtrière, Paris, 75013, France.

[85]Mental Health Research Centre, Russian Academy of Medical Sciences, 115522 Moscow, Russia.

[86]Neuroscience Therapeutic Area, Janssen Research and Development, Raritan, New Jersey 08869, USA.

[87]Queensland Brain Institute, The University of Queensland, Brisbane, Queensland, QLD 4072, Australia.

[88]Academic Medical Centre University of Amsterdam, Department of Psychiatry, 1105 AZ Amsterdam, The Netherlands.

[89]Illumina, La Jolla, California, California 92122, USA.

[90]Institute of Biological Psychiatry, Mental Health Centre Sct. Hans, Mental Health Services Copenhagen, DK-4000, Denmark.

[91]Friedman Brain Institute, Icahn School of Medicine at Mount Sinai, New York, New York 10029, USA.

[92]J. J. Peters VA Medical Center, Bronx, New York, New York 10468, USA.

[93]Priority Research Centre for Health Behaviour, University of Newcastle, Newcastle NSW 2308, Australia.

[94]School of Electrical Engineering and Computer Science, University of Newcastle, Newcastle NSW 2308, Australia.

[95]Division of Medical Genetics, Department of Biomedicine, University of Basel, Basel, CH-4058, Switzerland.

[96]Department of Genetics, Harvard Medical School, Boston, Massachusetts 02115, USA.

[97]Section of Neonatal Screening and Hormones, Department of Clinical Biochemistry, Immunology and Genetics, Statens Serum Institut, Copenhagen, DK-2300, Denmark.

[98]Department of Psychiatry, Fujita Health University School of Medicine, Toyoake, Aichi, 470-1192, Japan.

[99]Regional Centre for Clinical Research in Psychosis, Department of Psychiatry, Stavanger University Hospital, 4011 Stavanger, Norway.

[100]Rheumatology Research Group, Vall d'Hebron Research Institute, Barcelona, 08035, Spain.

[101]Centre for Medical Research, The University of Western Australia, Perth, WA 6009, Australia.

[102]The Perkins Institute for Medical Research, The University of Western Australia, Perth, WA 6009, Australia.

[103]Department of Medical Genetics, Medical University, Sofia1431, Bulgaria.

[104]Department of Psychology, University of Colorado Boulder, Boulder, Colorado 80309, USA.

[105]Campbell Family Mental Health Research Institute, Centre for Addiction and Mental Health, Toronto, Ontario, M5T 1R8, Canada.

[106]Department of Psychiatry, University of Toronto, Toronto, Ontario, M5T 1R8, Canada.

[107]Institute of Medical Science, University of Toronto, Toronto, Ontario, M5S 1A8, Canada.

[108]Institute of Molecular Genetics, Russian Academy of Sciences, Moscow123182, Russia.

[109]Latvian Biomedical Research and Study Centre, Riga, LV-1067, Latvia.

[110]Department of Psychiatry and Zilkha Neurogenetics Institute, Keck School of Medicine at University of Southern California, Los Angeles, California 90089, USA.

[111]Faculty of Medicine, Vilnius University, LT-01513 Vilnius, Lithuania.

[112] Department of Biology and Medical Genetics, 2nd Faculty of Medicine and University Hospital Motol, 150 06 Prague, Czech Republic.

[113] Department of Child and Adolescent Psychiatry, Pierre and Marie Curie Faculty of Medicine, Paris 75013, France.

[114]Duke-NUS Graduate Medical School, Singapore 169857, Singapore.

[115]Department of Psychiatry, Hadassah-Hebrew University Medical Center, Jerusalem 91120, Israel.

[116]Centre for Genomic Sciences, The University of Hong Kong, Hong Kong, China.

[117]Mental Health Centre and Psychiatric Laboratory, West China Hospital, Sichuan University, Chengdu, 610041, Sichuan, China.

[118]Department of Biostatistics, Johns Hopkins University Bloomberg School of Public Health, Baltimore, Maryland 21205, USA.

[119]Department of Psychiatry, Columbia University, New York, New York 10032, USA.

[120]Priority Centre for Translational Neuroscience and Mental Health, University of Newcastle, Newcastle NSW 2300, Australia.

[121]Department of Genetics and Pathology, International Hereditary Cancer Center, Pomeranian Medical University in Szczecin, 70-453 Szczecin, Poland.

[122]Department of Mental Health and Substance Abuse Services; National Institute for Health and Welfare, P.O. BOX 30, FI-00271 Helsinki, Finland

[123]Department of Mental Health, Bloomberg School of Public Health, Johns Hopkins University, Baltimore, Maryland 21205, USA.

[124]Department of Psychiatry, University of Bonn, D-53127 Bonn, Germany.

[125]Centre National de la Recherche Scientifique, Laboratoire de Génétique Moléculaire de la Neurotransmission et des Processus Neurodégénératifs, Hôpital de la Pitié Salpêtrière, 75013, Paris, France.

[126]Department of Genomics Mathematics, University of Bonn, D-53127 Bonn, Germany.

[127]Research Unit, Sørlandet Hospital, 4604 Kristiansand, Norway.

[128]Department of Psychiatry, Harvard Medical School, Boston, Massachusetts 02115, USA.

[129]VA Boston Health Care System, Brockton, Massachusetts 02301, USA.

[130]Department of Psychiatry, National University of Ireland Galway, Co. Galway, Ireland.

[131]Centre for Cognitive Ageing and Cognitive Epidemiology, University of Edinburgh, Edinburgh EH16 4SB, UK.

[132]Division of Psychiatry, University of Edinburgh, Edinburgh EH16 4SB, UK.

[133]Division of Mental Health and Addiction, Oslo University Hospital, 0424 Oslo, Norway.

[134]Massachusetts Mental Health Center Public Psychiatry Division of the Beth Israel Deaconess Medical Center, Boston, Massachusetts 02114, USA.

[135]Estonian Genome Center, University of Tartu, Tartu 50090, Estonia.

[136]School of Psychology, University of Newcastle, Newcastle NSW 2308, Australia.

[137]First Psychiatric Clinic, Medical University, Sofia 1431, Bulgaria.

[138]Department P, Aarhus University Hospital, DK-8240 Risskov, Denmark.

[139]Department of Psychiatry, Royal College of Surgeons in Ireland, Dublin 2, Ireland.

[140]King's College London, London SE5 8AF, UK.

[141]Maastricht University Medical Centre, South Limburg Mental Health Research and Teaching Network, EURON, 6229 HX Maastricht, The Netherlands.

[142]Institute of Translational Medicine, University of Liverpool, Liverpool L69 3BX, UK.

[143]Max Planck Institute of Psychiatry, 80336 Munich, Germany.

[144]Munich Cluster for Systems Neurology (SyNergy), 80336 Munich, Germany.

[145]Department of Psychiatry and Psychotherapy, Jena University Hospital, 07743 Jena, Germany.

[146]Department of Psychiatry, Queensland Brain Institute and Queensland Centre for Mental Health Research, University of Queensland, Brisbane, Queensland, St Lucia QLD 4072, Australia.

[147]Department of Psychiatry and Behavioral Sciences, Johns Hopkins University School of Medicine, Baltimore, Maryland 21205, USA.

[148]Department of Psychiatry, Trinity College Dublin, Dublin 2, Ireland.

[149]Eli Lilly and Company, Lilly Corporate Center, Indianapolis, 46285 Indiana, USA.

[150]Department of Clinical Sciences, Psychiatry, Umeå University, SE-901 87 Umeå, Sweden.

[151]DETECT Early Intervention Service for Psychosis, Blackrock, Co. Dublin, Ireland.

[152]Centre for Public Health, Institute of Clinical Sciences, Queen's University Belfast, Belfast BT12 6AB, UK.

[153]Lawrence Berkeley National Laboratory, University of California at Berkeley, Berkeley, California 94720, USA.

[154]Institute of Psychiatry, King's College London, London SE5 8AF, UK.

[155]A list of authors and affiliations appear in the Supplementary Information.

[156]Melbourne Neuropsychiatry Centre, University of Melbourne & Melbourne Health, Melbourne, Vic 3053, Australia.

[157]Department of Psychiatry, University of Helsinki, P.O. Box 590, FI-00029 HUS, Helsinki, Finland.

[158]Public Health Genomics Unit, National Institute for Health and Welfare, P.O. BOX 30, FI-00271 Helsinki, Finland.

[159]Medical Faculty, University of Belgrade, 11000 Belgrade, Serbia.

[160]Department of Psychiatry, University of North Carolina, Chapel Hill, North Carolina 27599-7160, USA.

[161]Institute for Molecular Medicine Finland, FIMM, University of Helsinki, P.O. Box 20

FI-00014, Helsinki, Finland.

[162]Department of Epidemiology, Harvard School of Public Health, Boston, Massachusetts 02115, USA.

[163]Department of Psychiatry, University of Oxford, Oxford, OX3 7JX, UK.

[164]Virginia Institute for Psychiatric and Behavioral Genetics, Virginia Commonwealth University, Richmond, Virginia 23298, USA.

[165]Institute for Multiscale Biology, Icahn School of Medicine at Mount Sinai, New York, New York 10029, USA.

[166]PharmaTherapeutics Clinical Research, Pfizer Worldwide Research and Development, Cambridge, Massachusetts 02139, USA.

[167]Department of Psychiatry and Psychotherapy, University of Gottingen, 37073 Göttingen, Germany.

[168]Psychiatry and Psychotherapy Clinic, University of Erlangen, 91054 Erlangen, Germany.

[169]Hunter New England Health Service, Newcastle NSW 2308, Australia.

[170]School of Biomedical Sciences and Pharmacy, University of Newcastle, Callaghan NSW 2308, Australia.

[171]Division of Cancer Epidemiology and Genetics, National Cancer Institute, Bethesda, Maryland 20892, USA.

[172]University of Iceland, Landspitali, National University Hospital, 101 Reykjavik, Iceland.

[173]Department of Psychiatry and Drug Addiction, Tbilisi State Medical University (TSMU), **N33, 0177** Tbilisi, Georgia.

[174]Research and Development, Bronx Veterans Affairs Medical Center, New York, New York 10468, USA.

[175]Wellcome Trust Centre for Human Genetics, Oxford, OX3 7BN, UK.

[176]deCODE Genetics, 101 Reykjavik, Iceland.

[177]Department of Clinical Neurology, Medical University of Vienna, 1090 Wien, Austria.

[178]Lieber Institute for Brain Development, Baltimore, Maryland 21205, USA.

[179]Department of Medical Genetics, University Medical Centre Utrecht, Universiteitsweg 100, 3584 CG, Utrecht, The Netherlands.

[180]Berkshire Healthcare NHS Foundation Trust, Bracknell RG12 1BQ, UK.

[181]Section of Psychiatry, University of Verona, 37134 Verona, Italy.

[182]Department of Psychiatry, University of Oulu, P.O. BOX 5000, 90014, Finland

[183]University Hospital of Oulu, P.O.BOX 20, 90029 OYS, Finland.

[184]Molecular and Cellular Therapeutics, Royal College of Surgeons in Ireland, Dublin 2, Ireland.

[185]Health Research Board, Dublin 2, Ireland.

[186]School of Psychiatry and Clinical Neurosciences, The University of Western Australia, Perth WA6009, Australia.

[187]Computational Sciences CoE, Pfizer Worldwide Research and Development, Cambridge, Massachusetts 02139, USA.

[188]Human Genetics, Genome Institute of Singapore, A*STAR, Singapore 138672, Singapore.

[189]A list of authors and affiliations appear in the Supplementary Information.

[190]University College London, London WC1E 6BT, UK.

[191]Department of Neuroscience, Icahn School of Medicine at Mount Sinai, New York, New York 10029, USA.

[192]Institute of Neuroscience and Medicine (INM-1), Research Center Juelich, 52428 Juelich, Germany.

[193]Department of Genetics, The Hebrew University of Jerusalem, 91905 Jerusalem, Israel.

[194]Neuroscience Discovery and Translational Area, Pharma Research and Early Development, F. Hoffman-La Roche, CH-4070 Basel, Switzerland.

[195]Centre for Clinical Research in Neuropsychiatry, School of Psychiatry and Clinical Neurosciences, The University of Western Australia, Medical Research Foundation Building, Perth WA 6000, Australia.

[196]Virginia Institute for Psychiatric and Behavioral Genetics, Departments of Psychiatry and Human and Molecular Genetics, Virginia Commonwealth University, Richmond, Virginia 23298, USA.

[197]The Feinstein Institute for Medical Research, Manhasset, New York, 11030 USA.

[198]The Hofstra NS-LIJ School of Medicine, Hempstead, New York, 11549 USA.

[199]The Zucker Hillside Hospital, Glen Oaks, New York,11004 USA.

[200]Saw Swee Hock School of Public Health, National University of Singapore, Singapore 117597, Singapore.

[201]Queensland Centre for Mental Health Research, University of Queensland, Brisbane 4076, Queensland, Australia.

[202]Center for Human Genetic Research and Department of Psychiatry, Massachusetts General Hospital, Boston, Massachusetts 02114, USA.

[203]Department of Child and Adolescent Psychiatry, Erasmus University Medical Centre, Rotterdam 3000, The Netherlands.

[204]Department of Complex Trait Genetics, Neuroscience Campus Amsterdam, VU University Medical Center Amsterdam, Amsterdam 1081, The Netherlands.

[205]Department of Functional Genomics, Center for Neurogenomics and Cognitive Research, Neuroscience Campus Amsterdam, VU University, Amsterdam 1081, The Netherlands.

[206]University of Aberdeen, Institute of Medical Sciences, Aberdeen, AB25 2ZD, UK.

[207]Departments of Psychiatry, Neurology, Neuroscience and Institute of Genetic Medicine, Johns Hopkins School of Medicine, Baltimore, Maryland 21205, USA.

[208]Department of Clinical Medicine, University of Copenhagen, Copenhagen 2200, Denmark.

[209]Departments of Psychiatry and Human Genetics, University of Chicago, Chicago, Illinois 60637, USA.

[210]University Hospital Marqués de Valdecilla, Instituto de Formación e Investigación Marqués de Valdecilla, University of Cantabria, E-39008 Santander, Spain.
